# Emergence and spread of a B.1.1.28-derived P.6 lineage with Q675H and Q677H Spike mutations in Uruguay

**DOI:** 10.1101/2021.07.27.21261150

**Authors:** Natalia Rego, Cecilia Salazar, Mercedes Paz, Alicia Costábile, Alvaro Fajardo, Ignacio Ferrés, Paula Perbolianachis, Tamara Fernández-Calero, Veronica Noya, Matias R. Machado, Rodrigo Arce, Mailen Arleo, Tania Possi, Natalia Reyes, María Noel Bentancor, Andrés Lizasoain, Viviana Bortagaray, Ana Moller, Odhille Chappos, Nicolas Nin, Javier Hurtado, Melissa Duquía, Belén González, Luciana Griffero, Mauricio Méndez, Ma Pía Techera, Juan Zanetti, Bernardina Rivera, Matías Maidana, Martina Alonso, Cecilia Alonso, Julio Medina, Henry Albornoz, Rodney Colina, Gonzalo Bello, Pilar Moreno, Gonzalo Moratorio, Gregorio Iraola, Lucía Spangenberg

## Abstract

Uruguay was able to control the viral dissemination during the first nine months of the SARS-CoV-2 pandemic. Unfortunately, towards the end of 2020, the number of daily new cases exponentially increased. Herein we analyzed the country-wide genetic diversity of SARS-CoV-2 between November, 2020 and April, 2021. Our findings identified that the most prevalent viral variant during late 2020 was a B.1.1.28 sublineage carrying mutations Q675H+Q677H in the viral Spike, now designated as lineage P.6. This new lineage P.6 probably arose around November 2020, in Montevideo, Uruguay’s capital department and rapidly spread to other Uruguayan departments, with evidence of further local transmission clusters, also spread sporadically to the USA and Spain. The Q675H and Q677H mutations are in the proximity of the polybasic cleavage site at the S1/S2 boundary and also arose independently in many SARS-CoV-2 lineages circulating worldwide. Although the lineage P.6 was replaced by the Variant of Concern (VOC) P.1 as the predominant viral strain in Uruguay since April 2021, the monitoring of the concurrent emergence of Q675H+Q677H in VOCs should be of worldwide interest.

## Introduction

Uruguay was able to control the early viral dissemination during the first nine months of the SARS-CoV-2 pandemic. Unfortunately, towards the end of 2020, the number of newly daily cases exponentially increased, from 60 cases per day on average during October and November to over 400 during December [1,2]. This coincided with the loss of the TETRIS (Test, Trace and Isolation strategy) safety zone [3,4]. With a 1,068 km long Uruguayan-Brazilian dry border, multiple introductions and successful dissemination of Brazilian lineages B.1.1.28, B.1.1.33, P.1 (Gamma) and P.2 have occurred during 2020 and 2021 [5–7]. Little is known, however, regarding the SARS-CoV-2 strains that circulated during the first exponential increase of COVID-19 cases by the end of 2020, before the arrival of the VOI P.2 and the VOC P.1 (which earliest samples are from January and February 2021, respectively) [6,7].

In a previous study we surveyed patients diagnosed between November 2020 and February 2021 in Rocha, an eastern Uruguayan department bordering Brazil, where we found that lineage B.1.1.28 was the most prevalent during November-December, 2020 in that location [7]. Many B.1.1.28 sequences branched in a clade highly prevalent in southern Brazil, while others branched in a second monophyletic cluster harboring several lineage-defining mutations including two non-synonymous changes in the Spike protein: Q675H and Q677H, so far not concurrently reported. The convergent appearance of S:Q677H in different viral lineages and its proximity to the S1/S2 cleavage site raised concerns about its functional relevance [8,9]. To understand the SARS-CoV-2 strains associated with the first COVID-19 epidemic wave in Uruguay, herein we analyzed the genetic diversity of viruses circulating in different country localities by the end of 2020 and first months of 2021.

## Methods

### Ethics statement

This work was done by the Inter-Institutional Working Group (IiWG) for SARS-CoV-2 genomic surveillance in Uruguay, which involves a diagnostic network, expertise and resources to handle large-scale sequencing, computational scientists for genomic analysis, and an affordable and decentralized “in house” qPCR test designed to detect known VOCs [6]. Residual de-identified RNA samples from SARS-CoV-2 positive patients were remitted to the Institut Pasteur de Montevideo (IPMon). IPMon was validated by the Ministry of Health of Uruguay as an approved center providing diagnostic testing for COVID-19. All samples were de-identified before receipt by the study investigators. All relevant ethical guidelines have been appropriately followed. Additionally, the project was approved by the Ethics Committee of the Sanatorio Americano SASA (Uruguay). Ethical approval was given and signed informed consent was obtained from the participants.

### SARS-CoV-2 samples

SARS-CoV-2 RNA samples were recovered from nasopharyngeal-throat combined swabs collected from clinically ill or asymptomatic individuals that reside in different Uruguayan departments and were diagnosed from December 2020 to April 2021 in Uruguay. While our target period of study is November 2020 to April 2021, residual patient samples from November were not available at any laboratory of the IiGW diagnostic network (the IiWG started working as such in March 2021). Positive RNA samples were reverse transcribed using SuperScript™ II Reverse Transcriptase (Thermo Fisher Scientific Inc., MA, USA) or the LunaScript® RT SuperMix Kit (New England Biolabs, Ipswich, MA, USA). A negative control was included at this point and carried throughout the protocol. Samples were initially screened by the VOC-qPCR assay, and those VOC-negative were further processed for whole-genome sequencing.

### Genome sequencing

Sequencing libraries were prepared according to the classic ARTIC protocol described by Quick J. [10,11] or the SARS-CoV2 genome sequencing protocol (1200bp amplicon “midnight” primer set, using Nanopore Rapid kit) described by Freed N. and Silander O. [12,13] with minor modifications (**Table S1**). Final library was eluted in EB buffer (ONT) and quantified using a fluorometric assay. Recommended amounts of library were loaded into a FLO-MIN106D R9.4.1 flowcell and sequenced on the GridION X5 sequencing platform (ONT). Basecalling and demultiplexing was performed with Guppy 4.3.2 or higher [14] using the high or super accuracy mode. Consensus genomes were generated using the poreCov pipeline [15–24] and Nanopolish was used for consensus generation. Complete sequences with up to 15% of Ns were kept for further analysis. All genomes obtained in this study were uploaded to the EpiCoV database in the GISAID initiative under the accession numbers EPI_ISL_XX to EPI_ISL_XX (**Table S1**).

### Phylogenetic and phylogeographic analysis

SARS-CoV-2 full-length consensus sequences were manually curated in specific genome positions, such as clade defining mutations. Genotyping was performed according to Rambaut et al. [25] using the Pangolin application [26] and later confirmed using maximum likelihood (ML) analysis. Uruguayan B.1.1.28 sequences (n=174) were next analyzed in the context of additional B.1.1.28 sequences from Uruguay and Brazil, downloaded from the EpiCoV database of the GISAID initiative [27] (**Table S3** with GISAID Acknowledgements). Downloaded B.1.1.28 sequences from Uruguay (n=184) were complete, with full collection date information and sampled before May 31, 2021. Sequences from Brazil (n=1,428) were complete, high quality, with full collection date information and sampled before May 31, 2021 (**Table S2**). Additionally, we downloaded four B.1.1.28 from USA (n=2), Spain (n=1) and Belgium (n=1) which also harbor both S:Q675H and S:Q677H mutations (Table S2 and Table S3). Finally, we added to the B.1.1.28 dataset six available sequences collected by the IiWG out of the target period of this work (**Table S1**). Alignment was performed with MAFFT v7.471 [28]. Maximum likelihood (ML) phylogenetic analysis of the 1,796 B.1.1.28 sequences was performed with IQ-TREE version 1.6.12 under the model GTR+F+R3 of nucleotide substitution selected by the built-in ModelFinder option [29]. Branch support was assessed by the approximate likelihood-ratio test based on a Shimodaira–Hasegawa-like procedure (SH-aLRT) with 1,000 replicates [30]. The tree root was established with the sequence EPI_ISL_416036 with the earliest collection date 2020-03-05. This tree was time-scaled using TreeTime [31] applying a fixed clock rate of 8×10^−4^ substitutions/site/year [32,33], keeping polytomies. The time-scaled tree was then employed for the ancestral character state reconstruction (ACR) of epidemic locations with PastML [34], using the Marginal Posterior Probabilites Approximation (MPPA) method with an F81-like model. Brazilian sequences were grouped according to the region: South, Southeast, Central West, North and Northeast. A time-scaled Bayesian phylogeographic analysis was next performed to infer the geographical source and dissemination pattern of the Uruguayan B.1.1.28+Q675H+Q677H samples and to estimate the time of their most recent common ancestors (T_MRCA_). Phylogenetic trees were estimated in BEAST v1.10 [35] using the GTR+F+I nucleotide substitution model, the non-parametric Bayesian skyline model as the coalescent tree prior [36], a strict molecular clock model with a uniform substitution rate prior (8-10 ×10^−4^ substitutions/site/year) and a reversible discrete phylogeographic model (using Uruguayan departments as epidemic locations) [37] with a continuous-time Markov chain (CTMC) rate reference prior [38]. MCMC chains were run for 100 million generations and convergence (Effective Sample Size > 200) in parameter estimates was assessed using Tracer v1.7 [39]. Maximum clade credibility (MCC) tree was summarized with TreeAnnotator v1.10 [40] and visualized using FigTree v1.4.4 [41]. Additional visualizations were implemented in the R environment with treeio and ggtree Bioconductor packages [42].

### Determination of prevalence of Q675H+Q677H

To assess the prevalence of co-occurring S:Q675H and S:Q677H in worldwide SARS-CoV-2 genomes, we downloaded from GISAID (accessed on July 7, 2021) 129 complete genomes, with high quality and full collection date information (**Table S2-S3**).

### Structural representation of the Spike protein

The molecular model of SARS-CoV-2 spike glycoprotein was taken from *D. E. Shaw Research database* (DESRES-ANTON-11021566) [43]. The visual rendering was done with VMD 1.9.3 [44].

## Results

The Uruguayan inter-institutional working group (IiWG) for SARS-CoV-2 genomic surveillance sequenced 567 SARS-CoV-2 positive samples detected in Uruguay between December 2020 and April 2021, that were classified in the following lineages: 180 B.1.1.28 (**Table S1**), 333 P.1, 36 P.2 and 36 other B.1-derived lineages. The mutational profile identified 163 B.1.1.28 sequences (91%) carrying both S:Q675H and S:Q677H amino acid changes that were widely spread throughout the country, being detected in 12 out of 19 Uruguayan departments from 2nd December, 2020 to 26th April, 2021 (**Figure 1A**). Analysis of all SARS-CoV Uruguayan sequences determined by the IiWG between November 2020 and April 2021 plus additional Uruguayan sequences obtained from GISAID database revealed that the relative frequency of B.1.1.28+Q675H+Q677H sequences increase from 26% in December 2020 up to 76% in February 2021 (**Figure 1B)** and was later replaced by the VOC P.1 as the most prevalent lineage through the country as previously described [6].

**Figure 1.**
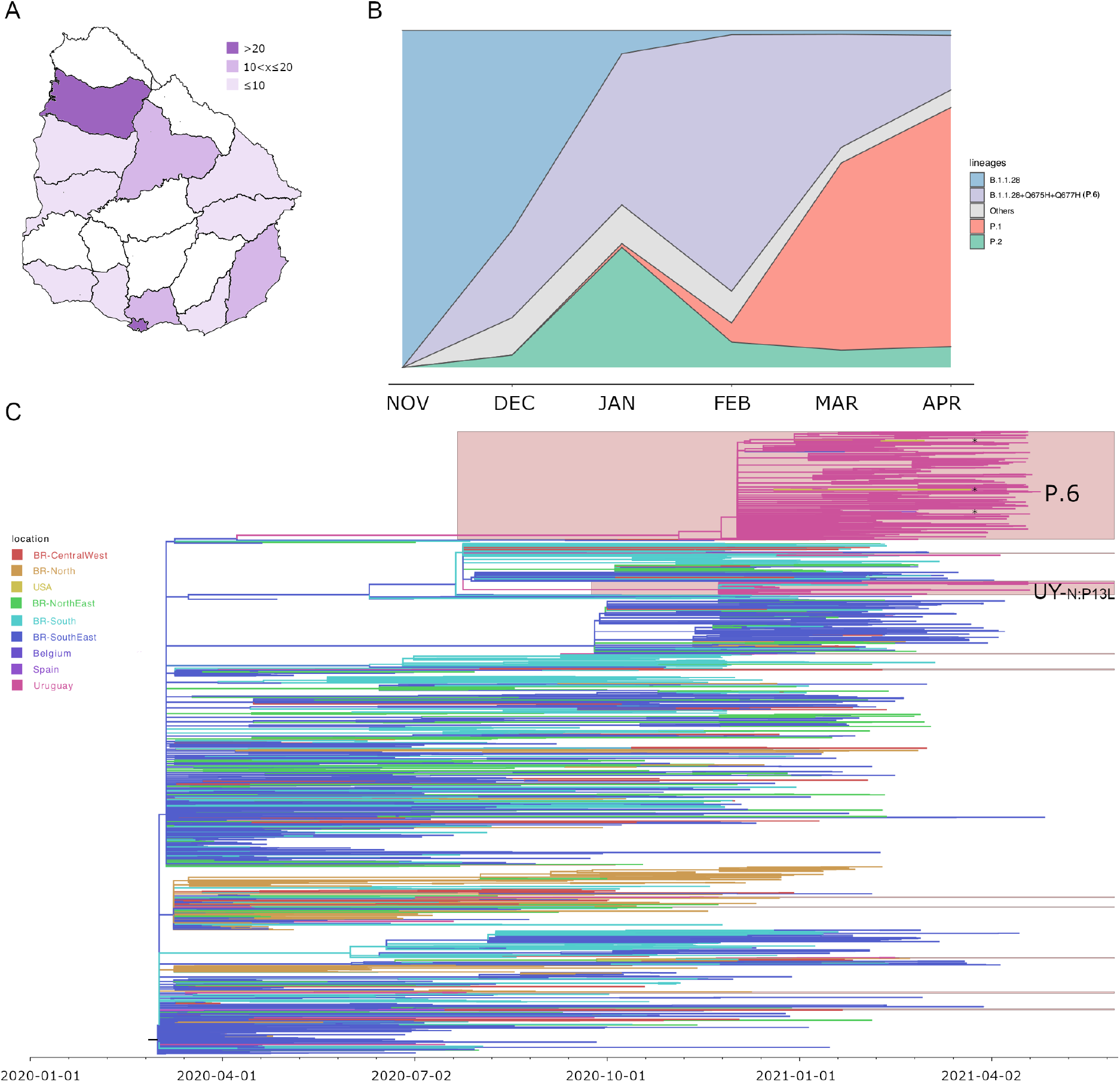
**A**. Map of Uruguay showing the number of sequences classified as P.6 (B.1.1.28+Q675H+Q677H) in every department (n=163). **B**. Lineage proportions of all available Uruguayan samples calculated monthly, from November, 2020 to April, 2021. The ‘Others’ category includes B.1, B.1.1, B.1.1.1, B.1.1.34, B.1.177, B.1.177.12. **C**. Maximum likelihood phylogeographic analysis of lineage B.1.1.28 samples (n=1,786) from Uruguay (n=364), Brazil (n=1,418) and USA, Spain and Belgium (n=4) inferred by ancestral character reconstruction method implemented in PastML. Tips and branches are colored according to sampling location and the most probable location state of their descendent nodes, respectively, as indicated in the legend. Shaded boxes highlight the major B.1.1.28 clades in Uruguay. P.6 is the assigned pango name to the clade B.1.1.28+Q675H+Q677H here discussed while UY-N:P13L is a B.1.1.28 clade carrying mutation N:P13L widely distributed in southern Brazil. Asterisks indicate the sequences dispersed from Uruguay to the USA and Spain. The time scaled tree was rooted with the earliest sequence (collection date 2020-03-05).

All B.1.1.28 Uruguayan sequences here obtained (**Table S1**) were combined with B.1.1.28 complete genome sequences available at the EpiCoV database in GISAID by June 07, 2021 sampled in Uruguay (n=184) and Brazil (n=1,428) and with all B.1.1.28 sequences sampled worldwide (USA=2, Spain=1 and Belgium=1) that carry mutations Q675H and Q677H (**Table S2**). The ML tree of the B.1.1.28 dataset supports at least 20 independent introductions of this lineage from Brazil into Uruguay (**Figure 1C** and **Figure S1**). Fourteen introductions resulted in singletons or dyads with no evidence of further dissemination. Six introductions resulted in highly supported clades (SH-aLRT>0.79) with at least three Uruguayan samples. All 314 Uruguayan sequences carrying mutations Q675H+Q677H branched in a highly supported (SH-aLRT>0.98) monophyletic clade, now designated as the new P.6 pango lineage [25]. This clade also comprises two sequences collected in the USA and one sequence sampled in Spain. According to the ML-based ACR of epidemic locations, the lineage P.6 was most likely introduced from the southeastern Brazilian region (ACR location marginal probability = 0.99) and was disseminated from Uruguay to the USA (two independent times, ACR location marginal probability = 0.99 and 1.00, respectively) and Spain (ACR location marginal probability = 0.99) (**Figure 1C** and **Figure S2**). The second largest monophyletic clade designated as UY-N:P13L (**Figure 1C** and **Figure S1**) is composed of 27 Uruguayan sequences and five Brazilian ones and is part of a B.1.1.28 clade characterized by the amino acid change P13L in the Nucleocapsid viral protein that is widely distributed in southern Brazil and already detected circulating in Rocha department by the end of 2020 [7,45,46].

We next performed a time-scaled phylogeographic analysis of all lineage P.6 Uruguayan sequences produced in this work (n=163) and six Brazilian basal sequences. The spatiotemporal reconstruction suggests that lineage P.6 probably arose around November 9, 2020 (HPD: October 20, 2020 to November 26, 2020) in Montevideo, the capital department of Uruguay (**Figure 2A**). Lineage P.6 spread from Montevideo to other Uruguayan departments, with evidence of local transmission clusters in Rocha, Salto, San Jose and Tacuarembo. The T_MRCA_ of local transmission clusters in Rocha and Salto was traced to December 23, 2020 (HPD: December 12, 2020 to December 30, 2020) and December 30, 2020 (HPD: December 19, 2020 to January 09, 2021), respectively (**Figure 2A-B**). The introduction and dispersion of this lineage in each department coincided with the increase in new COVID-19 cases reported daily (**Figure 2A**).

**Figure 2.**
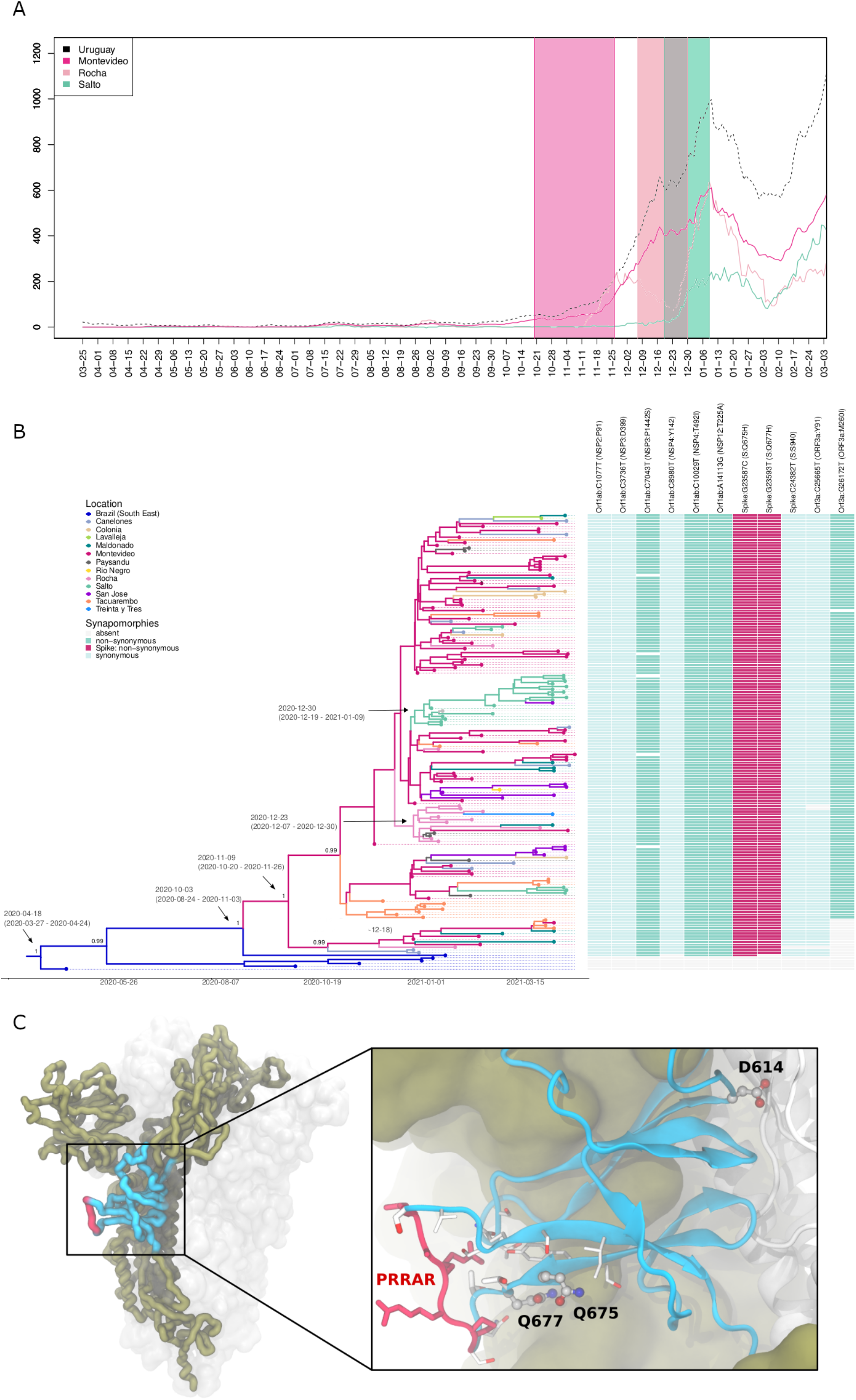
**A**. Number of daily new cases from March, 2020 to May, 2021 in the country (black), in Montevideo (fuchsia), Rocha (pink), Salto (green) and in Tacuarembó (orange). Daily new cases for Rocha, Salto and Tacuarembó were multiplied by a factor proportional to the population of that department in comparison to Montevideo (times 24,13 and 16 for Rocha, Salto and Tacuarembó, respectively) for visualization purposes. Confidence intervals of TMRCA of Montevideo (fuchsia), Rocha (pink), Salto (green) and Tacuarembó (orange) clades are shown as shaded areas. **B**. Bayesian phylogeographic analysis of P.6 clade in Uruguay, implemented in BEAST. Uruguayan sequences generated by the IiWG (n=163) were combined with six additional basal sequences from southeastern Brazil. Tips and branches of the time-scaled Bayesian tree are colored according to sampling location and the most probable location state of their descendent nodes, respectively, as indicated in the legend. Posterior probability support values and estimated TMRCAs are indicated at key nodes. Additionally, a heatmap represents the presence or absence of synapomorphic sites. The color scheme indicates the different mutations, as indicated in the legend. In each case, genomic position, nucleotide substitution, viral protein and amino acid is shown. **C**. One Spike protomer of the homotrimer is shown in thick ribbons, while the others are represented as transparent solvent accessible surfaces. The subdomain SD2 (residues 590-700) is indicated in blue, and the polybasic furin cleavage site (PRRAR, residues 681-685) is in red. The inset shows a zoom into the structural context of SD2, representing residues D614, Q675 and Q677 in balls and sticks, and nearby residues in sticks.

This new lineage P.6 spreading in Uruguay is characterized by 10 lineage-defining genetic changes, including five non-synonymous mutations, two of them in the Spike viral protein (**Figure 2B**). The amino acid change S:Q677H has been reported as a recurrent mutation arising independently in many SARS-CoV-2 lineages circulating worldwide by the end of 2020 [8,9]. In fact, a search in the EpiCoV database (accessed on July 7, 2021) reported 85 non-Uruguayan samples with high quality SARS-CoV-2 genome sequences with both Q675H and Q677H present. The pair Q675H+Q677H appeared distributed in 12 different countries (in descendent frequency order: Uruguay, England, USA, Belgium, India, Australia, Switzerland, Spain, Netherlands, Japan, Germany and France) and classified as 13 different pango lineages (in descendent frequency order: B.1.1.28, B.1.36, B.1.2, C.36, B.1.538, B.1.1.316, B.1.526, B.1.525, B.1.243, B.1.1.70, B.1.1.7, B.1.1.63 and B.1) (**Figure S3)**. Structural analysis of the SARS-CoV-2 Spike glycoprotein shows that residues Q675H and Q677H are within the subdomain SD2 of each protomer constituting the homotrimer (**Figure 2C**). The non-synonymous mutations Q675H and Q677H lay at the beginning of a very flexible loop (residues 675-690) [47], which embraces the solvent accessible polybasic furin cleavage site (**Figure 2C**, inset). These mutations are close to two experimentally observed O-glycosylation sites at T676 and T678 [48,49] and at the same domain of D614G mutation [50]. Particularly, D614G mutation is known to impact structural and thermodynamic aspects of the spike [51–54], and to enhance the protease cleavage [55], likely by allosterically increasing the binding to furin [56]. Recently, the increase in protease-mediated cleavage and the promotion of syncytium formation and virus infectivity has been shown for G614 viruses [57]. Knowing that histidine residues function as pH-sensors in other viruses [58], Q675H and Q677H mutations might also provide some synergic structural changes in the dynamics of the subdomain SD2. All in all, both Q675H and Q677H amino acid changes might trigger some effects on viral transmissibility by enhancing D614G effects.

## Discussion

Since March 2020, Uruguay had been successful at keeping the COVID-19 pandemic at check. Closed international borders and an aggressive contact-tracing system, among other government measures, were able to avoid virus transmission growing exponentially [1,2]. A few outbreaks occurred in departments bordering Brazil or in the southern departments, but related to those happening close to the Brazilian frontier [5]. Brazil has been a COVID-19 hotspot in South America and the 1,068 km long Uruguayan-Brazilian dry border allowed the rapid local establishment of SARS-CoV-2 Brazilian lineages B.1.1.28 and B.1.1.33. However, by the end of 2020, the pandemics worsened with a clear increase of daily cases in December. Summer-related social gatherings and relaxed vigilance are some of the proposed reasons to explain the epidemic growth that led to the loss of the TETRIS strategy [2]. However, we suggest that the emergence and spread of a B.1.1.28-derived lineage might have been involved.

In this study, we described a new B.1.1.28 sub-lineage designated P.6 that arose in Montevideo by November 2020 and spread throughout the country. Lineage P.6 comprises most Uruguayan virus genomes sequenced in the late 2020 and early 2021 and its emergence and local spread coincided with the national and local (as shown for Rocha and Salto departments) increase in daily SARS-CoV-2 cases. Lineage dispersion and prevalence changes can occur purely by chance and human behaviour, as was demonstrated for variant 20E(EU1) which emerged in Spain and spread through Europe in the boreal summer of 2020 [59]. While the human behaviour in Uruguay could have changed by the end of the austral spring and there was an ambiance of relaxed restrictions, we consider that the amino acid changes Q675H and Q677H in the Spike protein might alter SARS-CoV-2 fitness, contributing to the rapid increase in the lineage dominance observed between December 2020 and February 2021.

We have not conducted any experimental study on the effect on the viral fitness produced by these Q675H and Q677H Spike mutations and we are not aware of any experimental assay that assessed them. However, independent data might indicate that these amino acid changes (either one or both of them) could facilitate viral transmissibility. First, as explained above, they are in close proximity to the polybasic cleavage site at the S1/S2 boundary and the mutations are consistent with its potential functional relevance during cell entry. Second, a recent study that developed an innovative model on epidemiological variables integrating the effect of Spike amino acid changes in viral fitness, forecasted that both Q675H and Q677H could appear in emerging SARS-CoV-2 VOCs in the following months [60]. Third, and most importantly, convergent evolution is a hallmark of positive selection, and we identified the independent appearance of both Q675H and Q677H in 12 additional SARS-CoV-2 lineages. Moreover, Q677H alone has also been identified before as a recurrently appearing mutation in different lineages [8,9].

In summary, this study describes the emergence and local spread of lineage P.6, a new B.1.1.28-derived lineage carrying Spike mutations Q675H+Q677H, in Uruguay that coincided with the first exponential growth phase of the COVID-19 epidemic in the country that started by November, 2020 and lasted until mid-February 2021. The ancestral B.1.1.28 virus carrying mutation Q675H was probably introduced from southeastern Brazil into Montevideo, Uruguay’s capital city, by November 2020 and this virus rapidly fixed mutation Q677H and spread across the whole country. We propose that lineage P.6 has some altered biological properties which contributed to its swift dissemination in Uruguay and to the first COVID-19 wave that the country suffered by the end of 2020 and beginning of 2021. Although the lineage P.6 was rapidly substituted by the VOC P.1 as the most prevalent lineage in Uruguay since April 2021 [6], the monitoring of the concurrent emergence of Spike mutations Q675H+Q677H in VOIs and/or VOCs should be of worldwide interest.

## Data Availability

All SARS-CoV-2 genome sequences have been submitted to the EpiCoV/GISAID database with accession numbers indicated in Table S1.

## Acknowledgments

The National Ministry of Health (Uruguay) is the main health Institution in our country. It is a dedicated ethics oversight body and has granted us the ethical approval for this work. All necessary patient/participant consent has been obtained and the appropriate institutional forms have been archived.

The authors wish to thank all the health care workers and scientists who have worked hard to deal with this pandemic threat, the GISAID team, and all the EpiCoV database’s submitters (GISAID acknowledgment table containing the sequences used in this study is in supplementary **Table S3**). This work was supported by FOCEM-Fondo para la Convergencia Estructural del Mercosur (COF03/11).

We thank Christian Brandt from Institute for Infectious Diseases and Infection Control (Jena University Hospital), for adapting their poreCov Nextflow pipeline to our requirements and quickly fixing reported bugs.

## Data availability

All SARS-CoV-2 genome sequences have been submitted to the EpiCoV/GISAID database with accession numbers indicated in **Table S1**.

**Figure S1**. Maximum likelihood tree of 1,792 B.1.1.28 sequences found in Brazil and Uruguay. Additionally, four sequences from the USA, Spain and Belgium were included. The root of the tree was established using the earliest sequence from Brazil, with collection date 2020-03-05. The Uruguayan P.6 clade and Brazilian basal sequences are highlighted in purple and pink, respectively. Besides, taxon names for samples collected in the USA and Spain are shown in blue. Remaining Uruguayan sequences through the ML tree are indicated by a green taxon name, and supported clades are highlighted in green.

**Figure S2**. Schematic representation of migration events during dissemination of SARS-CoV-2 lineage B.1.1.28 between Uruguay and Brazil. The migration events were inferred by ancestral character reconstruction obtained through a maximum likelihood method implemented in PastML. Each node in the network is identified by location and number of sequences within different phylogenetic subclusters. Arrows indicate migration events deduced from location state changes across the B.1.1.28 ML tree. The shade of gray identifies marginal probabilities and the numbers quantify the migration events connecting respective locations (no numbers represent one single event). Nodes are colored according to their location. BR: Brazil.

**Figure S3**. Geographical and pango lineage distribution of worldwide SARS-CoV-2 genome sequences carrying mutations Q675H+Q677H. Barplots indicate the number of observed cases per country and lineage. The analysis is based on the 163 B.1.1.28+Q675H+Q677H sequences from this study and additional 129 complete and high quality genomes, collected worldwide and carrying Q675H+Q677H independently of the assigned lineage. These sequences were obtained from GISAID on July 7, 2021. B.1.1.28 corresponds to the newly designated P.6 pango lineage.

**Table S1**. Information on the B.1.1.28 samples from Uruguay used in this study. # indicate 11 samples already analyzed in Rego et al. [7], ^ show six samples here sequenced although they were not collected in our target period (November 2020 - April 2021) and * shows the samples processed using the “midnight” primer set, using Nanopore Rapid kit.

**Table S2**. Information about Uruguayan, Brazilian and worldwide samples obtained from GISAID (accessed on July 7, 2021).

**Table S3**. GISAID Acknowledgements Table.

## Notes

### Competing Interest Statement

The authors have declared no competing interest.

### Author Declarations

This work was done by the Inter-Institutional Working Group (IiWG) for SARS-CoV-2 genomic surveillance in Uruguay, which involves a diagnostic network, expertise and resources to handle large-scale sequencing, computational scientists for genomic analysis, and an affordable and decentralized in house qPCR test designed to detect known VOCs [6]. Residual de-identified RNA samples from SARS-CoV-2 positive patients were remitted to the Institut Pasteur de Montevideo (IPMon). IPMon was validated by the Ministry of Health of Uruguay as an approved center providing diagnostic testing for COVID-19. All samples were de-identified before receipt by the study investigators. All relevant ethical guidelines have been appropriately followed. Additionally, the project was approved by the Ethics Committee of the Sanatorio Americano SASA (Uruguay). Ethical approval was given and signed informed consent was obtained from the participants.

## References

1. Estadisticasuy Available online: https://guiad-covid.github.io/estadisticasuy.html (accessed on 26 July 2021).

2. Taylor, L. Why Uruguay Lost Control of COVID. Nature 2021, 595, 21–21, doi:10.1038/d41586-021-01714-4.

3. Fraser, C.; Riley, S.; Anderson, R.M.; Ferguson, N.M. Factors That Make an Infectious Disease Outbreak Controllable. Proc. Natl. Acad. Sci. 2004, 101, 6146–6151, doi:10.1073/pnas.0307506101.

4. Grantz, K.H.; Lee, E.C.; McGowan, L.D.; Lee, K.H.; Metcalf, C.J.E.; Gurley, E.S.; Lessler, J. Maximizing and Evaluating the Impact of Test-Trace-Isolate Programs: A Modeling Study. PLOS Med. 2021, 18, e1003585, doi:10.1371/journal.pmed.1003585.

5. Mir, D.; Rego, N.; Resende, P.C.; Tort, F.; Fernández-Calero, T.; Noya, V.; Brandes, M.; Possi, T.; Arleo, M.; Reyes, N.; et al. Recurrent Dissemination of SARS-CoV-2 Through the Uruguayan–Brazilian Border. Front. Microbiol. 2021, 0, doi:10.3389/fmicb.2021.653986.

6. Rego, N.; Costábile, A.; Paz, M.; Salazar, C.; Perbolianachis, P.; Spangenberg, L.; Ferrés, I.; Arce, R.; Fajardo, A.; Arleo, M.; et al. Implementation of a QPCR Assay Coupled with Genomic Surveillance for Real-Time Monitoring of SARS-CoV-2 Variants of Concern; Infectious Diseases (except HIV/AIDS), 2021;

7. Rego, N.; Fernández-Calero, T.; Arantes, I.; Noya, V.; Mir, D.; Brandes, M.; Zanetti, J.; Arleo, M.; Pereira, E.; Possi, T.; et al. Spatiotemporal Dissemination Pattern of SARS-CoV-2 B1.1.28-Derived Lineages Introduced into Uruguay across Its Southeastern Border with Brazil. medRxiv 2021, 2021.07.05.21259760, doi:10.1101/2021.07.05.21259760.

8. Detection of the Recurrent Substitution Q677H in the Spike Protein of SARS-CoV-2 in Cases Descended from the Lineage B.1.429 - SARS-CoV-2 Coronavirus / NCoV-2019 Genomic Epidemiology Available online: https://virological.org/t/detection-of-the-recurrent-substitution-q677h-in-the-spike-protein-of-sars-cov-2-in-cases-descended-from-the-lineage-b-1-429/660 (accessed on 22 July 2021).

9. Hodcroft, E.B.; Domman, D.B.; Snyder, D.J.; Oguntuyo, K.Y.; Diest, M.V.; Densmore, K.H.; Schwalm, K.C.; Femling, J.; Carroll, J.L.; Scott, R.S.; et al. Emergence in Late 2020 of Multiple Lineages of SARS-CoV-2 Spike Protein Variants Affecting Amino Acid Position 677. medRxiv 2021, 2021.02.12.21251658, doi:10.1101/2021.02.12.21251658.

10. Quick, J. NCoV-2019 Sequencing Protocol v3 (LoCost). 2020.

11. Tyson, J.R.; James, P.; Stoddart, D.; Sparks, N.; Wickenhagen, A.; Hall, G.; Choi, J.H.; Lapointe, H.; Kamelian, K.; Smith, A.D.; et al. Improvements to the ARTIC Multiplex PCR Method for SARS-CoV-2 Genome Sequencing Using Nanopore; Genomics, 2020;

12. Freed, N.; Silander, O. SARS-CoV2 Genome Sequencing Protocol (1200bp Amplicon &#34;Midnight&#34; Primer Set, Using Nanopore Rapid Kit) V5.

13. Freed, N.E.; Vlková, M.; Faisal, M.B.; Silander, O.K. Rapid and Inexpensive Whole-Genome Sequencing of SARS-CoV-2 Using 1200 Bp Tiled Amplicons and Oxford Nanopore Rapid Barcoding. Biol. Methods Protoc. 2020, 5, bpaa014, doi:10.1093/biomethods/bpaa014.

14. Oxford Nanopore Technologies Available online: https://nanoporetech.com/ (accessed on 2 June 2021).

15. GitHub - Replikation/PoreCov: SARS-CoV-2 Workflow for Nanopore Sequence Data Available online: https://github.com/replikation/poreCov (accessed on 23 July 2021).

16. Hufsky, F.; Lamkiewicz, K.; Almeida, A.; Aouacheria, A.; Arighi, C.; Bateman, A.; Baumbach, J.; Beerenwinkel, N.; Brandt, C.; Cacciabue, M.; et al. Computational Strategies to Combat COVID-19: Useful Tools to Accelerate SARS-CoV-2 and Coronavirus Research. Brief. Bioinform. 2021, 22, 642–663, doi:10.1093/bib/bbaa232.

17. Brandt, C.; Krautwurst, S.; Spott, R.; Lohde, M.; Jundzill, M.; Marquet, M.; Hölzer, M. PoreCov - an Easy to Use, Fast, and Robust Workflow for SARS-CoV-2 Genome Reconstruction via Nanopore Sequencing. bioRxiv 2021, 2021.05.07.443089, doi:10.1101/2021.05.07.443089.

18. Wood, D.E.; Lu, J.; Langmead, B. Improved Metagenomic Analysis with Kraken 2. Genome Biol. 2019, 20, 257, doi:10.1186/s13059-019-1891-0.

19. Loman, N.J.; Quick, J.; Simpson, J.T. A Complete Bacterial Genome Assembled de Novo Using Only Nanopore Sequencing Data. Nat. Methods 2015, 12, 733–735, doi:10.1038/nmeth.3444.

20. Di Tommaso, P.; Chatzou, M.; Floden, E.W.; Barja, P.P.; Palumbo, E.; Notredame, C. Nextflow Enables Reproducible Computational Workflows. Nat. Biotechnol. 2017, 35, 316–319, doi:10.1038/nbt.3820.

21. Kurtzer, G.M.; Sochat, V.; Bauer, M.W. Singularity: Scientific Containers for Mobility of Compute. PLOS ONE 2017, 12, e0177459, doi:10.1371/journal.pone.0177459.

22. Ferguson, J.M.; Gamaarachchi, H.; Nguyen, T.; Gollon, A.; Tong, S.; Aquilina-Reid, C.; Bowen-James, R.; Deveson, I.W. InterARTIC: An Interactive Web Application for Whole-Genome Nanopore Sequencing Analysis of SARS-CoV-2 and Other Viruses. bioRxiv 2021, 2021.04.21.440861, doi:10.1101/2021.04.21.440861.

23. Li, H. Minimap2: Pairwise Alignment for Nucleotide Sequences. Bioinformatics 2018, 34, 3094–3100, doi:10.1093/bioinformatics/bty191.

24. Ondov, B.D.; Bergman, N.H.; Phillippy, A.M. Interactive Metagenomic Visualization in a Web Browser. BMC Bioinformatics 2011, 12, 385, doi:10.1186/1471-2105-12-385.

25. Rambaut, A.; Holmes, E.C.; O’Toole, Á.; Hill, V.; McCrone, J.T.; Ruis, C.; du Plessis, L.; Pybus, O.G. A Dynamic Nomenclature Proposal for SARS-CoV-2 Lineages to Assist Genomic Epidemiology. Nat. Microbiol. 2020, 5, 1403–1407, doi:10.1038/s41564-020-0770-5.

26. COG-UK Available online: https://pangolin.cog-uk.io/ (accessed on 26 July 2021).

27. Shu, Y.; McCauley, J. GISAID: Global Initiative on Sharing All Influenza Data – from Vision to Reality. Eurosurveillance 2017, 22, 30494, doi:10.2807/1560-7917.ES.2017.22.13.30494.

28. Katoh, K.; Standley, D.M. MAFFT Multiple Sequence Alignment Software Version 7: Improvements in Performance and Usability. Mol. Biol. Evol. 2013, 30, 772–780, doi:10.1093/molbev/mst010.

29. Nguyen, L.-T.; Schmidt, H.A.; von Haeseler, A.; Minh, B.Q. IQ-TREE: A Fast and Effective Stochastic Algorithm for Estimating Maximum-Likelihood Phylogenies. Mol. Biol. Evol. 2015, 32, 268–274, doi:10.1093/molbev/msu300.

30. Anisimova, M.; Gascuel, O. Approximate Likelihood-Ratio Test for Branches: A Fast, Accurate, and Powerful Alternative. Syst. Biol. 2006, 55, 539–552, doi:10.1080/10635150600755453.

31. Sagulenko, P.; Puller, V.; Neher, R.A. TreeTime: Maximum-Likelihood Phylodynamic Analysis. Virus Evol. 2018, 4, doi:10.1093/ve/vex042.

32. Duchene, S.; Featherstone, L.; Haritopoulou-Sinanidou, M.; Rambaut, A.; Lemey, P.; Baele, G. Temporal Signal and the Phylodynamic Threshold of SARS-CoV-2. Virus Evol. 2020, 6, doi:10.1093/ve/veaa061.

33. Time Dependence of SARS-CoV-2 Substitution Rates - SARS-CoV-2 Coronavirus / NCoV-2019 Evolutionary History Available online: https://virological.org/t/time-dependence-of-sars-cov-2-substitution-rates/542 (accessed on 23 July 2021).

34. Ishikawa, S.A.; Zhukova, A.; Iwasaki, W.; Gascuel, O. A Fast Likelihood Method to Reconstruct and Visualize Ancestral Scenarios. Mol. Biol. Evol. 2019, 36, 2069–2085, doi:10.1093/molbev/msz131.

35. Suchard, M.A.; Lemey, P.; Baele, G.; Ayres, D.L.; Drummond, A.J.; Rambaut, A. Bayesian Phylogenetic and Phylodynamic Data Integration Using BEAST 1.10. Virus Evol. 2018, 4, doi:10.1093/ve/vey016.

36. Drummond, A.J.; Rambaut, A.; Shapiro, B.; Pybus, O.G. Bayesian Coalescent Inference of Past Population Dynamics from Molecular Sequences. Mol. Biol. Evol. 2005, 22, 1185–1192, doi:10.1093/molbev/msi103.

37. Lemey, P.; Rambaut, A.; Drummond, A.J.; Suchard, M.A. Bayesian Phylogeography Finds Its Roots. PLoS Comput. Biol. 2009, 5, e1000520, doi:10.1371/journal.pcbi.1000520.

38. Ferreira, M.A.R.; Suchard, M.A. Bayesian Analysis of Elapsed Times in Continuous-Time Markov Chains. Can. J. Stat. 2008, 36, 355–368, doi:10.1002/cjs.5550360302.

39. Rambaut, A.; Drummond, A.J.; Xie, D.; Baele, G.; Suchard, M.A. Posterior Summarization in Bayesian Phylogenetics Using Tracer 1.7. Syst. Biol. 2018, 67, 901– 904, doi:10.1093/sysbio/syy032.

40. Bouckaert, R.; Vaughan, T.G.; Barido-Sottani, J.; Duchêne, S.; Fourment, M.; Gavryushkina, A.; Heled, J.; Jones, G.; Kühnert, D.; Maio, N.D.; et al. BEAST 2.5: An Advanced Software Platform for Bayesian Evolutionary Analysis. PLOS Comput. Biol. 2019, 15, e1006650, doi:10.1371/journal.pcbi.1006650.

41. FigTree Available online: http://tree.bio.ed.ac.uk/software/figtree/ (accessed on 26 July 2021).

42. Yu, G.; Smith, D.K.; Zhu, H.; Guan, Y.; Lam, T.T.-Y. Ggtree: An r Package for Visualization and Annotation of Phylogenetic Trees with Their Covariates and Other Associated Data. Methods Ecol. Evol. 2017, 8, 28–36, doi:10.1111/2041-210X.12628.

43. D. E. Shaw Research Technical Data Molecular Dynamics Simulations Related to SARS-CoV-2 Available online: https://www.deshawresearch.com/downloads/download_trajectory_sarscov2.cgi/ (accessed on 26 July 2021).

44. Humphrey, W.; Dalke, A.; Schulten, K. VMD: Visual Molecular Dynamics. J. Mol. Graph. 1996, 14, 33–38, doi:10.1016/0263-7855(96)00018-5.

45. Francisco Jr, R. da S.; Benites, L.F.; Lamarca, A.P.; de Almeida, L.G.P.; Hansen, A.W.; Gularte, J.S.; Demoliner, M.; Gerber, A.L.; de C Guimarães, A.P.; Antunes, A.K.E.; et al. Pervasive Transmission of E484K and Emergence of VUI-NP13L with Evidence of SARS-CoV-2 Co-Infection Events by Two Different Lineages in Rio Grande Do Sul, Brazil. Virus Res. 2021, 296, 198345, doi:10.1016/j.virusres.2021.198345.

46. Sant’Anna, F.H.; Varela, A.P.M.; Prichula, J.; Comerlato, J.; Comerlato, C.B.; Roglio, V.S.; Pereira, G.F.M.; Moreno, F.; Seixas, A.; Wendland, E.M. Emergence of the Novel SARS-CoV-2 Lineage VUI-NP13L and Massive Spread of P.2 in South Brazil. Emerg. Microbes Infect. 2021, 10, 1431–1440, doi:10.1080/22221751.2021.1949948.

47. Lemmin, T.; Kalbermatter, D.; Harder, D.; Plattet, P.; Fotiadis, D. Structures and Dynamics of the Novel S1/S2 Protease Cleavage Site Loop of the SARS-CoV-2 Spike Glycoprotein. J. Struct. Biol. X 2020, 4, 100038, doi:10.1016/j.yjsbx.2020.100038.

48. Bagdonaite, I.; Thompson, A.J.; Wang, X.; Søgaard, M.; Fougeroux, C.; Frank, M.; Diedrich, J.K.; Yates, J.R.; Salanti, A.; Vakhrushev, S.Y.; et al. Site-Specific O-Glycosylation Analysis of SARS-CoV-2 Spike Protein Produced in Insect and Human Cells. Viruses 2021, 13, 551, doi:10.3390/v13040551.

49. Sanda, M.; Morrison, L.; Goldman, R. N- and O-Glycosylation of the SARS-CoV-2 Spike Protein. Anal. Chem. 2021, 93, 2003–2009, doi:10.1021/acs.analchem.0c03173.

50. Gobeil, S.M.-C.; Janowska, K.; McDowell, S.; Mansouri, K.; Parks, R.; Stalls, V.; Kopp, M.F.; Manne, K.; Li, D.; Wiehe, K.; et al. Effect of Natural Mutations of SARS-CoV-2 on Spike Structure, Conformation, and Antigenicity. Science 2021, doi:10.1126/science.abi6226.

51. Zhang, J.; Cai, Y.; Xiao, T.; Lu, J.; Peng, H.; Sterling, S.M.; Walsh, R.M.; Rits-Volloch, S.; Zhu, H.; Woosley, A.N.; et al. Structural Impact on SARS-CoV-2 Spike Protein by D614G Substitution. Science 2021, 372, 525–530, doi:10.1126/science.abf2303.

52. Mansbach, R.A.; Chakraborty, S.; Nguyen, K.; Montefiori, D.C.; Korber, B.; Gnanakaran, S. The SARS-CoV-2 Spike Variant D614G Favors an Open Conformational State. Sci. Adv. 2021, 7, eabf3671, doi:10.1126/sciadv.abf3671.

53. Yurkovetskiy, L.; Wang, X.; Pascal, K.E.; Tomkins-Tinch, C.; Nyalile, T.P.; Wang, Y.; Baum, A.; Diehl, W.E.; Dauphin, A.; Carbone, C.; et al. Structural and Functional Analysis of the D614G SARS-CoV-2 Spike Protein Variant. Cell 2020, 183, 739–751.e8, doi:10.1016/j.cell.2020.09.032.

54. Benton, D.J.; Wrobel, A.G.; Roustan, C.; Borg, A.; Xu, P.; Martin, S.R.; Rosenthal, P.B.; Skehel, J.J.; Gamblin, S.J. The Effect of the D614G Substitution on the Structure of the Spike Glycoprotein of SARS-CoV-2. Proc. Natl. Acad. Sci. 2021, 118, doi:10.1073/pnas.2022586118.

55. Gobeil, S.M.-C.; Janowska, K.; McDowell, S.; Mansouri, K.; Parks, R.; Manne, K.; Stalls, V.; Kopp, M.F.; Henderson, R.; Edwards, R.J.; et al. D614G Mutation Alters SARS-CoV-2 Spike Conformation and Enhances Protease Cleavage at the S1/S2 Junction. Cell Rep. 2021, 34, 108630, doi:10.1016/j.celrep.2020.108630.

56. Mohammad, A.; Alshawaf, E.; Marafie, S.K.; Abu-Farha, M.; Abubaker, J.; Al-Mulla, F. Higher Binding Affinity of Furin for SARS-CoV-2 Spike (S) Protein D614G Mutant Could Be Associated with Higher SARS-CoV-2 Infectivity. Int. J. Infect. Dis. 2021, 103, 611– 616, doi:10.1016/j.ijid.2020.10.033.

57. Cheng, Y.-W.; Chao, T.-L.; Li, C.-L.; Wang, S.-H.; Kao, H.-C.; Tsai, Y.-M.; Wang, H.-Y.; Hsieh, C.-L.; Lin, Y.-Y.; Chen, P.-J.; et al. D614G Substitution of SARS-CoV-2 Spike Protein Increases Syncytium Formation and Virus Titer via Enhanced Furin-Mediated Spike Cleavage. mBio 0, e00587–21, doi:10.1128/mBio.00587-21.

58. Kampmann, T.; Mueller, D.S.; Mark, A.E.; Young, P.R.; Kobe, B. The Role of Histidine Residues in Low-PH-Mediated Viral Membrane Fusion. Structure 2006, 14, 1481–1487, doi:10.1016/j.str.2006.07.011.

59. Hodcroft, E.B.; Zuber, M.; Nadeau, S.; Vaughan, T.G.; Crawford, K.H.D.; Althaus, C.L.; Reichmuth, M.L.; Bowen, J.E.; Walls, A.C.; Corti, D.; et al. Spread of a SARS-CoV-2 Variant through Europe in the Summer of 2020. Nature 2021, 1–6, doi:10.1038/s41586-021-03677-y.

60. Maher, M.C.; Bartha, I.; Weaver, S.; Iulio, J. di Ferri, E.; Soriaga, L.; Lempp, F.A.; Hie, B.L.; Bryson, B.; Berger, B.; et al. Predicting the Mutational Drivers of Future SARS-CoV-2 Variants of Concern. medRxiv 2021, 2021.06.21.21259286, doi:10.1101/2021.06.21.21259286.

