## Supplementary figures and images for "Emergence and spread of a B.1.1.28-derived P.6 lineage with Q675H and Q677H Spike mutations in Uruguay"

### Figure S1

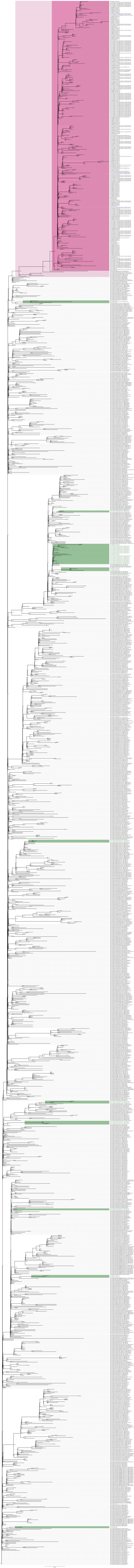

### Figure S2

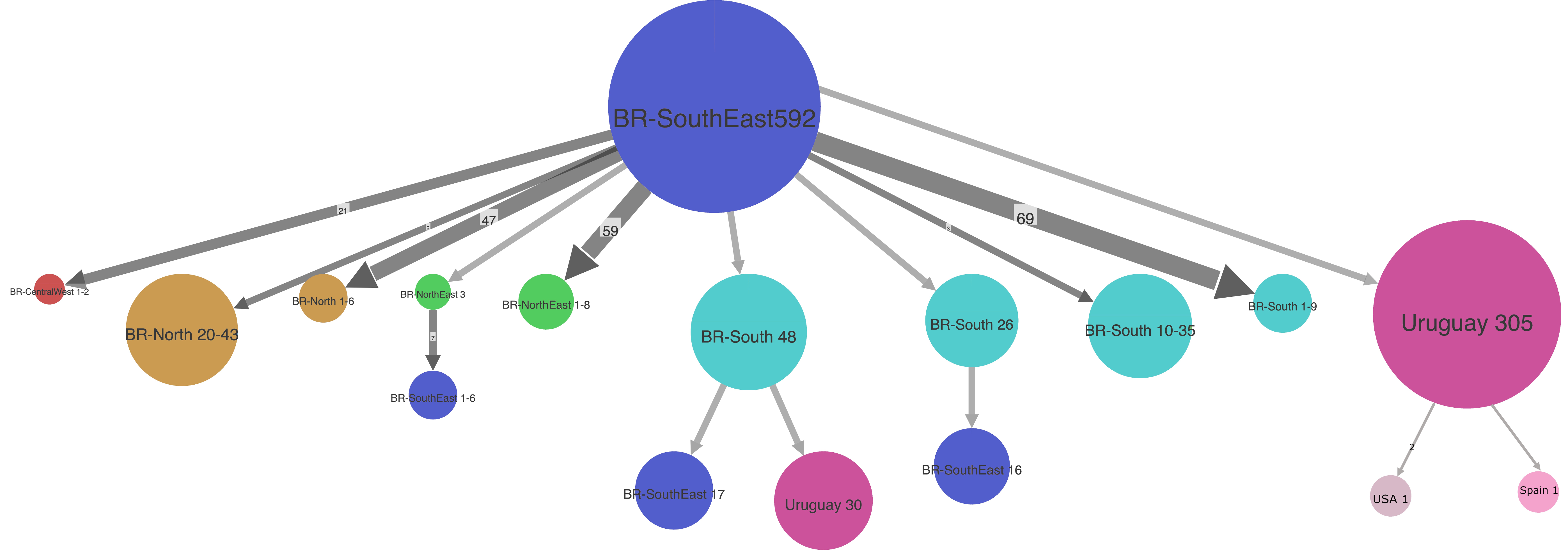

### Figure S3

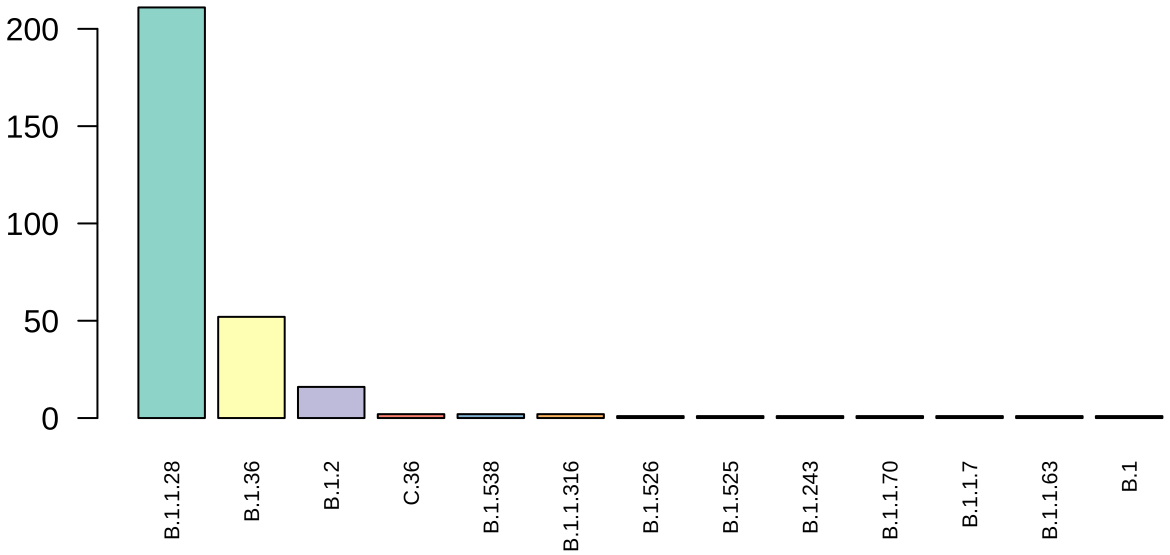

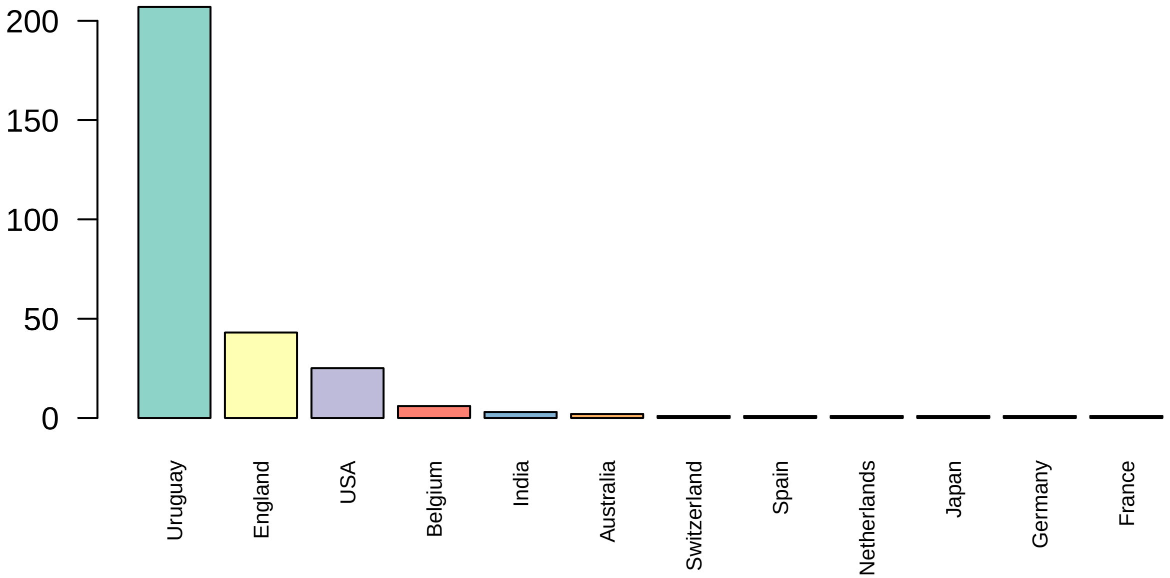
