## Supplementary material for "Emergence and spread of a B.1.1.28-derived P.6 lineage with Q675H and Q677H Spike mutations in Uruguay": Table S3

We gratefully acknowledge the following Authors from the Originating laboratories responsible for obtaining the specimens, as well as the Submitting laboratories where the genome data were generated and shared via GISAID, on which this research is based.

All Submitters of data may be contacted directly via www.gisaid.org

Authors are sorted alphabetically.

| Accession ID | Originating Laboratory | Submitting Laboratory | Authors |
|---|---|---|---|
| EPI_ISL_2427508, EPI_ISL_2427509, EPI_ISL_2427511, EPI_ISL_2427513, EPI_ISL_2427516, EPI_ISL_2427517, EPI_ISL_2427519, EPI_ISL_2427520, EPI_ISL_2427524, EPI_ISL_2427527, EPI_ISL_2427528, EPI_ISL_2427529, EPI_ISL_2427531, EPI_ISL_2427532, EPI_ISL_2427535, EPI_ISL_2427539, EPI_ISL_2427540, EPI_ISL_2427542, EPI_ISL_2427543, EPI_ISL_2427546, EPI_ISL_2427548, EPI_ISL_2427549, EPI_ISL_2427550, EPI_ISL_2427551, EPI_ISL_2427552, EPI_ISL_2427553, EPI_ISL_2427554, EPI_ISL_2427555, EPI_ISL_2427557, EPI_ISL_2427558, EPI_ISL_2427559, EPI_ISL_2427563, EPI_ISL_2427564, EPI_ISL_2427568, EPI_ISL_2427569, EPI_ISL_2427571, EPI_ISL_2427575, EPI_ISL_2427577, EPI_ISL_2427578, EPI_ISL_2427579, EPI_ISL_2427584, EPI_ISL_2427585, EPI_ISL_2427587, EPI_ISL_2427589, EPI_ISL_2427591, EPI_ISL_2427592, EPI_ISL_2427593, EPI_ISL_2427598, EPI_ISL_2427599, EPI_ISL_2427600, EPI_ISL_2427603, EPI_ISL_2427605, EPI_ISL_2427606, EPI_ISL_2427607, EPI_ISL_2427608, EPI_ISL_2427609, EPI_ISL_2427613, EPI_ISL_2427614, EPI_ISL_2427615, EPI_ISL_2427616, EPI_ISL_2427617, EPI_ISL_2427619, EPI_ISL_2427620, EPI_ISL_2427621, EPI_ISL_2427622, EPI_ISL_2427623, EPI_ISL_2427624, EPI_ISL_2427625, EPI_ISL_2427626, EPI_ISL_2427627, EPI_ISL_2427629, EPI_ISL_2427630, EPI_ISL_2427631, EPI_ISL_2427632, EPI_ISL_2427633, EPI_ISL_2427635, EPI_ISL_2427636, EPI_ISL_2427637, EPI_ISL_2427640, EPI_ISL_2427641, EPI_ISL_2427642, EPI_ISL_2427644, EPI_ISL_2427645, EPI_ISL_2427646, EPI_ISL_2427647, EPI_ISL_2427648, EPI_ISL_2427649, EPI_ISL_2427650, EPI_ISL_2427651, EPI_ISL_2427652, EPI_ISL_2427653, EPI_ISL_2427654, EPI_ISL_2427655, EPI_ISL_2427656, EPI_ISL_2427658, EPI_ISL_2427660, EPI_ISL_2427661, EPI_ISL_2427662, EPI_ISL_2427663, EPI_ISL_2427664, EPI_ISL_2427667, EPI_ISL_2427668, EPI_ISL_2427670, EPI_ISL_2427671, EPI_ISL_2427672, EPI_ISL_2427675, EPI_ISL_2427676, EPI_ISL_2427677, EPI_ISL_2427678, EPI_ISL_2427679, EPI_ISL_2427680, EPI_ISL_2427684, EPI_ISL_2427685, EPI_ISL_2427686, EPI_ISL_2427690, EPI_ISL_2427691, EPI_ISL_2427692, EPI_ISL_2427699, EPI_ISL_2427707, EPI_ISL_2427710, EPI_ISL_2427713, EPI_ISL_2427715, EPI_ISL_2427716, EPI_ISL_2427718, EPI_ISL_2427722, EPI_ISL_2427723, EPI_ISL_2427726, EPI_ISL_2427737, EPI_ISL_2427745, EPI_ISL_2427752, EPI_ISL_2427753, EPI_ISL_2427761, EPI_ISL_2427774, EPI_ISL_2427775 | | | |
| see above | Laboratorio de Biología Molecular Médica Uruguaya | Departments of Pathology and Medicine, New York University School of Medicine | Maria Victoria Elizondo, Maria Noel Zubillaga, Gonzalo Manrique, Cecilia Sorhouet, Maria Cristina Mogdasy, Paul Zappile, Dacia Dimartino, Christian Marier, Adriana Heguy |
| EPI_ISL_2753959, EPI_ISL_2753960, EPI_ISL_2753963, EPI_ISL_2753964, EPI_ISL_2754023, EPI_ISL_2754026, EPI_ISL_2754030, EPI_ISL_2754031, EPI_ISL_2754032, EPI_ISL_2754073, EPI_ISL_2754095, EPI_ISL_2754096, EPI_ISL_2754115, EPI_ISL_2754129, EPI_ISL_2754130, EPI_ISL_2754132, EPI_ISL_2754139, EPI_ISL_2754195, EPI_ISL_2754203, EPI_ISL_2754310, EPI_ISL_2754311 | | | |
| see above | Sanatorio Americano | Institut Pasteur de Montevideo | Natalia Rego,Tamara Fernández-Calero,Ighor Arantes,Verónica Noya,Daiana Mir,Mariana Brandes,Juan Zanetti,Mailen Arleo,Emiliano Pereira,Tania Possi,Odhille Chappos,Lucia Bilbao,Natalia Reyes,Melissa Duquía,Matías Victoria,Pía Techera,María José Benítez-Galeano,Luciana Griffero,Mauricio Méndez,Belén González,Pablo Smircich,Andres Lizasoain,Matías Castells,Matías Salvo,Rodney Colina,Cecilia Alonso,Gonzalo Bello,Lucía Spangenberg |
| EPI_ISL_2754313, EPI_ISL_2754314, EPI_ISL_2754315, EPI_ISL_2754316, EPI_ISL_2754317, EPI_ISL_2754318, EPI_ISL_2754370, EPI_ISL_2754398, EPI_ISL_2754608, EPI_ISL_2754628, EPI_ISL_2754806, EPI_ISL_2754960, EPI_ISL_2755057, EPI_ISL_2755733, EPI_ISL_2755857, EPI_ISL_2757850, EPI_ISL_2757851, EPI_ISL_2757855, EPI_ISL_2757856, EPI_ISL_2757857 | | | |
| see above | CURE | Institut Pasteur de Montevideo | Natalia Rego,Tamara Fernández-Calero,Ighor Arantes,Verónica Noya,Daiana Mir,Mariana Brandes,Juan Zanetti,Mailen Arleo,Emiliano Pereira,Tania Possi,Odhille Chappos,Lucia Bilbao,Natalia Reyes,Melissa Duquía,Matías Victoria,Pía Techera,María José Benítez-Galeano,Luciana Griffero,Mauricio Méndez,Belén González,Pablo Smircich,Andres Lizasoain,Matías Castells,Matías Salvo,Rodney Colina,Cecilia Alonso,Gonzalo Bello,Lucía Spangenberg |
| EPI_ISL_750175 | CENUR Este-Sede Rocha-UdelaR | Institut Pasteur de Montevideo | Daiana Mir, Natalia Rego, Paola Cristina Resende, Fernando Lopez-Tort, Tamara Fernandez-Calero, Veronica Noya, Mariana Brandes, Tania Possi, Mailen Arleo, Natalia Reyes, Matias Victoria, Andres Lizasoain, Matias Castells, Leticia Maya, Matías Salvo, Tatiana Schäffer Gregianini, Marilda Tereza Mar da Rosa, Letícia Garay Martins, Cecilia Alonso, Yasser Vega, Cecilia Salazar, Ignacio Ferrés, Pablo Smircich, Jose Sotelo, Ighor Arantes, Luciana Appolinario, Ana Carolina Mendonça, Maria Jose Benitez-Galeano, Martín Graña, Camila Simoes, Fernando Motta, Marilda Mendonça Siqueira, Gonzalo Bello, Rodney Colina, Lucia Spangenberg |
| EPI_ISL_750176, EPI_ISL_750177 | Sanatorio Americano | Institut Pasteur de Montevideo | Daiana Mir, Natalia Rego, Paola Cristina Resende, Fernando Lopez-Tort, Tamara Fernandez-Calero, Veronica Noya, Mariana Brandes, Tania Possi, Mailen Arleo, Natalia Reyes, Matias Victoria, Andres Lizasoain, Matias Castells, Leticia Maya, Matías Salvo, Tatiana Schäffer Gregianini, Marilda Tereza Mar da Rosa, Letícia Garay Martins, Cecilia Alonso, Yasser Vega, Cecilia Salazar, Ignacio Ferrés, Pablo Smircich, Jose Sotelo, Ighor Arantes, Luciana Appolinario, Ana Carolina Mendonça, Maria Jose Benitez-Galeano, Martín Graña, Camila Simoes, Fernando Motta, Marilda Mendonça Siqueira, Gonzalo Bello, Rodney Colina, Lucia Spangenberg |
| EPI_ISL_751184, EPI_ISL_751185, EPI_ISL_751186, EPI_ISL_751189, EPI_ISL_751190 | CENUR Litoral Norte - UdelaR, Salto, Uruguay | Institut Pasteur de Montevideo | Daiana Mir, Natalia Rego, Paola Cristina Resende, Fernando Lopez-Tort, Tamara Fernandez-Calero, Veronica Noya, Mariana Brandes, Tania Possi, Mailen Arleo, Natalia Reyes, Matias Victoria, Andres Lizasoain, Matias Castells, Leticia Maya, Matías Salvo, Tatiana Schäffer Gregianini, Marilda Tereza Mar da Rosa, Letícia Garay Martins, Cecilia Alonso, Yasser Vega, Cecilia Salazar, Ignacio Ferrés, Pablo Smircich, Jose Sotelo, Ighor Arantes, Luciana Appolinario, Ana Carolina Mendonça, Maria Jose Benitez-Galeano, Martín Graña, Camila Simoes, Fernando Motta, Marilda Mendonça Siqueira, Gonzalo Bello, Rodney Colina, Lucia Spangenberg |
| EPI_ISL_751201 | Laboratorio DILAVE/MGAP-INIA-UdelaR -Tacuarembó | Institut Pasteur de Montevideo | Daiana Mir, Natalia Rego, Paola Cristina Resende, Fernando Lopez-Tort, Tamara Fernandez-Calero, Veronica Noya, Mariana Brandes, Tania Possi, Mailen Arleo, Natalia Reyes, Matias Victoria, Andres Lizasoain, Matias Castells, Leticia Maya, Matías Salvo, Tatiana Schäffer Gregianini, Marilda Tereza Mar da Rosa, Letícia Garay Martins, Cecilia Alonso, Yasser Vega, Cecilia Salazar, Ignacio Ferrés, Pablo Smircich, Jose Sotelo, Ighor Arantes, Luciana Appolinario, Ana Carolina Mendonça, Maria Jose Benitez-Galeano, Martín Graña, Camila Simoes, Fernando Motta, Marilda Mendonça Siqueira, Gonzalo Bello, Rodney Colina, Lucia Spangenberg |

We gratefully acknowledge the following Authors from the Originating laboratories responsible for obtaining the specimens, as well as the Submitting laboratories where the genome data were generated and shared via GISAID, on which this research is based.

All Submitters of data may be contacted directly via www.gisaid.org

Authors are sorted alphabetically.

| Accession ID | Originating Laboratory | Submitting Laboratory | Authors |
|---|---|---|---|
| EPI_ISL_1000670 | Instituto de Biotecnologia - UNESP-Botucatu-SP | Instituto de Biotecnologia - UNESP-Botucatu-SP | Leila Sabrina Ullmann; Fábio Sossai Possebon, Camila Dantas Malossi, Paula Rahal, Paulo Inacio da Costa, João Pessoa Araújo Jr. |
| EPI_ISL_1039696 | Instituto Adolfo Lutz - Regional de Presidente Prudente | Instituto Adolfo Lutz, Interdiciplinary Procedures Center, Strategic Laboratory | Claudio Tavares Sacchi, Claudia Regina Gonçalves, Erica Valessa Ramos Gomes, Karoline Rodrigues Campos |
| EPI_ISL_1039697 | Instituto Adolfo Lutz Central | Instituto Adolfo Lutz, Interdiciplinary Procedures Center, Strategic Laboratory | Claudio Tavares Sacchi, Claudia Regina Gonçalves, Erica Valessa Ramos Gomes, Karoline Rodrigues Campos |
| EPI_ISL_1039698 | Lab Loc - Itapecerica da Serra | Instituto Adolfo Lutz, Interdiciplinary Procedures Center, Strategic Laboratory | Claudio Tavares Sacchi, Claudia Regina Gonçalves, Erica Valessa Ramos Gomes, Karoline Rodrigues Campos |
| EPI_ISL_1039699 | Instituto Adolfo Lutz - Regional de Taubate | Instituto Adolfo Lutz, Interdiciplinary Procedures Center, Strategic Laboratory | Claudio Tavares Sacchi, Claudia Regina Gonçalves, Erica Valessa Ramos Gomes, Karoline Rodrigues Campos |
| EPI_ISL_1039701 | Instituto Adolfo Lutz Central | Instituto Adolfo Lutz, Interdiciplinary Procedures Center, Strategic Laboratory | Claudio Tavares Sacchi, Claudia Regina Gonçalves, Erica Valessa Ramos Gomes, Karoline Rodrigues Campos |
| EPI_ISL_1039702 | Instituto Adolfo Lutz - Regional de Aracatuba | Instituto Adolfo Lutz, Interdiciplinary Procedures Center, Strategic Laboratory | Claudio Tavares Sacchi, Claudia Regina Gonçalves, Erica Valessa Ramos Gomes, Karoline Rodrigues Campos |
| EPI_ISL_1039703 | Instituto Adolfo Lutz - Regional de Taubate | Instituto Adolfo Lutz, Interdiciplinary Procedures Center, Strategic Laboratory | Claudio Tavares Sacchi, Claudia Regina Gonçalves, Erica Valessa Ramos Gomes, Karoline Rodrigues Campos |
| EPI_ISL_1039704 | Lab Loc - Itapecerica da Serra | Instituto Adolfo Lutz, Interdiciplinary Procedures Center, Strategic Laboratory | Claudio Tavares Sacchi, Claudia Regina Gonçalves, Erica Valessa Ramos Gomes, Karoline Rodrigues Campos |
| EPI_ISL_1039705, EPI_ISL_1039706, EPI_ISL_1039707, EPI_ISL_1039708, EPI_ISL_1039709, EPI_ISL_1039710 | Instituto Adolfo Lutz Central | Instituto Adolfo Lutz, Interdiciplinary Procedures Center, Strategic Laboratory | Claudio Tavares Sacchi, Claudia Regina Gonçalves, Erica Valessa Ramos Gomes, Karoline Rodrigues Campos |
| EPI_ISL_1040823 | Secretaria Municipal de Saude de Piracaia | Instituto Adolfo Lutz, Interdiciplinary Procedures Center, Strategic Laboratory | Claudio Tavares Sacchi, Claudia Regina Gonçalves, Erica Valessa Ramos Gomes, Karoline Rodrigues Campos |
| EPI_ISL_1040825, EPI_ISL_1040826, EPI_ISL_1040827, EPI_ISL_1040828, EPI_ISL_1040830, EPI_ISL_1040832, EPI_ISL_1040834, EPI_ISL_1040838, EPI_ISL_1040841, EPI_ISL_1040846, EPI_ISL_1040847, EPI_ISL_1040849, EPI_ISL_1040850 | | | |
| see above | LACEN do Mato Grosso do Sul | Instituto Adolfo Lutz, Interdiciplinary Procedures Center, Strategic Laboratory | Claudio Tavares Sacchi, Claudia Regina Gonçalves, Erica Valessa Ramos Gomes, Karoline Rodrigues Campos |
| EPI_ISL_1063789 | Evandro Chagas Institute | Evandro Chagas Institute Virology | Santos, M.C.; Silva, A.M.; Junior, W.D.C.; Barbagelata, L.S.; Ferreira, J.A.; Sousa, E.M.A.; da Silva, P.S.; Pinheiro, K.C.; L.C.; Sousa Junior, E.C. |
| EPI_ISL_1068082, EPI_ISL_1068089, EPI_ISL_1068090, EPI_ISL_1068093, EPI_ISL_1068095, EPI_ISL_1068096, EPI_ISL_1068101, EPI_ISL_1068102, EPI_ISL_1068107, EPI_ISL_1068127, EPI_ISL_1068132, EPI_ISL_1068134, EPI_ISL_1068135, EPI_ISL_1068137, EPI_ISL_1068146, EPI_ISL_1068148, EPI_ISL_1068152, EPI_ISL_1068161, EPI_ISL_1068168, EPI_ISL_1068172, EPI_ISL_1068175, EPI_ISL_1068182, EPI_ISL_1068190, EPI_ISL_1068192, EPI_ISL_1068197, EPI_ISL_1068205, EPI_ISL_1068206, EPI_ISL_1068208, EPI_ISL_1068209, EPI_ISL_1068210, EPI_ISL_1068211, EPI_ISL_1068213, EPI_ISL_1068214, EPI_ISL_1068217, EPI_ISL_1068218, EPI_ISL_1068223, EPI_ISL_1068242, EPI_ISL_1068244, EPI_ISL_1068245, EPI_ISL_1068246, EPI_ISL_1068247, EPI_ISL_1068250, EPI_ISL_1068253, EPI_ISL_1068254, EPI_ISL_1068257 | | | |
| see above | Laboratorio de Ecologia de Doencas Transmissiveis na Amazonia, Instituto Leonidas e Maria Deane - Fiocruz Amazonia | Laboratorio de Ecologia de Doencas Transmissiveis na Amazonia, Instituto Leonidas e Maria Deane - Fiocruz Amazonia | Valdinete Nascimento, Victor Souza, André Corado, Fernanda Nascimento, George Silva, Ágatha Costa, Debora Duarte, Karina Pessoa, Matilde Mejía, Luciana Gonçalves, Maria Júlia Brandão, Michele Jesus, Felipe Naveca on behalf of the Fiocruz COVID-19 Genomic Surveillance Network |
| EPI_ISL_1068319, EPI_ISL_1068363, EPI_ISL_1068364, EPI_ISL_1068369, EPI_ISL_1068371, EPI_ISL_1068373, EPI_ISL_1068376, EPI_ISL_1068377, EPI_ISL_1068378, EPI_ISL_1068380, EPI_ISL_1068394 | | | |
| see above | Central Public Health Laboratory - LACEN -Bahia, Salvador, Brazil | Central Public Health Laboratory - LACEN -Bahia, Salvador, Brazil | Stephane Tosta, Luciana Oliveira, Vanessa Nardy,Patrícia Cajado,Marcela Gómez, Breno Dominguez, Jaqueline Gomes, Vagner Fonseca,Marta Giovanetti,Luiz Alcantara, Felicidade Pereira, Arabela Leal |
| EPI_ISL_1078981, EPI_ISL_1078983, EPI_ISL_1078984, EPI_ISL_1078991, EPI_ISL_1078996, EPI_ISL_1079003, EPI_ISL_1079006, EPI_ISL_1079158, EPI_ISL_1079163, EPI_ISL_1079166 | IAL Regional de Bauru | Instituto Adolfo Lutz, Interdiciplinary Procedures Center, Strategic Laboratory | Claudio Tavares Sacchi, Claudia Regina Gonçalves, Erica Valessa Ramos Gomes, Karoline Rodrigues Campos |
| EPI_ISL_1086051, EPI_ISL_1086056 | IAL Regional de Bauru | Instituto Adolfo Lutz, Interdiciplinary Procedures Center, Strategic Laboratory | Claudio Tavares Sacchi, Claudia Regina Gonçalves, Erica Valessa Ramos Gomes, Karoline Rodrigues Campos, Caio Vinicius Dias Lopes |
| EPI_ISL_1086376 | LACEN - Laboratório Central de Saúde Pública do Rio Grande do Norte | Evandro Chagas Institute | Santos, M.C.; Silva, A.M.; Junior, W.D.C.; Barbagelata, L.S.; Ferreira, J.A.; Sousa, E.M.A.; da Silva, P.S.; Pinheiro, K.C.; L.C.; Sousa Junior, E.C. |
| EPI_ISL_1086377 | LACEN - Laboratório Central de Saúde Pública do Paraiba | Evandro Chagas Institute | Santos, M.C.; Silva, A.M.; Junior, W.D.C.; Barbagelata, L.S.; Ferreira, J.A.; Sousa, E.M.A.; da Silva, P.S.; Pinheiro, K.C.; L.C.; Sousa Junior, E.C. |
| EPI_ISL_1092725 | Diagnosticos da America - DASA | Instituto Adolfo Lutz, Interdiciplinary Procedures Center, Strategic Laboratory | Claudio Tavares Sacchi, Claudia Regina Gonçalves, Erica Valessa Ramos Gomes, Karoline Rodrigues Campos |
| EPI_ISL_1117384, EPI_ISL_1117388, EPI_ISL_1117408, EPI_ISL_1117429 | Nucleo de Pesquisa em Inovacao Terapeutica - UFPE | LABBE, Federal University of Pernambuco | Wilson Jose da Silva Junior, Marcos da Silveira Regueira Neto, Heidi Lacerda Alves da Cruz, Bruno Sampaio, Reginaldo Goncalves de Lima Neto, Maira Galdino da Rocha Pitta, Michelly Cristiny Pereira, Marco Katzenberger, Valdir de Queiroz Balbino |
| EPI_ISL_1121317 | IAL Regional de Bauru | Instituto Adolfo Lutz, Interdiciplinary Procedures Center, Strategic Laboratory | Claudio Tavares Sacchi, Claudia Regina Gonçalves, Erica Valessa Ramos Gomes, Karoline Rodrigues Campos, Caio Vinicius Dias Lopes |
| EPI_ISL_1121322 | Santa Casa de Santa Isabel | Instituto Adolfo Lutz, Interdiciplinary Procedures Center, Strategic Laboratory | Claudio Tavares Sacchi, Claudia Regina Gonçalves, Erica Valessa Ramos Gomes, Karoline Rodrigues Campos, Caio Vinicius Dias Lopes |
| EPI_ISL_1121323 | Complexo Hospitalar Padre Bentode Guarulhos | Instituto Adolfo Lutz, Interdiciplinary Procedures Center, Strategic Laboratory | Claudio Tavares Sacchi, Claudia Regina Gonçalves, Erica Valessa Ramos Gomes, Karoline Rodrigues Campos, Caio Vinicius Dias Lopes |
| EPI_ISL_1121326 | IAL Regional de Bauru | Instituto Adolfo Lutz, Interdiciplinary Procedures Center, Strategic Laboratory | Claudio Tavares Sacchi, Claudia Regina Gonçalves, Erica Valessa Ramos Gomes, Karoline Rodrigues Campos, Caio Vinicius Dias Lopes |
| EPI_ISL_1121329 | LACEN do Mato Grosso do Sul | Instituto Adolfo Lutz, Interdiciplinary Procedures Center, Strategic Laboratory | Claudio Tavares Sacchi, Claudia Regina Gonçalves, Erica Valessa Ramos Gomes, Karoline Rodrigues Campos, Caio Vinicius Dias Lopes |
| EPI_ISL_1123372 | UPA I Santa Isabel | Instituto Adolfo Lutz, Interdiciplinary Procedures Center, Strategic Laboratory | Claudio Tavares Sacchi, Claudia Regina Gonçalves, Erica Valessa Ramos Gomes, Karoline Rodrigues Campos, Caio Vinicius Dias Lopes |

| | | | |
|---|---|---|---|
| EPI_ISL_1123374 | IAL Regional de Santos | Instituto Adolfo Lutz, Interdiciplinary Procedures Center, Strategic Laboratory | Claudio Tavares Sacchi, Claudia Regina Gonçalves, Erica Valessa Ramos Gomes, Karoline Rodrigues Campos, Caio Vinicius Dias Lopes |
| EPI_ISL_1139052, EPI_ISL_1139054, EPI_ISL_1139056, EPI_ISL_1139057, EPI_ISL_1139060, EPI_ISL_1139067 | LACEN do Mato Grosso do Sul | Instituto Adolfo Lutz, Interdiciplinary Procedures Center, Strategic Laboratory | Claudio Tavares Sacchi, Claudia Regina Gonçalves, Erica Valessa Ramos Gomes, Karoline Rodrigues Campos, Caio Vinicius Dias Lopes |
| EPI_ISL_1164994 | LACEN - Laboratório Central de Saúde Pública do Paraiba | Evandro Chagas Institute | Santos, M.C.; Silva, A.M.; Junior, W.D.C.; Barbagelata, L.S.; Ferreira, J.A.; Sousa, E.M.A.; da Silva, P.S.; Pinheiro, K.C.; L.C.; Sousa Junior, E.C. |
| EPI_ISL_1164995 | LACEN - Laboratório Central de Saúde Pública do Ceará | Evandro Chagas Institute | Santos, M.C.; Silva, A.M.; Junior, W.D.C.; Barbagelata, L.S.; Ferreira, J.A.; Sousa, E.M.A.; da Silva, P.S.; Pinheiro, K.C.; L.C.; Sousa Junior, E.C. |
| EPI_ISL_1171620 | Instituto Adolfo Lutz Central | Instituto Adolfo Lutz, Interdiciplinary Procedures Center, Strategic Laboratory | Claudio Tavares Sacchi, Claudia Regina Gonçalves, Erica Valessa Ramos Gomes, Karoline Rodrigues Campos, Caio Vinicius Dias Lopes |
| EPI_ISL_1171621 | LACEN do Mato Grosso do Sul | Instituto Adolfo Lutz, Interdiciplinary Procedures Center, Strategic Laboratory | Claudio Tavares Sacchi, Claudia Regina Gonçalves, Erica Valessa Ramos Gomes, Karoline Rodrigues Campos, Caio Vinicius Dias Lopes |
| EPI_ISL_1171623, EPI_ISL_1171625, EPI_ISL_1171627, EPI_ISL_1171631, EPI_ISL_1171633, EPI_ISL_1171635, EPI_ISL_1171636 | IAL Regional de Santos | Instituto Adolfo Lutz, Interdiciplinary Procedures Center, Strategic Laboratory | Claudio Tavares Sacchi, Claudia Regina Gonçalves, Erica Valessa Ramos Gomes, Karoline Rodrigues Campos, Caio Vinicius Dias Lopes |
| EPI_ISL_1171646, EPI_ISL_1171647 | IAL Regional de Marilia | Instituto Adolfo Lutz, Interdiciplinary Procedures Center, Strategic Laboratory | Claudio Tavares Sacchi, Claudia Regina Gonçalves, Erica Valessa Ramos Gomes, Karoline Rodrigues Campos, Caio Vinicius Dias Lopes |
| EPI_ISL_1171664, EPI_ISL_1171668, EPI_ISL_1171669, EPI_ISL_1171671 | IAL Regional de Presidente Prudente | Instituto Adolfo Lutz, Interdiciplinary Procedures Center, Strategic Laboratory | Claudio Tavares Sacchi, Claudia Regina Gonçalves, Erica Valessa Ramos Gomes, Karoline Rodrigues Campos, Caio Vinicius Dias Lopes |
| EPI_ISL_1172014 | HC_FMUSP | Laboratório de Parasitologia Médica - Instituto de Medicina Tropical - Universidade de São Paulo | Brazil-UK Centre for Arbovirus Discovery Diagnosis Genomics and Epidemiology (CADDE) Genomic Network - Instituto de Medicina Tropical |
| EPI_ISL_1182550 | Fundação Ezequiel Dias (FUNED) | Coordenação Geral de Laboratórios de Saúde Pública (CGLAB/DAEVS/SVS/MS) | Vagner Fonseca, et al. |
| EPI_ISL_1182554 | Laboratório Central do Estado do Rio de Janeiro | Coordenação Geral de Laboratórios de Saúde Pública (CGLAB/DAEVS/SVS/MS) | Vagner Fonseca, et al. |
| EPI_ISL_1182567, EPI_ISL_1182584 | Laboratório Central do Estado do Paraná | Coordenação Geral de Laboratórios de Saúde Pública (CGLAB/DAEVS/SVS/MS) | Vagner Fonseca, et al. |
| EPI_ISL_1182587 | Fundação Ezequiel Dias (FUNED) | Coordenação Geral de Laboratórios de Saúde Pública (CGLAB/DAEVS/SVS/MS) | Vagner Fonseca, et al. |
| EPI_ISL_1182595 | Laboratório Central do Estado do Paraná | Coordenação Geral de Laboratórios de Saúde Pública (CGLAB/DAEVS/SVS/MS) | Vagner Fonseca, et al. |
| EPI_ISL_1182599 | Laboratório Central de Saúde Pública do Rio Grande do Sul | Coordenação Geral de Laboratórios de Saúde Pública (CGLAB/DAEVS/SVS/MS) | Vagner Fonseca, et al. |
| EPI_ISL_1182601, EPI_ISL_1182602 | Fundação Ezequiel Dias (FUNED) | Coordenação Geral de Laboratórios de Saúde Pública (CGLAB/DAEVS/SVS/MS) | Vagner Fonseca, et al. |
| EPI_ISL_1182603 | Laboratório Central do Estado do Paraná | Coordenação Geral de Laboratórios de Saúde Pública (CGLAB/DAEVS/SVS/MS) | Vagner Fonseca, et al. |
| EPI_ISL_1182609 | Fundação Ezequiel Dias (FUNED) | Coordenação Geral de Laboratórios de Saúde Pública (CGLAB/DAEVS/SVS/MS) | Vagner Fonseca, et al. |
| EPI_ISL_1182610 | Laboratório Central de Saúde Pública do Rio Grande do Sul | Coordenação Geral de Laboratórios de Saúde Pública (CGLAB/DAEVS/SVS/MS) | Vagner Fonseca, et al. |
| EPI_ISL_1182612 | Fundação Ezequiel Dias (FUNED) | Coordenação Geral de Laboratórios de Saúde Pública (CGLAB/DAEVS/SVS/MS) | Vagner Fonseca, et al. |
| EPI_ISL_1182613, EPI_ISL_1182614 | Laboratório Central do Estado do Paraná | Coordenação Geral de Laboratórios de Saúde Pública (CGLAB/DAEVS/SVS/MS) | Vagner Fonseca, et al. |
| EPI_ISL_1182621, EPI_ISL_1182623 | Laboratório Central de Saúde Pública do Rio Grande do Sul | Coordenação Geral de Laboratórios de Saúde Pública (CGLAB/DAEVS/SVS/MS) | Vagner Fonseca, et al. |
| EPI_ISL_1195275, EPI_ISL_1195276 | CENTRO DE REFERENCIA EM SINDROMES GRIPAIS | Epiclin | Fernando Hayashi Sant'Anna, Ana Paula Muterle, Janira Prichula, Juliana Comerlato, Carolina Comerlato, Eliana Márcia Da Ros Wendland |
| EPI_ISL_1195277 | Unidade de Atendimento DST AIDS TB e Han | Epiclin | Fernando Hayashi Sant'Anna, Ana Paula Muterle, Janira Prichula, Juliana Comerlato, Carolina Comerlato, Eliana Márcia Da Ros Wendland |
| EPI_ISL_1195278 | DIRETORIA DE VIGILANCIA EM SAUDE | Epiclin | Fernando Hayashi Sant'Anna, Ana Paula Muterle, Janira Prichula, Juliana Comerlato, Carolina Comerlato, Eliana Márcia Da Ros Wendland |
| EPI_ISL_1195279 | CENTRO DE REFERENCIA EM SINDROMES GRIPAIS | Epiclin | Fernando Hayashi Sant'Anna, Ana Paula Muterle, Janira Prichula, Juliana Comerlato, Carolina Comerlato, Eliana Márcia Da Ros Wendland |
| EPI_ISL_1195280 | SECRETARIA MUNICIPAL DE SAUDE DE ARARICA | Epiclin | Fernando Hayashi Sant'Anna, Ana Paula Muterle, Janira Prichula, Juliana Comerlato, Carolina Comerlato, Eliana Márcia Da Ros Wendland |
| EPI_ISL_1195281 | FUNDACAO DE SAUDE PUBLICA SAO CAMILO DE ESTEIO | Epiclin | Fernando Hayashi Sant'Anna, Ana Paula Muterle, Janira Prichula, Juliana Comerlato, Carolina Comerlato, Eliana Márcia Da Ros Wendland |
| EPI_ISL_1195282 | UNIDADE SANITARIA DE IGREJINHA | Epiclin | Fernando Hayashi Sant'Anna, Ana Paula Muterle, Janira Prichula, Juliana Comerlato, Carolina Comerlato, Eliana Márcia Da Ros Wendland |
| EPI_ISL_1195283 | SECRETARIA MUNICIPAL DE SAUDE DE TRES COROAS | Epiclin | Fernando Hayashi Sant'Anna, Ana Paula Muterle, Janira Prichula, Juliana Comerlato, Carolina Comerlato, Eliana Márcia Da Ros Wendland |
| EPI_ISL_1195284 | Centro de Especialidades Triunfo | Epiclin | Fernando Hayashi Sant'Anna, Ana Paula Muterle, Janira Prichula, Juliana Comerlato, Carolina Comerlato, Eliana Márcia Da Ros Wendland |
| EPI_ISL_1195285, EPI_ISL_1195286 | SECRETARIA MUNICIPAL DE SAUDE DE TRES COROAS | Epiclin | Fernando Hayashi Sant'Anna, Ana Paula Muterle, Janira Prichula, Juliana Comerlato, Carolina Comerlato, Eliana Márcia Da Ros Wendland |
| EPI_ISL_1195287 | SECRETARIA MUNICIPAL DE SAUDE DE SAO LEOPOLDO | Epiclin | Fernando Hayashi Sant'Anna, Ana Paula Muterle, Janira Prichula, Juliana Comerlato, Carolina Comerlato, Eliana Márcia Da Ros Wendland |
| EPI_ISL_1195288 | DIRETORIA DE VIGILANCIA EM SAUDE | Epiclin | Fernando Hayashi Sant'Anna, Ana Paula Muterle, Janira Prichula, Juliana Comerlato, Carolina Comerlato, Eliana Márcia Da Ros Wendland |
| EPI_ISL_1195289 | SECRETARIA MUNICIPAL DE SAUDE DE SAO LEOPOLDO | Epiclin | Fernando Hayashi Sant'Anna, Ana Paula Muterle, Janira Prichula, Juliana Comerlato, Carolina Comerlato, Eliana Márcia Da Ros Wendland |
| EPI_ISL_1195290 | SECRETARIA MUNICIPAL DE SAUDE DE TRES COROAS | Epiclin | Fernando Hayashi Sant'Anna, Ana Paula Muterle, Janira Prichula, Juliana Comerlato, Carolina Comerlato, Eliana Márcia Da Ros Wendland |
| EPI_ISL_1195291 | DIRETORIA DE VIGILANCIA EM SAUDE | Epiclin | Fernando Hayashi Sant'Anna, Ana Paula Muterle, Janira Prichula, Juliana Comerlato, Carolina Comerlato, Eliana Márcia Da Ros Wendland |
| EPI_ISL_1195292 | SECRETARIA MUNICIPAL DE SAUDE DE TRES COROAS | Epiclin | Fernando Hayashi Sant'Anna, Ana Paula Muterle, Janira Prichula, Juliana Comerlato, Carolina Comerlato, Eliana Márcia Da Ros Wendland |
| EPI_ISL_1195293 | SECRETARIA MUNICIPAL DE SAUDE DE TAQUARA | Epiclin | Fernando Hayashi Sant'Anna, Ana Paula Muterle, Janira Prichula, Juliana Comerlato, Carolina Comerlato, Eliana Márcia Da Ros Wendland |
| EPI_ISL_1196287, EPI_ISL_1196288, EPI_ISL_1196291, EPI_ISL_1196293 | LACEN do Distrito Federal | Instituto Adolfo Lutz, Interdiciplinary Procedures Center, Strategic Laboratory | Claudio Tavares Sacchi, Claudia Regina Gonçalves, Erica Valessa Ramos Gomes, Karoline Rodrigues Campos, Caio Vinicius Dias Lopes |
| EPI_ISL_1196297, EPI_ISL_1196298 | IAL Regional de Marilia | Instituto Adolfo Lutz, Interdiciplinary Procedures Center, | Claudio Tavares Sacchi, Claudia Regina Gonçalves, Erica Valessa Ramos Gomes, Karoline Rodrigues Campos, Caio Vinicius Dias Lopes |

| | | | |
|---|---|---|---|
| | | Strategic Laboratory | |
| EPI_ISL_1201886 | Aeroporto Internacional de Guarulhos | Instituto Adolfo Lutz, Interdiciplinary Procedures Center, Strategic Laboratory | Claudio Tavares Sacchi, Claudia Regina Gonçalves, Erica Valessa Ramos Gomes, Karoline Rodrigues Campos, Caio Vinicius Dias Lopes |
| EPI_ISL_1213220, EPI_ISL_1213226, EPI_ISL_1213237, EPI_ISL_1213269, EPI_ISL_1213294 | LAFEM/UESC | Bioinformatics Laboratory / LNCC | Alessandra P Lamarca, Luiz G P de Almeida, Ronaldo da Silva Francisco Jr, Lucymara Fassarella Agnez Lima, Kátia Castanho Scortecci, Vinícius Pietta Perez, Otavio J. Brustolini, Eduardo Sérgio Soares Sousa, Danielle Angst Secco, Angela Maria Guimarães Santos, George Rego Albuquerque, Ana Paula Melo Mariano, Bianca Mendes Maciel, Alexandra L Gerber, Ana Paula de C Guimarães, Paulo Ricardo Nascimento, Francisco Paulo Freire Neto, Sandra Rocha Gadelha, Luís Cristóvão Porto, Eloiza Helena Campana, Selma Maria Bezerra Jeronimo, Ana Tereza R Vasconcelos |
| EPI_ISL_1213309 | Laboratório HLA/UERJ | Bioinformatics Laboratory / LNCC | Alessandra P Lamarca, Luiz G P de Almeida, Ronaldo da Silva Francisco Jr, Lucymara Fassarella Agnez Lima, Kátia Castanho Scortecci, Vinícius Pietta Perez, Otavio J. Brustolini, Eduardo Sérgio Soares Sousa, Danielle Angst Secco, Angela Maria Guimarães Santos, George Rego Albuquerque, Ana Paula Melo Mariano, Bianca Mendes Maciel, Alexandra L Gerber, Ana Paula de C Guimarães, Paulo Ricardo Nascimento, Francisco Paulo Freire Neto, Sandra Rocha Gadelha, Luís Cristóvão Porto, Eloiza Helena Campana, Selma Maria Bezerra Jeronimo, Ana Tereza R Vasconcelos |
| EPI_ISL_1213317 | IMT-UFRN/RN | Bioinformatics Laboratory / LNCC | Alessandra P Lamarca, Luiz G P de Almeida, Ronaldo da Silva Francisco Jr, Lucymara Fassarella Agnez Lima, Kátia Castanho Scortecci, Vinícius Pietta Perez, Otavio J. Brustolini, Eduardo Sérgio Soares Sousa, Danielle Angst Secco, Angela Maria Guimarães Santos, George Rego Albuquerque, Ana Paula Melo Mariano, Bianca Mendes Maciel, Alexandra L Gerber, Ana Paula de C Guimarães, Paulo Ricardo Nascimento, Francisco Paulo Freire Neto, Sandra Rocha Gadelha, Luís Cristóvão Porto, Eloiza Helena Campana, Selma Maria Bezerra Jeronimo, Ana Tereza R Vasconcelos |
| EPI_ISL_1213397 | Laboratório HLA/UERJ | Bioinformatics Laboratory / LNCC | Alessandra P Lamarca, Luiz G P de Almeida, Ronaldo da Silva Francisco Jr, Lucymara Fassarella Agnez Lima, Kátia Castanho Scortecci, Vinícius Pietta Perez, Otavio J. Brustolini, Eduardo Sérgio Soares Sousa, Danielle Angst Secco, Angela Maria Guimarães Santos, George Rego Albuquerque, Ana Paula Melo Mariano, Bianca Mendes Maciel, Alexandra L Gerber, Ana Paula de C Guimarães, Paulo Ricardo Nascimento, Francisco Paulo Freire Neto, Sandra Rocha Gadelha, Luís Cristóvão Porto, Eloiza Helena Campana, Selma Maria Bezerra Jeronimo, Ana Tereza R Vasconcelos |
| EPI_ISL_1213401 | LAFEM/UESC | Bioinformatics Laboratory / LNCC | Alessandra P Lamarca, Luiz G P de Almeida, Ronaldo da Silva Francisco Jr, Lucymara Fassarella Agnez Lima, Kátia Castanho Scortecci, Vinícius Pietta Perez, Otavio J. Brustolini, Eduardo Sérgio Soares Sousa, Danielle Angst Secco, Angela Maria Guimarães Santos, George Rego Albuquerque, Ana Paula Melo Mariano, Bianca Mendes Maciel, Alexandra L Gerber, Ana Paula de C Guimarães, Paulo Ricardo Nascimento, Francisco Paulo Freire Neto, Sandra Rocha Gadelha, Luís Cristóvão Porto, Eloiza Helena Campana, Selma Maria Bezerra Jeronimo, Ana Tereza R Vasconcelos |
| EPI_ISL_1213429, EPI_ISL_1213433, EPI_ISL_1213443, EPI_ISL_1213453, EPI_ISL_1213454, EPI_ISL_1213458, EPI_ISL_1213459, EPI_ISL_1213461 | LBM/UFPB | Bioinformatics Laboratory / LNCC | Alessandra P Lamarca, Luiz G P de Almeida, Ronaldo da Silva Francisco Jr, Lucymara Fassarella Agnez Lima, Kátia Castanho Scortecci, Vinícius Pietta Perez, Otavio J. Brustolini, Eduardo Sérgio Soares Sousa, Danielle Angst Secco, Angela Maria Guimarães Santos, George Rego Albuquerque, Ana Paula Melo Mariano, Bianca Mendes Maciel, Alexandra L Gerber, Ana Paula de C Guimarães, Paulo Ricardo Nascimento, Francisco Paulo Freire Neto, Sandra Rocha Gadelha, Luís Cristóvão Porto, Eloiza Helena Campana, Selma Maria Bezerra Jeronimo, Ana Tereza R Vasconcelos |
| EPI_ISL_1219028, EPI_ISL_1219032 | Aeroporto Internacional de Guarulhos | Instituto Adolfo Lutz, Interdiciplinary Procedures Center, Strategic Laboratory | Claudio Tavares Sacchi, Claudia Regina Gonçalves, Erica Valessa Ramos Gomes, Karoline Rodrigues Campos, Caio Vinicius Dias Lopes |
| EPI_ISL_1239116, EPI_ISL_1239117 | Laboratório Central de Saúde Pública do Espírito Santo | Coordenação Geral de Laboratórios de Saúde Pública (CGLAB) | Vagner Fonseca et al, |
| EPI_ISL_1261698 | LACEN - Laboratório Central de Saúde Pública de Pernambuco | Evandro Chagas Institute | Santos, M.C.; Silva, A.M.; Junior, W.D.C.; Barbagelata, L.S.; Ferreira, J.A.; Sousa, E.M.A.; da Silva, P.S.; Pinheiro, K.C.; L.C.; Sousa Junior, E.C. |
| EPI_ISL_1293052 | LACEN de Rondonia | Instituto Adolfo Lutz, Interdiciplinary Procedures Center, Strategic Laboratory | Claudio Tavares Sacchi, Claudia Regina Gonçalves, Erica Valessa Ramos Gomes, Karoline Rodrigues Campos, Caio Vinicius Dias Lopes |
| EPI_ISL_1293056, EPI_ISL_1293057, EPI_ISL_1293064, EPI_ISL_1293069, EPI_ISL_1293072, EPI_ISL_1293081 | IAL Regional de Sorocaba | Instituto Adolfo Lutz, Interdiciplinary Procedures Center, Strategic Laboratory | Claudio Tavares Sacchi, Claudia Regina Gonçalves, Erica Valessa Ramos Gomes, Karoline Rodrigues Campos, Caio Vinicius Dias Lopes |
| EPI_ISL_1303499 | LACEN de Rondonia | Instituto Adolfo Lutz, Interdiciplinary Procedures Center, Strategic Laboratory | Claudio Tavares Sacchi, Claudia Regina Gonçalves, Erica Valessa Ramos Gomes, Karoline Rodrigues Campos, Caio Vinicius Dias Lopes |
| EPI_ISL_1303510 | LACEN do Estado de Goias | Instituto Adolfo Lutz, Interdiciplinary Procedures Center, Strategic Laboratory | Claudio Tavares Sacchi, Claudia Regina Gonçalves, Erica Valessa Ramos Gomes, Karoline Rodrigues Campos, Caio Vinicius Dias Lopes |
| EPI_ISL_1303527, EPI_ISL_1303528 | IAL Regional de São Jose do Rio Preto | Instituto Adolfo Lutz, Interdiciplinary Procedures Center, Strategic Laboratory | Claudio Tavares Sacchi, Claudia Regina Gonçalves, Erica Valessa Ramos Gomes, Karoline Rodrigues Campos, Caio Vinicius Dias Lopes |
| EPI_ISL_1303544 | Hospital Municipal Cidade Tiradentes Carmen Prudente | Instituto Adolfo Lutz, Interdiciplinary Procedures Center, Strategic Laboratory | Claudio Tavares Sacchi, Claudia Regina Gonçalves, Erica Valessa Ramos Gomes, Karoline Rodrigues Campos, Caio Vinicius Dias Lopes |
| EPI_ISL_1324137, EPI_ISL_1324140 | UW Virology Lab | UW Virology Lab | Pavitra Roychoudhury, Hong Xie, Lasata Shrestha, Shah Mohamed Bakhash, Michelle Lin, Margaret Mills, Noah Baker, Sean Ellis, Saraswathi Sathees, Meei-Li Huang, Keith R Jerome, Alexander Greninger |
| EPI_ISL_1358296, EPI_ISL_1358297 | IAL Regional de Santo Andre | Instituto Adolfo Lutz, Interdiciplinary Procedures Center, Strategic Laboratory | Claudio Tavares Sacchi, Claudia Regina Gonçalves, Erica Valessa Ramos Gomes, Karoline Rodrigues Campos, Caio Vinicius Dias Lopes |
| EPI_ISL_1358302 | Lacen de Tocantins | Instituto Adolfo Lutz, Interdiciplinary Procedures Center, Strategic Laboratory | Claudio Tavares Sacchi, Claudia Regina Gonçalves, Erica Valessa Ramos Gomes, Karoline Rodrigues Campos, Caio Vinicius Dias Lopes |
| EPI_ISL_1358305, EPI_ISL_1358307, EPI_ISL_1358308, EPI_ISL_1358309, EPI_ISL_1358311, EPI_ISL_1358314 | LACEN do Mato Grosso do Sul | Instituto Adolfo Lutz, Interdiciplinary Procedures Center, Strategic Laboratory | Claudio Tavares Sacchi, Claudia Regina Gonçalves, Erica Valessa Ramos Gomes, Karoline Rodrigues Campos, Caio Vinicius Dias Lopes |
| EPI_ISL_1358322 | UPA Vila Santa Catarina | Instituto Adolfo Lutz, Interdiciplinary Procedures Center, Strategic Laboratory | Claudio Tavares Sacchi, Claudia Regina Gonçalves, Erica Valessa Ramos Gomes, Karoline Rodrigues Campos, Caio Vinicius Dias Lopes |
| EPI_ISL_1381044, EPI_ISL_1381046, EPI_ISL_1381049, EPI_ISL_1381064 | IAL Regional de Santo Andre | Instituto Adolfo Lutz, Interdiciplinary Procedures Center, Strategic Laboratory | Claudio Tavares Sacchi, Claudia Regina Gonçalves, Erica Valessa Ramos Gomes, Karoline Rodrigues Campos, Caio Vinicius Dias Lopes |
| EPI_ISL_1445089 | CENTRO DE SAUDE II MAIRINQUE MAIRINQUE | Instituto Butantan / Mendelics | Dimas Tadeu Covas, Sandra Coccuzzo Sampaio, Maria Carolina Elias, José Salvatore Leister Patané, Vincent Louis Viala, Antonio Jorge Martins, Ricardo Haddad, Claudia Renata dos Santos Barros, Elaine Cristina Marqueze, Raul Machado Neto, Debora Botequio Moretti, Bibiana Santos, João Paulo Kitajima, Erika Freitas, David Schlesinger, Simone Kashima, Evandra Strazza Rodrigues, Svetoslav Nanev Slavov, Elaine Vieira dos Santos, Rafael dos Santos Bezerra, Luiz Carlos Junior de Alcantara, Marta Giovanetti, Vagner Fonseca, Flavia Aburjaile, Rodrigo Tocantins Calado. |
| EPI_ISL_1445090 | POLICLINICA COVID 19 ITAPETININGA | Instituto Butantan / Mendelics | Dimas Tadeu Covas, Sandra Coccuzzo Sampaio, Maria Carolina Elias, José Salvatore Leister Patané, Vincent Louis Viala, Antonio Jorge Martins, Ricardo Haddad, Claudia Renata dos Santos Barros, Elaine Cristina Marqueze, Raul Machado Neto, Debora Botequio Moretti, Bibiana Santos, João Paulo Kitajima, Erika Freitas, David Schlesinger, Simone Kashima, Evandra Strazza Rodrigues, Svetoslav Nanev Slavov, Elaine Vieira dos Santos, Rafael dos Santos Bezerra, Luiz Carlos Junior de Alcantara, Marta Giovanetti, Vagner Fonseca, Flavia Aburjaile, Rodrigo Tocantins Calado. |
| EPI_ISL_1445134 | SECRETARIA MUNICIPAL DE SAUDE SOROCABA | Instituto Butantan / Mendelics | Dimas Tadeu Covas, Sandra Coccuzzo Sampaio, Maria Carolina Elias, José Salvatore Leister Patané, Vincent Louis Viala, Antonio Jorge Martins, Ricardo Haddad, Claudia Renata dos Santos Barros, Elaine Cristina Marqueze, Raul Machado Neto, Debora Botequio Moretti, Bibiana Santos, João Paulo Kitajima, Erika Freitas, David Schlesinger, Simone Kashima, Evandra Strazza Rodrigues, Svetoslav Nanev Slavov, Elaine Vieira dos Santos, Rafael dos Santos Bezerra, Luiz Carlos Junior de Alcantara, Marta Giovanetti, Vagner Fonseca, Flavia Aburjaile, Rodrigo Tocantins Calado. |
| EPI_ISL_1445195 | CENTRO DE SAUDE DE BORA | Instituto Butantan / Mendelics | Dimas Tadeu Covas, Sandra Coccuzzo Sampaio, Maria Carolina Elias, José Salvatore Leister Patané, Vincent Louis Viala, Antonio Jorge Martins, Ricardo Haddad, Claudia Renata dos Santos Barros, Elaine Cristina Marqueze, Raul Machado Neto, Debora Botequio Moretti, Bibiana Santos, João Paulo Kitajima, Erika Freitas, David Schlesinger, Simone Kashima, Evandra Strazza Rodrigues, Svetoslav Nanev Slavov, Elaine Vieira dos Santos, Rafael dos |

| | | | |
|---|---|---|---|
| | | | Santos Bezerra, Luiz Carlos Junior de Alcantara, Marta Giovanetti, Vagner Fonseca, Flavia Aburjaile, Rodrigo Tocantins Calado. |
| EPI_ISL_1445235 | VIGILANCIA EPIDEMIOLOGICA | Instituto Butantan / Mendelics | Dimas Tadeu Covas, Sandra Coccuzzo Sampaio, Maria Carolina Elias, José Salvatore Leister Patané, Vincent Louis Viala, Antonio Jorge Martins, Ricardo Haddad, Claudia Renata dos Santos Barros, Elaine Cristina Marqueze, Raul Machado Neto, Debora Botequio Moretti, Bibiana Santos, João Paulo Kitajima, Erika Freitas, David Schlesinger, Simone Kashima, Evandra Strazza Rodrigues, Svetoslav Nanev Slavov, Elaine Vieira dos Santos, Rafael dos Santos Bezerra, Luiz Carlos Junior de Alcantara, Marta Giovanetti, Vagner Fonseca, Flavia Aburjaile, Rodrigo Tocantins Calado. |
| EPI_ISL_1445242 | SECAO CENTRO DE DIAGNOSTICO SECEDI | Instituto Butantan / Mendelics | Dimas Tadeu Covas, Sandra Coccuzzo Sampaio, Maria Carolina Elias, José Salvatore Leister Patané, Vincent Louis Viala, Antonio Jorge Martins, Ricardo Haddad, Claudia Renata dos Santos Barros, Elaine Cristina Marqueze, Raul Machado Neto, Debora Botequio Moretti, Bibiana Santos, João Paulo Kitajima, Erika Freitas, David Schlesinger, Simone Kashima, Evandra Strazza Rodrigues, Svetoslav Nanev Slavov, Elaine Vieira dos Santos, Rafael dos Santos Bezerra, Luiz Carlos Junior de Alcantara, Marta Giovanetti, Vagner Fonseca, Flavia Aburjaile, Rodrigo Tocantins Calado. |
| EPI_ISL_1445249, EPI_ISL_1445270 | VIGILANCIA EPIDEMIOLOGICA | Instituto Butantan / Mendelics | Dimas Tadeu Covas, Sandra Coccuzzo Sampaio, Maria Carolina Elias, José Salvatore Leister Patané, Vincent Louis Viala, Antonio Jorge Martins, Ricardo Haddad, Claudia Renata dos Santos Barros, Elaine Cristina Marqueze, Raul Machado Neto, Debora Botequio Moretti, Bibiana Santos, João Paulo Kitajima, Erika Freitas, David Schlesinger, Simone Kashima, Evandra Strazza Rodrigues, Svetoslav Nanev Slavov, Elaine Vieira dos Santos, Rafael dos Santos Bezerra, Luiz Carlos Junior de Alcantara, Marta Giovanetti, Vagner Fonseca, Flavia Aburjaile, Rodrigo Tocantins Calado. |
| EPI_ISL_1464675, EPI_ISL_1464677 | Laboratório de Virologia - UNIFESP | Laboratory of Respiratory Viruses and Measles, Oswaldo Cruz Institute, FIOCRUZ | Paola Resende, Nancy Belei, Luciana Appolinario, Fernando Motta, Anna Carolina Paixao, Ana Carolina Mendonca, Alice Sampaio Rocha, Renata Serrano Lopes, Marilda Siqueira on behalf of the Fiocruz COVID-19 Genomic Surveillance Network |
| EPI_ISL_1465225 | Laboratorio Central de Saude Publica do Estado do Maranhao (LACEN-MA) | Laboratory of Respiratory Viruses and Measles, Oswaldo Cruz Institute, FIOCRUZ | Paola Resende, Luciana Appolinario, Fernando Motta, Anna Carolina Paixao, Ana Carolina Mendonca, Alice Sampaio Rocha, Renata Serrano Lopes, Lidio Gonçalves Lima Neto, Marilda Siqueira on behalf of the Fiocruz COVID-19 Genomic Surveillance Network |
| EPI_ISL_1468437 | LACEN do Mato Grosso do Sul | Instituto Adolfo Lutz, Interdiciplinary Procedures Center, Strategic Laboratory | Claudio Tavares Sacchi, Claudia Regina Gonçalves, Erica Valessa Ramos Gomes, Karoline Rodrigues Campos, Caio Vinicius Dias Lopes |
| EPI_ISL_1468442 | Santa Casa de Birigui | Instituto Adolfo Lutz, Interdiciplinary Procedures Center, Strategic Laboratory | Claudio Tavares Sacchi, Claudia Regina Gonçalves, Erica Valessa Ramos Gomes, Karoline Rodrigues Campos, Caio Vinicius Dias Lopes |
| EPI_ISL_1468449 | Santa Casa de Aracatuba Hospital Sagrado Coracao de Jesus | Instituto Adolfo Lutz, Interdiciplinary Procedures Center, Strategic Laboratory | Claudio Tavares Sacchi, Claudia Regina Gonçalves, Erica Valessa Ramos Gomes, Karoline Rodrigues Campos, Caio Vinicius Dias Lopes |
| EPI_ISL_1468453 | Santa Casa de Misericordia de Pereira Barreto | Instituto Adolfo Lutz, Interdiciplinary Procedures Center, Strategic Laboratory | Claudio Tavares Sacchi, Claudia Regina Gonçalves, Erica Valessa Ramos Gomes, Karoline Rodrigues Campos, Caio Vinicius Dias Lopes |
| EPI_ISL_1468454 | Santa Casa de Andradina | Instituto Adolfo Lutz, Interdiciplinary Procedures Center, Strategic Laboratory | Claudio Tavares Sacchi, Claudia Regina Gonçalves, Erica Valessa Ramos Gomes, Karoline Rodrigues Campos, Caio Vinicius Dias Lopes |
| EPI_ISL_1468459 | Secretaria Municipal de Saude de Valparaiso SP | Instituto Adolfo Lutz, Interdiciplinary Procedures Center, Strategic Laboratory | Claudio Tavares Sacchi, Claudia Regina Gonçalves, Erica Valessa Ramos Gomes, Karoline Rodrigues Campos, Caio Vinicius Dias Lopes |
| EPI_ISL_1468466 | Santa Casa de Sao Carlos | Instituto Adolfo Lutz, Interdiciplinary Procedures Center, Strategic Laboratory | Claudio Tavares Sacchi, Claudia Regina Gonçalves, Erica Valessa Ramos Gomes, Karoline Rodrigues Campos, Caio Vinicius Dias Lopes |
| EPI_ISL_1468469 | Secretaria Municipal de Saude Porto Ferreira | Instituto Adolfo Lutz, Interdiciplinary Procedures Center, Strategic Laboratory | Claudio Tavares Sacchi, Claudia Regina Gonçalves, Erica Valessa Ramos Gomes, Karoline Rodrigues Campos, Caio Vinicius Dias Lopes |
| EPI_ISL_1468470, EPI_ISL_1468471 | Secretaria Municipal de Saude Descalvado | Instituto Adolfo Lutz, Interdiciplinary Procedures Center, Strategic Laboratory | Claudio Tavares Sacchi, Claudia Regina Gonçalves, Erica Valessa Ramos Gomes, Karoline Rodrigues Campos, Caio Vinicius Dias Lopes |
| EPI_ISL_1469555 | FUNDACAO DE SAUDE PUBLICA DE NOVO HAMBURGO FSNH | Epiclin | Fernando Hayashi Sant'Anna, Ana Paula Muterle, Janira Prichula, Juliana Comerlato, Carolina Comerlato, Eliana Márcia Da Ros Wendland |
| EPI_ISL_1469560 | Diretoria de Vigilância em Saúde | Epiclin | Fernando Hayashi Sant'Anna, Ana Paula Muterle, Janira Prichula, Juliana Comerlato, Carolina Comerlato, Eliana Márcia Da Ros Wendland |
| EPI_ISL_1469561 | Fundação de Saúde Pública São Camilo de Esteio | Epiclin | Fernando Hayashi Sant'Anna, Ana Paula Muterle, Janira Prichula, Juliana Comerlato, Carolina Comerlato, Eliana Márcia Da Ros Wendland |
| EPI_ISL_1469570 | Hospital Universitário | Epiclin | Fernando Hayashi Sant'Anna, Ana Paula Muterle, Janira Prichula, Juliana Comerlato, Carolina Comerlato, Eliana Márcia Da Ros Wendland |
| EPI_ISL_1469571 | Unidade de Pronto Atendimento de Sapucaia do Sul | Epiclin | Fernando Hayashi Sant'Anna, Ana Paula Muterle, Janira Prichula, Juliana Comerlato, Carolina Comerlato, Eliana Márcia Da Ros Wendland |
| EPI_ISL_1469572 | FUNDACAO DE SAUDE PUBLICA DE NOVO HAMBURGO FSNH | Epiclin | Fernando Hayashi Sant'Anna, Ana Paula Muterle, Janira Prichula, Juliana Comerlato, Carolina Comerlato, Eliana Márcia Da Ros Wendland |
| EPI_ISL_1469573 | VIGILANCIA EM SAUDE NH | Epiclin | Fernando Hayashi Sant'Anna, Ana Paula Muterle, Janira Prichula, Juliana Comerlato, Carolina Comerlato, Eliana Márcia Da Ros Wendland |
| EPI_ISL_1469576 | Unidade Sanitária de Igrejinha | Epiclin | Fernando Hayashi Sant'Anna, Ana Paula Muterle, Janira Prichula, Juliana Comerlato, Carolina Comerlato, Eliana Márcia Da Ros Wendland |
| EPI_ISL_1469579 | DIRETORIA DE VIGILANCIA EM SAUDE | Epiclin | Fernando Hayashi Sant'Anna, Ana Paula Muterle, Janira Prichula, Juliana Comerlato, Carolina Comerlato, Eliana Márcia Da Ros Wendland |
| EPI_ISL_1469580 | Unidade de Pronto Atendimento de Sapucaia do Sul | Epiclin | Fernando Hayashi Sant'Anna, Ana Paula Muterle, Janira Prichula, Juliana Comerlato, Carolina Comerlato, Eliana Márcia Da Ros Wendland |
| EPI_ISL_1469584 | CENTRO MUNICIPAL DE SAUDE DE ROLANTE | Epiclin | Fernando Hayashi Sant'Anna, Ana Paula Muterle, Janira Prichula, Juliana Comerlato, Carolina Comerlato, Eliana Márcia Da Ros Wendland |
| EPI_ISL_1469586 | FUNDACAO HOSPITALAR SAO JOSE | Epiclin | Fernando Hayashi Sant'Anna, Ana Paula Muterle, Janira Prichula, Juliana Comerlato, Carolina Comerlato, Eliana Márcia Da Ros Wendland |
| EPI_ISL_1469588 | Secretaria Municipal de Saúde de Taquara | Epiclin | Fernando Hayashi Sant'Anna, Ana Paula Muterle, Janira Prichula, Juliana Comerlato, Carolina Comerlato, Eliana Márcia Da Ros Wendland |
| EPI_ISL_1469593 | Hospital Sapiranga | Epiclin | Fernando Hayashi Sant'Anna, Ana Paula Muterle, Janira Prichula, Juliana Comerlato, Carolina Comerlato, Eliana Márcia Da Ros Wendland |
| EPI_ISL_1469604, EPI_ISL_1469608 | COORDENADORIA GERAL DE VIGILANCIA EM SAUDE | Epiclin | Fernando Hayashi Sant'Anna, Ana Paula Muterle, Janira Prichula, Juliana Comerlato, Carolina Comerlato, Eliana Márcia Da Ros Wendland |
| EPI_ISL_1469609, EPI_ISL_1469610 | Diretoria de Vigilância em Saúde | Epiclin | Fernando Hayashi Sant'Anna, Ana Paula Muterle, Janira Prichula, Juliana Comerlato, Carolina Comerlato, Eliana Márcia Da Ros Wendland |
| EPI_ISL_1469615 | Pronto Atendimento Campo Bom | Epiclin | Fernando Hayashi Sant'Anna, Ana Paula Muterle, Janira Prichula, Juliana Comerlato, Carolina Comerlato, Eliana Márcia Da Ros Wendland |
| EPI_ISL_1469616 | Diretoria de Vigilância em Saúde | Epiclin | Fernando Hayashi Sant'Anna, Ana Paula Muterle, Janira Prichula, Juliana Comerlato, Carolina Comerlato, Eliana Márcia Da Ros Wendland |
| EPI_ISL_1469620 | FUNDACAO DE SAUDE PUBLICA DE NOVO HAMBURGO FSNH | Epiclin | Fernando Hayashi Sant'Anna, Ana Paula Muterle, Janira Prichula, Juliana Comerlato, Carolina Comerlato, Eliana Márcia Da Ros Wendland |
| EPI_ISL_1469623 | Diretoria de Vigilância em Saúde | Epiclin | Fernando Hayashi Sant'Anna, Ana Paula Muterle, Janira Prichula, Juliana Comerlato, Carolina Comerlato, Eliana Márcia Da Ros Wendland |
| EPI_ISL_1469624 | HOSPITAL MUNICIPAL GETULIO VARGAS | Epiclin | Fernando Hayashi Sant'Anna, Ana Paula Muterle, Janira Prichula, Juliana Comerlato, Carolina Comerlato, Eliana Márcia Da Ros Wendland |
| EPI_ISL_1469625, EPI_ISL_1469627 | Diretoria de Vigilância em Saúde | Epiclin | Fernando Hayashi Sant'Anna, Ana Paula Muterle, Janira Prichula, Juliana Comerlato, Carolina Comerlato, Eliana Márcia Da Ros Wendland |
| EPI_ISL_1469629 | CENTRO MUNICIPAL DE SAUDE DE ROLANTE | Epiclin | Fernando Hayashi Sant'Anna, Ana Paula Muterle, Janira Prichula, Juliana Comerlato, Carolina Comerlato, Eliana Márcia Da Ros Wendland |
| EPI_ISL_1469631 | HOSPITAL MUNICIPAL GETULIO VARGAS | Epiclin | Fernando Hayashi Sant'Anna, Ana Paula Muterle, Janira Prichula, Juliana Comerlato, Carolina Comerlato, Eliana Márcia Da Ros Wendland |
| EPI_ISL_1469633 | DIRETORIA DE VIGILANCIA EM SAUDE | Epiclin | Fernando Hayashi Sant'Anna, Ana Paula Muterle, Janira Prichula, Juliana Comerlato, Carolina Comerlato, Eliana Márcia Da Ros Wendland |
| EPI_ISL_1469636 | COORDENADORIA GERAL DE VIGILANCIA EM SAUDE | Epiclin | Fernando Hayashi Sant'Anna, Ana Paula Muterle, Janira Prichula, Juliana Comerlato, Carolina Comerlato, Eliana Márcia Da Ros Wendland |
| EPI_ISL_1469637, EPI_ISL_1469638 | DIRETORIA DE VIGILANCIA EM SAUDE | Epiclin | Fernando Hayashi Sant'Anna, Ana Paula Muterle, Janira Prichula, Juliana Comerlato, Carolina Comerlato, Eliana Márcia Da Ros Wendland |
| EPI_ISL_1469641 | COORDENADORIA GERAL DE VIGILANCIA EM SAUDE | Epiclin | Fernando Hayashi Sant'Anna, Ana Paula Muterle, Janira Prichula, Juliana Comerlato, Carolina Comerlato, Eliana Márcia Da Ros Wendland |

| | | | |
|---|---|---|---|
| EPI_ISL_1469642 | CENTRO DE REFERENCIA EM SINDROMES GRIPAIS | Epiclin | Fernando Hayashi Sant'Anna, Ana Paula Muterle, Janira Prichula, Juliana Comerlato, Carolina Comerlato, Eliana Márcia Da Ros Wendland |
| EPI_ISL_1469647 | Secretaria Municipal de Saúde de Taquara | Epiclin | Fernando Hayashi Sant'Anna, Ana Paula Muterle, Janira Prichula, Juliana Comerlato, Carolina Comerlato, Eliana Márcia Da Ros Wendland |
| EPI_ISL_1469656 | Diretoria de Vigilância em Saúde | Epiclin | Fernando Hayashi Sant'Anna, Ana Paula Muterle, Janira Prichula, Juliana Comerlato, Carolina Comerlato, Eliana Márcia Da Ros Wendland |
| EPI_ISL_1469657 | Pronto Atendimento Campo Bom | Epiclin | Fernando Hayashi Sant'Anna, Ana Paula Muterle, Janira Prichula, Juliana Comerlato, Carolina Comerlato, Eliana Márcia Da Ros Wendland |
| EPI_ISL_1469658 | CENTRO DE ESPECIALIDADES TRIUNFO | Epiclin | Fernando Hayashi Sant'Anna, Ana Paula Muterle, Janira Prichula, Juliana Comerlato, Carolina Comerlato, Eliana Márcia Da Ros Wendland |
| EPI_ISL_1469661 | FUNDACAO DE SAUDE PUBLICA DE NOVO HAMBURGO FSNH | Epiclin | Fernando Hayashi Sant'Anna, Ana Paula Muterle, Janira Prichula, Juliana Comerlato, Carolina Comerlato, Eliana Márcia Da Ros Wendland |
| EPI_ISL_1469664 | Unidade de Atendimento DST AIDS TB e Han | Epiclin | Fernando Hayashi Sant'Anna, Ana Paula Muterle, Janira Prichula, Juliana Comerlato, Carolina Comerlato, Eliana Márcia Da Ros Wendland |
| EPI_ISL_1469669 | Diretoria de Vigilância em Saúde | Epiclin | Fernando Hayashi Sant'Anna, Ana Paula Muterle, Janira Prichula, Juliana Comerlato, Carolina Comerlato, Eliana Márcia Da Ros Wendland |
| EPI_ISL_1469675, EPI_ISL_1469677 | DIRETORIA DE VIGILANCIA EM SAUDE | Epiclin | Fernando Hayashi Sant'Anna, Ana Paula Muterle, Janira Prichula, Juliana Comerlato, Carolina Comerlato, Eliana Márcia Da Ros Wendland |
| EPI_ISL_1469683 | SECRETARIA MUNICIPAL DE SAUDE DE TAQUARA | Epiclin | Fernando Hayashi Sant'Anna, Ana Paula Muterle, Janira Prichula, Juliana Comerlato, Carolina Comerlato, Eliana Márcia Da Ros Wendland |
| EPI_ISL_1469684 | DIRETORIA DE VIGILANCIA EM SAUDE | Epiclin | Fernando Hayashi Sant'Anna, Ana Paula Muterle, Janira Prichula, Juliana Comerlato, Carolina Comerlato, Eliana Márcia Da Ros Wendland |
| EPI_ISL_1469687 | SECRETARIA MUNICIPAL DE SAUDE DE TAQUARA | Epiclin | Fernando Hayashi Sant'Anna, Ana Paula Muterle, Janira Prichula, Juliana Comerlato, Carolina Comerlato, Eliana Márcia Da Ros Wendland |
| EPI_ISL_1469692 | DIRETORIA DE VIGILANCIA EM SAUDE | Epiclin | Fernando Hayashi Sant'Anna, Ana Paula Muterle, Janira Prichula, Juliana Comerlato, Carolina Comerlato, Eliana Márcia Da Ros Wendland |
| EPI_ISL_1469696 | Vigilância em Saúde de Sapucaia do Sul | Epiclin | Fernando Hayashi Sant'Anna, Ana Paula Muterle, Janira Prichula, Juliana Comerlato, Carolina Comerlato, Eliana Márcia Da Ros Wendland |
| EPI_ISL_1469704 | FUNDACAO DE SAUDE PUBLICA SAO CAMILO DE ESTEIO | Epiclin | Fernando Hayashi Sant'Anna, Ana Paula Muterle, Janira Prichula, Juliana Comerlato, Carolina Comerlato, Eliana Márcia Da Ros Wendland |
| EPI_ISL_1469708 | Diretoria de Vigilância em Saúde | Epiclin | Fernando Hayashi Sant'Anna, Ana Paula Muterle, Janira Prichula, Juliana Comerlato, Carolina Comerlato, Eliana Márcia Da Ros Wendland |
| EPI_ISL_1469713 | Fundação Hospitalar de Sapucaia do Sul | Epiclin | Fernando Hayashi Sant'Anna, Ana Paula Muterle, Janira Prichula, Juliana Comerlato, Carolina Comerlato, Eliana Márcia Da Ros Wendland |
| EPI_ISL_1469714 | Unidade de Atendimento DST AIDS TB e Han | Epiclin | Fernando Hayashi Sant'Anna, Ana Paula Muterle, Janira Prichula, Juliana Comerlato, Carolina Comerlato, Eliana Márcia Da Ros Wendland |
| EPI_ISL_1469719 | UNIDADE DE PRONTO ATENDIMENTO DE SAPUCAIA DO SUL UPA | Epiclin | Fernando Hayashi Sant'Anna, Ana Paula Muterle, Janira Prichula, Juliana Comerlato, Carolina Comerlato, Eliana Márcia Da Ros Wendland |
| EPI_ISL_1469720 | UNIDADE SANITARIA DE IGREJINHA | Epiclin | Fernando Hayashi Sant'Anna, Ana Paula Muterle, Janira Prichula, Juliana Comerlato, Carolina Comerlato, Eliana Márcia Da Ros Wendland |
| EPI_ISL_1469721 | Pronto Atendimento Cruzeiro do Sul | Epiclin | Fernando Hayashi Sant'Anna, Ana Paula Muterle, Janira Prichula, Juliana Comerlato, Carolina Comerlato, Eliana Márcia Da Ros Wendland |
| EPI_ISL_1469722 | UNIDADE SANITARIA DE IGREJINHA | Epiclin | Fernando Hayashi Sant'Anna, Ana Paula Muterle, Janira Prichula, Juliana Comerlato, Carolina Comerlato, Eliana Márcia Da Ros Wendland |
| EPI_ISL_1469723 | FUNDACAO DE SAUDE PUBLICA SAO CAMILO DE ESTEIO | Epiclin | Fernando Hayashi Sant'Anna, Ana Paula Muterle, Janira Prichula, Juliana Comerlato, Carolina Comerlato, Eliana Márcia Da Ros Wendland |
| EPI_ISL_1469730 | CENTRO DE REFERENCIA EM SINDROMES GRIPAIS | Epiclin | Fernando Hayashi Sant'Anna, Ana Paula Muterle, Janira Prichula, Juliana Comerlato, Carolina Comerlato, Eliana Márcia Da Ros Wendland |
| EPI_ISL_1469731 | DIRETORIA DE VIGILANCIA EM SAUDE | Epiclin | Fernando Hayashi Sant'Anna, Ana Paula Muterle, Janira Prichula, Juliana Comerlato, Carolina Comerlato, Eliana Márcia Da Ros Wendland |
| EPI_ISL_1469733 | UNIDADE DE PRONTO ATENDIMENTO DE SAPUCAIA DO SUL UPA | Epiclin | Fernando Hayashi Sant'Anna, Ana Paula Muterle, Janira Prichula, Juliana Comerlato, Carolina Comerlato, Eliana Márcia Da Ros Wendland |
| EPI_ISL_1469737 | CENTRO DE REFERENCIA EM SINDROMES GRIPAIS | Epiclin | Fernando Hayashi Sant'Anna, Ana Paula Muterle, Janira Prichula, Juliana Comerlato, Carolina Comerlato, Eliana Márcia Da Ros Wendland |
| EPI_ISL_1469740 | SECRETARIA MUNICIPAL DE SAUDE DE TRES COROAS | Epiclin | Fernando Hayashi Sant'Anna, Ana Paula Muterle, Janira Prichula, Juliana Comerlato, Carolina Comerlato, Eliana Márcia Da Ros Wendland |
| EPI_ISL_1469746 | HOSPITAL MONTENEGRO | Epiclin | Fernando Hayashi Sant'Anna, Ana Paula Muterle, Janira Prichula, Juliana Comerlato, Carolina Comerlato, Eliana Márcia Da Ros Wendland |
| EPI_ISL_1469748 | Diretoria de Vigilância em Saúde | Epiclin | Fernando Hayashi Sant'Anna, Ana Paula Muterle, Janira Prichula, Juliana Comerlato, Carolina Comerlato, Eliana Márcia Da Ros Wendland |
| EPI_ISL_1469749 | Centro de Referência em Síndromes Gripais | Epiclin | Fernando Hayashi Sant'Anna, Ana Paula Muterle, Janira Prichula, Juliana Comerlato, Carolina Comerlato, Eliana Márcia Da Ros Wendland |
| EPI_ISL_1469754 | Hospital Municipal Getúlio Vargas | Epiclin | Fernando Hayashi Sant'Anna, Ana Paula Muterle, Janira Prichula, Juliana Comerlato, Carolina Comerlato, Eliana Márcia Da Ros Wendland |
| EPI_ISL_1469774 | Hospital Universitário de Canoas | Epiclin | Fernando Hayashi Sant'Anna, Ana Paula Muterle, Janira Prichula, Juliana Comerlato, Carolina Comerlato, Eliana Márcia Da Ros Wendland |
| EPI_ISL_1469778 | Hospital Municipal Getúlio Vargas | Epiclin | Fernando Hayashi Sant'Anna, Ana Paula Muterle, Janira Prichula, Juliana Comerlato, Carolina Comerlato, Eliana Márcia Da Ros Wendland |
| EPI_ISL_1469781 | DIRETORIA DE VIGILANCIA EM SAUDE | Epiclin | Fernando Hayashi Sant'Anna, Ana Paula Muterle, Janira Prichula, Juliana Comerlato, Carolina Comerlato, Eliana Márcia Da Ros Wendland |
| EPI_ISL_1469783, EPI_ISL_1469792, EPI_ISL_1469796, EPI_ISL_1469802 | Diretoria de Vigilância em Saúde | Epiclin | Fernando Hayashi Sant'Anna, Ana Paula Muterle, Janira Prichula, Juliana Comerlato, Carolina Comerlato, Eliana Márcia Da Ros Wendland |
| EPI_ISL_1469803 | Secretaria Municipal de Saúde de Três Coroas | Epiclin | Fernando Hayashi Sant'Anna, Ana Paula Muterle, Janira Prichula, Juliana Comerlato, Carolina Comerlato, Eliana Márcia Da Ros Wendland |
| EPI_ISL_1469808, EPI_ISL_1469823, EPI_ISL_1469835 | Diretoria de Vigilância em Saúde | Epiclin | Fernando Hayashi Sant'Anna, Ana Paula Muterle, Janira Prichula, Juliana Comerlato, Carolina Comerlato, Eliana Márcia Da Ros Wendland |
| EPI_ISL_1469845 | Fundação Hospitalar de Sapucaia do Sul | Epiclin | Fernando Hayashi Sant'Anna, Ana Paula Muterle, Janira Prichula, Juliana Comerlato, Carolina Comerlato, Eliana Márcia Da Ros Wendland |
| EPI_ISL_1479121 | DIRETORIA DE VIGILANCIA EM SAUDE | Epiclin | Fernando Hayashi Sant'Anna, Ana Paula Muterle, Janira Prichula, Juliana Comerlato, Carolina Comerlato, Eliana Márcia Da Ros Wendland |
| EPI_ISL_1493590 | Hospital Sao Marcos da Samamorro Agudo | Instituto Adolfo Lutz, Interdiciplinary Procedures Center, Strategic Laboratory | Claudio Tavares Sacchi, Claudia Regina Gonçalves, Erica Valessa Ramos Gomes, Karoline Rodrigues Campos, Caio Vinicius Dias Lopes |
| EPI_ISL_1493591 | Centro de Saude II Dr Alcides Facundo Arroyo | Instituto Adolfo Lutz, Interdiciplinary Procedures Center, Strategic Laboratory | Claudio Tavares Sacchi, Claudia Regina Gonçalves, Erica Valessa Ramos Gomes, Karoline Rodrigues Campos, Caio Vinicius Dias Lopes |
| EPI_ISL_1493592 | Santa Casa de Guaira | Instituto Adolfo Lutz, Interdiciplinary Procedures Center, Strategic Laboratory | Claudio Tavares Sacchi, Claudia Regina Gonçalves, Erica Valessa Ramos Gomes, Karoline Rodrigues Campos, Caio Vinicius Dias Lopes |
| EPI_ISL_1494970, EPI_ISL_1495004 | Laboratório de Biologia Integrativa | Laboratório de Biologia Integrativa | Filipe Romero Rebello Moreira, Diego Menezes Bonfim, Victor Emmanuel Viana Geddes, Danielle Alves Gomes Zauli, Joice do Prado Silva, Aline Brito de Lima, Frederico Scott Varella Malta, Alessandro Clayton de Souza Ferreira, Victor Cavalcanti Pardini, Daniel Costa Queiroz, Rafael Marques de Souza, Lucyene Miguita Luiz, Paula Luize Camargos Fonseca, Rennan Garcias Moreira, Nuno Rodrigues Faria, Carolina Moreira Voloch, Renan Pedra de Souza, Renato Santana Aguiar |
| EPI_ISL_1499020, EPI_ISL_1499114, EPI_ISL_1499297 | Associação Fundo de Incentivo à Pesquisa (AFIP) | Associação Fundo de Incentivo à Pesquisa (AFIP) | Priscila Farias Tempaku, Juliana Nogueira Martins Rodrigues, Erika Rodrigues de Oliveira, Debora R. Ramadan, Soraya Sgambatti de Andrade, Sergio Tufik. |
| EPI_ISL_1511641 | Laboratorio de Patologia Clinica - UNICAMP | Laboratorio de Estudos de Virus Emergentes | Mariene R. Amorim, William M. Souza, Antonio C. G. Carlos Jr, Daniel A. Toledo-Teixeira, Karina Bispo-dos-Santos, Camila L. Simeoni, Pierina L. Parise, Aline Vieira, Julia Forato, Ingra M. Claro, Luciana S. Mofatto, Natalia S. Brunetti, Emerson S.S. França, Gisele A. Pedroso, Barbara F. N. Carvalho, Tania R. Zaccariotto, Kamila C. S. Krywacz, André S. Vieira, Marcelo A. Mori, Alessandro S. Farias, Maria H. P. Pavan, Luís Felipe Bachur, Luís G. O. Cardoso, Fernando R. Spilki, Ester C. Sabino, Nuno R. Faria, Magnun N. N. Santos, Rodrigo Angerami, Patricia A. F. Leme, Angelica Schreiber, Maria L. Moretti, Fabiana Granja, José Luiz Proenca-Modena |
| EPI_ISL_1520110 | Hospital Municipal Reynaldo Guerra Cajati | Instituto Adolfo Lutz, Interdiciplinary Procedures Center, Strategic Laboratory | Claudio Tavares Sacchi, Claudia Regina Gonçalves, Erica Valessa Ramos Gomes, Karoline Rodrigues Campos, Caio Vinicius Dias Lopes |

| | | | |
|---|---|---|---|
| EPI_ISL_1520134, EPI_ISL_1520135 | Centro de Saude II Dr Jose Paione Mococa | Instituto Adolfo Lutz, Interdiciplinary Procedures Center, Strategic Laboratory | Claudio Tavares Sacchi, Claudia Regina Gonçalves, Erica Valessa Ramos Gomes, Karoline Rodrigues Campos, Caio Vinicius Dias Lopes |
| EPI_ISL_1533691 | Centro de Saude II Dr. Jose Paione Mococa | Instituto Adolfo Lutz, Interdiciplinary Procedures Center, Strategic Laboratory | Claudio Tavares Sacchi, Claudia Regina Gonçalves, Erica Valessa Ramos Gomes, Karoline Rodrigues Campos, Caio Vinicius Dias Lopes, Leonardo Jose Tadeu de Araujo |
| EPI_ISL_1533692 | Santa Casa de Sao Paulo | Instituto Adolfo Lutz, Interdiciplinary Procedures Center, Strategic Laboratory | Claudio Tavares Sacchi, Claudia Regina Gonçalves, Erica Valessa Ramos Gomes, Karoline Rodrigues Campos, Caio Vinicius Dias Lopes, Leonardo Jose Tadeu de Araujo |
| EPI_ISL_1533697 | Hospital Estadual de Vila Alpina | Instituto Adolfo Lutz, Interdiciplinary Procedures Center, Strategic Laboratory | Claudio Tavares Sacchi, Claudia Regina Gonçalves, Erica Valessa Ramos Gomes, Karoline Rodrigues Campos, Caio Vinicius Dias Lopes, Leonardo Jose Tadeu de Araujo |
| EPI_ISL_1533724 | Diretoria Municipal de Saude | Instituto Adolfo Lutz, Interdiciplinary Procedures Center, Strategic Laboratory | Claudio Tavares Sacchi, Claudia Regina Gonçalves, Erica Valessa Ramos Gomes, Karoline Rodrigues Campos, Caio Vinicius Dias Lopes, Leonardo Jose Tadeu de Araujo |
| EPI_ISL_1580504 | Laboratório de Biologia Molecular do Hospital das Clínicas da Faculdade de Medicina de Botucatu/SP | Laboratórios de Genômica Funcional (FCA/UNESP) e Biologia Molecular (FMB-HC/UNESP) - Rede de Vigilância Genômica (Vigenômica)/UNESP | Patrícia Akemi Assato; Felipe Allan da Silva da Costa; Bianca Cechetto Carlos; Flavia Hebner Barbosa Trovão; Guilherme Targino Valente; Rejane Maria Tommasini Grotto; Jayme A. Souza-Neto. |
| EPI_ISL_1583642, EPI_ISL_1583643, EPI_ISL_1583648, EPI_ISL_1583649, EPI_ISL_1583651, EPI_ISL_1583657, EPI_ISL_1583658, EPI_ISL_1583659, EPI_ISL_1583660, EPI_ISL_1583668, EPI_ISL_1583671, EPI_ISL_1583686, EPI_ISL_1583727 | | | |
| see above | Central Public Health Laboratory - LACEN -Bahia, Salvador, Brazil | Central Public Health Laboratory - LACEN -Bahia, Salvador, Brazil | Stephane Tosta, Luciana Oliveira, Vanessa Nardy,Patrícia Cajado,Marcela Gómez, Breno Dominguez, Jaqueline Gomes, Vagner Fonseca,Marta Giovanetti,Luiz Alcantara, Felicidade Pereira, Arabela Leal |
| EPI_ISL_1625985, EPI_ISL_1625996, EPI_ISL_1626008, EPI_ISL_1628346 | Instituto Adolfo Lutz - Regional de Rio Claro | Instituto Adolfo Lutz, Interdiciplinary Procedures Center, Strategic Laboratory | Claudio Tavares Sacchi, Claudia Regina Gonçalves, Erica Valessa Ramos Gomes, Karoline Rodrigues Campos, Caio Vinicius Dias Lopes, Leonardo Jose Tadeu de Araujo, Katia Correa de Oliveira Santos |
| EPI_ISL_1661251 | Laboratorio de Ecologia de Doencas Transmissiveis na Amazonia, Instituto Leonidas e Maria Deane - Fiocruz Amazonia | Laboratorio de Ecologia de Doencas Transmissiveis na Amazonia, Instituto Leonidas e Maria Deane - Fiocruz Amazonia | Valdinete Nascimento, Victor Souza, André Corado, Fernanda Nascimento, George Silva, Ágatha Costa, Debora Duarte, Karina Pessoa, Matilde Mejía, Luciana Gonçalves, Maria Júlia Brandão, Michele Jesus, Felipe Naveca |
| EPI_ISL_1731593, EPI_ISL_1731606 | Instituto Adolfo Lutz Central | Instituto Adolfo Lutz, Interdiciplinary Procedures Center, Strategic Laboratory | Claudio Tavares Sacchi, Claudia Regina Gonçalves, Erica Valessa Ramos Gomes, Karoline Rodrigues Campos, Caio Vinicius Dias Lopes, Leonardo Jose Tadeu de Araujo, Katia Correa de Oliveira Santos |
| EPI_ISL_1785610, EPI_ISL_1785612 | Laboratório de Pesquisa em Virologia, FAMERP, SJRP | Laboratório de Pesquisa em Virologia, FAMERP, SJRP | Fábio Sossai Possebon; Leila Sabrina Ullmann; Cecília Artico Banho; Cíntia Bittar; Guilherme Campos; Helena Lage Ferreira; Jorge A. Petroli Marchesi; Lívia Sacchetto; Maisa C. Pereira Parra; Marília Moraes; Maurício L. Nogueira; Paula Rahal; Paulo Inacio da Costa; João Pessoa Araújo Jr. |
| EPI_ISL_1795398 | SMS SECRETARIA MUNICIPAL DE SAUDE DE BOITUVA | Instituto Butantan / ESALQ-Piracicaba | Instituto Butantan: Alexander Roberto Precioso, Dimas Tadeu Covas, Sandra Coccuzzo Sampaio, Maria Carolina Elias, José Salvatore Leister Patané, Vincent Louis Viala, Antonio Jorge Martins, Ricardo Haddad, Claudia Renata dos Santos Barros, Elaine Cristina Marqueze, Raul Machado Neto, Debora Botequio Moretti. Centro de Genômica Funcional da ESALQ: Luiz Lehmann Coutinho, Ricardo Augusto Brassaloti, Raquel de Lello Rocha Campos Cassano. NGS Soluções Genômicas: Pilar Drummond Sampaio Corrêa Mariani. FZEA-USP Pirassununga: Mirele Daiana Poleti, Jessika Cristina Chagas Lesbon, Elisangela Chicaroni Mattos, Heidge Fukumasu. USP-Botucatu: Rejane Maria Tommasini Grotto, Jayme A. Souza-Neto, Guilherme Targino Valente, Patricia Akemi Assato, Felipe Allan da Silva da Costa, Bianca Cechetto Carlos. Mendelics: Bibiana Santos, João Paulo Kitajima, Erika Freitas, David Schlesinger. Hemocentro Ribeirão Preto: Simone Kashima, Evandra Strazza Rodrigues, Svetoslav Nanev Slavov, Elaine Vieira dos Santos, Rafael dos Santos Bezerra, Luiz Carlos Junior de Alcantara, Marta Giovanetti, Vagner Fonseca, Flavia Aburjaile, Rodrigo Tocantins Calado. |
| EPI_ISL_1795399 | POLICLINICA HORTOLANDIA | Instituto Butantan / ESALQ-Piracicaba | Instituto Butantan: Alexander Roberto Precioso, Dimas Tadeu Covas, Sandra Coccuzzo Sampaio, Maria Carolina Elias, José Salvatore Leister Patané, Vincent Louis Viala, Antonio Jorge Martins, Ricardo Haddad, Claudia Renata dos Santos Barros, Elaine Cristina Marqueze, Raul Machado Neto, Debora Botequio Moretti. Centro de Genômica Funcional da ESALQ: Luiz Lehmann Coutinho, Ricardo Augusto Brassaloti, Raquel de Lello Rocha Campos Cassano. NGS Soluções Genômicas: Pilar Drummond Sampaio Corrêa Mariani. FZEA-USP Pirassununga: Mirele Daiana Poleti, Jessika Cristina Chagas Lesbon, Elisangela Chicaroni Mattos, Heidge Fukumasu. USP-Botucatu: Rejane Maria Tommasini Grotto, Jayme A. Souza-Neto, Guilherme Targino Valente, Patricia Akemi Assato, Felipe Allan da Silva da Costa, Bianca Cechetto Carlos. Mendelics: Bibiana Santos, João Paulo Kitajima, Erika Freitas, David Schlesinger. Hemocentro Ribeirão Preto: Simone Kashima, Evandra Strazza Rodrigues, Svetoslav Nanev Slavov, Elaine Vieira dos Santos, Rafael dos Santos Bezerra, Luiz Carlos Junior de Alcantara, Marta Giovanetti, Vagner Fonseca, Flavia Aburjaile, Rodrigo Tocantins Calado. |
| EPI_ISL_1795414 | HOSPITAL MUNICIPAL DE IBIUNA IBIUNA SP | Instituto Butantan / ESALQ-Piracicaba | Instituto Butantan: Alexander Roberto Precioso, Dimas Tadeu Covas, Sandra Coccuzzo Sampaio, Maria Carolina Elias, José Salvatore Leister Patané, Vincent Louis Viala, Antonio Jorge Martins, Ricardo Haddad, Claudia Renata dos Santos Barros, Elaine Cristina Marqueze, Raul Machado Neto, Debora Botequio Moretti. Centro de Genômica Funcional da ESALQ: Luiz Lehmann Coutinho, Ricardo Augusto Brassaloti, Raquel de Lello Rocha Campos Cassano. NGS Soluções Genômicas: Pilar Drummond Sampaio Corrêa Mariani. FZEA-USP Pirassununga: Mirele Daiana Poleti, Jessika Cristina Chagas Lesbon, Elisangela Chicaroni Mattos, Heidge Fukumasu. USP-Botucatu: Rejane Maria Tommasini Grotto, Jayme A. Souza-Neto, Guilherme Targino Valente, Patricia Akemi Assato, Felipe Allan da Silva da Costa, Bianca Cechetto Carlos. Mendelics: Bibiana Santos, João Paulo Kitajima, Erika Freitas, David Schlesinger. Hemocentro Ribeirão Preto: Simone Kashima, Evandra Strazza Rodrigues, Svetoslav Nanev Slavov, Elaine Vieira dos Santos, Rafael dos Santos Bezerra, Luiz Carlos Junior de Alcantara, Marta Giovanetti, Vagner Fonseca, Flavia Aburjaile, Rodrigo Tocantins Calado. |
| EPI_ISL_1795418, EPI_ISL_1795420, EPI_ISL_1795421, EPI_ISL_1795422 | LABORATORIO DE FRANCA | Instituto Butantan / ESALQ-Piracicaba | Instituto Butantan: Alexander Roberto Precioso, Dimas Tadeu Covas, Sandra Coccuzzo Sampaio, Maria Carolina Elias, José Salvatore Leister Patané, Vincent Louis Viala, Antonio Jorge Martins, Ricardo Haddad, Claudia Renata dos Santos Barros, Elaine Cristina Marqueze, Raul Machado Neto, Debora Botequio Moretti. Centro de Genômica Funcional da ESALQ: Luiz Lehmann Coutinho, Ricardo Augusto Brassaloti, Raquel de Lello Rocha Campos Cassano. NGS Soluções Genômicas: Pilar Drummond Sampaio Corrêa Mariani. FZEA-USP Pirassununga: Mirele Daiana Poleti, Jessika Cristina Chagas Lesbon, Elisangela Chicaroni Mattos, Heidge Fukumasu. USP-Botucatu: Rejane Maria Tommasini Grotto, Jayme A. Souza-Neto, Guilherme Targino Valente, Patricia Akemi Assato, Felipe Allan da Silva da Costa, Bianca Cechetto Carlos. Mendelics: Bibiana Santos, João Paulo Kitajima, Erika Freitas, David Schlesinger. Hemocentro Ribeirão Preto: Simone Kashima, Evandra Strazza Rodrigues, Svetoslav Nanev Slavov, Elaine Vieira dos Santos, Rafael dos Santos Bezerra, Luiz Carlos Junior de Alcantara, Marta Giovanetti, Vagner Fonseca, Flavia Aburjaile, Rodrigo Tocantins Calado. |
| EPI_ISL_1799499, EPI_ISL_1799505 | Laboratório de Microbiologia Molecular - Universidade FEEVALE | Molecular Microbiology Laboratory | Alana Witt Hansen, Fágner Henrique Heldt, Fernando Rosado Spilki, Flávio Silveira, Juliana Schons Gulart, Juliane Deise Fleck, Mariana Soares da Silva, Meriane Demoliner, Matheus Nunes Weber, Paula Rodrigues de Almeida, Michele Filippi. |
| EPI_ISL_1821229, EPI_ISL_1821231, EPI_ISL_1821238 | Instituto Adolfo Lutz - Regional de Marilia | Instituto Adolfo Lutz, Interdiciplinary Procedures Center, Strategic Laboratory | Claudio Tavares Sacchi, Claudia Regina Gonçalves, Erica Valessa Ramos Gomes, Karoline Rodrigues Campos, Caio Vinicius Dias Lopes, Leonardo Jose Tadeu de Araujo |
| EPI_ISL_1966067 | VIGILANCIA EPIDEMIOLOGICA | Instituto Butantan / Mendelics | Instituto Butantan: Dimas Tadeu Covas, Sandra Coccuzzo Sampaio, Maria Carolina Elias, José Salvatore Leister Patané, Vincent Louis Viala, Antonio Jorge Martins, Ricardo Haddad, Claudia Renata dos Santos Barros, Elaine Cristina Marqueze, Raul Machado Neto, Debora Botequio Moretti, Jardelina de Souza Todao Bernardino, Loyze Paola Oliveira de Lima, Luiz Aurelio de Campos Crispin. Centro de Genômica Funcional da ESALQ: Luiz Lehmann Coutinho, Ricardo Augusto Brassaloti, Raquel de Lello Rocha Campos Cassano. NGS Soluções Genômicas: Pilar Drummond Sampaio Corrêa Mariani. FZEA-USP Pirassununga: Mirele Daiana Poleti, Jessika Cristina Chagas Lesbon, Elisangela Chicaroni Mattos, Heidge Fukumasu. USP-Botucatu: Rejane Maria Tommasini Grotto, Jayme A. Souza-Neto, Guilherme Targino Valente, Patricia Akemi Assato, Felipe Allan da Silva da Costa, Bianca Cechetto Carlos. Mendelics: Bibiana Santos, João Paulo Kitajima, Erika Freitas, David Schlesinger. Hemocentro Ribeirão Preto: Simone Kashima, Evandra Strazza Rodrigues, Svetoslav Nanev Slavov, Elaine Vieira dos Santos, Rafael dos Santos Bezerra, Luiz Carlos Junior de Alcantara, Marta Giovanetti, Vagner Fonseca, Flavia Aburjaile, Rodrigo Tocantins Calado. FAMERP-SJRP: Cecília Artico Banho, Lívia Sacchetto, Fábio Sossai Possebon, Leila Sabrina Ullmann, Cíntia Bittar, Guilherme Campos, Helena Lage Ferreira, Jorge A. Petroli Marchesi, Maisa C. Pereira Parra, Marília Moraes, Paula Rahal, Paulo Inacio da Costa, João Pessoa Araújo Jr., Maurício Lacerda Nogueira. Prefeitura de Sao Paulo: Melissa Palmieri. |
| EPI_ISL_1966073 | SECAO CENTRO DE DIAGNOSTICO SECEDI | Instituto Butantan / Mendelics | Instituto Butantan: Dimas Tadeu Covas, Sandra Coccuzzo Sampaio, Maria Carolina Elias, José Salvatore Leister Patané, Vincent Louis Viala, Antonio Jorge Martins, Ricardo Haddad, Claudia Renata dos Santos Barros, Elaine Cristina Marqueze, Raul Machado Neto, Debora Botequio Moretti, Jardelina de Souza Todao Bernardino, Loyze Paola Oliveira de Lima, Luiz Aurelio de Campos Crispin. Centro de Genômica Funcional da ESALQ: Luiz Lehmann Coutinho, Ricardo Augusto Brassaloti, Raquel de Lello Rocha Campos Cassano. NGS Soluções Genômicas: Pilar Drummond Sampaio Corrêa Mariani. FZEA-USP Pirassununga: Mirele Daiana Poleti, Jessika Cristina Chagas Lesbon, Elisangela Chicaroni Mattos, Heidge Fukumasu. USP-Botucatu: Rejane Maria Tommasini Grotto, Jayme A. Souza-Neto, Guilherme Targino Valente, Patricia Akemi Assato, Felipe Allan da Silva da Costa, Bianca Cechetto |

| | | | |
|---|---|---|---|
| | | | Carlos. Mendelics: Bibiana Santos, João Paulo Kitajima, Erika Freitas, David Schlesinger. Hemocentro Ribeirão Preto: Simone Kashima, Evandra Strazza Rodrigues, Svetoslav Nanev Slavov, Elaine Vieira dos Santos, Rafael dos Santos Bezerra, Luiz Carlos Junior de Alcantara, Marta Giovanetti, Vagner Fonseca, Flavia Aburjaile, Rodrigo Tocantins Calado. FAMERP-SJRP: Cecília Artico Banho, Lívia Sacchetto, Fábio Sossai Possebon, Leila Sabrina Ullmann, Cíntia Bittar, Guilherme Campos, Helena Lage Ferreira, Jorge A. Petroli Marchesi, Maisa C. Pereira Parra, Marília Moraes, Paula Rahal, Paulo Inacio da Costa, João Pessoa Araújo Jr., Maurício Lacerda Nogueira. Prefeitura de Sao Paulo: Melissa Palmieri. |
| EPI_ISL_1966091 | VIGILANCIA EPIDEMIOLOGICA | Instituto Butantan / Mendelics | Instituto Butantan: Dimas Tadeu Covas, Sandra Coccuzzo Sampaio, Maria Carolina Elias, José Salvatore Leister Patané, Vincent Louis Viala, Antonio Jorge Martins, Ricardo Haddad, Claudia Renata dos Santos Barros, Elaine Cristina Marqueze, Raul Machado Neto, Debora Botequio Moretti, Jardelina de Souza Todao Bernardino, Loyze Paola Oliveira de Lima, Luiz Aurelio de Campos Crispin. Centro de Genômica Funcional da ESALQ: Luiz Lehmann Coutinho, Ricardo Augusto Brassaloti, Raquel de Lello Rocha Campos Cassano. NGS Soluções Genômicas: Pilar Drummond Sampaio Corrêa Mariani. FZEA-USP Pirassununga: Mirele Daiana Poleti, Jessika Cristina Chagas Lesbon, Elisangela Chicaroni Mattos, Heidge Fukumasu. USP-Botucatu: Rejane Maria Tommasini Grotto, Jayme A. Souza-Neto, Guilherme Targino Valente, Patricia Akemi Assato, Felipe Allan da Silva da Costa, Bianca Cechetto Carlos. Mendelics: Bibiana Santos, João Paulo Kitajima, Erika Freitas, David Schlesinger. Hemocentro Ribeirão Preto: Simone Kashima, Evandra Strazza Rodrigues, Svetoslav Nanev Slavov, Elaine Vieira dos Santos, Rafael dos Santos Bezerra, Luiz Carlos Junior de Alcantara, Marta Giovanetti, Vagner Fonseca, Flavia Aburjaile, Rodrigo Tocantins Calado. FAMERP-SJRP: Cecília Artico Banho, Lívia Sacchetto, Fábio Sossai Possebon, Leila Sabrina Ullmann, Cíntia Bittar, Guilherme Campos, Helena Lage Ferreira, Jorge A. Petroli Marchesi, Maisa C. Pereira Parra, Marília Moraes, Paula Rahal, Paulo Inacio da Costa, João Pessoa Araújo Jr., Maurício Lacerda Nogueira. Prefeitura de Sao Paulo: Melissa Palmieri. |
| EPI_ISL_1966104 | POLICLINICA COVID 19 ITAPETININGA | Instituto Butantan / Mendelics | Instituto Butantan: Dimas Tadeu Covas, Sandra Coccuzzo Sampaio, Maria Carolina Elias, José Salvatore Leister Patané, Vincent Louis Viala, Antonio Jorge Martins, Ricardo Haddad, Claudia Renata dos Santos Barros, Elaine Cristina Marqueze, Raul Machado Neto, Debora Botequio Moretti, Jardelina de Souza Todao Bernardino, Loyze Paola Oliveira de Lima, Luiz Aurelio de Campos Crispin. Centro de Genômica Funcional da ESALQ: Luiz Lehmann Coutinho, Ricardo Augusto Brassaloti, Raquel de Lello Rocha Campos Cassano. NGS Soluções Genômicas: Pilar Drummond Sampaio Corrêa Mariani. FZEA-USP Pirassununga: Mirele Daiana Poleti, Jessika Cristina Chagas Lesbon, Elisangela Chicaroni Mattos, Heidge Fukumasu. USP-Botucatu: Rejane Maria Tommasini Grotto, Jayme A. Souza-Neto, Guilherme Targino Valente, Patricia Akemi Assato, Felipe Allan da Silva da Costa, Bianca Cechetto Carlos. Mendelics: Bibiana Santos, João Paulo Kitajima, Erika Freitas, David Schlesinger. Hemocentro Ribeirão Preto: Simone Kashima, Evandra Strazza Rodrigues, Svetoslav Nanev Slavov, Elaine Vieira dos Santos, Rafael dos Santos Bezerra, Luiz Carlos Junior de Alcantara, Marta Giovanetti, Vagner Fonseca, Flavia Aburjaile, Rodrigo Tocantins Calado. FAMERP-SJRP: Cecília Artico Banho, Lívia Sacchetto, Fábio Sossai Possebon, Leila Sabrina Ullmann, Cíntia Bittar, Guilherme Campos, Helena Lage Ferreira, Jorge A. Petroli Marchesi, Maisa C. Pereira Parra, Marília Moraes, Paula Rahal, Paulo Inacio da Costa, João Pessoa Araújo Jr., Maurício Lacerda Nogueira. Prefeitura de Sao Paulo: Melissa Palmieri. |
| EPI_ISL_1966131 | CENTRO DE SAUDE II MAIRINQUE MAIRINQUE | Instituto Butantan / Mendelics | Instituto Butantan: Dimas Tadeu Covas, Sandra Coccuzzo Sampaio, Maria Carolina Elias, José Salvatore Leister Patané, Vincent Louis Viala, Antonio Jorge Martins, Ricardo Haddad, Claudia Renata dos Santos Barros, Elaine Cristina Marqueze, Raul Machado Neto, Debora Botequio Moretti, Jardelina de Souza Todao Bernardino, Loyze Paola Oliveira de Lima, Luiz Aurelio de Campos Crispin. Centro de Genômica Funcional da ESALQ: Luiz Lehmann Coutinho, Ricardo Augusto Brassaloti, Raquel de Lello Rocha Campos Cassano. NGS Soluções Genômicas: Pilar Drummond Sampaio Corrêa Mariani. FZEA-USP Pirassununga: Mirele Daiana Poleti, Jessika Cristina Chagas Lesbon, Elisangela Chicaroni Mattos, Heidge Fukumasu. USP-Botucatu: Rejane Maria Tommasini Grotto, Jayme A. Souza-Neto, Guilherme Targino Valente, Patricia Akemi Assato, Felipe Allan da Silva da Costa, Bianca Cechetto Carlos. Mendelics: Bibiana Santos, João Paulo Kitajima, Erika Freitas, David Schlesinger. Hemocentro Ribeirão Preto: Simone Kashima, Evandra Strazza Rodrigues, Svetoslav Nanev Slavov, Elaine Vieira dos Santos, Rafael dos Santos Bezerra, Luiz Carlos Junior de Alcantara, Marta Giovanetti, Vagner Fonseca, Flavia Aburjaile, Rodrigo Tocantins Calado. FAMERP-SJRP: Cecília Artico Banho, Lívia Sacchetto, Fábio Sossai Possebon, Leila Sabrina Ullmann, Cíntia Bittar, Guilherme Campos, Helena Lage Ferreira, Jorge A. Petroli Marchesi, Maisa C. Pereira Parra, Marília Moraes, Paula Rahal, Paulo Inacio da Costa, João Pessoa Araújo Jr., Maurício Lacerda Nogueira. Prefeitura de Sao Paulo: Melissa Palmieri. |
| EPI_ISL_1966178 | SECRETARIA MUNICIPAL DE SAUDE SOROCABA | Instituto Butantan / Mendelics | Instituto Butantan: Dimas Tadeu Covas, Sandra Coccuzzo Sampaio, Maria Carolina Elias, José Salvatore Leister Patané, Vincent Louis Viala, Antonio Jorge Martins, Ricardo Haddad, Claudia Renata dos Santos Barros, Elaine Cristina Marqueze, Raul Machado Neto, Debora Botequio Moretti, Jardelina de Souza Todao Bernardino, Loyze Paola Oliveira de Lima, Luiz Aurelio de Campos Crispin. Centro de Genômica Funcional da ESALQ: Luiz Lehmann Coutinho, Ricardo Augusto Brassaloti, Raquel de Lello Rocha Campos Cassano. NGS Soluções Genômicas: Pilar Drummond Sampaio Corrêa Mariani. FZEA-USP Pirassununga: Mirele Daiana Poleti, Jessika Cristina Chagas Lesbon, Elisangela Chicaroni Mattos, Heidge Fukumasu. USP-Botucatu: Rejane Maria Tommasini Grotto, Jayme A. Souza-Neto, Guilherme Targino Valente, Patricia Akemi Assato, Felipe Allan da Silva da Costa, Bianca Cechetto Carlos. Mendelics: Bibiana Santos, João Paulo Kitajima, Erika Freitas, David Schlesinger. Hemocentro Ribeirão Preto: Simone Kashima, Evandra Strazza Rodrigues, Svetoslav Nanev Slavov, Elaine Vieira dos Santos, Rafael dos Santos Bezerra, Luiz Carlos Junior de Alcantara, Marta Giovanetti, Vagner Fonseca, Flavia Aburjaile, Rodrigo Tocantins Calado. FAMERP-SJRP: Cecília Artico Banho, Lívia Sacchetto, Fábio Sossai Possebon, Leila Sabrina Ullmann, Cíntia Bittar, Guilherme Campos, Helena Lage Ferreira, Jorge A. Petroli Marchesi, Maisa C. Pereira Parra, Marília Moraes, Paula Rahal, Paulo Inacio da Costa, João Pessoa Araújo Jr., Maurício Lacerda Nogueira. Prefeitura de Sao Paulo: Melissa Palmieri. |
| EPI_ISL_1966219 | CENTRO DE SAUDE DE BORA | Instituto Butantan / Mendelics | Instituto Butantan: Dimas Tadeu Covas, Sandra Coccuzzo Sampaio, Maria Carolina Elias, José Salvatore Leister Patané, Vincent Louis Viala, Antonio Jorge Martins, Ricardo Haddad, Claudia Renata dos Santos Barros, Elaine Cristina Marqueze, Raul Machado Neto, Debora Botequio Moretti, Jardelina de Souza Todao Bernardino, Loyze Paola Oliveira de Lima, Luiz Aurelio de Campos Crispin. Centro de Genômica Funcional da ESALQ: Luiz Lehmann Coutinho, Ricardo Augusto Brassaloti, Raquel de Lello Rocha Campos Cassano. NGS Soluções Genômicas: Pilar Drummond Sampaio Corrêa Mariani. FZEA-USP Pirassununga: Mirele Daiana Poleti, Jessika Cristina Chagas Lesbon, Elisangela Chicaroni Mattos, Heidge Fukumasu. USP-Botucatu: Rejane Maria Tommasini Grotto, Jayme A. Souza-Neto, Guilherme Targino Valente, Patricia Akemi Assato, Felipe Allan da Silva da Costa, Bianca Cechetto Carlos. Mendelics: Bibiana Santos, João Paulo Kitajima, Erika Freitas, David Schlesinger. Hemocentro Ribeirão Preto: Simone Kashima, Evandra Strazza Rodrigues, Svetoslav Nanev Slavov, Elaine Vieira dos Santos, Rafael dos Santos Bezerra, Luiz Carlos Junior de Alcantara, Marta Giovanetti, Vagner Fonseca, Flavia Aburjaile, Rodrigo Tocantins Calado. FAMERP-SJRP: Cecília Artico Banho, Lívia Sacchetto, Fábio Sossai Possebon, Leila Sabrina Ullmann, Cíntia Bittar, Guilherme Campos, Helena Lage Ferreira, Jorge A. Petroli Marchesi, Maisa C. Pereira Parra, Marília Moraes, Paula Rahal, Paulo Inacio da Costa, João Pessoa Araújo Jr., Maurício Lacerda Nogueira. Prefeitura de Sao Paulo: Melissa Palmieri. |
| EPI_ISL_1966241 | CENTRO DE SAUDE DR MARIO DIAS DE AGUIAR CAPIVARI | Instituto Butantan / Mendelics | Instituto Butantan: Dimas Tadeu Covas, Sandra Coccuzzo Sampaio, Maria Carolina Elias, José Salvatore Leister Patané, Vincent Louis Viala, Antonio Jorge Martins, Ricardo Haddad, Claudia Renata dos Santos Barros, Elaine Cristina Marqueze, Raul Machado Neto, Debora Botequio Moretti, Jardelina de Souza Todao Bernardino, Loyze Paola Oliveira de Lima, Luiz Aurelio de Campos Crispin. Centro de Genômica Funcional da ESALQ: Luiz Lehmann Coutinho, Ricardo Augusto Brassaloti, Raquel de Lello Rocha Campos Cassano. NGS Soluções Genômicas: Pilar Drummond Sampaio Corrêa Mariani. FZEA-USP Pirassununga: Mirele Daiana Poleti, Jessika Cristina Chagas Lesbon, Elisangela Chicaroni Mattos, Heidge Fukumasu. USP-Botucatu: Rejane Maria Tommasini Grotto, Jayme A. Souza-Neto, Guilherme Targino Valente, Patricia Akemi Assato, Felipe Allan da Silva da Costa, Bianca Cechetto Carlos. Mendelics: Bibiana Santos, João Paulo Kitajima, Erika Freitas, David Schlesinger. Hemocentro Ribeirão Preto: Simone Kashima, Evandra Strazza Rodrigues, Svetoslav Nanev Slavov, Elaine Vieira dos Santos, Rafael dos Santos Bezerra, Luiz Carlos Junior de Alcantara, Marta Giovanetti, Vagner Fonseca, Flavia Aburjaile, Rodrigo Tocantins Calado. FAMERP-SJRP: Cecília Artico Banho, Lívia Sacchetto, Fábio Sossai Possebon, Leila Sabrina Ullmann, Cíntia Bittar, Guilherme Campos, Helena Lage Ferreira, Jorge A. Petroli Marchesi, Maisa C. Pereira Parra, Marília Moraes, Paula Rahal, Paulo Inacio da Costa, João Pessoa Araújo Jr., Maurício Lacerda Nogueira. Prefeitura de Sao Paulo: Melissa Palmieri. |
| EPI_ISL_1966261 | SMS SECRETARIA MUNICIPAL DE SAUDE DE BOITUVA | Instituto Butantan / Mendelics | Instituto Butantan: Dimas Tadeu Covas, Sandra Coccuzzo Sampaio, Maria Carolina Elias, José Salvatore Leister Patané, Vincent Louis Viala, Antonio Jorge Martins, Ricardo Haddad, Claudia Renata dos Santos Barros, Elaine Cristina Marqueze, Raul Machado Neto, Debora Botequio Moretti, Jardelina de Souza Todao Bernardino, Loyze Paola Oliveira de Lima, Luiz Aurelio de Campos Crispin. Centro de Genômica Funcional da ESALQ: Luiz Lehmann Coutinho, Ricardo Augusto Brassaloti, Raquel de Lello Rocha Campos Cassano. NGS Soluções Genômicas: Pilar Drummond Sampaio Corrêa Mariani. FZEA-USP Pirassununga: Mirele Daiana Poleti, Jessika Cristina Chagas Lesbon, Elisangela Chicaroni Mattos, Heidge Fukumasu. USP-Botucatu: Rejane Maria Tommasini Grotto, Jayme A. Souza-Neto, Guilherme Targino Valente, Patricia Akemi Assato, Felipe Allan da Silva da Costa, Bianca Cechetto Carlos. Mendelics: Bibiana Santos, João Paulo Kitajima, Erika Freitas, David Schlesinger. Hemocentro Ribeirão Preto: Simone Kashima, Evandra Strazza Rodrigues, Svetoslav Nanev Slavov, Elaine Vieira dos Santos, Rafael dos Santos Bezerra, Luiz Carlos Junior de Alcantara, Marta Giovanetti, Vagner Fonseca, Flavia Aburjaile, Rodrigo Tocantins Calado. FAMERP-SJRP: Cecília Artico Banho, Lívia Sacchetto, Fábio Sossai Possebon, Leila Sabrina Ullmann, Cíntia Bittar, Guilherme Campos, Helena Lage Ferreira, Jorge A. Petroli Marchesi, Maisa C. Pereira Parra, Marília Moraes, Paula Rahal, Paulo Inacio da Costa, João Pessoa Araújo Jr., Maurício Lacerda Nogueira. Prefeitura de Sao Paulo: Melissa Palmieri. |
| EPI_ISL_1966342 | UPA 24 HORAS CENTRO | Instituto Butantan / Mendelics | Instituto Butantan: Dimas Tadeu Covas, Sandra Coccuzzo Sampaio, Maria Carolina Elias, José Salvatore Leister Patané, Vincent Louis Viala, Antonio |

| | | | |
|---|---|---|---|
| | | | Jorge Martins, Ricardo Haddad, Claudia Renata dos Santos Barros, Elaine Cristina Marqueze, Raul Machado Neto, Debora Botequio Moretti, Jardelina de Souza Todao Bernardino, Loyze Paola Oliveira de Lima, Luiz Aurelio de Campos Crispin. Centro de Genômica Funcional da ESALQ: Luiz Lehmann Coutinho, Ricardo Augusto Brassaloti, Raquel de Lello Rocha Campos Cassano. NGS Soluções Genômicas: Pilar Drummond Sampaio Corrêa Mariani. FZEA-USP Pirassununga: Mirele Daiana Poleti, Jessika Cristina Chagas Lesbon, Elisangela Chicaroni Mattos, Heidge Fukumasu. USP-Botucatu: Rejane Maria Tommasini Grotto, Jayme A. Souza-Neto, Guilherme Targino Valente, Patricia Akemi Assato, Felipe Allan da Silva da Costa, Bianca Cechetto Carlos. Mendelics: Bibiana Santos, João Paulo Kitajima, Erika Freitas, David Schlesinger. Hemocentro Ribeirão Preto: Simone Kashima, Evandra Strazza Rodrigues, Svetoslav Nanev Slavov, Elaine Vieira dos Santos, Rafael dos Santos Bezerra, Luiz Carlos Junior de Alcantara, Marta Giovanetti, Vagner Fonseca, Flavia Aburjaile, Rodrigo Tocantins Calado. FAMERP-SJRP: Cecília Artico Banho, Lívia Sacchetto, Fábio Sossai Possebon, Leila Sabrina Ullmann, Cíntia Bittar, Guilherme Campos, Helena Lage Ferreira, Jorge A. Petroli Marchesi, Maisa C. Pereira Parra, Marília Moraes, Paula Rahal, Paulo Inacio da Costa, João Pessoa Araújo Jr., Maurício Lacerda Nogueira. Prefeitura de Sao Paulo: Melissa Palmieri. |
| EPI_ISL_1966488 | SECAO CENTRO DE DIAGNOSTICO SECEDI | Instituto Butantan / Mendelics | Instituto Butantan: Dimas Tadeu Covas, Sandra Coccuzzo Sampaio, Maria Carolina Elias, José Salvatore Leister Patané, Vincent Louis Viala, Antonio Jorge Martins, Ricardo Haddad, Claudia Renata dos Santos Barros, Elaine Cristina Marqueze, Raul Machado Neto, Debora Botequio Moretti, Jardelina de Souza Todao Bernardino, Loyze Paola Oliveira de Lima, Luiz Aurelio de Campos Crispin. Centro de Genômica Funcional da ESALQ: Luiz Lehmann Coutinho, Ricardo Augusto Brassaloti, Raquel de Lello Rocha Campos Cassano. NGS Soluções Genômicas: Pilar Drummond Sampaio Corrêa Mariani. FZEA-USP Pirassununga: Mirele Daiana Poleti, Jessika Cristina Chagas Lesbon, Elisangela Chicaroni Mattos, Heidge Fukumasu. USP-Botucatu: Rejane Maria Tommasini Grotto, Jayme A. Souza-Neto, Guilherme Targino Valente, Patricia Akemi Assato, Felipe Allan da Silva da Costa, Bianca Cechetto Carlos. Mendelics: Bibiana Santos, João Paulo Kitajima, Erika Freitas, David Schlesinger. Hemocentro Ribeirão Preto: Simone Kashima, Evandra Strazza Rodrigues, Svetoslav Nanev Slavov, Elaine Vieira dos Santos, Rafael dos Santos Bezerra, Luiz Carlos Junior de Alcantara, Marta Giovanetti, Vagner Fonseca, Flavia Aburjaile, Rodrigo Tocantins Calado. FAMERP-SJRP: Cecília Artico Banho, Lívia Sacchetto, Fábio Sossai Possebon, Leila Sabrina Ullmann, Cíntia Bittar, Guilherme Campos, Helena Lage Ferreira, Jorge A. Petroli Marchesi, Maisa C. Pereira Parra, Marília Moraes, Paula Rahal, Paulo Inacio da Costa, João Pessoa Araújo Jr., Maurício Lacerda Nogueira. Prefeitura de Sao Paulo: Melissa Palmieri. |
| EPI_ISL_1966553 | UBS II DE TANABI MILTON MARTINS PERCHES | Instituto Butantan / FZEA-USP (Pirassununga) | Instituto Butantan: Dimas Tadeu Covas, Sandra Coccuzzo Sampaio, Maria Carolina Elias, José Salvatore Leister Patané, Vincent Louis Viala, Antonio Jorge Martins, Ricardo Haddad, Claudia Renata dos Santos Barros, Elaine Cristina Marqueze, Raul Machado Neto, Debora Botequio Moretti, Jardelina de Souza Todao Bernardino, Loyze Paola Oliveira de Lima, Luiz Aurelio de Campos Crispin. Centro de Genômica Funcional da ESALQ: Luiz Lehmann Coutinho, Ricardo Augusto Brassaloti, Raquel de Lello Rocha Campos Cassano. NGS Soluções Genômicas: Pilar Drummond Sampaio Corrêa Mariani. FZEA-USP Pirassununga: Mirele Daiana Poleti, Jessika Cristina Chagas Lesbon, Elisangela Chicaroni Mattos, Heidge Fukumasu. USP-Botucatu: Rejane Maria Tommasini Grotto, Jayme A. Souza-Neto, Guilherme Targino Valente, Patricia Akemi Assato, Felipe Allan da Silva da Costa, Bianca Cechetto Carlos. Mendelics: Bibiana Santos, João Paulo Kitajima, Erika Freitas, David Schlesinger. Hemocentro Ribeirão Preto: Simone Kashima, Evandra Strazza Rodrigues, Svetoslav Nanev Slavov, Elaine Vieira dos Santos, Rafael dos Santos Bezerra, Luiz Carlos Junior de Alcantara, Marta Giovanetti, Vagner Fonseca, Flavia Aburjaile, Rodrigo Tocantins Calado. FAMERP-SJRP: Cecília Artico Banho, Lívia Sacchetto, Fábio Sossai Possebon, Leila Sabrina Ullmann, Cíntia Bittar, Guilherme Campos, Helena Lage Ferreira, Jorge A. Petroli Marchesi, Maisa C. Pereira Parra, Marília Moraes, Paula Rahal, Paulo Inacio da Costa, João Pessoa Araújo Jr., Maurício Lacerda Nogueira. Prefeitura de Sao Paulo: Melissa Palmieri. |
| EPI_ISL_1966563 | NUCLEO DE SAUDE VILA FALCAO DE BAURU | Instituto Butantan / Mendelics | Instituto Butantan: Dimas Tadeu Covas, Sandra Coccuzzo Sampaio, Maria Carolina Elias, José Salvatore Leister Patané, Vincent Louis Viala, Antonio Jorge Martins, Ricardo Haddad, Claudia Renata dos Santos Barros, Elaine Cristina Marqueze, Raul Machado Neto, Debora Botequio Moretti, Jardelina de Souza Todao Bernardino, Loyze Paola Oliveira de Lima, Luiz Aurelio de Campos Crispin. Centro de Genômica Funcional da ESALQ: Luiz Lehmann Coutinho, Ricardo Augusto Brassaloti, Raquel de Lello Rocha Campos Cassano. NGS Soluções Genômicas: Pilar Drummond Sampaio Corrêa Mariani. FZEA-USP Pirassununga: Mirele Daiana Poleti, Jessika Cristina Chagas Lesbon, Elisangela Chicaroni Mattos, Heidge Fukumasu. USP-Botucatu: Rejane Maria Tommasini Grotto, Jayme A. Souza-Neto, Guilherme Targino Valente, Patricia Akemi Assato, Felipe Allan da Silva da Costa, Bianca Cechetto Carlos. Mendelics: Bibiana Santos, João Paulo Kitajima, Erika Freitas, David Schlesinger. Hemocentro Ribeirão Preto: Simone Kashima, Evandra Strazza Rodrigues, Svetoslav Nanev Slavov, Elaine Vieira dos Santos, Rafael dos Santos Bezerra, Luiz Carlos Junior de Alcantara, Marta Giovanetti, Vagner Fonseca, Flavia Aburjaile, Rodrigo Tocantins Calado. FAMERP-SJRP: Cecília Artico Banho, Lívia Sacchetto, Fábio Sossai Possebon, Leila Sabrina Ullmann, Cíntia Bittar, Guilherme Campos, Helena Lage Ferreira, Jorge A. Petroli Marchesi, Maisa C. Pereira Parra, Marília Moraes, Paula Rahal, Paulo Inacio da Costa, João Pessoa Araújo Jr., Maurício Lacerda Nogueira. Prefeitura de Sao Paulo: Melissa Palmieri. |
| EPI_ISL_1966691 | PRONTO ATENDIMENTO MUNICIPAL DE JACUPIRANGA | Instituto Butantan / Mendelics | Instituto Butantan: Dimas Tadeu Covas, Sandra Coccuzzo Sampaio, Maria Carolina Elias, José Salvatore Leister Patané, Vincent Louis Viala, Antonio Jorge Martins, Ricardo Haddad, Claudia Renata dos Santos Barros, Elaine Cristina Marqueze, Raul Machado Neto, Debora Botequio Moretti, Jardelina de Souza Todao Bernardino, Loyze Paola Oliveira de Lima, Luiz Aurelio de Campos Crispin. Centro de Genômica Funcional da ESALQ: Luiz Lehmann Coutinho, Ricardo Augusto Brassaloti, Raquel de Lello Rocha Campos Cassano. NGS Soluções Genômicas: Pilar Drummond Sampaio Corrêa Mariani. FZEA-USP Pirassununga: Mirele Daiana Poleti, Jessika Cristina Chagas Lesbon, Elisangela Chicaroni Mattos, Heidge Fukumasu. USP-Botucatu: Rejane Maria Tommasini Grotto, Jayme A. Souza-Neto, Guilherme Targino Valente, Patricia Akemi Assato, Felipe Allan da Silva da Costa, Bianca Cechetto Carlos. Mendelics: Bibiana Santos, João Paulo Kitajima, Erika Freitas, David Schlesinger. Hemocentro Ribeirão Preto: Simone Kashima, Evandra Strazza Rodrigues, Svetoslav Nanev Slavov, Elaine Vieira dos Santos, Rafael dos Santos Bezerra, Luiz Carlos Junior de Alcantara, Marta Giovanetti, Vagner Fonseca, Flavia Aburjaile, Rodrigo Tocantins Calado. FAMERP-SJRP: Cecília Artico Banho, Lívia Sacchetto, Fábio Sossai Possebon, Leila Sabrina Ullmann, Cíntia Bittar, Guilherme Campos, Helena Lage Ferreira, Jorge A. Petroli Marchesi, Maisa C. Pereira Parra, Marília Moraes, Paula Rahal, Paulo Inacio da Costa, João Pessoa Araújo Jr., Maurício Lacerda Nogueira. Prefeitura de Sao Paulo: Melissa Palmieri. |
| EPI_ISL_1966752 | UNIDADE SENTINELA COVID19 | Instituto Butantan / Mendelics | Instituto Butantan: Dimas Tadeu Covas, Sandra Coccuzzo Sampaio, Maria Carolina Elias, José Salvatore Leister Patané, Vincent Louis Viala, Antonio Jorge Martins, Ricardo Haddad, Claudia Renata dos Santos Barros, Elaine Cristina Marqueze, Raul Machado Neto, Debora Botequio Moretti, Jardelina de Souza Todao Bernardino, Loyze Paola Oliveira de Lima, Luiz Aurelio de Campos Crispin. Centro de Genômica Funcional da ESALQ: Luiz Lehmann Coutinho, Ricardo Augusto Brassaloti, Raquel de Lello Rocha Campos Cassano. NGS Soluções Genômicas: Pilar Drummond Sampaio Corrêa Mariani. FZEA-USP Pirassununga: Mirele Daiana Poleti, Jessika Cristina Chagas Lesbon, Elisangela Chicaroni Mattos, Heidge Fukumasu. USP-Botucatu: Rejane Maria Tommasini Grotto, Jayme A. Souza-Neto, Guilherme Targino Valente, Patricia Akemi Assato, Felipe Allan da Silva da Costa, Bianca Cechetto Carlos. Mendelics: Bibiana Santos, João Paulo Kitajima, Erika Freitas, David Schlesinger. Hemocentro Ribeirão Preto: Simone Kashima, Evandra Strazza Rodrigues, Svetoslav Nanev Slavov, Elaine Vieira dos Santos, Rafael dos Santos Bezerra, Luiz Carlos Junior de Alcantara, Marta Giovanetti, Vagner Fonseca, Flavia Aburjaile, Rodrigo Tocantins Calado. FAMERP-SJRP: Cecília Artico Banho, Lívia Sacchetto, Fábio Sossai Possebon, Leila Sabrina Ullmann, Cíntia Bittar, Guilherme Campos, Helena Lage Ferreira, Jorge A. Petroli Marchesi, Maisa C. Pereira Parra, Marília Moraes, Paula Rahal, Paulo Inacio da Costa, João Pessoa Araújo Jr., Maurício Lacerda Nogueira. Prefeitura de Sao Paulo: Melissa Palmieri. |
| EPI_ISL_1966922 | HOSPITAL MUNICIPAL DE ITABERA | Instituto Butantan / ESALQ-USP (Piracicaba) | Instituto Butantan: Dimas Tadeu Covas, Sandra Coccuzzo Sampaio, Maria Carolina Elias, José Salvatore Leister Patané, Vincent Louis Viala, Antonio Jorge Martins, Ricardo Haddad, Claudia Renata dos Santos Barros, Elaine Cristina Marqueze, Raul Machado Neto, Debora Botequio Moretti, Jardelina de Souza Todao Bernardino, Loyze Paola Oliveira de Lima, Luiz Aurelio de Campos Crispin. Centro de Genômica Funcional da ESALQ: Luiz Lehmann Coutinho, Ricardo Augusto Brassaloti, Raquel de Lello Rocha Campos Cassano. NGS Soluções Genômicas: Pilar Drummond Sampaio Corrêa Mariani. FZEA-USP Pirassununga: Mirele Daiana Poleti, Jessika Cristina Chagas Lesbon, Elisangela Chicaroni Mattos, Heidge Fukumasu. USP-Botucatu: Rejane Maria Tommasini Grotto, Jayme A. Souza-Neto, Guilherme Targino Valente, Patricia Akemi Assato, Felipe Allan da Silva da Costa, Bianca Cechetto Carlos. Mendelics: Bibiana Santos, João Paulo Kitajima, Erika Freitas, David Schlesinger. Hemocentro Ribeirão Preto: Simone Kashima, Evandra Strazza Rodrigues, Svetoslav Nanev Slavov, Elaine Vieira dos Santos, Rafael dos Santos Bezerra, Luiz Carlos Junior de Alcantara, Marta Giovanetti, Vagner Fonseca, Flavia Aburjaile, Rodrigo Tocantins Calado. FAMERP-SJRP: Cecília Artico Banho, Lívia Sacchetto, Fábio Sossai Possebon, Leila Sabrina Ullmann, Cíntia Bittar, Guilherme Campos, Helena Lage Ferreira, Jorge A. Petroli Marchesi, Maisa C. Pereira Parra, Marília Moraes, Paula Rahal, Paulo Inacio da Costa, João Pessoa Araújo Jr., Maurício Lacerda Nogueira. Prefeitura de Sao Paulo: Melissa Palmieri. |
| EPI_ISL_1967009 | CSII DR WASHINGTON LUIS M RODRIGUES DA SILVA PITANGUEIRAS | Instituto Butantan / ESALQ-USP (Piracicaba) | Instituto Butantan: Dimas Tadeu Covas, Sandra Coccuzzo Sampaio, Maria Carolina Elias, José Salvatore Leister Patané, Vincent Louis Viala, Antonio Jorge Martins, Ricardo Haddad, Claudia Renata dos Santos Barros, Elaine Cristina Marqueze, Raul Machado Neto, Debora Botequio Moretti, Jardelina de Souza Todao Bernardino, Loyze Paola Oliveira de Lima, Luiz Aurelio de Campos Crispin. Centro de Genômica Funcional da ESALQ: Luiz Lehmann Coutinho, Ricardo Augusto Brassaloti, Raquel de Lello Rocha Campos Cassano. NGS Soluções Genômicas: Pilar Drummond Sampaio Corrêa Mariani. FZEA-USP Pirassununga: Mirele Daiana Poleti, Jessika Cristina Chagas Lesbon, Elisangela Chicaroni Mattos, Heidge Fukumasu. USP-Botucatu: Rejane Maria Tommasini Grotto, Jayme A. Souza-Neto, Guilherme Targino Valente, Patricia Akemi Assato, Felipe Allan da Silva da Costa, Bianca Cechetto Carlos. Mendelics: Bibiana Santos, João Paulo Kitajima, Erika Freitas, David Schlesinger. Hemocentro Ribeirão Preto: Simone Kashima, Evandra Strazza Rodrigues, Svetoslav Nanev Slavov, Elaine Vieira dos Santos, Rafael dos Santos Bezerra, Luiz Carlos Junior de Alcantara, Marta Giovanetti, Vagner |

| | | | |
|---|---|---|---|
| | | | Fonseca, Flavia Aburjaile, Rodrigo Tocantins Calado. FAMERP-SJRP: Cecília Artico Banho, Lívia Sacchetto, Fábio Sossai Possebon, Leila Sabrina Ullmann, Cíntia Bittar, Guilherme Campos, Helena Lage Ferreira, Jorge A. Petroli Marchesi, Maisa C. Pereira Parra, Marília Moraes, Paula Rahal, Paulo Inacio da Costa, João Pessoa Araújo Jr., Maurício Lacerda Nogueira. Prefeitura de Sao Paulo: Melissa Palmieri. |
| EPI_ISL_2003156, EPI_ISL_2003160 | Instituto Adolfo Lutz Central | Instituto Adolfo Lutz, Interdiciplinary Procedures Center, Strategic Laboratory | Claudio Tavares Sacchi, Claudia Regina Gonçalves, Erica Valessa Ramos Gomes, Karoline Rodrigues Campos, Caio Vinicius Dias Lopes, Leonardo Jose Tadeu de Araujo |
| EPI_ISL_2008937, EPI_ISL_2008939 | Laboratorio de Pesquisa em Virologia, FAMERP, SJRP | Laboratorio de Pesquisa em Virologia, FAMERP, SJRP | Cecília Artico Banho; Lívia Sacchetto; Fábio Sossai Possebon; Leila Sabrina Ullmann; Cíntia Bittar; Guilherme Campos; Helena Lage Ferreira; Jorge A. Petroli Marchesi; Maisa C. Pereira Parra; Marília Moraes; Paula Rahal; Paulo Inacio da Costa; João Pessoa Araújo Jr.; Maurício L. Nogueira. |
| EPI_ISL_2017243, EPI_ISL_2017247, EPI_ISL_2017251, EPI_ISL_2017253, EPI_ISL_2017254, EPI_ISL_2017258, EPI_ISL_2017259, EPI_ISL_2017264, EPI_ISL_2017266, EPI_ISL_2017268, EPI_ISL_2017271, EPI_ISL_2017275, EPI_ISL_2017276, EPI_ISL_2017309 | | | |
| see above | HLAGYN - Laboratorio de Imunologia de Transplantes de Goias | HLAGYN - Laboratorio de Imunologia de Transplantes de Goias | Fernando Antonio Vinhal dos Santos, Erika Lopes Rocha Batista, Alessandro Leonardo Alvares Magalhaes, Raphael Bessa Parmigiane, Frederico Rodrigues Vinhal, Sabrina Sara Moreira Duarte, Danielle de Paiva Rezende, Lucas Carlos Gomes Pereira, Paola Cristina Resende Silva |
| EPI_ISL_2017450, EPI_ISL_2017457, EPI_ISL_2102517 | HLAGYN - Laboratorio de Imunologia de Transplantes de Goias | HLAGYN - Laboratorio de Imunologia de Transplantes de Goias | Fernando Antonio Vinhal dos Santos, Erika Lopes Rocha Batista, Alessandro Leonardo Alvares Magalhaes, Frederico Rodrigues Vinhal, Sabrina Sara Moreira Duarte, Danielle de Paiva Rezende, Lucas Carlos Gomes Pereira, Paola Cristina Resende Silva |
| EPI_ISL_2107304 | Laboratorio de Pesquisa em Virologia, FAMERP, SJRP | Laboratorio de Pesquisa em Virologia, FAMERP, SJRP | Cecília Artico Banho; Lívia Sacchetto; Fábio Sossai Possebon; Leila Sabrina Ullmann; Cíntia Bittar; Guilherme Campos; Helena Lage Ferreira; Jorge A. Petroli Marchesi; Maisa C. Pereira Parra; Marília Moraes; Paula Rahal; Paulo Inacio da Costa; João Pessoa Araújo Jr.; Maurício L. Nogueira. |
| EPI_ISL_2157343 | Lboratorio Central de Saude Publica do Estado do Parana (LACEN/PR) | Laboratory of Respiratory Viruses and Measles, Oswaldo Cruz Institute, FIOCRUZ | Paola Resende, Luciana Appolinario, Fernando Motta, Anna Carolina Paixao, Ana Carolina Mendonca, Alice Sampaio Rocha, Taina Venas, Elisa Cavalcante Pereira, Renata Serrano Lopes, Irina Riediger, Marilda Siqueira on behalf of the Fiocruz COVID-19 Genomic Surveillance Network |
| EPI_ISL_2157550 | Laboratorio Central de Saude Publica do Estado da Paraiba (LACEN-PB) | Laboratory of Respiratory Viruses and Measles, Oswaldo Cruz Institute, FIOCRUZ | Paola Resende, Luciana Appolinario, Fernando Motta, Anna Carolina Paixao, Ana Carolina Mendonca, Alice Sampaio Rocha, Taina Venas, Elisa Cavalcante Pereira, Renata Serrano Lopes, Joao Felipe Bezerra, Dalane Loudal Florentino Teixeira, Marilda Siqueira on behalf of the Fiocruz COVID-19 Genomic Surveillance Network |
| EPI_ISL_2157572 | Laboratório Central de Saude Publica do Estado de Santa Catarina (LACEN/SC) | Laboratory of Respiratory Viruses and Measles, Oswaldo Cruz Institute, FIOCRUZ | Paola Resende, Luciana Appolinario, Fernando Motta, Anna Carolina Paixao, Ana Carolina Mendonca, Alice Sampaio Rocha, Taina Venas, Elisa Cavalcante Pereira, Renata Serrano Lopes, Darcita Buerger Rovaris, Sandra Bianchini Fernandes, Marilda Siqueira on behalf of the Fiocruz COVID-19 Genomic Surveillance Network |
| EPI_ISL_2157578 | Laboratorio Central de Saude Publica do Estado da Paraiba (LACEN-PB) | Laboratory of Respiratory Viruses and Measles, Oswaldo Cruz Institute, FIOCRUZ | Paola Resende, Luciana Appolinario, Fernando Motta, Anna Carolina Paixao, Ana Carolina Mendonca, Alice Sampaio Rocha, Taina Venas, Elisa Cavalcante Pereira, Renata Serrano Lopes, Joao Felipe Bezerra, Dalane Loudal Florentino Teixeira, Marilda Siqueira on behalf of the Fiocruz COVID-19 Genomic Surveillance Network |
| EPI_ISL_2157587 | Laboratorio Central de Saude Publica do Estado de Sergipe (LACEN/SE) | Laboratory of Respiratory Viruses and Measles, Oswaldo Cruz Institute, FIOCRUZ | Paola Resende, Luciana Appolinario, Fernando Motta, Anna Carolina Paixao, Ana Carolina Mendonca, Alice Sampaio Rocha, Tainá Moreira Martins Venas, Elisa Cavalcante Pereira, Renata Serrano Lopes, Cliomar Alves dos Santos, Marilda Siqueira on behalf of the Fiocruz COVID-19 Genomic Surveillance Network |
| EPI_ISL_2157592 | Laboratório Central de Saude Publica do Estado de Santa Catarina (LACEN/SC) | Laboratory of Respiratory Viruses and Measles, Oswaldo Cruz Institute, FIOCRUZ | Paola Resende, Luciana Appolinario, Fernando Motta, Anna Carolina Paixao, Ana Carolina Mendonca, Alice Sampaio Rocha, Taina Venas, Elisa Cavalcante Pereira, Renata Serrano Lopes, Darcita Buerger Rovaris, Sandra Bianchini Fernandes, Marilda Siqueira on behalf of the Fiocruz COVID-19 Genomic Surveillance Network |
| EPI_ISL_2170898 | VIGILANCIA EPIDEMIOLOGICA | Instituto Butantan / Mendelics | Instituto Butantan: Dimas Tadeu Covas, Sandra Coccuzzo Sampaio, Maria Carolina Elias, José Salvatore Leister Patané, Vincent Louis Viala, Antonio Jorge Martins, Ricardo Haddad, Claudia Renata dos Santos Barros, Elaine Cristina Marqueze, Raul Machado Neto, Debora Botequio Moretti, Jardelina de Souza Todao Bernardino, Loyze Paola Oliveira de Lima, Luiz Aurelio de Campos Crispin. Centro de Genômica Funcional da ESALQ: Luiz Lehmann Coutinho, Ricardo Augusto Brassaloti, Raquel de Lello Rocha Campos Cassano. NGS Soluções Genômicas: Pilar Drummond Sampaio Corrêa Mariani. FZEA-USP Pirassununga: Mirele Daiana Poleti, Jessika Cristina Chagas Lesbon, Elisangela Chicaroni Mattos, Heidge Fukumasu. USP-Botucatu: Rejane Maria Tommasini Grotto, Jayme A. Souza-Neto, Guilherme Targino Valente, Patricia Akemi Assato, Felipe Allan da Silva da Costa, Bianca Cechetto Carlos. Mendelics: Bibiana Santos, João Paulo Kitajima, Erika Freitas, David Schlesinger. Hemocentro Ribeirão Preto: Simone Kashima, Evandra Strazza Rodrigues, Svetoslav Nanev Slavov, Elaine Vieira dos Santos, Rafael dos Santos Bezerra, Luiz Carlos Junior de Alcantara, Marta Giovanetti, Vagner Fonseca, Flavia Aburjaile, Rodrigo Tocantins Calado. FAMERP-SJRP: Cecília Artico Banho, Lívia Sacchetto, Fábio Sossai Possebon, Leila Sabrina Ullmann, Cíntia Bittar, Guilherme Campos, Helena Lage Ferreira, Jorge A. Petroli Marchesi, Maisa C. Pereira Parra, Marília Moraes, Paula Rahal, Paulo Inacio da Costa, João Pessoa Araújo Jr., Maurício Lacerda Nogueira. Prefeitura de Sao Paulo: Melissa Palmieri. |
| EPI_ISL_2187684, EPI_ISL_2187703, EPI_ISL_2187714, EPI_ISL_2187725, EPI_ISL_2187726, EPI_ISL_2187735, EPI_ISL_2187746, EPI_ISL_2187747, EPI_ISL_2187783 | HLAGYN - Laboratorio de Imunologia de Transplantes de Goias | HLAGYN - Laboratorio de Imunologia de Transplantes de Goias | Fernando Antonio Vinhal dos Santos, Erika Lopes Rocha Batista, Alessandro Leonardo Alvares Magalhaes, Frederico Rodrigues Vinhal, Sabrina Sara Moreira Duarte, Lucas Carlos Gomes Pereira, Daniel Ferreira de Sousa |
| EPI_ISL_2196238 | Laboratorio Central de Saude Publica do Estado de Sergipe (LACEN/SE) | Laboratory of Respiratory Viruses and Measles, Oswaldo Cruz Institute, FIOCRUZ | Paola Resende, Luciana Appolinario, Fernando Motta, Anna Carolina Paixao, Ana Carolina Mendonca, Alice Sampaio Rocha, Tainá Moreira Martins Venas, Elisa Cavalcante Pereira, Renata Serrano Lopes, Cliomar Alves dos Santos, Marilda Siqueira on behalf of the Fiocruz COVID-19 Genomic Surveillance Network |
| EPI_ISL_2196357, EPI_ISL_2196360 | Labortorio Central de Saude Publica do Estado de Santa Catarina (LACEN/SC) | Laboratory of Respiratory Viruses and Measles, Oswaldo Cruz Institute, FIOCRUZ | Paola Resende, Luciana Appolinario, Fernando Motta, Anna Carolina Paixao, Ana Carolina Mendonca, Alice Sampaio Rocha, Taina Venas, Elisa Cavalcante Pereira, Renata Serrano Lopes, Darcita Buerger Rovaris, Sandra Bianchini Fernandes, Marilda Siqueira on behalf of the Fiocruz COVID-19 Genomic Surveillance Network |
| EPI_ISL_2196362 | Lboratorio Central de Saude Publica do Estado do Parana (LACEN/PR) | Laboratory of Respiratory Viruses and Measles, Oswaldo Cruz Institute, FIOCRUZ | Paola Resende, Luciana Appolinario, Fernando Motta, Anna Carolina Paixao, Ana Carolina Mendonca, Alice Sampaio Rocha, Taina Venas, Elisa Cavalcante Pereira, Renata Serrano Lopes, Irina Riediger, Marilda Siqueira on behalf of the Fiocruz COVID-19 Genomic Surveillance Network |
| EPI_ISL_2209413 | PRONTO ATENDIMENTO MUNICIPAL ITALO SANTUCCI | Instituto Butantan | Dimas Tadeu Covas, Antonio Jorge Martins, Claudia Renata dos Santos Barros, David Schlesinger, Debora Botequio Moretti, Elaine Cristina Marqueze, Elaine Vieira Santos, Evandra Strazza Rodrigues, Heidge Fukumasu, Jayme Augusto de Souza-Neto, José Salvatore Leister Patané, Luiz Alcantara, Luiz Lehmann Coutinho, Maria Carolina Elias, Maurício Lacerda Nogueira, Rafael dos Santos Bezerra, Raul Machado Neto, Rejane Maria Tommasini Grotto, Ricardo Haddad, Sandra Coccuzzo Sampaio Vessoni, Simone Kashima, Svetoslav Nanev Slavov, Vincent Louis Viala. |
| EPI_ISL_2209422 | PRONTO SOCORRO MUNICIPAL DE TAUBATE | Instituto Butantan | Dimas Tadeu Covas, Antonio Jorge Martins, Claudia Renata dos Santos Barros, David Schlesinger, Debora Botequio Moretti, Elaine Cristina Marqueze, Elaine Vieira Santos, Evandra Strazza Rodrigues, Heidge Fukumasu, Jayme Augusto de Souza-Neto, José Salvatore Leister Patané, Luiz Alcantara, Luiz Lehmann Coutinho, Maria Carolina Elias, Maurício Lacerda Nogueira, Rafael dos Santos Bezerra, Raul Machado Neto, Rejane Maria Tommasini Grotto, Ricardo Haddad, Sandra Coccuzzo Sampaio Vessoni, Simone Kashima, Svetoslav Nanev Slavov, Vincent Louis Viala. |
| EPI_ISL_2209934 | UNIDADE SENTINELA COVID19 | Instituto Butantan | Dimas Tadeu Covas, Antonio Jorge Martins, Claudia Renata dos Santos Barros, David Schlesinger, Debora Botequio Moretti, Elaine Cristina Marqueze, Elaine Vieira Santos, Evandra Strazza Rodrigues, Heidge Fukumasu, Jayme Augusto de Souza-Neto, José Salvatore Leister Patané, Luiz Alcantara, Luiz Lehmann Coutinho, Maria Carolina Elias, Maurício Lacerda Nogueira, Rafael dos Santos Bezerra, Raul Machado Neto, Rejane Maria Tommasini Grotto, Ricardo Haddad, Sandra Coccuzzo Sampaio Vessoni, Simone Kashima, Svetoslav Nanev Slavov, Vincent Louis Viala. |
| EPI_ISL_2227560 | HLAGYN - Laboratorio de Imunologia de Transplantes de Goias | HLAGYN - Laboratorio de Imunologia de Transplantes de Goias | Fernando Antonio Vinhal dos Santos, Erika Lopes Rocha Batista, Alessandro Leonardo Alvares Magalhaes, Frederico Rodrigues Vinhal, Sabrina Sara Moreira Duarte, Lucas Carlos Gomes Pereira, Daniel Ferreira de Sousa |
| EPI_ISL_2241497, EPI_ISL_2241498 | Laboratório Central de Saúde Pública da Paraiba | Coordenação Geral de Laboratórios de Saúde Pública (CGLAB/DAEVS/SVS/MS) | Vagner Fonseca, et al. |
| EPI_ISL_2241508 | Laboratório Central de Saúde Pública do Rio Grande do Norte | Coordenação Geral de Laboratórios de Saúde Pública (CGLAB/DAEVS/SVS/MS) | Vagner Fonseca, et al. |
| EPI_ISL_2241514 | Laboratório Central de Saúde Pública da Paraiba | Coordenação Geral de Laboratórios de Saúde Pública (CGLAB/DAEVS/SVS/MS) | Vagner Fonseca, et al. |

| | | | |
|---|---|---|---|
| EPI_ISL_2241528 | Laboratório Central de Saúde Pública da Bahia | Coordenação Geral de Laboratórios de Saúde Pública (CGLAB/DAEVS/SVS/MS) | Vagner Fonseca, et al. |
| EPI_ISL_2241529 | Laboratório Central de Saúde Pública da Paraiba | Coordenação Geral de Laboratórios de Saúde Pública (CGLAB/DAEVS/SVS/MS) | Vagner Fonseca, et al. |
| EPI_ISL_2241557 | Laboratório Central de Saúde Pública de Sergipe | Coordenação Geral de Laboratórios de Saúde Pública (CGLAB/DAEVS/SVS/MS) | Vagner Fonseca, et al. |
| EPI_ISL_2241566, EPI_ISL_2241567, EPI_ISL_2241572 | Laboratório Central de Saúde Pública da Paraiba | Coordenação Geral de Laboratórios de Saúde Pública (CGLAB/DAEVS/SVS/MS) | Vagner Fonseca, et al. |
| EPI_ISL_2241593 | Laboratório Central de Saúde Pública do Piaui | Coordenação Geral de Laboratórios de Saúde Pública (CGLAB/DAEVS/SVS/MS) | Vagner Fonseca, et al. |
| EPI_ISL_2241596 | Laboratório Central de Saúde Pública da Paraiba | Coordenação Geral de Laboratórios de Saúde Pública (CGLAB/DAEVS/SVS/MS) | Vagner Fonseca, et al. |
| EPI_ISL_2241607 | Laboratório Central de Saúde Pública do Piaui | Coordenação Geral de Laboratórios de Saúde Pública (CGLAB/DAEVS/SVS/MS) | Vagner Fonseca, et al. |
| EPI_ISL_2241609 | Laboratório Central de Saúde Pública de Sergipe | Coordenação Geral de Laboratórios de Saúde Pública (CGLAB/DAEVS/SVS/MS) | Vagner Fonseca, et al. |
| EPI_ISL_2245069, EPI_ISL_2245071 | Laboratório Central de Saúde Pública do Amapá | Coordenação Geral de Laboratórios de Saúde Pública (CGLAB/DAEVS/SVS/MS) | Vagner Fonseca, et al. |
| EPI_ISL_2245187, EPI_ISL_2245188 | Instituto Adolfo Lutz | Coordenação Geral de Laboratórios de Saúde Pública (CGLAB/DAEVS/SVS/MS) | Vagner Fonseca, et al. |
| EPI_ISL_2246287 | Laboratório Central de Saúde Pública do Piaui | Coordenação Geral de Laboratórios de Saúde Pública (CGLAB/DAEVS/SVS/MS) | Vagner Fonseca, et al. |
| EPI_ISL_2248770 | Laboratório Central de Saúde Pública do Maranhão | Coordenação Geral de Laboratórios de Saúde Pública (CGLAB/DAEVS/SVS/MS) | Vagner Fonseca, et al. |
| EPI_ISL_2249348, EPI_ISL_2249352, EPI_ISL_2249353, EPI_ISL_2249355, EPI_ISL_2249362, EPI_ISL_2249379, EPI_ISL_2249382, EPI_ISL_2249386 | Laboratório Central de Saúde Pública do Rio Grande do Sul | Coordenação Geral de Laboratórios de Saúde Pública (CGLAB/DAEVS/SVS/MS) | Vagner Fonseca, et al. |
| EPI_ISL_2249387 | Laboratório Central de Saúde Pública de Santa Catarina | Coordenação Geral de Laboratórios de Saúde Pública (CGLAB/DAEVS/SVS/MS) | Vagner Fonseca, et al. |
| EPI_ISL_2249404, EPI_ISL_2249405, EPI_ISL_2249407, EPI_ISL_2249409, EPI_ISL_2249426, EPI_ISL_2249428 | Laboratório Central de Saúde Pública do Rio de Janeiro | Coordenação Geral de Laboratórios de Saúde Pública (CGLAB/DAEVS/SVS/MS) | Vagner Fonseca, et al. |
| EPI_ISL_2292996, EPI_ISL_2293009 | Laboratório Central de Saúde Pública de Santa Catarina | Coordenação Geral de Laboratórios de Saúde Pública (CGLAB/DAEVS/SVS/MS) | Vagner Fonseca, et al. |
| EPI_ISL_2298748 | Laboratório Central de Saúde Pública de Roraima | Coordenação Geral de Laboratórios de Saúde Pública (CGLAB/DAEVS/SVS/MS) | Vagner Fonseca, et al. |
| EPI_ISL_2298750 | Laboratório Central de Saúde Pública do Maranhão | Coordenação Geral de Laboratórios de Saúde Pública (CGLAB/DAEVS/SVS/MS) | Vagner Fonseca, et al. |
| EPI_ISL_2298865 | Laboratório Central de Saúde Pública do Amazonas | Coordenação Geral de Laboratórios de Saúde Pública (CGLAB/DAEVS/SVS/MS) | Vagner Fonseca, et al. |
| EPI_ISL_2308452, EPI_ISL_2308469 | Laboratório Central de Saúde Pública de Alagoas | Coordenação Geral de Laboratórios de Saúde Pública (CGLAB/DAEVS/SVS/MS) | Vagner Fonseca, et al. |
| EPI_ISL_2344421, EPI_ISL_2344422, EPI_ISL_2344426, EPI_ISL_2344429, EPI_ISL_2344430, EPI_ISL_2344431, EPI_ISL_2344433, EPI_ISL_2344435, EPI_ISL_2344438, EPI_ISL_2344439, EPI_ISL_2344445, EPI_ISL_2344448, EPI_ISL_2344450, EPI_ISL_2344451, EPI_ISL_2344453, EPI_ISL_2344454, EPI_ISL_2344457, EPI_ISL_2344459 | | | |
| see above | Instituto Butantan | Instituto de Medicina Tropical de Sao Paulo | Brazil-UK Centre for Arbovirus Discovery Diagnosis Genomics and Epidemiology (CADDE) Genomic Network - Instituto de Medicina Tropical |
| EPI_ISL_2344592 | PRONTO ATENDIMENTO MUNICIPAL ITALO SANTUCCI | Instituto Butantan / UNESP-Botucatu | Dimas Tadeu Covas, Antonio Jorge Martins, Claudia Renata dos Santos Barros, David Schlesinger, Debora Botequio Moretti, Elaine Cristina Marqueze, Elaine Vieira Santos, Evandra Strazza Rodrigues, Heidge Fukumasu, Jayme Augusto de Souza-Neto, José Salvatore Leister Patané, Luiz Alcantara, Luiz Lehmann Coutinho, Maria Carolina Elias, Maurício Lacerda Nogueira, Rafael dos Santos Bezerra, Raul Machado Neto, Rejane Maria Tommasini Grotto, Ricardo Haddad, Sandra Coccuzzo Sampaio Vessoni, Simone Kashima, Svetoslav Nanev Slavov, Vincent Louis Viala |
| EPI_ISL_2344658 | UNIDADE SENTINELA COVID19 | Instituto Butantan / UNESP-Botucatu | Dimas Tadeu Covas, Antonio Jorge Martins, Claudia Renata dos Santos Barros, David Schlesinger, Debora Botequio Moretti, Elaine Cristina Marqueze, Elaine Vieira Santos, Evandra Strazza Rodrigues, Heidge Fukumasu, Jayme Augusto de Souza-Neto, José Salvatore Leister Patané, Luiz Alcantara, Luiz Lehmann Coutinho, Maria Carolina Elias, Maurício Lacerda Nogueira, Rafael dos Santos Bezerra, Raul Machado Neto, Rejane Maria Tommasini Grotto, Ricardo Haddad, Sandra Coccuzzo Sampaio Vessoni, Simone Kashima, Svetoslav Nanev Slavov, Vincent Louis Viala |
| EPI_ISL_2344686 | PRONTO SOCORRO MUNICIPAL DE TAUBATE | Instituto Butantan / UNESP-Botucatu | Dimas Tadeu Covas, Antonio Jorge Martins, Claudia Renata dos Santos Barros, David Schlesinger, Debora Botequio Moretti, Elaine Cristina Marqueze, Elaine Vieira Santos, Evandra Strazza Rodrigues, Heidge Fukumasu, Jayme Augusto de Souza-Neto, José Salvatore Leister Patané, Luiz Alcantara, Luiz Lehmann Coutinho, Maria Carolina Elias, Maurício Lacerda Nogueira, Rafael dos Santos Bezerra, Raul Machado Neto, Rejane Maria Tommasini Grotto, Ricardo Haddad, Sandra Coccuzzo Sampaio Vessoni, Simone Kashima, Svetoslav Nanev Slavov, Vincent Louis Viala |
| EPI_ISL_2344691 | DIRETORIA MUNICIPAL DE SAUDE DE ENGENHEIRO COELHO | Instituto Butantan / FZEA-USP-Pirassununga | Dimas Tadeu Covas, Antonio Jorge Martins, Claudia Renata dos Santos Barros, David Schlesinger, Debora Botequio Moretti, Elaine Cristina Marqueze, Elaine Vieira Santos, Evandra Strazza Rodrigues, Heidge Fukumasu, Jayme Augusto de Souza-Neto, José Salvatore Leister Patané, Luiz Alcantara, Luiz Lehmann Coutinho, Maria Carolina Elias, Maurício Lacerda Nogueira, Rafael dos Santos Bezerra, Raul Machado Neto, Rejane Maria Tommasini Grotto, Ricardo Haddad, Sandra Coccuzzo Sampaio Vessoni, Simone Kashima, Svetoslav Nanev Slavov, Vincent Louis Viala |
| EPI_ISL_2345349 | POLICLINICA HORTOLANDIA | Instituto Butantan / ESALQ-Piracicaba | Dimas Tadeu Covas, Antonio Jorge Martins, Claudia Renata dos Santos Barros, David Schlesinger, Debora Botequio Moretti, Elaine Cristina Marqueze, Elaine Vieira Santos, Evandra Strazza Rodrigues, Heidge Fukumasu, Jayme Augusto de Souza-Neto, José Salvatore Leister Patané, Luiz Alcantara, Luiz Lehmann Coutinho, Maria Carolina Elias, Maurício Lacerda Nogueira, Rafael dos Santos Bezerra, Raul Machado Neto, Rejane Maria Tommasini Grotto, Ricardo Haddad, Sandra Coccuzzo Sampaio Vessoni, Simone Kashima, Svetoslav Nanev Slavov, Vincent Louis Viala |
| EPI_ISL_2345448 | HOSPITAL MUNICIPAL DE IBIUNA IBIUNA SP | Instituto Butantan / ESALQ-Piracicaba | Dimas Tadeu Covas, Antonio Jorge Martins, Claudia Renata dos Santos Barros, David Schlesinger, Debora Botequio Moretti, Elaine Cristina Marqueze, Elaine Vieira Santos, Evandra Strazza Rodrigues, Heidge Fukumasu, Jayme Augusto de Souza-Neto, José Salvatore Leister Patané, Luiz Alcantara, Luiz Lehmann Coutinho, Maria Carolina Elias, Maurício Lacerda Nogueira, Rafael dos Santos Bezerra, Raul Machado Neto, Rejane Maria Tommasini Grotto, Ricardo Haddad, Sandra Coccuzzo Sampaio Vessoni, Simone Kashima, Svetoslav Nanev Slavov, Vincent Louis Viala |
| EPI_ISL_2345454 | SMS SECRETARIA MUNICIPAL DE SAUDE DE BOITUVA | Instituto Butantan / ESALQ-Piracicaba | Dimas Tadeu Covas, Antonio Jorge Martins, Claudia Renata dos Santos Barros, David Schlesinger, Debora Botequio Moretti, Elaine Cristina Marqueze, Elaine Vieira Santos, Evandra Strazza Rodrigues, Heidge Fukumasu, Jayme Augusto de Souza-Neto, José Salvatore Leister Patané, Luiz Alcantara, Luiz Lehmann Coutinho, Maria Carolina Elias, Maurício Lacerda Nogueira, Rafael dos Santos Bezerra, Raul Machado Neto, Rejane Maria Tommasini Grotto, |

| | | | |
|---|---|---|---|
| | | | Ricardo Haddad, Sandra Coccuzzo Sampaio Vessoni, Simone Kashima, Svetoslav Nanev Slavov, Vincent Louis Viala |
| EPI_ISL_2345552, EPI_ISL_2345573, EPI_ISL_2345574, EPI_ISL_2345580 | LABORATORIO DE FRANCA | Instituto Butantan / ESALQ-Piracicaba | Dimas Tadeu Covas, Antonio Jorge Martins, Claudia Renata dos Santos Barros, David Schlesinger, Debora Botequio Moretti, Elaine Cristina Marqueze, Elaine Vieira Santos, Evandra Strazza Rodrigues, Heidge Fukumasu, Jayme Augusto de Souza-Neto, José Salvatore Leister Patané, Luiz Alcantara, Luiz Lehmann Coutinho, Maria Carolina Elias, Maurício Lacerda Nogueira, Rafael dos Santos Bezerra, Raul Machado Neto, Rejane Maria Tommasini Grotto, Ricardo Haddad, Sandra Coccuzzo Sampaio Vessoni, Simone Kashima, Svetoslav Nanev Slavov, Vincent Louis Viala |
| EPI_ISL_2345734 | VIGILANCIA EPIDEMIOLOGICA JARDINOPOLIS SP | Instituto Butantan / Mendelics | Dimas Tadeu Covas, Antonio Jorge Martins, Claudia Renata dos Santos Barros, David Schlesinger, Debora Botequio Moretti, Elaine Cristina Marqueze, Elaine Vieira Santos, Evandra Strazza Rodrigues, Heidge Fukumasu, Jayme Augusto de Souza-Neto, José Salvatore Leister Patané, Luiz Alcantara, Luiz Lehmann Coutinho, Maria Carolina Elias, Maurício Lacerda Nogueira, Rafael dos Santos Bezerra, Raul Machado Neto, Rejane Maria Tommasini Grotto, Ricardo Haddad, Sandra Coccuzzo Sampaio Vessoni, Simone Kashima, Svetoslav Nanev Slavov, Vincent Louis Viala |
| EPI_ISL_2346035 | UPA DR FRANCO DA ROCHA | Instituto Butantan | Dimas Tadeu Covas, Antonio Jorge Martins, Claudia Renata dos Santos Barros, David Schlesinger, Debora Botequio Moretti, Elaine Cristina Marqueze, Elaine Vieira Santos, Evandra Strazza Rodrigues, Heidge Fukumasu, Jayme Augusto de Souza-Neto, José Salvatore Leister Patané, Luiz Alcantara, Luiz Lehmann Coutinho, Maria Carolina Elias, Maurício Lacerda Nogueira, Rafael dos Santos Bezerra, Raul Machado Neto, Rejane Maria Tommasini Grotto, Ricardo Haddad, Sandra Coccuzzo Sampaio Vessoni, Simone Kashima, Svetoslav Nanev Slavov, Vincent Louis Viala |
| EPI_ISL_2346065, EPI_ISL_2346068 | SERRANA | Instituto Butantan / Mendelics | Dimas Tadeu Covas, Antonio Jorge Martins, Claudia Renata dos Santos Barros, David Schlesinger, Debora Botequio Moretti, Elaine Cristina Marqueze, Elaine Vieira Santos, Evandra Strazza Rodrigues, Heidge Fukumasu, Jayme Augusto de Souza-Neto, José Salvatore Leister Patané, Luiz Alcantara, Luiz Lehmann Coutinho, Maria Carolina Elias, Maurício Lacerda Nogueira, Rafael dos Santos Bezerra, Raul Machado Neto, Rejane Maria Tommasini Grotto, Ricardo Haddad, Sandra Coccuzzo Sampaio Vessoni, Simone Kashima, Svetoslav Nanev Slavov, Vincent Louis Viala |
| EPI_ISL_2348593 | HLAGYN - Laboratorio de Imunologia de Transplantes de Goias | HLAGYN - Laboratorio de Imunologia de Transplantes de Goias | Fernando Antonio Vinhal dos Santos, Erika Lopes Rocha Batista, Alessandro Leonardo Alvares Magalhaes, Frederico Rodrigues Vinhal, Sabrina Sara Moreira Duarte, Lucas Carlos Gomes Pereira, Daniel Ferreira de Sousa |
| EPI_ISL_2399436 | Laboratório de Microbiologia Molecular - Universidade FEEVALE | Molecular Microbiology Laboratory | Alana Witt Hansen, Fágner Henrique Heldt, Fernando Rosado Spilki, Flávio Silveira, Juliana Schons Gularte, Juliane Deise Fleck, Mariana Soares da Silva, Meriane Demoliner, Matheus Nunes Weber, Paula Rodrigues de Almeida, Micheli Filippi. |
| EPI_ISL_2425442, EPI_ISL_2425443 | Instituto Butantan | Instituto de Medicina Tropical de Sao Paulo | Brazil-UK Centre for Arbovirus Discovery Diagnosis Genomics and Epidemiology (CADDE) Genomic Network - Instituto de Medicina Tropical |
| EPI_ISL_2431429, EPI_ISL_2431431, EPI_ISL_2431433 | Laboratório de Microbiologia Molecular - Universidade FEEVALE | Molecular Microbiology Laboratory | Alana Witt Hansen, Fágner Henrique Heldt, Fernando Rosado Spilki, Flávio Silveira, Juliana Schons Gularte, Juliane Deise Fleck, Mariana Soares da Silva, Meriane Demoliner, Matheus Nunes Weber, Paula Rodrigues de Almeida, Micheli Filippi. |
| EPI_ISL_2443595, EPI_ISL_2443635, EPI_ISL_2443636 | Laboratory of Respiratory Viruses and Measles, Oswaldo Cruz Institute, FIOCRUZ | Laboratory of Respiratory Viruses and Measles, Oswaldo Cruz Institute, FIOCRUZ | Paola Resende, Luciana Appolinario, Fernando Motta, Anna Carolina Paixao, Ana Carolina Mendonca, Alice Sampaio Rocha, Taina Venas, Elisa Cavalcante Pereira, Renata Serrano Lopes, Marilda Siqueira on behalf of the Fiocruz COVID-19 Genomic Surveillance Network |
| EPI_ISL_2466150, EPI_ISL_2466153, EPI_ISL_2466157, EPI_ISL_2466161, EPI_ISL_2466172, EPI_ISL_2466176, EPI_ISL_2466186, EPI_ISL_2466188, EPI_ISL_2466189, EPI_ISL_2466229 | Laboratório de Biologia Molecular de Doenças Infecciosas e do Câncer (LADIC - UFRN) | Laboratory of Respiratory Viruses and Measles, Oswaldo Cruz Institute, FIOCRUZ | Paola Resende, Josélio Araújo, Luciana Appolinario, Fernando Motta, Anna Carolina Paixao, Ana Carolina Mendonca, Alice Sampaio Rocha, Taina Venas, Elisa Cavalcante Pereira, Renata Serrano Lopes, Marilda Siqueira on behalf of the Fiocruz COVID-19 Genomic Surveillance Network |
| EPI_ISL_2488770 | LACEN - Laboratório Central de Saúde Pública do Ceará | Evandro Chagas Institute | Santos, M.C.; Silva, A.M.; Junior, W.D.C.; Barbagelata, L.S.; Ferreira, J.A.; Sousa, E.M.A.; da Silva, P.S.; Pinheiro, K.C.; L.C.; Sousa Junior, E.C. |
| EPI_ISL_2488773, EPI_ISL_2488810 | LACEN - Laboratório Central de Saúde Pública do Maranhao | Evandro Chagas Institute | Santos, M.C.; Silva, A.M.; Junior, W.D.C.; Barbagelata, L.S.; Ferreira, J.A.; Sousa, E.M.A.; da Silva, P.S.; Pinheiro, K.C.; L.C.; Sousa Junior, E.C. |
| EPI_ISL_2491693 | Centro de Pesquisa Gonçalo Moniz (CPqGM - FIOCRUZ/BA) | Laboratory of Respiratory Viruses and Measles, Oswaldo Cruz Institute, FIOCRUZ | Paola Resende, Luciana Appolinario, Fernando Motta, Anna Carolina Paixao, Ana Carolina Mendonca, Alice Sampaio Rocha, Taina Venas, Elisa Cavalcante Pereira, Renata Serrano Lopes, Ricardo Khouri, Camila I. de Oliveira, Marilda Siqueira on behalf of the Fiocruz COVID-19 Genomic Surveillance Network |
| EPI_ISL_2491721 | Laboratorio Central de Saude Publica do Estado da Bahia (LACEN/BA) | Laboratory of Respiratory Viruses and Measles, Oswaldo Cruz Institute, FIOCRUZ | Paola Resende, Luciana Appolinario, Fernando Motta, Anna Carolina Paixao, Ana Carolina Mendonca, Alice Sampaio Rocha, Taina Venas, Elisa Cavalcante Pereira, Renata Serrano Lopes,Felicidade Pereira, Marilda Siqueira on behalf of the Fiocruz COVID-19 Genomic Surveillance Network |
| EPI_ISL_2491722 | Universidade Federal do Sul da Bahia (UFSB) | Laboratory of Respiratory Viruses and Measles, Oswaldo Cruz Institute, FIOCRUZ | Paola Resende, Luciana Appolinario, Fernando Motta, Anna Carolina Paixao, Ana Carolina Mendonca, Alice Sampaio Rocha, Taina Venas, Elisa Cavalcante Pereira, Renata Serrano Lopes,Felicidade Pereira, Thiago Mafra, Marilda Siqueira on behalf of the Fiocruz COVID-19 Genomic Surveillance Network |
| EPI_ISL_2491723, EPI_ISL_2491724, EPI_ISL_2491725, EPI_ISL_2491730, EPI_ISL_2491731, EPI_ISL_2491751 | Laboratorio Central de Saude Publica do Estado da Bahia (LACEN/BA) | Laboratory of Respiratory Viruses and Measles, Oswaldo Cruz Institute, FIOCRUZ | Paola Resende, Luciana Appolinario, Fernando Motta, Anna Carolina Paixao, Ana Carolina Mendonca, Alice Sampaio Rocha, Taina Venas, Elisa Cavalcante Pereira, Renata Serrano Lopes,Felicidade Pereira, Marilda Siqueira on behalf of the Fiocruz COVID-19 Genomic Surveillance Network |
| EPI_ISL_2491756, EPI_ISL_2491758, EPI_ISL_2491760, EPI_ISL_2491762, EPI_ISL_2491780, EPI_ISL_2491781 | Centro de Pesquisa Gonçalo Moniz (CPqGM - FIOCRUZ/BA) | Laboratory of Respiratory Viruses and Measles, Oswaldo Cruz Institute, FIOCRUZ | Paola Resende, Luciana Appolinario, Fernando Motta, Anna Carolina Paixao, Ana Carolina Mendonca, Alice Sampaio Rocha, Taina Venas, Elisa Cavalcante Pereira, Renata Serrano Lopes, Ricardo Khouri, Camila I. de Oliveira, Marilda Siqueira on behalf of the Fiocruz COVID-19 Genomic Surveillance Network |
| EPI_ISL_2497434 | HLAGYN - Laboratorio de Imunologia de Transplantes de Goias | HLAGYN - Laboratorio de Imunologia de Transplantes de Goias | Fernando Antonio Vinhal dos Santos, Erika Lopes Rocha Batista, Alessandro Leonardo Alvares Magalhaes, Frederico Rodrigues Vinhal, Sabrina Sara Moreira Duarte, Lucas Carlos Gomes Pereira, Daniel Ferreira de Sousa |
| EPI_ISL_2536330, EPI_ISL_2536352 | Laboratorio Central de Saude Publica do Estado da Paraiba (LACEN-PB) | Laboratory of Respiratory Viruses and Measles, Oswaldo Cruz Institute, FIOCRUZ | Paola Resende, Luciana Appolinario, Fernando Motta, Anna Carolina Paixao, Ana Carolina Mendonca, Alice Sampaio Rocha, Taina Venas, Elisa Cavalcante Pereira, Renata Serrano Lopes, Joao Felipe Bezerra, Dalane Loudal Florentino Teixeira, Marilda Siqueira on behalf of the Fiocruz COVID-19 Genomic Surveillance Network |
| EPI_ISL_2544838, EPI_ISL_2544848, EPI_ISL_2544849, EPI_ISL_2544851, EPI_ISL_2544857, EPI_ISL_2544858, EPI_ISL_2544862, EPI_ISL_2544873, EPI_ISL_2544879, EPI_ISL_2544880, EPI_ISL_2544894, EPI_ISL_2544895 | | | |
| see above | Laboratorio de Pesquisa em Virologia, FAMERP, SJRP | Laboratorio de Pesquisa em Virologia, FAMERP, SJRP | Cecília Artico Banho; Lívia Sacchetto; Guilherme Campos; Fábio Sossai Possebon; Leila Sabrina Ullmann; Cíntia Bittar; Helena Lage Ferreira; Jorge A. Petroli Marchesi; Maisa C. Pereira Parra; Marília Moraes; Paula Rahal; Paulo Inacio da Costa; João Pessoa Araújo Jr.; Maurício L. Nogueira. |
| EPI_ISL_2557340, EPI_ISL_2557341, EPI_ISL_2557342, EPI_ISL_2557346, EPI_ISL_2557350, EPI_ISL_2557353, EPI_ISL_2557358 | Laboratorio Central de Saude Publica do Estado de Minas Gerais (LACEN/MG) | Laboratory of Respiratory Viruses and Measles, Oswaldo Cruz Institute, FIOCRUZ | Paola Resende, Luciana Appolinario, Fernando Motta, Anna Carolina Paixao, Ana Carolina Mendonca, Alice Sampaio Rocha, Taina Venas, Elisa Cavalcante Pereira, Renata Serrano Lopes, Andre Felipe Leal Bernardes, Marilda Siqueira on behalf of the Fiocruz COVID-19 Genomic Surveillance Network |
| EPI_ISL_2557388, EPI_ISL_2557400, EPI_ISL_2603472, EPI_ISL_2603477, EPI_ISL_2603487, EPI_ISL_2603499, EPI_ISL_2603501, EPI_ISL_2603505, EPI_ISL_2603512, EPI_ISL_2603516, EPI_ISL_2603517, EPI_ISL_2603520 | | | |
| see above | Laboratory of Respiratory Viruses and Measles, Oswaldo Cruz Institute, FIOCRUZ | Laboratory of Respiratory Viruses and Measles, Oswaldo Cruz Institute, FIOCRUZ | Paola Resende, Luciana Appolinario, Fernando Motta, Anna Carolina Paixao, Ana Carolina Mendonca, Alice Sampaio Rocha, Taina Venas, Elisa Cavalcante Pereira, Renata Serrano Lopes, Marilda Siqueira on behalf of the Fiocruz COVID-19 Genomic Surveillance Network |
| EPI_ISL_2603526 | Laboratorio Central de Saude Publica do Estado do Rio Grande do Sul (LACEN-RS) | Laboratory of Respiratory Viruses and Measles, Oswaldo Cruz Institute, FIOCRUZ | Paola Resende, Luciana Appolinario, Fernando Motta, Anna Carolina Paixao, Ana Carolina Mendonca, Alice Sampaio Rocha, Taina Venas, Elisa Cavalcante Pereira, Renata Serrano Lopes, Anderson Brandao Leite, Marilda Siqueira on behalf of the Fiocruz COVID-19 Genomic Surveillance Network |
| EPI_ISL_2612324, EPI_ISL_2612328, EPI_ISL_2612348, EPI_ISL_2612349, EPI_ISL_2612353, EPI_ISL_2612355, EPI_ISL_2612361, EPI_ISL_2612364, EPI_ISL_2612393, EPI_ISL_2612394, EPI_ISL_2612395 | | | |
| see above | Centro de Infectologia Charles Mérieux/ Laboratório Rodolphe Mérieux, FUNDHACRE | Bioinformatics Laboratory / LNCC | Alessandra P Lamarca, Luiz G P de Almeida, Ronaldo da Silva F Jr,Douglas Terra Machado, Alexandra L Gerber, Ana Paula de C Guimarães, Cirley Maria de Oliveira Lobato, Andreas Stocker, Luiz Fellype Alves de Souza, Ana Tereza R Vasconcelos |
| EPI_ISL_2614072 | Laboratory of Molecular Virology, Federal University of Rio de Janeito, UFRJ | Laboratory of Respiratory Viruses and Measles, Oswaldo Cruz Institute, FIOCRUZ | Paola Resende, Amilcar Tanuri, Luciana Appolinario, Fernando Motta, Anna Carolina Paixao, Ana Carolina Mendonca, Alice Sampaio Rocha, Taina Venas, Elisa Cavalcante Pereira, Renata Serrano Lopes, Marilda Siqueira on behalf of the Fiocruz COVID-19 Genomic Surveillance Network |
| EPI_ISL_2614095, EPI_ISL_2614103, EPI_ISL_2614104, EPI_ISL_2614144, EPI_ISL_2614156, EPI_ISL_2614160, EPI_ISL_2614169, EPI_ISL_2614181 | Laboratory of Respiratory Viruses and Measles, Oswaldo Cruz Institute, FIOCRUZ | Laboratory of Respiratory Viruses and Measles, Oswaldo Cruz Institute, FIOCRUZ | Paola Resende, Luciana Appolinario, Fernando Motta, Anna Carolina Paixao, Ana Carolina Mendonca, Alice Sampaio Rocha, Taina Venas, Elisa Cavalcante Pereira, Renata Serrano Lopes, Marilda Siqueira on behalf of the Fiocruz COVID-19 Genomic Surveillance Network |

| | | | |
|---|---|---|---|
| EPI_ISL_2614351 | Labortorio Central de Saude Publica do Estado do Rio de Janeiro (LACEN/RJ) | Laboratory of Respiratory Viruses and Measles, Oswaldo Cruz Institute, FIOCRUZ | Paola Resende, Luciana Appolinario, Fernando Motta, Anna Carolina Paixao, Ana Carolina Mendonca, Alice Sampaio Rocha, Taina Venas, Elisa Cavalcante Pereira, Renata Serrano Lopes, Andrea Cony Cavalcanti, Marilda Siqueira on behalf of the Fiocruz COVID-19 Genomic Surveillance Network |
| EPI_ISL_2614514, EPI_ISL_2614516, EPI_ISL_2614517 | Instituto Adolfo Lutz Central | Instituto Adolfo Lutz, Interdiciplinary Procedures Center, Strategic Laboratory | Claudio Tavares Sacchi, Claudia Regina Gonçalves, Erica Valessa Ramos Gomes, Karoline Rodrigues Campos, Caio Vinicius Dias Lopes, Leonardo Jose Tadeu de Araujo |
| EPI_ISL_2617626 | HLAGYN - Laboratorio de Imunologia de Transplantes de Goias | HLAGYN - Laboratorio de Imunologia de Transplantes de Goias | Fernando Antonio Vinhal dos Santos, Erika Lopes Rocha Batista, Alessandro Leonardo Alvares Magalhaes, Frederico Rodrigues Vinhal, Sabrina Sara Moreira Duarte, Lucas Carlos Gomes Pereira, Daniel Ferreira de Sousa |
| EPI_ISL_2629608, EPI_ISL_2629624, EPI_ISL_2629634 | Laboratório de Virologia Molecular - Universidade Federal do Rio de Janeiro | Laboratório de Virologia Molecular - Universidade Federal do Rio de Janeiro | Filipe Romero Rebello Moreira, Mirela D'arc, Diana Mariani, Alice Laschuk Herlinger, Francine Bittencourt Schiffler, Átila Duque Rossi, Isabela de Carvalho Leitão, Thamiris dos Santos Miranda, Matheus Augusto Calvano Cosentino, Marcelo Calado de Paula Tôrres, Raíssa Mirella dos Santos Cunha da Costa, Cássia Cristina Alves Gonçalves, Débora Souza Faffe, Rafael Mello Galliez, Orlando da Costa Ferreira Junior, Renato Santana de Aguiar,, André Felipe Andrade dos Santos, Carolina Moreira Voloch, Terezinha Marta Pereira Pinto Castineiras, Amilcar Tanuri |
| EPI_ISL_2645420 | Laboratorio Central de Saude Publica do Estado de Minas Gerais (LACEN-MG) | Laboratory of Respiratory Viruses and Measles, Oswaldo Cruz Institute, FIOCRUZ | Paola Resende, Luciana Appolinario, Fernando Motta, Anna Carolina Paixao, Ana Carolina Mendonca, Alice Sampaio Rocha, Taina Venas, Elisa Cavalcante Pereira, Renata Serrano Lopes, Andre Felipe Leal Bernardes, Marilda Siqueira on behalf of the Fiocruz COVID-19 Genomic Surveillance Network |
| EPI_ISL_2645546, EPI_ISL_2645549, EPI_ISL_2645551, EPI_ISL_2645554, EPI_ISL_2645561, EPI_ISL_2645564, EPI_ISL_2645569, EPI_ISL_2645572, EPI_ISL_2645574, EPI_ISL_2645578, EPI_ISL_2645579, EPI_ISL_2645586, EPI_ISL_2645588, EPI_ISL_2645606, EPI_ISL_2645613, EPI_ISL_2645614, EPI_ISL_2645626, EPI_ISL_2645627, EPI_ISL_2645633 | | | |
| see above | Laboratorio Central de Saude Publica do Estado do Espirito Santo (LACEN/ES) | Laboratory of Respiratory Viruses and Measles, Oswaldo Cruz Institute, FIOCRUZ | Paola Resende, Luciana Appolinario, Fernando Motta, Anna Carolina Paixao, Ana Carolina Mendonca, Alice Sampaio Rocha, Taina Venas, Eilisa Cavalcante Pereira, Renata Serrano Lopes, Rodrigo Ribeiro Rodrigues, Marilda Siqueira on behalf of the Fiocruz COVID-19 Genomic Surveillance Network |
| EPI_ISL_2645637, EPI_ISL_2645638, EPI_ISL_2645656, EPI_ISL_2645685, EPI_ISL_2645688, EPI_ISL_2645695 | Laboratorio Central de Saude Publica do Estado de Alagoas (LACEN/AL) | Laboratory of Respiratory Viruses and Measles, Oswaldo Cruz Institute, FIOCRUZ | Paola Resende, Luciana Appolinario, Fernando Motta, Anna Carolina Paixao, Ana Carolina Mendonca, Alice Sampaio Rocha, Taina Venas, Elisa Cavalcante Pereira, Renata Serrano Lopes, Anderson Brandao Leite, Marilda Siqueira on behalf of the Fiocruz COVID-19 Genomic Surveillance Network |
| EPI_ISL_2645843, EPI_ISL_2645855, EPI_ISL_2645860, EPI_ISL_2645861, EPI_ISL_2645904 | Laboratorio Central de Saude Publica do Estado do Para (LACEN/PA) | Laboratory of Respiratory Viruses and Measles, Oswaldo Cruz Institute, FIOCRUZ | Paola Resende, Luciana Appolinario, Fernando Motta, Anna Carolina Paixao, Ana Carolina Mendonca, Alice Sampaio Rocha, Taina Venas, Elisa Cavalcante Pereira, Renata Serrano Lopes, Valnete Andrade, Marilda Siqueira on behalf of the Fiocruz COVID-19 Genomic Surveillance Network |
| EPI_ISL_2660459, EPI_ISL_2660473, EPI_ISL_2660476, EPI_ISL_2660493, EPI_ISL_2660496, EPI_ISL_2660498, EPI_ISL_2660500, EPI_ISL_2660502, EPI_ISL_2660504 | Laboratorio Central de Saude Publica do Estado de Minas Gerais (LACEN/MG) | Laboratory of Respiratory Viruses and Measles, Oswaldo Cruz Institute, FIOCRUZ | Paola Resende, Luciana Appolinario, Fernando Motta, Anna Carolina Paixao, Ana Carolina Mendonca, Alice Sampaio Rocha, Taina Venas, Elisa Cavalcante Pereira, Renata Serrano Lopes, Andre Felipe Leal Bernardes, Marilda Siqueira on behalf of the Fiocruz COVID-19 Genomic Surveillance Network |
| EPI_ISL_2660600, EPI_ISL_2660637, EPI_ISL_2660638, EPI_ISL_2660663, EPI_ISL_2660664, EPI_ISL_2660670, EPI_ISL_2660678, EPI_ISL_2660689, EPI_ISL_2660690 | Laboratorio Central de Saude Publica do Estado de Sergipe (LACEN/SE) | Laboratory of Respiratory Viruses and Measles, Oswaldo Cruz Institute, FIOCRUZ | Paola Resende, Luciana Appolinario, Fernando Motta, Anna Carolina Paixao, Ana Carolina Mendonca, Alice Sampaio Rocha, Tainá Moreira Martins Venas, Elisa Cavalcante Pereira, Renata Serrano Lopes, Cliomar Alves dos Santos, Marilda Siqueira on behalf of the Fiocruz COVID-19 Genomic Surveillance Network |
| EPI_ISL_2661758, EPI_ISL_2661782, EPI_ISL_2661786, EPI_ISL_2661788, EPI_ISL_2661795, EPI_ISL_2661797, EPI_ISL_2661798, EPI_ISL_2661799, EPI_ISL_2661801, EPI_ISL_2661802, EPI_ISL_2661804, EPI_ISL_2661809, EPI_ISL_2661810, EPI_ISL_2661811, EPI_ISL_2661813, EPI_ISL_2661814, EPI_ISL_2661819, EPI_ISL_2661827, EPI_ISL_2661831, EPI_ISL_2661832, EPI_ISL_2661834, EPI_ISL_2661836, EPI_ISL_2661838, EPI_ISL_2661840, EPI_ISL_2661864, EPI_ISL_2661865 | | | |
| see above | Laboratorio Central de Saude Publica do Estado do Rio Grande do Sul (LACEN-RS) | Laboratory of Respiratory Viruses and Measles, Oswaldo Cruz Institute, FIOCRUZ | Paola Resende, Luciana Appolinario, Fernando Motta, Anna Carolina Paixao, Ana Carolina Mendonca, Alice Sampaio Rocha, Taina Venas, Elisa Cavalcante Pereira, Renata Serrano Lopes, Tatiana Schaffer Gregianini, Richard Salvato, Marilda Siqueira on behalf of the Fiocruz COVID-19 Genomic Surveillance Network |
| EPI_ISL_2661879, EPI_ISL_2661883, EPI_ISL_2661885, EPI_ISL_2661893, EPI_ISL_2661894, EPI_ISL_2661903 | Oswaldo Cruz Institute, FIOCRUZ/CE | Laboratory of Respiratory Viruses and Measles, Oswaldo Cruz Institute, FIOCRUZ | Paola Resende, Fabio Miyajima, Luciana Appolinario, Fernando Motta, Anna Carolina Paixao, Ana Carolina Mendonca, Alice Sampaio Rocha, Taina Venas, Elisa Cavalcante Pereira, Renata Serrano Lopes, Marilda Siqueira on behalf of the Fiocruz COVID-19 Genomic Surveillance Network |
| EPI_ISL_2663256, EPI_ISL_2663286 | Plataforma de Vigilancia Molecular (PVM) - FIOCRUZ/BA | Plataforma de Vigilancia Molecular (PVM) - FIOCRUZ/BA | Ricardo Khouri, Marina Cucco, Tiago Graf, Clarissa Araújo Gurgel, Leonardo Paiva Farias, Bruno Bezerril Andrade, Camila I. de Oliveira on behalf of the Fiocruz COVID-19 Genomic Surveillance Network. |
| EPI_ISL_2677085, EPI_ISL_2677096, EPI_ISL_2677099, EPI_ISL_2677120, EPI_ISL_2677126 | Labortorio Central de Saude Publica do Estado de Santa Catarina (LACEN/SC) | Laboratory of Respiratory Viruses and Measles, Oswaldo Cruz Institute, FIOCRUZ | Paola Resende, Luciana Appolinario, Fernando Motta, Anna Carolina Paixao, Ana Carolina Mendonca, Alice Sampaio Rocha, Taina Venas, Elisa Cavalcante Pereira, Renata Serrano Lopes, Darcita Buerger Rovaris, Sandra Bianchini Fernandes, Marilda Siqueira on behalf of the Fiocruz COVID-19 Genomic Surveillance Network |
| EPI_ISL_2677218 | Laboratory of Respiratory Viruses and Measles, Oswaldo Cruz Institute, FIOCRUZ | Laboratory of Respiratory Viruses and Measles, Oswaldo Cruz Institute, FIOCRUZ | Paola Resende, Luciana Appolinario, Fernando Motta, Anna Carolina Paixao, Ana Carolina Mendonca, Alice Sampaio Rocha, Taina Venas, Elisa Cavalcante Pereira, Renata Serrano Lopes, Marilda Siqueira on behalf of the Fiocruz COVID-19 Genomic Surveillance Network |
| EPI_ISL_2677244, EPI_ISL_2677255, EPI_ISL_2677271, EPI_ISL_2677274, EPI_ISL_2677275, EPI_ISL_2677285, EPI_ISL_2677289, EPI_ISL_2677291, EPI_ISL_2677292, EPI_ISL_2677294, EPI_ISL_2677299, EPI_ISL_2677300, EPI_ISL_2677307, EPI_ISL_2677308 | | | |
| see above | Labortorio Central de Saude Publica do Estado de Santa Catarina (LACEN/SC) | Laboratory of Respiratory Viruses and Measles, Oswaldo Cruz Institute, FIOCRUZ | Paola Resende, Luciana Appolinario, Fernando Motta, Anna Carolina Paixao, Ana Carolina Mendonca, Alice Sampaio Rocha, Taina Venas, Elisa Cavalcante Pereira, Renata Serrano Lopes, Darcita Buerger Rovaris, Sandra Bianchini Fernandes, Marilda Siqueira on behalf of the Fiocruz COVID-19 Genomic Surveillance Network |
| EPI_ISL_2691099 | Instituto Adolfo Lutz - Regional de Marilia | Instituto Adolfo Lutz, Interdiciplinary Procedures Center, Strategic Laboratory | Claudio Tavares Sacchi, Claudia Regina Gonçalves, Erica Valessa Ramos Gomes, Karoline Rodrigues Campos, Caio Vinicius Dias Lopes, Leonardo Jose Tadeu de Araujo |
| EPI_ISL_2698098 | Biology, UFLA | Biology, UFLA | Pylro,V., Cherem,J., Luciano,P., Fernandes,G., Melo,D., Barcante,J. |
| EPI_ISL_2731459 | Lboratorio Central de Saude Publica do Estado do Parana (LACEN/PR) | Laboratory of Respiratory Viruses and Measles, Oswaldo Cruz Institute, FIOCRUZ | Paola Resende, Luciana Appolinario, Fernando Motta, Anna Carolina Paixao, Ana Carolina Mendonca, Alice Sampaio Rocha, Taina Venas, Elisa Cavalcante Pereira, Renata Serrano Lopes, Irina Riediger, Marilda Siqueira on behalf of the Fiocruz COVID-19 Genomic Surveillance Network |
| EPI_ISL_2731460 | Laboratory of Respiratory Viruses and Measles, Oswaldo Cruz Institute, FIOCRUZ | Laboratory of Respiratory Viruses and Measles, Oswaldo Cruz Institute, FIOCRUZ | Paola Resende, Luciana Appolinario, Fernando Motta, Anna Carolina Paixao, Ana Carolina Mendonca, Alice Sampaio Rocha, Taina Venas, Elisa Cavalcante Pereira, Renata Serrano Lopes, Marilda Siqueira on behalf of the Fiocruz COVID-19 Genomic Surveillance Network |
| EPI_ISL_2731461 | Lboratorio Central de Saude Publica do Estado do Parana (LACEN/PR) | Laboratory of Respiratory Viruses and Measles, Oswaldo Cruz Institute, FIOCRUZ | Paola Resende, Luciana Appolinario, Fernando Motta, Anna Carolina Paixao, Ana Carolina Mendonca, Alice Sampaio Rocha, Taina Venas, Elisa Cavalcante Pereira, Renata Serrano Lopes, Irina Riediger, Marilda Siqueira on behalf of the Fiocruz COVID-19 Genomic Surveillance Network |
| EPI_ISL_2731462, EPI_ISL_2731463, EPI_ISL_2731464, EPI_ISL_2731465 | Laboratory of Respiratory Viruses and Measles, Oswaldo Cruz Institute, FIOCRUZ | Laboratory of Respiratory Viruses and Measles, Oswaldo Cruz Institute, FIOCRUZ | Paola Resende, Luciana Appolinario, Fernando Motta, Anna Carolina Paixao, Ana Carolina Mendonca, Alice Sampaio Rocha, Taina Venas, Elisa Cavalcante Pereira, Renata Serrano Lopes, Marilda Siqueira on behalf of the Fiocruz COVID-19 Genomic Surveillance Network |
| EPI_ISL_2731467 | Lboratorio Central de Saude Publica do Estado do Parana (LACEN/PR) | Laboratory of Respiratory Viruses and Measles, Oswaldo Cruz Institute, FIOCRUZ | Paola Resende, Luciana Appolinario, Fernando Motta, Anna Carolina Paixao, Ana Carolina Mendonca, Alice Sampaio Rocha, Taina Venas, Elisa Cavalcante Pereira, Renata Serrano Lopes, Irina Riediger, Marilda Siqueira on behalf of the Fiocruz COVID-19 Genomic Surveillance Network |
| EPI_ISL_2731471 | Labortorio Central de Saude Publica do Estado do Rio de Janeiro (LACEN/RJ) | Laboratory of Respiratory Viruses and Measles, Oswaldo Cruz Institute, FIOCRUZ | Paola Resende, Luciana Appolinario, Fernando Motta, Anna Carolina Paixao, Ana Carolina Mendonca, Alice Sampaio Rocha, Taina Venas, Elisa Cavalcante Pereira, Renata Serrano Lopes, Andrea Cony Cavalcanti, Marilda Siqueira on behalf of the Fiocruz COVID-19 Genomic Surveillance Network |
| EPI_ISL_2731477, EPI_ISL_2731479, EPI_ISL_2731484, EPI_ISL_2731485, EPI_ISL_2731489, EPI_ISL_2731490, EPI_ISL_2731493, EPI_ISL_2731496, EPI_ISL_2731497, EPI_ISL_2731500, EPI_ISL_2731502, EPI_ISL_2731503, EPI_ISL_2731505, EPI_ISL_2731678 | | | |
| see above | Laboratory of Respiratory Viruses and Measles, Oswaldo Cruz Institute, FIOCRUZ | Laboratory of Respiratory Viruses and Measles, Oswaldo Cruz Institute, FIOCRUZ | Paola Resende, Luciana Appolinario, Fernando Motta, Anna Carolina Paixao, Ana Carolina Mendonca, Alice Sampaio Rocha, Taina Venas, Elisa Cavalcante Pereira, Renata Serrano Lopes, Marilda Siqueira on behalf of the Fiocruz COVID-19 Genomic Surveillance Network |
| EPI_ISL_2756442, EPI_ISL_2756452 | Instituto Adolfo Lutz Central | Instituto Adolfo Lutz, Interdiciplinary Procedures Center, Strategic Laboratory | Claudio Tavares Sacchi, Claudia Regina Gonçalves, Erica Valessa Ramos Gomes, Karoline Rodrigues Campos, Caio Vinicius Dias Lopes, Leonardo Jose Tadeu de Araujo |

| | | | |
|---|---|---|---|
| EPI_ISL_2758653, EPI_ISL_2758655, EPI_ISL_2758659, EPI_ISL_2758660, EPI_ISL_2758663, EPI_ISL_2758664, EPI_ISL_2758665, EPI_ISL_2758666, EPI_ISL_2758667 | UEL | IPEC Guarapuava | NAPI-Genômica (Novos Arranjo de Pesquisa e Inovação em Genômica): Ademar Dantas da Cunha Júnior Adriano Ferrasa Adriano Mondini Aldo Przybysz Alessandra Lourenço Cecchini Armani Alex Sandro Jorge Alexandra Ivo de Medeiros Alexandre Maller Aline Cristina Batista Rodrigues Johann Ana Lucia Ferreira Ana Marisa Fusco Almeida Anderson Joel Martino Andrade André Luís Laforga Vanzela Andrea Duarte Doetzer Andrea Name Colado Simao Andressa Pereira de Souza Anelisa Ramão Angelica Beate Winter Boldt Anna Hermínia Castro Gomes de Amorim Anna Silvia Penteado Setti da Rocha Antonio Camilo da Silva Filho Antonio Stabelini Neto Arthur Hirata Bertachi Barbara Mendes Paz Chao Betty Cristiane Kuhn Bruno Ambrozio Galindo Bruno Ribeiro Cruz Camilla Reginatto De Pierri Carla Fredrichsen Moya Araujo Carla Fredrichsen Moya Araujo Carlos Alberto Oliveira de Biagi Junior Carlos Augusto Nassar Carlos Eduardo Buss Carlos Gilberto Carlotti Junior Carlos Henrique Schneider Carolina Panis Carolina Weigert Galvão Caroline de Jesus Coelho Donha Caroline Guisantes de Salvo Toni Caryna Eurich Mazur Catiuscie Cabreira da Silva Tortorella Celso F. D. Doliveira Cesar Luiz Boguszewski Christiane Pienna Soares Chung Man Chin Claudia Moro Cleverson Busso Cristiane Cominetti Daiane Priscila Simão-Silva Dalila Luciola Zanette Daniel de Paula Daniel de Paula Daniel Rech Daniela Fiori Gradia Daniela Pretti da Cunha Tirapelli Daniela Viganó Zanoti Jeronymo Daniele Ukan Danielle Malheiros Ferreira Danielle Venturini Deborah Catharine de Assis Leite Deivid Calebe de Souza Dennis Armando Bertolini Edenir Inez Pamero Edna Maria Vissoci Reiche Edson Roberto Arpini Miguel Eduardo José de Almeida Araújo Eliana Carolina Vespero Eliandro Reis Tavares Elza Kimura Grimshaw Emanuel Maltempi de Souza Emanuele Cristina Gustani Buss Émerson Carraro Emiliana Cristina Melo ENILze Maria de Souza Fonseca Ribeiro Enilze Maria de Souza Fonseca Ribeiro Erika Izumi Erika Seki Kioshima Cotica Evani Marques Pereira Fabio Negretti Fábio Rodrigues Ferreira Seiva Felipe Dunin dos Santos Felipe Tuon Fernanda Andreia Rosa Fernanda Cestaro Prado Cortez Fernanda Ivanski Fernanda Maris Peria Flavia Regina Oliveira de Barros Franciele Aní Caovilla Follador Franciele Mara Lucca Zanardo Bohm Francinete Ramos Campos Fulviana Silva Nishiyama GABRIEL RIBEIRO CORDEIRO Gabriela Datsch Bennemann Gisele Santos de Oliveira Glaucio Valdameri Glauco Akelinghton Freire Vitiello Glauco Vieira Miranda Glaura Scantamburlo ALves Fernandes Guilherme Ferreira Silveira Gustavo Bianchini Porfírio Gustavo Lenci Marques Hélito Volpato Hildebrando Masshiro Nagai Huei Diana Lee Ilce Mara de Syllos Cólus Iris Rabinovich Israel Gomy Jackson Kawakami Jacques Duílio Brancher Jaime Luís Lopes Rocha Jaqueline Carvalho de Oliveira Jean Henrique da Silva Rodrigues Jean Leandro dos Santos Jeane Eliete Laguila Visentainer João Paulo Bianchi Ximenez Joaquim Manoel da Silva Jociani Ascari Joel Donazzolo Jorge Luis Maria Ruiz Jose Knopfholz José Luis da Conceição Silva José Sebastião dos Santos Joseane Carla Schabarum Juliana Cheleski Wiggers Juliana Mara Serpeloni Juliana Morini Küpper Cardoso Perseguini Karen Brajão de Oliveira Karin Braun Prado Karine Aparecida de Lima Katiany Rizzieri Caleffi Ferracioli Katiuscia de Oliveira Francisco Gabriel Kelvinson Fernandes Viana Larissa Beatriz Cossalter Larissa Danielle Bahls Pinto Laurival Antonio Vilas Boas Léia Carolina Lucio Líbero Mezzadri Neto Ligia Carla Faccin Galhardi Lirane Elize Defante Ferreto Luciana Furlaneto Maia Luciana Oliveira de Fariña Luciana Reis Azevedo Alanis Luciane Regina Cavalli Lucy Megumi Yamauchi Lioni Luis Paulo Gomes Mascarenhas Luis Paulo Gomes Mascarenhas Luís Paulo Mascarenhas Lupe Furtado Alle Lyvia Regina Biagi Silva Bertachi Mara Antonia Ramos Costa Mara L. Cordeiro Marcela Maria Birolim Marcelo Ricardo Vicari Marcia Edilaine Lopes Consolaro Marcia Holsbach Beltrame Marcia Regina Eches Perugini Marcos Abdo Arbex Marcos Pileggi MARCOS TADEU GRZELCZAK Marcus Peikriszwili Tartaruga Maria Angelica Ehara Watanabe Maria Antonia Ramos Costa Maria Claudia Gross Maria José Soares Mendes Giannini Maria Leandra Terencio Maria Lúcia Bonfleur Maria Luiza Guimarães de Oliveira Maria Luiza Petzl-Erler Mariana Abe Vicente Cavagnari Marina Kimiko Kadowaki Marise Fonseca dos Santos Marla Karine Amarante Maurício Turkiewicz Mauro Antonio Alves Castro Michel Rodrigo Zambrano Passarini Michele Potrich Michelle Orane Schemberger Milena Massumi Kozonoe Mônica Degraf Cavallin Monica Tereza Suldofski Mucio Luiz de Assis Cirino Nadia Graciele Krohn Najeh Maissar Khalil Nédia de Castilhos Ghisi Neide Tomimura Costa Neiva Leite Neyva Maria Lopes Romeiro Patrícia Amâncio da Rosa Patricia Dayane Carvalho Schaker Patricia Oehlmeyer Nassar Patrícia Savio de Araújo-Souza Patricia Silva Lucio Paulo Henrique Couto Souza Paulo Roberto Donadio Percy Nohama Quirino Alves de Lima Neto Rafael Deminice Rafael dos Santos Bezerra Raquel Alves dos Santos Renan Manozzo Galante Renata Ernlund Freitas de Macedo Rita de Cássia Garcia Simão Roberta Losi Guembarovski Roberto H. Herai Roberto Rosati Rodrigo Ferreira Rodrigo Rodrigues Matiello Rogério Neri Shinsato Rogério Pincela Mateus Rosane Aparecida Ribeiro Rosilene Fressatti Cardoso Rosilene Fressatti Cardoso Sandra Mara Guse Scós Venske Selene Elifio Esposito Sérgio Ossamu Ioshii Silvana Giuliatti Silvia Mara de Souza Halick Silvio Henrique Maia de Almeida Simone Neumann Wendt Spencer Luiz Marques Payão Stefan Wolanski Negrão Stephane Janaina de Moura Escobar Sueli Fumie Yamada Ogatta SUELI PERCIO QUINAIA Taciane Finatto Tatiana Mayumi Veiga Iriyoda Tayza Katelline Danilau Ostroski Tony Alexander Hild Valeria Valente Vanessa Nascimento Kozak Vanessa Santos Sotomaior Victor Breno Pedrosa Victoria Zeghbi Cochenski Borba Vivian Rotuno Moure Valdameri Wander Rogerio Pavanelli Weber Cláudio Francisco Nunes da Silva Willian Augusto de Melo Yohandra Reyes Torres |
| EPI_ISL_2758668, EPI_ISL_2758669, EPI_ISL_2758671, EPI_ISL_2758672 | IPEC Guarapuava | IPEC Guarapuava | NAPI-Genômica (Novos Arranjo de Pesquisa e Inovação em Genômica): Ademar Dantas da Cunha Júnior Adriano Ferrasa Adriano Mondini Aldo Przybysz Alessandra Lourenço Cecchini Armani Alex Sandro Jorge Alexandra Ivo de Medeiros Alexandre Maller Aline Cristina Batista Rodrigues Johann Ana Lucia Ferreira Ana Marisa Fusco Almeida Anderson Joel Martino Andrade André Luís Laforga Vanzela Andrea Duarte Doetzer Andrea Name Colado Simao Andressa Pereira de Souza Anelisa Ramão Angelica Beate Winter Boldt Anna Hermínia Castro Gomes de Amorim Anna Silvia Penteado Setti da Rocha Antonio Camilo da Silva Filho Antonio Stabelini Neto Arthur Hirata Bertachi Barbara Mendes Paz Chao Betty Cristiane Kuhn Bruno Ambrozio Galindo Bruno Ribeiro Cruz Camilla Reginatto De Pierri Carla Fredrichsen Moya Araujo Carla Fredrichsen Moya Araujo Carlos Alberto Oliveira de Biagi Junior Carlos Augusto Nassar Carlos Eduardo Buss Carlos Gilberto Carlotti Junior Carlos Henrique Schneider Carolina Panis Carolina Weigert Galvão Caroline de Jesus Coelho Donha Caroline Guisantes de Salvo Toni Caryna Eurich Mazur Catiuscie Cabreira da Silva Tortorella Celso F. D. Doliveira Cesar Luiz Boguszewski Christiane Pienna Soares Chung Man Chin Claudia Moro Cleverson Busso Cristiane Cominetti Daiane Priscila Simão-Silva Dalila Luciola Zanette Daniel de Paula Daniel de Paula Daniel Rech Daniela Fiori Gradia Daniela Pretti da Cunha Tirapelli Daniela Viganó Zanoti Jeronymo Daniele Ukan Danielle Malheiros Ferreira Danielle Venturini Deborah Catharine de Assis Leite Deivid Calebe de Souza Dennis Armando Bertolini Edenir Inez Pamero Edna Maria Vissoci Reiche Edson Roberto Arpini Miguel Eduardo José de Almeida Araújo Eliana Carolina Vespero Eliandro Reis Tavares Elza Kimura Grimshaw Emanuel Maltempi de Souza Emanuele Cristina Gustani Buss Émerson Carraro Emiliana Cristina Melo ENILze Maria de Souza Fonseca Ribeiro Enilze Maria de Souza Fonseca Ribeiro Erika Izumi Erika Seki Kioshima Cotica Evani Marques Pereira Fabio Negretti Fábio Rodrigues Ferreira Seiva Felipe Dunin dos Santos Felipe Tuon Fernanda Andreia Rosa Fernanda Cestaro Prado Cortez Fernanda Ivanski Fernanda Maris Peria Flavia Regina Oliveira de Barros Franciele Aní Caovilla Follador Franciele Mara Lucca Zanardo Bohm Francinete Ramos Campos Fulviana Silva Nishiyama GABRIEL RIBEIRO CORDEIRO Gabriela Datsch Bennemann Gisele Santos de Oliveira Glaucio Valdameri Glauco Akelinghton Freire Vitiello Glauco Vieira Miranda Glaura Scantamburlo ALves Fernandes Guilherme Ferreira Silveira Gustavo Bianchini Porfírio Gustavo Lenci Marques Hélito Volpato Hildebrando Masshiro Nagai Huei Diana Lee Ilce Mara de Syllos Cólus Iris Rabinovich Israel Gomy Jackson Kawakami Jacques Duílio Brancher Jaime Luís Lopes Rocha Jaqueline Carvalho de Oliveira Jean Henrique da Silva Rodrigues Jean Leandro dos Santos Jeane Eliete Laguila Visentainer João Paulo Bianchi Ximenez Joaquim Manoel da Silva Jociani Ascari Joel Donazzolo Jorge Luis Maria Ruiz Jose Knopfholz José Luis da Conceição Silva José Sebastião dos Santos Joseane Carla Schabarum Juliana Cheleski Wiggers Juliana Mara Serpeloni Juliana Morini Küpper Cardoso Perseguini Karen Brajão de Oliveira Karin Braun Prado Karine Aparecida de Lima Katiany Rizzieri Caleffi Ferracioli Katiuscia de Oliveira Francisco Gabriel Kelvinson Fernandes Viana Larissa Beatriz Cossalter Larissa Danielle Bahls Pinto Laurival Antonio Vilas Boas Léia Carolina Lucio Líbero Mezzadri Neto Ligia Carla Faccin Galhardi Lirane Elize Defante Ferreto Luciana Furlaneto Maia Luciana Oliveira de Fariña Luciana Reis Azevedo Alanis Luciane Regina Cavalli Lucy Megumi Yamauchi Lioni Luis Paulo Gomes Mascarenhas Luis Paulo Gomes Mascarenhas Luís Paulo Mascarenhas Lupe Furtado Alle Lyvia Regina Biagi Silva Bertachi Mara Antonia Ramos Costa Mara L. Cordeiro Marcela Maria Birolim Marcelo Ricardo Vicari Marcia Edilaine Lopes Consolaro Marcia Holsbach Beltrame Marcia Regina Eches Perugini Marcos Abdo Arbex Marcos Pileggi MARCOS TADEU GRZELCZAK Marcus Peikriszwili Tartaruga Maria Angelica Ehara Watanabe Maria Antonia Ramos Costa Maria Claudia Gross Maria José Soares Mendes Giannini Maria Leandra Terencio Maria Lúcia Bonfleur Maria Luiza Guimarães de Oliveira Maria Luiza Petzl-Erler Mariana Abe Vicente Cavagnari Marina Kimiko Kadowaki Marise Fonseca dos Santos Marla Karine Amarante Maurício Turkiewicz Mauro Antonio Alves Castro Michel Rodrigo Zambrano Passarini Michele Potrich Michelle Orane Schemberger Milena Massumi Kozonoe Mônica Degraf Cavallin Monica Tereza Suldofski Mucio Luiz de Assis Cirino Nadia Graciele Krohn Najeh Maissar Khalil Nédia de Castilhos Ghisi Neide Tomimura Costa Neiva Leite Neyva Maria Lopes Romeiro Patrícia Amâncio da Rosa Patricia Dayane Carvalho Schaker Patricia Oehlmeyer Nassar Patrícia Savio de Araújo-Souza Patricia Silva Lucio Paulo Henrique Couto Souza Paulo Roberto Donadio Percy Nohama Quirino Alves de Lima Neto Rafael Deminice Rafael dos Santos Bezerra Raquel Alves dos Santos Renan Manozzo Galante Renata Ernlund Freitas de Macedo Rita de Cássia Garcia Simão Roberta Losi Guembarovski Roberto H. Herai Roberto Rosati Rodrigo Ferreira Rodrigo Rodrigues Matiello Rogério Neri Shinsato Rogério Pincela Mateus Rosane Aparecida Ribeiro Rosilene Fressatti Cardoso Rosilene Fressatti Cardoso Sandra Mara Guse Scós Venske Selene Elifio Esposito Sérgio Ossamu Ioshii Silvana Giuliatti Silvia Mara de Souza Halick Silvio Henrique Maia de Almeida Simone Neumann Wendt Spencer Luiz Marques Payão Stefan Wolanski Negrão Stephane Janaina de Moura Escobar Sueli Fumie Yamada Ogatta SUELI PERCIO QUINAIA Taciane Finatto Tatiana Mayumi Veiga Iriyoda Tayza Katelline Danilau Ostroski Tony Alexander Hild Valeria Valente Vanessa Nascimento Kozak Vanessa Santos Sotomaior Victor Breno Pedrosa Victoria Zeghbi Cochenski Borba Vivian Rotuno Moure Valdameri Wander Rogerio Pavanelli Weber Cláudio Francisco Nunes da Silva Willian Augusto de Melo Yohandra Reyes Torres |
| EPI_ISL_2758675 | HUEM/IBMP | IPEC Guarapuava | NAPI-Genômica (Novos Arranjo de Pesquisa e Inovação em Genômica): Ademar Dantas da Cunha Júnior Adriano Ferrasa Adriano Mondini Aldo Przybysz Alessandra Lourenço Cecchini Armani Alex Sandro Jorge Alexandra Ivo de Medeiros Alexandre Maller Aline Cristina Batista Rodrigues Johann |

| | | | |
|---|---|---|---|
| | | | Ana Lucia Ferreira Ana Marisa Fusco Almeida Anderson Joel Martino Andrade André Luís Laforga Vanzela Andrea Duarte Doetzer Andrea Name Colado Simao Andressa Pereira de Souza Anelisa Ramão Angelica Beate Winter Boldt Anna Hermínia Castro Gomes de Amorim Anna Silvia Penteado Setti da Rocha Antonio Camilo da Silva Filho Antonio Stabelini Neto Arthur Hirata Bertachi Barbara Mendes Paz Chao Betty Cristiane Kuhn Bruno Ambrozio Galindo Bruno Ribeiro Cruz Camilla Reginatto De Pierri Carla Fredrichsen Moya Araujo Carla Fredrichsen Moya Araujo Carlos Alberto Oliveira de Biagi Junior Carlos Augusto Nassar Carlos Eduardo Buss Carlos Gilberto Carlotti Junior Carlos Henrique Schneider Carolina Panis Carolina Weigert Galvão Caroline de Jesus Coelho Donha Caroline Guisantes de Salvo Toni Caryna Eurich Mazur Catiuscie Cabreira da Silva Tortorella Celso F. D. Doliveira Cesar Luiz Boguszewski Christiane Pienna Soares Chung Man Chin Claudia Moro Cleverson Busso Cristiane Cominetti Daiane Priscila Simão-Silva Dalila Luciola Zanette Daniel de Paula Daniel de Paula Daniel Rech Daniela Fiori Gradia Daniela Pretti da Cunha Tirapelli Daniela Viganó Zanoti Jeronymo Daniele Ukan Danielle Malheiros Ferreira Danielle Venturini Deborah Catharine de Assis Leite Deivid Calebe de Souza Dennis Armando Bertolini Edenir Inez Pamero Edna Maria Vissoci Reiche Edson Roberto Arpini Miguel Eduardo José de Almeida Araújo Eliana Carolina Vespero Eliandro Reis Tavares Elza Kimura Grimshaw Emanuel Maltempi de Souza Emanuele Cristina Gustani Buss Émerson Carraro Emiliana Cristina Melo ENILze Maria de Souza Fonseca Ribeiro Enilze Maria de Souza Fonseca Ribeiro Erika Izumi Erika Seki Kioshima Cotica Evani Marques Pereira Fabio Negretti Fábio Rodrigues Ferreira Seiva Felipe Dunin dos Santos Felipe Tuon Fernanda Andreia Rosa Fernanda Cestaro Prado Cortez Fernanda Ivanski Fernanda Maris Peria Flavia Regina Oliveira de Barros Franciele Aní Caovilla Follador Franciele Mara Lucca Zanardo Bohm Francinete Ramos Campos Fulviana Silva Nishiyama GABRIEL RIBEIRO CORDEIRO Gabriela Datsch Bennemann Gisele Santos de Oliveira Glaucio Valdameri Glauco Akelinghton Freire Vitiello Glauco Vieira Miranda Glaura Scantamburlo ALves Fernandes Guilherme Ferreira Silveira Gustavo Bianchini Porfírio Gustavo Lenci Marques Hélito Volpato Hildebrando Masshiro Nagai Huei Diana Lee Ilce Mara de Syllos Cólus Iris Rabinovich Israel Gomy Jackson Kawakami Jacques Duílio Brancher Jaime Luís Lopes Rocha Jaqueline Carvalho de Oliveira Jean Henrique da Silva Rodrigues Jean Leandro dos Santos Jeane Eliete Laguila Visentainer João Paulo Bianchi Ximenez Joaquim Manoel da Silva Jociani Ascari Joel Donazzolo Jorge Luis Maria Ruiz Jose Knopfholz José Luis da Conceição Silva José Sebastião dos Santos Joseane Carla Schabarum Juliana Cheleski Wiggers Juliana Mara Serpeloni Juliana Morini Küpper Cardoso Perseguini Karen Brajão de Oliveira Karin Braun Prado Karine Aparecida de Lima Katiany Rizzieri Caleffi Ferracioli Katiuscia de Oliveira Francisco Gabriel Kelvinson Fernandes Viana Larissa Beatriz Cossalter Larissa Danielle Bahls Pinto Laurival Antonio Vilas Boas Léia Carolina Lucio Líbero Mezzadri Neto Ligia Carla Faccin Galhardi Lirane Elize Defante Ferreto Luciana Furlaneto Maia Luciana Oliveira de Fariña Luciana Reis Azevedo Alanis Luciane Regina Cavalli Lucy Megumi Yamauchi Lioni Luis Paulo Gomes Mascarenhas Luis Paulo Gomes Mascarenhas Luís Paulo Mascarenhas Lupe Furtado Alle Lyvia Regina Biagi Silva Bertachi Mara Antonia Ramos Costa Mara L. Cordeiro Marcela Maria Birolim Marcelo Ricardo Vicari Marcia Edilaine Lopes Consolaro Marcia Holsbach Beltrame Marcia Regina Eches Perugini Marcos Abdo Arbex Marcos Pileggi MARCOS TADEU GRZELCZAK Marcus Peikriszwili Tartaruga Maria Angelica Ehara Watanabe Maria Antonia Ramos Costa Maria Claudia Gross Maria José Soares Mendes Giannini Maria Leandra Terencio Maria Lúcia Bonfleur Maria Luiza Guimarães de Oliveira Maria Luiza Petzl-Erler Mariana Abe Vicente Cavagnari Marina Kimiko Kadowaki Marise Fonseca dos Santos Marla Karine Amarante Maurício Turkiewicz Mauro Antonio Alves Castro Michel Rodrigo Zambrano Passarini Michele Potrich Michelle Orane Schemberger Milena Massumi Kozonoe Mônica Degraf Cavallin Monica Tereza Suldofski Mucio Luiz de Assis Cirino Nadia Graciele Krohn Najeh Maissar Khalil Nédia de Castilhos Ghisi Neide Tomimura Costa Neiva Leite Neyva Maria Lopes Romeiro Patrícia Amâncio da Rosa Patricia Dayane Carvalho Schaker Patricia Oehlmeyer Nassar Patrícia Savio de Araújo-Souza Patricia Silva Lucio Paulo Henrique Couto Souza Paulo Roberto Donadio Percy Nohama Quirino Alves de Lima Neto Rafael Deminice Rafael dos Santos Bezerra Raquel Alves dos Santos Renan Manozzo Galante Renata Ernlund Freitas de Macedo Rita de Cássia Garcia Simão Roberta Losi Guembarovski Roberto H. Herai Roberto Rosati Rodrigo Ferreira Rodrigo Rodrigues Matiello Rogério Neri Shinsato Rogério Pincela Mateus Rosane Aparecida Ribeiro Rosilene Fressatti Cardoso Rosilene Fressatti Cardoso Sandra Mara Guse Scós Venske Selene Elifio Esposito Sérgio Ossamu Ioshii Silvana Giuliatti Silvia Mara de Souza Halick Silvio Henrique Maia de Almeida Simone Neumann Wendt Spencer Luiz Marques Payão Stefan Wolanski Negrão Stephane Janaina de Moura Escobar Sueli Fumie Yamada Ogatta SUELI PERCIO QUINAIA Taciane Finatto Tatiana Mayumi Veiga Iriyoda Tayza Katelline Danilau Ostroski Tony Alexander Hild Valeria Valente Vanessa Nascimento Kozak Vanessa Santos Sotomaior Victor Breno Pedrosa Victoria Zeghbi Cochenski Borba Vivian Rotuno Moure Valdameri Wander Rogerio Pavanelli Weber Cláudio Francisco Nunes da Silva Willian Augusto de Melo Yohandra Reyes Torres |
| EPI_ISL_2758680, EPI_ISL_2758689, EPI_ISL_2758690, EPI_ISL_2758691, EPI_ISL_2758697 | UEL | IPEC Guarapuava | NAPI-Genômica (Novos Arranjo de Pesquisa e Inovação em Genômica): Ademar Dantas da Cunha Júnior Adriano Ferrasa Adriano Mondini Aldo Przybysz Alessandra Lourenço Cecchini Armani Alex Sandro Jorge Alexandra Ivo de Medeiros Alexandre Maller Aline Cristina Batista Rodrigues Johann Ana Lucia Ferreira Ana Marisa Fusco Almeida Anderson Joel Martino Andrade André Luís Laforga Vanzela Andrea Duarte Doetzer Andrea Name Colado Simao Andressa Pereira de Souza Anelisa Ramão Angelica Beate Winter Boldt Anna Hermínia Castro Gomes de Amorim Anna Silvia Penteado Setti da Rocha Antonio Camilo da Silva Filho Antonio Stabelini Neto Arthur Hirata Bertachi Barbara Mendes Paz Chao Betty Cristiane Kuhn Bruno Ambrozio Galindo Bruno Ribeiro Cruz Camilla Reginatto De Pierri Carla Fredrichsen Moya Araujo Carla Fredrichsen Moya Araujo Carlos Alberto Oliveira de Biagi Junior Carlos Augusto Nassar Carlos Eduardo Buss Carlos Gilberto Carlotti Junior Carlos Henrique Schneider Carolina Panis Carolina Weigert Galvão Caroline de Jesus Coelho Donha Caroline Guisantes de Salvo Toni Caryna Eurich Mazur Catiuscie Cabreira da Silva Tortorella Celso F. D. Doliveira Cesar Luiz Boguszewski Christiane Pienna Soares Chung Man Chin Claudia Moro Cleverson Busso Cristiane Cominetti Daiane Priscila Simão-Silva Dalila Luciola Zanette Daniel de Paula Daniel de Paula Daniel Rech Daniela Fiori Gradia Daniela Pretti da Cunha Tirapelli Daniela Viganó Zanoti Jeronymo Daniele Ukan Danielle Malheiros Ferreira Danielle Venturini Deborah Catharine de Assis Leite Deivid Calebe de Souza Dennis Armando Bertolini Edenir Inez Pamero Edna Maria Vissoci Reiche Edson Roberto Arpini Miguel Eduardo José de Almeida Araújo Eliana Carolina Vespero Eliandro Reis Tavares Elza Kimura Grimshaw Emanuel Maltempi de Souza Emanuele Cristina Gustani Buss Émerson Carraro Emiliana Cristina Melo ENILze Maria de Souza Fonseca Ribeiro Enilze Maria de Souza Fonseca Ribeiro Erika Izumi Erika Seki Kioshima Cotica Evani Marques Pereira Fabio Negretti Fábio Rodrigues Ferreira Seiva Felipe Dunin dos Santos Felipe Tuon Fernanda Andreia Rosa Fernanda Cestaro Prado Cortez Fernanda Ivanski Fernanda Maris Peria Flavia Regina Oliveira de Barros Franciele Aní Caovilla Follador Franciele Mara Lucca Zanardo Bohm Francinete Ramos Campos Fulviana Silva Nishiyama GABRIEL RIBEIRO CORDEIRO Gabriela Datsch Bennemann Gisele Santos de Oliveira Glaucio Valdameri Glauco Akelinghton Freire Vitiello Glauco Vieira Miranda Glaura Scantamburlo ALves Fernandes Guilherme Ferreira Silveira Gustavo Bianchini Porfírio Gustavo Lenci Marques Hélito Volpato Hildebrando Masshiro Nagai Huei Diana Lee Ilce Mara de Syllos Cólus Iris Rabinovich Israel Gomy Jackson Kawakami Jacques Duílio Brancher Jaime Luís Lopes Rocha Jaqueline Carvalho de Oliveira Jean Henrique da Silva Rodrigues Jean Leandro dos Santos Jeane Eliete Laguila Visentainer João Paulo Bianchi Ximenez Joaquim Manoel da Silva Jociani Ascari Joel Donazzolo Jorge Luis Maria Ruiz Jose Knopfholz José Luis da Conceição Silva José Sebastião dos Santos Joseane Carla Schabarum Juliana Cheleski Wiggers Juliana Mara Serpeloni Juliana Morini Küpper Cardoso Perseguini Karen Brajão de Oliveira Karin Braun Prado Karine Aparecida de Lima Katiany Rizzieri Caleffi Ferracioli Katiuscia de Oliveira Francisco Gabriel Kelvinson Fernandes Viana Larissa Beatriz Cossalter Larissa Danielle Bahls Pinto Laurival Antonio Vilas Boas Léia Carolina Lucio Líbero Mezzadri Neto Ligia Carla Faccin Galhardi Lirane Elize Defante Ferreto Luciana Furlaneto Maia Luciana Oliveira de Fariña Luciana Reis Azevedo Alanis Luciane Regina Cavalli Lucy Megumi Yamauchi Lioni Luis Paulo Gomes Mascarenhas Luis Paulo Gomes Mascarenhas Luís Paulo Mascarenhas Lupe Furtado Alle Lyvia Regina Biagi Silva Bertachi Mara Antonia Ramos Costa Mara L. Cordeiro Marcela Maria Birolim Marcelo Ricardo Vicari Marcia Edilaine Lopes Consolaro Marcia Holsbach Beltrame Marcia Regina Eches Perugini Marcos Abdo Arbex Marcos Pileggi MARCOS TADEU GRZELCZAK Marcus Peikriszwili Tartaruga Maria Angelica Ehara Watanabe Maria Antonia Ramos Costa Maria Claudia Gross Maria José Soares Mendes Giannini Maria Leandra Terencio Maria Lúcia Bonfleur Maria Luiza Guimarães de Oliveira Maria Luiza Petzl-Erler Mariana Abe Vicente Cavagnari Marina Kimiko Kadowaki Marise Fonseca dos Santos Marla Karine Amarante Maurício Turkiewicz Mauro Antonio Alves Castro Michel Rodrigo Zambrano Passarini Michele Potrich Michelle Orane Schemberger Milena Massumi Kozonoe Mônica Degraf Cavallin Monica Tereza Suldofski Mucio Luiz de Assis Cirino Nadia Graciele Krohn Najeh Maissar Khalil Nédia de Castilhos Ghisi Neide Tomimura Costa Neiva Leite Neyva Maria Lopes Romeiro Patrícia Amâncio da Rosa Patricia Dayane Carvalho Schaker Patricia Oehlmeyer Nassar Patrícia Savio de Araújo-Souza Patricia Silva Lucio Paulo Henrique Couto Souza Paulo Roberto Donadio Percy Nohama Quirino Alves de Lima Neto Rafael Deminice Rafael dos Santos Bezerra Raquel Alves dos Santos Renan Manozzo Galante Renata Ernlund Freitas de Macedo Rita de Cássia Garcia Simão Roberta Losi Guembarovski Roberto H. Herai Roberto Rosati Rodrigo Ferreira Rodrigo Rodrigues Matiello Rogério Neri Shinsato Rogério Pincela Mateus Rosane Aparecida Ribeiro Rosilene Fressatti Cardoso Rosilene Fressatti Cardoso Sandra Mara Guse Scós Venske Selene Elifio Esposito Sérgio Ossamu Ioshii Silvana Giuliatti Silvia Mara de Souza Halick Silvio Henrique Maia de Almeida Simone Neumann Wendt Spencer Luiz Marques Payão Stefan Wolanski Negrão Stephane Janaina de Moura Escobar Sueli Fumie Yamada Ogatta SUELI PERCIO QUINAIA Taciane Finatto Tatiana Mayumi Veiga Iriyoda Tayza Katelline Danilau Ostroski Tony Alexander Hild Valeria Valente Vanessa Nascimento Kozak Vanessa Santos Sotomaior Victor Breno Pedrosa Victoria Zeghbi Cochenski Borba Vivian Rotuno Moure Valdameri Wander Rogerio Pavanelli Weber Cláudio Francisco Nunes da Silva Willian Augusto de Melo Yohandra Reyes Torres |
| EPI_ISL_2758699, EPI_ISL_2758700, EPI_ISL_2758702, EPI_ISL_2758703, EPI_ISL_2758704, EPI_ISL_2758708, EPI_ISL_2758712 | HUEM/IBMP | IPEC Guarapuava | NAPI-Genômica (Novos Arranjo de Pesquisa e Inovação em Genômica): Ademar Dantas da Cunha Júnior Adriano Ferrasa Adriano Mondini Aldo Przybysz Alessandra Lourenço Cecchini Armani Alex Sandro Jorge Alexandra Ivo de Medeiros Alexandre Maller Aline Cristina Batista Rodrigues Johann Ana Lucia Ferreira Ana Marisa Fusco Almeida Anderson Joel Martino Andrade André Luís Laforga Vanzela Andrea Duarte Doetzer Andrea Name Colado Simao Andressa Pereira de Souza Anelisa Ramão Angelica Beate Winter Boldt Anna Hermínia Castro Gomes de Amorim Anna Silvia Penteado Setti da |

| | | | |
|---|---|---|---|
| | | | Rocha Antonio Camilo da Silva Filho Antonio Stabelini Neto Arthur Hirata Bertachi Barbara Mendes Paz Chao Betty Cristiane Kuhn Bruno Ambrozio Galindo Bruno Ribeiro Cruz Camilla Reginatto De Pierri Carla Fredrichsen Moya Araujo Carla Fredrichsen Moya Araujo Carlos Alberto Oliveira de Biagi Junior Carlos Augusto Nassar Carlos Eduardo Buss Carlos Gilberto Carlotti Junior Carlos Henrique Schneider Carolina Panis Carolina Weigert Galvão Caroline de Jesus Coelho Donha Caroline Guisantes de Salvo Toni Caryna Eurich Mazur Catiuscie Cabreira da Silva Tortorella Celso F. D. Doliveira Cesar Luiz Boguszewski Christiane Pienna Soares Chung Man Chin Claudia Moro Cleverson Busso Cristiane Cominetti Daiane Priscila Simão-Silva Dalila Luciola Zanette Daniel de Paula Daniel de Paula Daniel Rech Daniela Fiori Gradia Daniela Pretti da Cunha Tirapelli Daniela Viganó Zanoti Jeronymo Daniele Ukan Danielle Malheiros Ferreira Danielle Venturini Deborah Catharine de Assis Leite Deivid Calebe de Souza Dennis Armando Bertolini Edenir Inez Pamero Edna Maria Vissoci Reiche Edson Roberto Arpini Miguel Eduardo José de Almeida Araújo Eliana Carolina Vespero Eliandro Reis Tavares Elza Kimura Grimshaw Emanuel Maltempi de Souza Emanuele Cristina Gustani Buss Émerson Carraro Emiliana Cristina Melo ENILze Maria de Souza Fonseca Ribeiro Enilze Maria de Souza Fonseca Ribeiro Erika Izumi Erika Seki Kioshima Cotica Evani Marques Pereira Fabio Negretti Fábio Rodrigues Ferreira Seiva Felipe Dunin dos Santos Felipe Tuon Fernanda Andreia Rosa Fernanda Cestaro Prado Cortez Fernanda Ivanski Fernanda Maris Peria Flavia Regina Oliveira de Barros Franciele Aní Caovilla Follador Franciele Mara Lucca Zanardo Bohm Francinete Ramos Campos Fulviana Silva Nishiyama GABRIEL RIBEIRO CORDEIRO Gabriela Datsch Bennemann Gisele Santos de Oliveira Glaucio Valdameri Glauco Akelinghton Freire Vitiello Glauco Vieira Miranda Glaura Scantamburlo ALves Fernandes Guilherme Ferreira Silveira Gustavo Bianchini Porfírio Gustavo Lenci Marques Hélito Volpato Hildebrando Masshiro Nagai Huei Diana Lee Ilce Mara de Syllos Cólus Iris Rabinovich Israel Gomy Jackson Kawakami Jacques Duílio Brancher Jaime Luís Lopes Rocha Jaqueline Carvalho de Oliveira Jean Henrique da Silva Rodrigues Jean Leandro dos Santos Jeane Eliete Laguila Visentainer João Paulo Bianchi Ximenez Joaquim Manoel da Silva Jociani Ascari Joel Donazzolo Jorge Luis Maria Ruiz Jose Knopfholz José Luis da Conceição Silva José Sebastião dos Santos Joseane Carla Schabarum Juliana Cheleski Wiggers Juliana Mara Serpeloni Juliana Morini Küpper Cardoso Perseguini Karen Brajão de Oliveira Karin Braun Prado Karine Aparecida de Lima Katiany Rizzieri Caleffi Ferracioli Katiuscia de Oliveira Francisco Gabriel Kelvinson Fernandes Viana Larissa Beatriz Cossalter Larissa Danielle Bahls Pinto Laurival Antonio Vilas Boas Léia Carolina Lucio Líbero Mezzadri Neto Ligia Carla Faccin Galhardi Lirane Elize Defante Ferreto Luciana Furlaneto Maia Luciana Oliveira de Fariña Luciana Reis Azevedo Alanis Luciane Regina Cavalli Lucy Megumi Yamauchi Lioni Luis Paulo Gomes Mascarenhas Luis Paulo Gomes Mascarenhas Luís Paulo Mascarenhas Lupe Furtado Alle Lyvia Regina Biagi Silva Bertachi Mara Antonia Ramos Costa Mara L. Cordeiro Marcela Maria Birolim Marcelo Ricardo Vicari Marcia Edilaine Lopes Consolaro Marcia Holsbach Beltrame Marcia Regina Eches Perugini Marcos Abdo Arbex Marcos Pileggi MARCOS TADEU GRZELCZAK Marcus Peikriszwili Tartaruga Maria Angelica Ehara Watanabe Maria Antonia Ramos Costa Maria Claudia Gross Maria José Soares Mendes Giannini Maria Leandra Terencio Maria Lúcia Bonfleur Maria Luiza Guimarães de Oliveira Maria Luiza Petzl-Erler Mariana Abe Vicente Cavagnari Marina Kimiko Kadowaki Marise Fonseca dos Santos Marla Karine Amarante Maurício Turkiewicz Mauro Antonio Alves Castro Michel Rodrigo Zambrano Passarini Michele Potrich Michelle Orane Schemberger Milena Massumi Kozonoe Mônica Degraf Cavallin Monica Tereza Suldofski Mucio Luiz de Assis Cirino Nadia Graciele Krohn Najeh Maissar Khalil Nédia de Castilhos Ghisi Neide Tomimura Costa Neiva Leite Neyva Maria Lopes Romeiro Patrícia Amâncio da Rosa Patricia Dayane Carvalho Schaker Patricia Oehlmeyer Nassar Patrícia Savio de Araújo-Souza Patricia Silva Lucio Paulo Henrique Couto Souza Paulo Roberto Donadio Percy Nohama Quirino Alves de Lima Neto Rafael Deminice Rafael dos Santos Bezerra Raquel Alves dos Santos Renan Manozzo Galante Renata Ernlund Freitas de Macedo Rita de Cássia Garcia Simão Roberta Losi Guembarovski Roberto H. Herai Roberto Rosati Rodrigo Ferreira Rodrigo Rodrigues Matiello Rogério Neri Shinsato Rogério Pincela Mateus Rosane Aparecida Ribeiro Rosilene Fressatti Cardoso Rosilene Fressatti Cardoso Sandra Mara Guse Scós Venske Selene Elifio Esposito Sérgio Ossamu Ioshii Silvana Giuliatti Silvia Mara de Souza Halick Silvio Henrique Maia de Almeida Simone Neumann Wendt Spencer Luiz Marques Payão Stefan Wolanski Negrão Stephane Janaina de Moura Escobar Sueli Fumie Yamada Ogatta SUELI PERCIO QUINAIA Taciane Finatto Tatiana Mayumi Veiga Iriyoda Tayza Katelline Danilau Ostroski Tony Alexander Hild Valeria Valente Vanessa Nascimento Kozak Vanessa Santos Sotomaior Victor Breno Pedrosa Victoria Zeghbi Cochenski Borba Vivian Rotuno Moure Valdameri Wander Rogerio Pavanelli Weber Cláudio Francisco Nunes da Silva Willian Augusto de Melo Yohandra Reyes Torres |
| EPI_ISL_2758715, EPI_ISL_2758721, EPI_ISL_2758731, EPI_ISL_2758738, EPI_ISL_2758751, EPI_ISL_2758753, EPI_ISL_2758755, EPI_ISL_2758766 | UEL | IPEC Guarapuava | NAPI-Genômica (Novos Arranjo de Pesquisa e Inovação em Genômica): Ademar Dantas da Cunha Júnior Adriano Ferrasa Adriano Mondini Aldo Przybysz Alessandra Lourenço Cecchini Armani Alex Sandro Jorge Alexandra Ivo de Medeiros Alexandre Maller Aline Cristina Batista Rodrigues Johann Ana Lucia Ferreira Ana Marisa Fusco Almeida Anderson Joel Martino Andrade André Luís Laforga Vanzela Andrea Duarte Doetzer Andrea Name Colado Simao Andressa Pereira de Souza Anelisa Ramão Angelica Beate Winter Boldt Anna Hermínia Castro Gomes de Amorim Anna Silvia Penteado Setti da Rocha Antonio Camilo da Silva Filho Antonio Stabelini Neto Arthur Hirata Bertachi Barbara Mendes Paz Chao Betty Cristiane Kuhn Bruno Ambrozio Galindo Bruno Ribeiro Cruz Camilla Reginatto De Pierri Carla Fredrichsen Moya Araujo Carla Fredrichsen Moya Araujo Carlos Alberto Oliveira de Biagi Junior Carlos Augusto Nassar Carlos Eduardo Buss Carlos Gilberto Carlotti Junior Carlos Henrique Schneider Carolina Panis Carolina Weigert Galvão Caroline de Jesus Coelho Donha Caroline Guisantes de Salvo Toni Caryna Eurich Mazur Catiuscie Cabreira da Silva Tortorella Celso F. D. Doliveira Cesar Luiz Boguszewski Christiane Pienna Soares Chung Man Chin Claudia Moro Cleverson Busso Cristiane Cominetti Daiane Priscila Simão-Silva Dalila Luciola Zanette Daniel de Paula Daniel de Paula Daniel Rech Daniela Fiori Gradia Daniela Pretti da Cunha Tirapelli Daniela Viganó Zanoti Jeronymo Daniele Ukan Danielle Malheiros Ferreira Danielle Venturini Deborah Catharine de Assis Leite Deivid Calebe de Souza Dennis Armando Bertolini Edenir Inez Pamero Edna Maria Vissoci Reiche Edson Roberto Arpini Miguel Eduardo José de Almeida Araújo Eliana Carolina Vespero Eliandro Reis Tavares Elza Kimura Grimshaw Emanuel Maltempi de Souza Emanuele Cristina Gustani Buss Émerson Carraro Emiliana Cristina Melo ENILze Maria de Souza Fonseca Ribeiro Enilze Maria de Souza Fonseca Ribeiro Erika Izumi Erika Seki Kioshima Cotica Evani Marques Pereira Fabio Negretti Fábio Rodrigues Ferreira Seiva Felipe Dunin dos Santos Felipe Tuon Fernanda Andreia Rosa Fernanda Cestaro Prado Cortez Fernanda Ivanski Fernanda Maris Peria Flavia Regina Oliveira de Barros Franciele Aní Caovilla Follador Franciele Mara Lucca Zanardo Bohm Francinete Ramos Campos Fulviana Silva Nishiyama GABRIEL RIBEIRO CORDEIRO Gabriela Datsch Bennemann Gisele Santos de Oliveira Glaucio Valdameri Glauco Akelinghton Freire Vitiello Glauco Vieira Miranda Glaura Scantamburlo ALves Fernandes Guilherme Ferreira Silveira Gustavo Bianchini Porfírio Gustavo Lenci Marques Hélito Volpato Hildebrando Masshiro Nagai Huei Diana Lee Ilce Mara de Syllos Cólus Iris Rabinovich Israel Gomy Jackson Kawakami Jacques Duílio Brancher Jaime Luís Lopes Rocha Jaqueline Carvalho de Oliveira Jean Henrique da Silva Rodrigues Jean Leandro dos Santos Jeane Eliete Laguila Visentainer João Paulo Bianchi Ximenez Joaquim Manoel da Silva Jociani Ascari Joel Donazzolo Jorge Luis Maria Ruiz Jose Knopfholz José Luis da Conceição Silva José Sebastião dos Santos Joseane Carla Schabarum Juliana Cheleski Wiggers Juliana Mara Serpeloni Juliana Morini Küpper Cardoso Perseguini Karen Brajão de Oliveira Karin Braun Prado Karine Aparecida de Lima Katiany Rizzieri Caleffi Ferracioli Katiuscia de Oliveira Francisco Gabriel Kelvinson Fernandes Viana Larissa Beatriz Cossalter Larissa Danielle Bahls Pinto Laurival Antonio Vilas Boas Léia Carolina Lucio Líbero Mezzadri Neto Ligia Carla Faccin Galhardi Lirane Elize Defante Ferreto Luciana Furlaneto Maia Luciana Oliveira de Fariña Luciana Reis Azevedo Alanis Luciane Regina Cavalli Lucy Megumi Yamauchi Lioni Luis Paulo Gomes Mascarenhas Luis Paulo Gomes Mascarenhas Luís Paulo Mascarenhas Lupe Furtado Alle Lyvia Regina Biagi Silva Bertachi Mara Antonia Ramos Costa Mara L. Cordeiro Marcela Maria Birolim Marcelo Ricardo Vicari Marcia Edilaine Lopes Consolaro Marcia Holsbach Beltrame Marcia Regina Eches Perugini Marcos Abdo Arbex Marcos Pileggi MARCOS TADEU GRZELCZAK Marcus Peikriszwili Tartaruga Maria Angelica Ehara Watanabe Maria Antonia Ramos Costa Maria Claudia Gross Maria José Soares Mendes Giannini Maria Leandra Terencio Maria Lúcia Bonfleur Maria Luiza Guimarães de Oliveira Maria Luiza Petzl-Erler Mariana Abe Vicente Cavagnari Marina Kimiko Kadowaki Marise Fonseca dos Santos Marla Karine Amarante Maurício Turkiewicz Mauro Antonio Alves Castro Michel Rodrigo Zambrano Passarini Michele Potrich Michelle Orane Schemberger Milena Massumi Kozonoe Mônica Degraf Cavallin Monica Tereza Suldofski Mucio Luiz de Assis Cirino Nadia Graciele Krohn Najeh Maissar Khalil Nédia de Castilhos Ghisi Neide Tomimura Costa Neiva Leite Neyva Maria Lopes Romeiro Patrícia Amâncio da Rosa Patricia Dayane Carvalho Schaker Patricia Oehlmeyer Nassar Patrícia Savio de Araújo-Souza Patricia Silva Lucio Paulo Henrique Couto Souza Paulo Roberto Donadio Percy Nohama Quirino Alves de Lima Neto Rafael Deminice Rafael dos Santos Bezerra Raquel Alves dos Santos Renan Manozzo Galante Renata Ernlund Freitas de Macedo Rita de Cássia Garcia Simão Roberta Losi Guembarovski Roberto H. Herai Roberto Rosati Rodrigo Ferreira Rodrigo Rodrigues Matiello Rogério Neri Shinsato Rogério Pincela Mateus Rosane Aparecida Ribeiro Rosilene Fressatti Cardoso Rosilene Fressatti Cardoso Sandra Mara Guse Scós Venske Selene Elifio Esposito Sérgio Ossamu Ioshii Silvana Giuliatti Silvia Mara de Souza Halick Silvio Henrique Maia de Almeida Simone Neumann Wendt Spencer Luiz Marques Payão Stefan Wolanski Negrão Stephane Janaina de Moura Escobar Sueli Fumie Yamada Ogatta SUELI PERCIO QUINAIA Taciane Finatto Tatiana Mayumi Veiga Iriyoda Tayza Katelline Danilau Ostroski Tony Alexander Hild Valeria Valente Vanessa Nascimento Kozak Vanessa Santos Sotomaior Victor Breno Pedrosa Victoria Zeghbi Cochenski Borba Vivian Rotuno Moure Valdameri Wander Rogerio Pavanelli Weber Cláudio Francisco Nunes da Silva Willian Augusto de Melo Yohandra Reyes Torres |
| EPI_ISL_2758767, EPI_ISL_2758768, EPI_ISL_2758769, EPI_ISL_2758770 | HUEM/IBMP | IPEC Guarapuava | NAPI-Genômica (Novos Arranjo de Pesquisa e Inovação em Genômica): Ademar Dantas da Cunha Júnior Adriano Ferrasa Adriano Mondini Aldo Przybysz Alessandra Lourenço Cecchini Armani Alex Sandro Jorge Alexandra Ivo de Medeiros Alexandre Maller Aline Cristina Batista Rodrigues Johann Ana Lucia Ferreira Ana Marisa Fusco Almeida Anderson Joel Martino Andrade André Luís Laforga Vanzela Andrea Duarte Doetzer Andrea Name Colado Simao Andressa Pereira de Souza Anelisa Ramão Angelica Beate Winter Boldt Anna Hermínia Castro Gomes de Amorim Anna Silvia Penteado Setti da Rocha Antonio Camilo da Silva Filho Antonio Stabelini Neto Arthur Hirata Bertachi Barbara Mendes Paz Chao Betty Cristiane Kuhn Bruno Ambrozio Galindo Bruno Ribeiro Cruz Camilla Reginatto De Pierri Carla Fredrichsen Moya Araujo Carla Fredrichsen Moya Araujo Carlos Alberto Oliveira de Biagi |

| | | | |
|---|---|---|---|
| | | | Junior Carlos Augusto Nassar Carlos Eduardo Buss Carlos Gilberto Carlotti Junior Carlos Henrique Schneider Carolina Panis Carolina Weigert Galvão Caroline de Jesus Coelho Donha Caroline Guisantes de Salvo Toni Caryna Eurich Mazur Catiuscie Cabreira da Silva Tortorella Celso F. D. Doliveira Cesar Luiz Boguszewski Christiane Pienna Soares Chung Man Chin Claudia Moro Cleverson Busso Cristiane Cominetti Daiane Priscila Simão-Silva Dalila Luciola Zanette Daniel de Paula Daniel de Paula Daniel Rech Daniela Fiori Gradia Daniela Pretti da Cunha Tirapelli Daniela Viganó Zanoti Jeronymo Daniele Ukan Danielle Malheiros Ferreira Danielle Venturini Deborah Catharine de Assis Leite Deivid Calebe de Souza Dennis Armando Bertolini Edenir Inez Pamero Edna Maria Vissoci Reiche Edson Roberto Arpini Miguel Eduardo José de Almeida Araújo Eliana Carolina Vespero Eliandro Reis Tavares Elza Kimura Grimshaw Emanuel Maltempi de Souza Emanuele Cristina Gustani Buss Émerson Carraro Emiliana Cristina Melo ENILze Maria de Souza Fonseca Ribeiro Enilze Maria de Souza Fonseca Ribeiro Erika Izumi Erika Seki Kioshima Cotica Evani Marques Pereira Fabio Negretti Fábio Rodrigues Ferreira Seiva Felipe Dunin dos Santos Felipe Tuon Fernanda Andreia Rosa Fernanda Cestaro Prado Cortez Fernanda Ivanski Fernanda Maris Peria Flavia Regina Oliveira de Barros Franciele Aní Caovilla Follador Franciele Mara Lucca Zanardo Bohm Francinete Ramos Campos Fulviana Silva Nishiyama GABRIEL RIBEIRO CORDEIRO Gabriela Datsch Bennemann Gisele Santos de Oliveira Glaucio Valdameri Glauco Akelinghton Freire Vitiello Glauco Vieira Miranda Glaura Scantamburlo ALves Fernandes Guilherme Ferreira Silveira Gustavo Bianchini Porfírio Gustavo Lenci Marques Hélito Volpato Hildebrando Masshiro Nagai Huei Diana Lee Ilce Mara de Syllos Cólus Iris Rabinovich Israel Gomy Jackson Kawakami Jacques Duílio Brancher Jaime Luís Lopes Rocha Jaqueline Carvalho de Oliveira Jean Henrique da Silva Rodrigues Jean Leandro dos Santos Jeane Eliete Laguila Visentainer João Paulo Bianchi Ximenez Joaquim Manoel da Silva Jociani Ascari Joel Donazzolo Jorge Luis Maria Ruiz Jose Knopfholz José Luis da Conceição Silva José Sebastião dos Santos Joseane Carla Schabarum Juliana Cheleski Wiggers Juliana Mara Serpeloni Juliana Morini Küpper Cardoso Perseguini Karen Brajão de Oliveira Karin Braun Prado Karine Aparecida de Lima Katiany Rizzieri Caleffi Ferracioli Katiuscia de Oliveira Francisco Gabriel Kelvinson Fernandes Viana Larissa Beatriz Cossalter Larissa Danielle Bahls Pinto Laurival Antonio Vilas Boas Léia Carolina Lucio Líbero Mezzadri Neto Ligia Carla Faccin Galhardi Lirane Elize Defante Ferreto Luciana Furlaneto Maia Luciana Oliveira de Fariña Luciana Reis Azevedo Alanis Luciane Regina Cavalli Lucy Megumi Yamauchi Lioni Luis Paulo Gomes Mascarenhas Luis Paulo Gomes Mascarenhas Luís Paulo Mascarenhas Lupe Furtado Alle Lyvia Regina Biagi Silva Bertachi Mara Antonia Ramos Costa Mara L. Cordeiro Marcela Maria Birolim Marcelo Ricardo Vicari Marcia Edilaine Lopes Consolaro Marcia Holsbach Beltrame Marcia Regina Eches Perugini Marcos Abdo Arbex Marcos Pileggi MARCOS TADEU GRZELCZAK Marcus Peikriszwili Tartaruga Maria Angelica Ehara Watanabe Maria Antonia Ramos Costa Maria Claudia Gross Maria José Soares Mendes Giannini Maria Leandra Terencio Maria Lúcia Bonfleur Maria Luiza Guimarães de Oliveira Maria Luiza Petzl-Erler Mariana Abe Vicente Cavagnari Marina Kimiko Kadowaki Marise Fonseca dos Santos Marla Karine Amarante Maurício Turkiewicz Mauro Antonio Alves Castro Michel Rodrigo Zambrano Passarini Michele Potrich Michelle Orane Schemberger Milena Massumi Kozonoe Mônica Degraf Cavallin Monica Tereza Suldofski Mucio Luiz de Assis Cirino Nadia Graciele Krohn Najeh Maissar Khalil Nédia de Castilhos Ghisi Neide Tomimura Costa Neiva Leite Neyva Maria Lopes Romeiro Patrícia Amâncio da Rosa Patricia Dayane Carvalho Schaker Patricia Oehlmeyer Nassar Patrícia Savio de Araújo-Souza Patricia Silva Lucio Paulo Henrique Couto Souza Paulo Roberto Donadio Percy Nohama Quirino Alves de Lima Neto Rafael Deminice Rafael dos Santos Bezerra Raquel Alves dos Santos Renan Manozzo Galante Renata Ernlund Freitas de Macedo Rita de Cássia Garcia Simão Roberta Losi Guembarovski Roberto H. Herai Roberto Rosati Rodrigo Ferreira Rodrigo Rodrigues Matiello Rogério Neri Shinsato Rogério Pincela Mateus Rosane Aparecida Ribeiro Rosilene Fressatti Cardoso Rosilene Fressatti Cardoso Sandra Mara Guse Scós Venske Selene Elifio Esposito Sérgio Ossamu Ioshii Silvana Giuliatti Silvia Mara de Souza Halick Silvio Henrique Maia de Almeida Simone Neumann Wendt Spencer Luiz Marques Payão Stefan Wolanski Negrão Stephane Janaina de Moura Escobar Sueli Fumie Yamada Ogatta SUELI PERCIO QUINAIA Taciane Finatto Tatiana Mayumi Veiga Iriyoda Tayza Katelline Danilau Ostroski Tony Alexander Hild Valeria Valente Vanessa Nascimento Kozak Vanessa Santos Sotomaior Victor Breno Pedrosa Victoria Zeghbi Cochenski Borba Vivian Rotuno Moure Valdameri Wander Rogerio Pavanelli Weber Cláudio Francisco Nunes da Silva Willian Augusto de Melo Yohandra Reyes Torres |
| EPI_ISL_2758771 | HUEM/LACEN | IPEC Guarapuava | NAPI-Genômica (Novos Arranjo de Pesquisa e Inovação em Genômica): Ademar Dantas da Cunha Júnior Adriano Ferrasa Adriano Mondini Aldo Przybysz Alessandra Lourenço Cecchini Armani Alex Sandro Jorge Alexandra Ivo de Medeiros Alexandre Maller Aline Cristina Batista Rodrigues Johann Ana Lucia Ferreira Ana Marisa Fusco Almeida Anderson Joel Martino Andrade André Luís Laforga Vanzela Andrea Duarte Doetzer Andrea Name Colado Simao Andressa Pereira de Souza Anelisa Ramão Angelica Beate Winter Boldt Anna Hermínia Castro Gomes de Amorim Anna Silvia Penteado Setti da Rocha Antonio Camilo da Silva Filho Antonio Stabelini Neto Arthur Hirata Bertachi Barbara Mendes Paz Chao Betty Cristiane Kuhn Bruno Ambrozio Galindo Bruno Ribeiro Cruz Camilla Reginatto De Pierri Carla Fredrichsen Moya Araujo Carla Fredrichsen Moya Araujo Carlos Alberto Oliveira de Biagi Junior Carlos Augusto Nassar Carlos Eduardo Buss Carlos Gilberto Carlotti Junior Carlos Henrique Schneider Carolina Panis Carolina Weigert Galvão Caroline de Jesus Coelho Donha Caroline Guisantes de Salvo Toni Caryna Eurich Mazur Catiuscie Cabreira da Silva Tortorella Celso F. D. Doliveira Cesar Luiz Boguszewski Christiane Pienna Soares Chung Man Chin Claudia Moro Cleverson Busso Cristiane Cominetti Daiane Priscila Simão-Silva Dalila Luciola Zanette Daniel de Paula Daniel de Paula Daniel Rech Daniela Fiori Gradia Daniela Pretti da Cunha Tirapelli Daniela Viganó Zanoti Jeronymo Daniele Ukan Danielle Malheiros Ferreira Danielle Venturini Deborah Catharine de Assis Leite Deivid Calebe de Souza Dennis Armando Bertolini Edenir Inez Pamero Edna Maria Vissoci Reiche Edson Roberto Arpini Miguel Eduardo José de Almeida Araújo Eliana Carolina Vespero Eliandro Reis Tavares Elza Kimura Grimshaw Emanuel Maltempi de Souza Emanuele Cristina Gustani Buss Émerson Carraro Emiliana Cristina Melo ENILze Maria de Souza Fonseca Ribeiro Enilze Maria de Souza Fonseca Ribeiro Erika Izumi Erika Seki Kioshima Cotica Evani Marques Pereira Fabio Negretti Fábio Rodrigues Ferreira Seiva Felipe Dunin dos Santos Felipe Tuon Fernanda Andreia Rosa Fernanda Cestaro Prado Cortez Fernanda Ivanski Fernanda Maris Peria Flavia Regina Oliveira de Barros Franciele Aní Caovilla Follador Franciele Mara Lucca Zanardo Bohm Francinete Ramos Campos Fulviana Silva Nishiyama GABRIEL RIBEIRO CORDEIRO Gabriela Datsch Bennemann Gisele Santos de Oliveira Glaucio Valdameri Glauco Akelinghton Freire Vitiello Glauco Vieira Miranda Glaura Scantamburlo ALves Fernandes Guilherme Ferreira Silveira Gustavo Bianchini Porfírio Gustavo Lenci Marques Hélito Volpato Hildebrando Masshiro Nagai Huei Diana Lee Ilce Mara de Syllos Cólus Iris Rabinovich Israel Gomy Jackson Kawakami Jacques Duílio Brancher Jaime Luís Lopes Rocha Jaqueline Carvalho de Oliveira Jean Henrique da Silva Rodrigues Jean Leandro dos Santos Jeane Eliete Laguila Visentainer João Paulo Bianchi Ximenez Joaquim Manoel da Silva Jociani Ascari Joel Donazzolo Jorge Luis Maria Ruiz Jose Knopfholz José Luis da Conceição Silva José Sebastião dos Santos Joseane Carla Schabarum Juliana Cheleski Wiggers Juliana Mara Serpeloni Juliana Morini Küpper Cardoso Perseguini Karen Brajão de Oliveira Karin Braun Prado Karine Aparecida de Lima Katiany Rizzieri Caleffi Ferracioli Katiuscia de Oliveira Francisco Gabriel Kelvinson Fernandes Viana Larissa Beatriz Cossalter Larissa Danielle Bahls Pinto Laurival Antonio Vilas Boas Léia Carolina Lucio Líbero Mezzadri Neto Ligia Carla Faccin Galhardi Lirane Elize Defante Ferreto Luciana Furlaneto Maia Luciana Oliveira de Fariña Luciana Reis Azevedo Alanis Luciane Regina Cavalli Lucy Megumi Yamauchi Lioni Luis Paulo Gomes Mascarenhas Luis Paulo Gomes Mascarenhas Luís Paulo Mascarenhas Lupe Furtado Alle Lyvia Regina Biagi Silva Bertachi Mara Antonia Ramos Costa Mara L. Cordeiro Marcela Maria Birolim Marcelo Ricardo Vicari Marcia Edilaine Lopes Consolaro Marcia Holsbach Beltrame Marcia Regina Eches Perugini Marcos Abdo Arbex Marcos Pileggi MARCOS TADEU GRZELCZAK Marcus Peikriszwili Tartaruga Maria Angelica Ehara Watanabe Maria Antonia Ramos Costa Maria Claudia Gross Maria José Soares Mendes Giannini Maria Leandra Terencio Maria Lúcia Bonfleur Maria Luiza Guimarães de Oliveira Maria Luiza Petzl-Erler Mariana Abe Vicente Cavagnari Marina Kimiko Kadowaki Marise Fonseca dos Santos Marla Karine Amarante Maurício Turkiewicz Mauro Antonio Alves Castro Michel Rodrigo Zambrano Passarini Michele Potrich Michelle Orane Schemberger Milena Massumi Kozonoe Mônica Degraf Cavallin Monica Tereza Suldofski Mucio Luiz de Assis Cirino Nadia Graciele Krohn Najeh Maissar Khalil Nédia de Castilhos Ghisi Neide Tomimura Costa Neiva Leite Neyva Maria Lopes Romeiro Patrícia Amâncio da Rosa Patricia Dayane Carvalho Schaker Patricia Oehlmeyer Nassar Patrícia Savio de Araújo-Souza Patricia Silva Lucio Paulo Henrique Couto Souza Paulo Roberto Donadio Percy Nohama Quirino Alves de Lima Neto Rafael Deminice Rafael dos Santos Bezerra Raquel Alves dos Santos Renan Manozzo Galante Renata Ernlund Freitas de Macedo Rita de Cássia Garcia Simão Roberta Losi Guembarovski Roberto H. Herai Roberto Rosati Rodrigo Ferreira Rodrigo Rodrigues Matiello Rogério Neri Shinsato Rogério Pincela Mateus Rosane Aparecida Ribeiro Rosilene Fressatti Cardoso Rosilene Fressatti Cardoso Sandra Mara Guse Scós Venske Selene Elifio Esposito Sérgio Ossamu Ioshii Silvana Giuliatti Silvia Mara de Souza Halick Silvio Henrique Maia de Almeida Simone Neumann Wendt Spencer Luiz Marques Payão Stefan Wolanski Negrão Stephane Janaina de Moura Escobar Sueli Fumie Yamada Ogatta SUELI PERCIO QUINAIA Taciane Finatto Tatiana Mayumi Veiga Iriyoda Tayza Katelline Danilau Ostroski Tony Alexander Hild Valeria Valente Vanessa Nascimento Kozak Vanessa Santos Sotomaior Victor Breno Pedrosa Victoria Zeghbi Cochenski Borba Vivian Rotuno Moure Valdameri Wander Rogerio Pavanelli Weber Cláudio Francisco Nunes da Silva Willian Augusto de Melo Yohandra Reyes Torres |
| EPI_ISL_2758772, EPI_ISL_2758774, EPI_ISL_2758775 | HUEM/IBMP | IPEC Guarapuava | NAPI-Genômica (Novos Arranjo de Pesquisa e Inovação em Genômica): Ademar Dantas da Cunha Júnior Adriano Ferrasa Adriano Mondini Aldo Przybysz Alessandra Lourenço Cecchini Armani Alex Sandro Jorge Alexandra Ivo de Medeiros Alexandre Maller Aline Cristina Batista Rodrigues Johann Ana Lucia Ferreira Ana Marisa Fusco Almeida Anderson Joel Martino Andrade André Luís Laforga Vanzela Andrea Duarte Doetzer Andrea Name Colado Simao Andressa Pereira de Souza Anelisa Ramão Angelica Beate Winter Boldt Anna Hermínia Castro Gomes de Amorim Anna Silvia Penteado Setti da Rocha Antonio Camilo da Silva Filho Antonio Stabelini Neto Arthur Hirata Bertachi Barbara Mendes Paz Chao Betty Cristiane Kuhn Bruno Ambrozio Galindo Bruno Ribeiro Cruz Camilla Reginatto De Pierri Carla Fredrichsen Moya Araujo Carla Fredrichsen Moya Araujo Carlos Alberto Oliveira de Biagi Junior Carlos Augusto Nassar Carlos Eduardo Buss Carlos Gilberto Carlotti Junior Carlos Henrique Schneider Carolina Panis Carolina Weigert Galvão Caroline de Jesus Coelho Donha Caroline Guisantes de Salvo Toni Caryna Eurich Mazur Catiuscie Cabreira da Silva Tortorella Celso F. D. Doliveira |

| | | | |
|---|---|---|---|
| | | | Cesar Luiz Boguszewski Christiane Pienna Soares Chung Man Chin Claudia Moro Cleverson Busso Cristiane Cominetti Daiane Priscila Simão-Silva Dalila Luciola Zanette Daniel de Paula Daniel de Paula Daniel Rech Daniela Fiori Gradia Daniela Pretti da Cunha Tirapelli Daniela Viganó Zanoti Jeronymo Daniele Ukan Danielle Malheiros Ferreira Danielle Venturini Deborah Catharine de Assis Leite Deivid Calebe de Souza Dennis Armando Bertolini Edenir Inez Pamero Edna Maria Vissoci Reiche Edson Roberto Arpini Miguel Eduardo José de Almeida Araújo Eliana Carolina Vespero Eliandro Reis Tavares Elza Kimura Grimshaw Emanuel Maltempi de Souza Emanuele Cristina Gustani Buss Émerson Carraro Emiliana Cristina Melo ENILze Maria de Souza Fonseca Ribeiro Enilze Maria de Souza Fonseca Ribeiro Erika Izumi Erika Seki Kioshima Cotica Evani Marques Pereira Fabio Negretti Fábio Rodrigues Ferreira Seiva Felipe Dunin dos Santos Felipe Tuon Fernanda Andreia Rosa Fernanda Cestaro Prado Cortez Fernanda Ivanski Fernanda Maris Peria Flavia Regina Oliveira de Barros Franciele Aní Caovilla Follador Franciele Mara Lucca Zanardo Bohm Francinete Ramos Campos Fulviana Silva Nishiyama GABRIEL RIBEIRO CORDEIRO Gabriela Datsch Bennemann Gisele Santos de Oliveira Glaucio Valdameri Glauco Akelinghton Freire Vitiello Glauco Vieira Miranda Glaura Scantamburlo ALves Fernandes Guilherme Ferreira Silveira Gustavo Bianchini Porfírio Gustavo Lenci Marques Hélito Volpato Hildebrando Masshiro Nagai Huei Diana Lee Ilce Mara de Syllos Cólus Iris Rabinovich Israel Gomy Jackson Kawakami Jacques Duílio Brancher Jaime Luís Lopes Rocha Jaqueline Carvalho de Oliveira Jean Henrique da Silva Rodrigues Jean Leandro dos Santos Jeane Eliete Laguila Visentainer João Paulo Bianchi Ximenez Joaquim Manoel da Silva Jociani Ascari Joel Donazzolo Jorge Luis Maria Ruiz Jose Knopfholz José Luis da Conceição Silva José Sebastião dos Santos Joseane Carla Schabarum Juliana Cheleski Wiggers Juliana Mara Serpeloni Juliana Morini Küpper Cardoso Perseguini Karen Brajão de Oliveira Karin Braun Prado Karine Aparecida de Lima Katiany Rizzieri Caleffi Ferracioli Katiuscia de Oliveira Francisco Gabriel Kelvinson Fernandes Viana Larissa Beatriz Cossalter Larissa Danielle Bahls Pinto Laurival Antonio Vilas Boas Léia Carolina Lucio Líbero Mezzadri Neto Ligia Carla Faccin Galhardi Lirane Elize Defante Ferreto Luciana Furlaneto Maia Luciana Oliveira de Fariña Luciana Reis Azevedo Alanis Luciane Regina Cavalli Lucy Megumi Yamauchi Lioni Luis Paulo Gomes Mascarenhas Luis Paulo Gomes Mascarenhas Luís Paulo Mascarenhas Lupe Furtado Alle Lyvia Regina Biagi Silva Bertachi Mara Antonia Ramos Costa Mara L. Cordeiro Marcela Maria Birolim Marcelo Ricardo Vicari Marcia Edilaine Lopes Consolaro Marcia Holsbach Beltrame Marcia Regina Eches Perugini Marcos Abdo Arbex Marcos Pileggi MARCOS TADEU GRZELCZAK Marcus Peikriszwili Tartaruga Maria Angelica Ehara Watanabe Maria Antonia Ramos Costa Maria Claudia Gross Maria José Soares Mendes Giannini Maria Leandra Terencio Maria Lúcia Bonfleur Maria Luiza Guimarães de Oliveira Maria Luiza Petzl-Erler Mariana Abe Vicente Cavagnari Marina Kimiko Kadowaki Marise Fonseca dos Santos Marla Karine Amarante Maurício Turkiewicz Mauro Antonio Alves Castro Michel Rodrigo Zambrano Passarini Michele Potrich Michelle Orane Schemberger Milena Massumi Kozonoe Mônica Degraf Cavallin Monica Tereza Suldofski Mucio Luiz de Assis Cirino Nadia Graciele Krohn Najeh Maissar Khalil Nédia de Castilhos Ghisi Neide Tomimura Costa Neiva Leite Neyva Maria Lopes Romeiro Patrícia Amâncio da Rosa Patricia Dayane Carvalho Schaker Patricia Oehlmeyer Nassar Patrícia Savio de Araújo-Souza Patricia Silva Lucio Paulo Henrique Couto Souza Paulo Roberto Donadio Percy Nohama Quirino Alves de Lima Neto Rafael Deminice Rafael dos Santos Bezerra Raquel Alves dos Santos Renan Manozzo Galante Renata Ernlund Freitas de Macedo Rita de Cássia Garcia Simão Roberta Losi Guembarovski Roberto H. Herai Roberto Rosati Rodrigo Ferreira Rodrigo Rodrigues Matiello Rogério Neri Shinsato Rogério Pincela Mateus Rosane Aparecida Ribeiro Rosilene Fressatti Cardoso Rosilene Fressatti Cardoso Sandra Mara Guse Scós Venske Selene Elifio Esposito Sérgio Ossamu Ioshii Silvana Giuliatti Silvia Mara de Souza Halick Silvio Henrique Maia de Almeida Simone Neumann Wendt Spencer Luiz Marques Payão Stefan Wolanski Negrão Stephane Janaina de Moura Escobar Sueli Fumie Yamada Ogatta SUELI PERCIO QUINAIA Taciane Finatto Tatiana Mayumi Veiga Iriyoda Tayza Katelline Danilau Ostroski Tony Alexander Hild Valeria Valente Vanessa Nascimento Kozak Vanessa Santos Sotomaior Victor Breno Pedrosa Victoria Zeghbi Cochenski Borba Vivian Rotuno Moure Valdameri Wander Rogerio Pavanelli Weber Cláudio Francisco Nunes da Silva Willian Augusto de Melo Yohandra Reyes Torres |
| EPI_ISL_2758777, EPI_ISL_2758779, EPI_ISL_2758783, EPI_ISL_2758784, EPI_ISL_2758785, EPI_ISL_2758788, EPI_ISL_2758789, EPI_ISL_2758792, EPI_ISL_2758799 | UEL | IPEC Guarapuava | NAPI-Genômica (Novos Arranjo de Pesquisa e Inovação em Genômica): Ademar Dantas da Cunha Júnior Adriano Ferrasa Adriano Mondini Aldo Przybysz Alessandra Lourenço Cecchini Armani Alex Sandro Jorge Alexandra Ivo de Medeiros Alexandre Maller Aline Cristina Batista Rodrigues Johann Ana Lucia Ferreira Ana Marisa Fusco Almeida Anderson Joel Martino Andrade André Luís Laforga Vanzela Andrea Duarte Doetzer Andrea Name Colado Simao Andressa Pereira de Souza Anelisa Ramão Angelica Beate Winter Boldt Anna Hermínia Castro Gomes de Amorim Anna Silvia Penteado Setti da Rocha Antonio Camilo da Silva Filho Antonio Stabelini Neto Arthur Hirata Bertachi Barbara Mendes Paz Chao Betty Cristiane Kuhn Bruno Ambrozio Galindo Bruno Ribeiro Cruz Camilla Reginatto De Pierri Carla Fredrichsen Moya Araujo Carla Fredrichsen Moya Araujo Carlos Alberto Oliveira de Biagi Junior Carlos Augusto Nassar Carlos Eduardo Buss Carlos Gilberto Carlotti Junior Carlos Henrique Schneider Carolina Panis Carolina Weigert Galvão Caroline de Jesus Coelho Donha Caroline Guisantes de Salvo Toni Caryna Eurich Mazur Catiuscie Cabreira da Silva Tortorella Celso F. D. Doliveira Cesar Luiz Boguszewski Christiane Pienna Soares Chung Man Chin Claudia Moro Cleverson Busso Cristiane Cominetti Daiane Priscila Simão-Silva Dalila Luciola Zanette Daniel de Paula Daniel de Paula Daniel Rech Daniela Fiori Gradia Daniela Pretti da Cunha Tirapelli Daniela Viganó Zanoti Jeronymo Daniele Ukan Danielle Malheiros Ferreira Danielle Venturini Deborah Catharine de Assis Leite Deivid Calebe de Souza Dennis Armando Bertolini Edenir Inez Pamero Edna Maria Vissoci Reiche Edson Roberto Arpini Miguel Eduardo José de Almeida Araújo Eliana Carolina Vespero Eliandro Reis Tavares Elza Kimura Grimshaw Emanuel Maltempi de Souza Emanuele Cristina Gustani Buss Émerson Carraro Emiliana Cristina Melo ENILze Maria de Souza Fonseca Ribeiro Enilze Maria de Souza Fonseca Ribeiro Erika Izumi Erika Seki Kioshima Cotica Evani Marques Pereira Fabio Negretti Fábio Rodrigues Ferreira Seiva Felipe Dunin dos Santos Felipe Tuon Fernanda Andreia Rosa Fernanda Cestaro Prado Cortez Fernanda Ivanski Fernanda Maris Peria Flavia Regina Oliveira de Barros Franciele Aní Caovilla Follador Franciele Mara Lucca Zanardo Bohm Francinete Ramos Campos Fulviana Silva Nishiyama GABRIEL RIBEIRO CORDEIRO Gabriela Datsch Bennemann Gisele Santos de Oliveira Glaucio Valdameri Glauco Akelinghton Freire Vitiello Glauco Vieira Miranda Glaura Scantamburlo ALves Fernandes Guilherme Ferreira Silveira Gustavo Bianchini Porfírio Gustavo Lenci Marques Hélito Volpato Hildebrando Masshiro Nagai Huei Diana Lee Ilce Mara de Syllos Cólus Iris Rabinovich Israel Gomy Jackson Kawakami Jacques Duílio Brancher Jaime Luís Lopes Rocha Jaqueline Carvalho de Oliveira Jean Henrique da Silva Rodrigues Jean Leandro dos Santos Jeane Eliete Laguila Visentainer João Paulo Bianchi Ximenez Joaquim Manoel da Silva Jociani Ascari Joel Donazzolo Jorge Luis Maria Ruiz Jose Knopfholz José Luis da Conceição Silva José Sebastião dos Santos Joseane Carla Schabarum Juliana Cheleski Wiggers Juliana Mara Serpeloni Juliana Morini Küpper Cardoso Perseguini Karen Brajão de Oliveira Karin Braun Prado Karine Aparecida de Lima Katiany Rizzieri Caleffi Ferracioli Katiuscia de Oliveira Francisco Gabriel Kelvinson Fernandes Viana Larissa Beatriz Cossalter Larissa Danielle Bahls Pinto Laurival Antonio Vilas Boas Léia Carolina Lucio Líbero Mezzadri Neto Ligia Carla Faccin Galhardi Lirane Elize Defante Ferreto Luciana Furlaneto Maia Luciana Oliveira de Fariña Luciana Reis Azevedo Alanis Luciane Regina Cavalli Lucy Megumi Yamauchi Lioni Luis Paulo Gomes Mascarenhas Luis Paulo Gomes Mascarenhas Luís Paulo Mascarenhas Lupe Furtado Alle Lyvia Regina Biagi Silva Bertachi Mara Antonia Ramos Costa Mara L. Cordeiro Marcela Maria Birolim Marcelo Ricardo Vicari Marcia Edilaine Lopes Consolaro Marcia Holsbach Beltrame Marcia Regina Eches Perugini Marcos Abdo Arbex Marcos Pileggi MARCOS TADEU GRZELCZAK Marcus Peikriszwili Tartaruga Maria Angelica Ehara Watanabe Maria Antonia Ramos Costa Maria Claudia Gross Maria José Soares Mendes Giannini Maria Leandra Terencio Maria Lúcia Bonfleur Maria Luiza Guimarães de Oliveira Maria Luiza Petzl-Erler Mariana Abe Vicente Cavagnari Marina Kimiko Kadowaki Marise Fonseca dos Santos Marla Karine Amarante Maurício Turkiewicz Mauro Antonio Alves Castro Michel Rodrigo Zambrano Passarini Michele Potrich Michelle Orane Schemberger Milena Massumi Kozonoe Mônica Degraf Cavallin Monica Tereza Suldofski Mucio Luiz de Assis Cirino Nadia Graciele Krohn Najeh Maissar Khalil Nédia de Castilhos Ghisi Neide Tomimura Costa Neiva Leite Neyva Maria Lopes Romeiro Patrícia Amâncio da Rosa Patricia Dayane Carvalho Schaker Patricia Oehlmeyer Nassar Patrícia Savio de Araújo-Souza Patricia Silva Lucio Paulo Henrique Couto Souza Paulo Roberto Donadio Percy Nohama Quirino Alves de Lima Neto Rafael Deminice Rafael dos Santos Bezerra Raquel Alves dos Santos Renan Manozzo Galante Renata Ernlund Freitas de Macedo Rita de Cássia Garcia Simão Roberta Losi Guembarovski Roberto H. Herai Roberto Rosati Rodrigo Ferreira Rodrigo Rodrigues Matiello Rogério Neri Shinsato Rogério Pincela Mateus Rosane Aparecida Ribeiro Rosilene Fressatti Cardoso Rosilene Fressatti Cardoso Sandra Mara Guse Scós Venske Selene Elifio Esposito Sérgio Ossamu Ioshii Silvana Giuliatti Silvia Mara de Souza Halick Silvio Henrique Maia de Almeida Simone Neumann Wendt Spencer Luiz Marques Payão Stefan Wolanski Negrão Stephane Janaina de Moura Escobar Sueli Fumie Yamada Ogatta SUELI PERCIO QUINAIA Taciane Finatto Tatiana Mayumi Veiga Iriyoda Tayza Katelline Danilau Ostroski Tony Alexander Hild Valeria Valente Vanessa Nascimento Kozak Vanessa Santos Sotomaior Victor Breno Pedrosa Victoria Zeghbi Cochenski Borba Vivian Rotuno Moure Valdameri Wander Rogerio Pavanelli Weber Cláudio Francisco Nunes da Silva Willian Augusto de Melo Yohandra Reyes Torres |
| EPI_ISL_2758806 | IPEC Guarapuava | IPEC Guarapuava | NAPI-Genômica (Novos Arranjo de Pesquisa e Inovação em Genômica): Ademar Dantas da Cunha Júnior Adriano Ferrasa Adriano Mondini Aldo Przybysz Alessandra Lourenço Cecchini Armani Alex Sandro Jorge Alexandra Ivo de Medeiros Alexandre Maller Aline Cristina Batista Rodrigues Johann Ana Lucia Ferreira Ana Marisa Fusco Almeida Anderson Joel Martino Andrade André Luís Laforga Vanzela Andrea Duarte Doetzer Andrea Name Colado Simao Andressa Pereira de Souza Anelisa Ramão Angelica Beate Winter Boldt Anna Hermínia Castro Gomes de Amorim Anna Silvia Penteado Setti da Rocha Antonio Camilo da Silva Filho Antonio Stabelini Neto Arthur Hirata Bertachi Barbara Mendes Paz Chao Betty Cristiane Kuhn Bruno Ambrozio Galindo Bruno Ribeiro Cruz Camilla Reginatto De Pierri Carla Fredrichsen Moya Araujo Carla Fredrichsen Moya Araujo Carlos Alberto Oliveira de Biagi Junior Carlos Augusto Nassar Carlos Eduardo Buss Carlos Gilberto Carlotti Junior Carlos Henrique Schneider Carolina Panis Carolina Weigert Galvão Caroline de Jesus Coelho Donha Caroline Guisantes de Salvo Toni Caryna Eurich Mazur Catiuscie Cabreira da Silva Tortorella Celso F. D. Doliveira Cesar Luiz Boguszewski Christiane Pienna Soares Chung Man Chin Claudia Moro Cleverson Busso Cristiane Cominetti Daiane Priscila Simão-Silva Dalila Luciola Zanette Daniel de Paula Daniel de Paula Daniel Rech Daniela Fiori Gradia Daniela Pretti da Cunha Tirapelli Daniela Viganó Zanoti Jeronymo |

| | | | |
|---|---|---|---|
| | | | Daniele Ukan Danielle Malheiros Ferreira Danielle Venturini Deborah Catharine de Assis Leite Deivid Calebe de Souza Dennis Armando Bertolini Edenir Inez Pamero Edna Maria Vissoci Reiche Edson Roberto Arpini Miguel Eduardo José de Almeida Araújo Eliana Carolina Vespero Eliandro Reis Tavares Elza Kimura Grimshaw Emanuel Maltempi de Souza Emanuele Cristina Gustani Buss Émerson Carraro Emiliana Cristina Melo ENILze Maria de Souza Fonseca Ribeiro Enilze Maria de Souza Fonseca Ribeiro Erika Izumi Erika Seki Kioshima Cotica Evani Marques Pereira Fabio Negretti Fábio Rodrigues Ferreira Seiva Felipe Dunin dos Santos Felipe Tuon Fernanda Andreia Rosa Fernanda Cestaro Prado Cortez Fernanda Ivanski Fernanda Maris Peria Flavia Regina Oliveira de Barros Franciele Aní Caovilla Follador Franciele Mara Lucca Zanardo Bohm Francinete Ramos Campos Fulviana Silva Nishiyama GABRIEL RIBEIRO CORDEIRO Gabriela Datsch Bennemann Gisele Santos de Oliveira Glaucio Valdameri Glauco Akelinghton Freire Vitiello Glauco Vieira Miranda Glaura Scantamburlo ALves Fernandes Guilherme Ferreira Silveira Gustavo Bianchini Porfírio Gustavo Lenci Marques Hélito Volpato Hildebrando Masshiro Nagai Huei Diana Lee Ilce Mara de Syllos Cólus Iris Rabinovich Israel Gomy Jackson Kawakami Jacques Duílio Brancher Jaime Luís Lopes Rocha Jaqueline Carvalho de Oliveira Jean Henrique da Silva Rodrigues Jean Leandro dos Santos Jeane Eliete Laguila Visentainer João Paulo Bianchi Ximenez Joaquim Manoel da Silva Jociani Ascari Joel Donazzolo Jorge Luis Maria Ruiz Jose Knopfholz José Luis da Conceição Silva José Sebastião dos Santos Joseane Carla Schabarum Juliana Cheleski Wiggers Juliana Mara Serpeloni Juliana Morini Küpper Cardoso Perseguini Karen Brajão de Oliveira Karin Braun Prado Karine Aparecida de Lima Katiany Rizzieri Caleffi Ferracioli Katiuscia de Oliveira Francisco Gabriel Kelvinson Fernandes Viana Larissa Beatriz Cossalter Larissa Danielle Bahls Pinto Laurival Antonio Vilas Boas Léia Carolina Lucio Líbero Mezzadri Neto Ligia Carla Faccin Galhardi Lirane Elize Defante Ferreto Luciana Furlaneto Maia Luciana Oliveira de Fariña Luciana Reis Azevedo Alanis Luciane Regina Cavalli Lucy Megumi Yamauchi Lioni Luis Paulo Gomes Mascarenhas Luis Paulo Gomes Mascarenhas Luís Paulo Mascarenhas Lupe Furtado Alle Lyvia Regina Biagi Silva Bertachi Mara Antonia Ramos Costa Mara L. Cordeiro Marcela Maria Birolim Marcelo Ricardo Vicari Marcia Edilaine Lopes Consolaro Marcia Holsbach Beltrame Marcia Regina Eches Perugini Marcos Abdo Arbex Marcos Pileggi MARCOS TADEU GRZELCZAK Marcus Peikriszwili Tartaruga Maria Angelica Ehara Watanabe Maria Antonia Ramos Costa Maria Claudia Gross Maria José Soares Mendes Giannini Maria Leandra Terencio Maria Lúcia Bonfleur Maria Luiza Guimarães de Oliveira Maria Luiza Petzl-Erler Mariana Abe Vicente Cavagnari Marina Kimiko Kadowaki Marise Fonseca dos Santos Marla Karine Amarante Maurício Turkiewicz Mauro Antonio Alves Castro Michel Rodrigo Zambrano Passarini Michele Potrich Michelle Orane Schemberger Milena Massumi Kozonoe Mônica Degraf Cavallin Monica Tereza Suldofski Mucio Luiz de Assis Cirino Nadia Graciele Krohn Najeh Maissar Khalil Nédia de Castilhos Ghisi Neide Tomimura Costa Neiva Leite Neyva Maria Lopes Romeiro Patrícia Amâncio da Rosa Patricia Dayane Carvalho Schaker Patricia Oehlmeyer Nassar Patrícia Savio de Araújo-Souza Patricia Silva Lucio Paulo Henrique Couto Souza Paulo Roberto Donadio Percy Nohama Quirino Alves de Lima Neto Rafael Deminice Rafael dos Santos Bezerra Raquel Alves dos Santos Renan Manozzo Galante Renata Ernlund Freitas de Macedo Rita de Cássia Garcia Simão Roberta Losi Guembarovski Roberto H. Herai Roberto Rosati Rodrigo Ferreira Rodrigo Rodrigues Matiello Rogério Neri Shinsato Rogério Pincela Mateus Rosane Aparecida Ribeiro Rosilene Fressatti Cardoso Rosilene Fressatti Cardoso Sandra Mara Guse Scós Venske Selene Elifio Esposito Sérgio Ossamu Ioshii Silvana Giuliatti Silvia Mara de Souza Halick Silvio Henrique Maia de Almeida Simone Neumann Wendt Spencer Luiz Marques Payão Stefan Wolanski Negrão Stephane Janaina de Moura Escobar Sueli Fumie Yamada Ogatta SUELI PERCIO QUINAIA Taciane Finatto Tatiana Mayumi Veiga Iriyoda Tayza Katelline Danilau Ostroski Tony Alexander Hild Valeria Valente Vanessa Nascimento Kozak Vanessa Santos Sotomaior Victor Breno Pedrosa Victoria Zeghbi Cochenski Borba Vivian Rotuno Moure Valdameri Wander Rogerio Pavanelli Weber Cláudio Francisco Nunes da Silva Willian Augusto de Melo Yohandra Reyes Torres |
| EPI_ISL_2775416, EPI_ISL_2775424, EPI_ISL_2775425, EPI_ISL_2775437, EPI_ISL_2775444, EPI_ISL_2775452, EPI_ISL_2775454, EPI_ISL_2775455, EPI_ISL_2775456, EPI_ISL_2775458, EPI_ISL_2775463, EPI_ISL_2775466, EPI_ISL_2775469, EPI_ISL_2775470, EPI_ISL_2775472, EPI_ISL_2775473, EPI_ISL_2775474, EPI_ISL_2775476, EPI_ISL_2775477, EPI_ISL_2775478, EPI_ISL_2775479, EPI_ISL_2775480, EPI_ISL_2775481, EPI_ISL_2775485, EPI_ISL_2775487, EPI_ISL_2775492 | | | |
| see above | Laboratorio Central de Saude Publica do Estado do Parana (Instituto de Biologia Molecular do Paraná (LAC)EN-PR) | Instituto Carlos Chagas - Fiocruz | Mauro de Medeiros Oliveira, Michelle Orane Schemberger, Andreia Akemi Suzukawa, Irina Nastassja Riediger, Maria do Carmo Debur, Guilherme Becker, Paola Cristina Resende, Tiago Gräf, Eduardo Balsanelli, Valter Antônio de Baura, Emanuel Maltempi de Souza, Fábio de Oliveira Pedrosa, Lysangela Ronalte Alves, Lucas Blanes, Sheila Cristina Nardeli, Alessandra De Melo Aguiar, Letusa Albrecht, Dalila Zanette, Andréa Rodrigues Ávila, Luis Gustavo Morello, Fabricio Klerynton Marchini, Hellen Geremias dos Santos, Fabio Passetti, Bruno Dallagiovanna, Helisson Faoro |
| EPI_ISL_2777360, EPI_ISL_2777371, EPI_ISL_2777387, EPI_ISL_2777390, EPI_ISL_2777391, EPI_ISL_2777392, EPI_ISL_2777394 | Laboratorio de Ecologia de Doencas Transmissiveis na Amazonia, Instituto Leonidas e Maria Deane - Fiocruz Amazonia | Laboratorio de Ecologia de Doencas Transmissiveis na Amazonia, Instituto Leonidas e Maria Deane - Fiocruz Amazonia | Valdinete Nascimento, Victor Souza, André Corado, Fernanda Nascimento, George Silva, Ágatha Costa, Debora Duarte, Karina Pessoa, Matilde Mejía, Luciana Gonçalves, Maria Júlia Brandão, Michele Jesus, Felipe Naveca |
| EPI_ISL_2821285, EPI_ISL_2821287, EPI_ISL_2821291, EPI_ISL_2821293, EPI_ISL_2821298, EPI_ISL_2821305, EPI_ISL_2821311, EPI_ISL_2821316, EPI_ISL_2821317, EPI_ISL_2821318, EPI_ISL_2821320, EPI_ISL_2821321, EPI_ISL_2821323, EPI_ISL_2821324, EPI_ISL_2821325 | | | |
| see above | LACEN/PE | WallauLab on behalf of Fiocruz COVID-19 Genomic Surveillance Network | Marcelo Henrique dos Santos Paiva, Duschinka Ribeiro Duarte Guedes, Cássia Docena, Matheus Filgueira Bezerra, Filipe Zimmer Dezordi, Laís Ceschini Machado, Larissa Krokovsky, Elisama Helvecio, Alexandre Freitas da Silva, Antonio Mauro Rezende, Sinval Pinto Brandão Filho, Constância Flávia Junqueira Ayres, Gabriel Luz Wallau |
| EPI_ISL_416036 | National Influenza Center - Instituto Adolfo Lutz | Instituto Adolfo Lutz, Interdiciplinary Procedures Center, Strategic Laboratory | Claudio Tavares Sacchi, Claudia Regina Gonçalves, Carlos Henrique Camargo, Erica Valessa Ramos Gomes, Fabiana Cristina Pereira dos Santos, Daniela Bernardes Borges da Silva, Simone Guadagnucci Morillo, Adriano Abbud, Adriana Bugno, Maria do Carmo Sampaio Tavares Timenetsky, Terezinha Maria de Paiva |
| EPI_ISL_427292 | Laboratório Central de Saúde Pública do Estado de Alagoas (LACEN-AL) | Laboratory of Respiratory Viruses and Measles, Oswaldo Cruz Institute, FIOCRUZ | Paola Resende, Fernando Motta, Luciana Appolinario, Sunando Roy, Aline Mattos, Milene Miranda, Cristiana Garcia, Braulia Caetano, Maria Ogrzewalska, Priscila Born, Jonathan Lopes, Marilda Siqueira on behalf of the Fiocruz COVID-19 Genomic Surveillance Network |
| EPI_ISL_456088 | Laboratório Central de Saúde Pública Noel Nutels (LACEN-RJ) | Laboratory of Respiratory Viruses and Measles, Oswaldo Cruz Institute, FIOCRUZ | Paola Resende, Luciana Appolinario, Fernando Motta, Aline Mattos, Milene Miranda, Cristiana Garcia, Braulia Caetano, Maria Ogrzewalska, Jonathan Lopes, Marilda Siqueira on behalf of the Fiocruz COVID-19 Genomic Surveillance Network |
| EPI_ISL_458140, EPI_ISL_458141, EPI_ISL_458146, EPI_ISL_458147 | Evandro Chagas Institute | Evandro Chagas Institute | Santos, M.C.; Silva, A.M.; Junior, W.D.C.; Barbagelata, L.S.; Ferreira, J.A.; Sousa, E.M.A.; da Silva, P.S.; Resque, H.R; Martins, L.C.; Sousa Junior, E.C.;Viana, G.M.R |
| EPI_ISL_467356, EPI_ISL_467359, EPI_ISL_467366 | Laboratory of Respiratory Viruses and Measles, Oswaldo Cruz Institute, FIOCRUZ | Laboratory of Respiratory Viruses and Measles, Oswaldo Cruz Institute, FIOCRUZ | Paola Resende, Luciana Appolinario, Fernando Motta, Anna Carolina Paixão, Ana Carolina Mendonça, Aline Mattos, Milene Miranda, Cristiana Garcia, Braulia Caetano, Maria Ogrzewalska, Jonathan Lopes, Marilda Siqueira on behalf of the Fiocruz COVID-19 Genomic Surveillance Network |
| EPI_ISL_468305, EPI_ISL_468307 | Centro de Vigilancia a Saude de Diadema | Instituto Adolfo Lutz, Interdiciplinary Procedures Center, Strategic Laboratory | Claudio Tavares Sacchi, Claudia Regina Gonçalves, Erica Valessa Ramos Gomes |
| EPI_ISL_468308 | Hospital Municipal do Tatuape Carmino Caricchio | Instituto Adolfo Lutz, Interdiciplinary Procedures Center, Strategic Laboratory | Claudio Tavares Sacchi, Claudia Regina Gonçalves, Erica Valessa Ramos Gomes |
| EPI_ISL_468311, EPI_ISL_468312 | Hospital Municipal Dr Ignacio Proenca de Gouvea | Instituto Adolfo Lutz, Interdiciplinary Procedures Center, Strategic Laboratory | Claudio Tavares Sacchi, Claudia Regina Gonçalves, Erica Valessa Ramos Gomes |
| EPI_ISL_468313 | Vigilancia Epidemiologica de São Bernardo do Campo | Instituto Adolfo Lutz, Interdiciplinary Procedures Center, Strategic Laboratory | Claudio Tavares Sacchi, Claudia Regina Gonçalves, Erica Valessa Ramos Gomes |
| EPI_ISL_468314 | CTA Centro de Testagem e Aconselhamento | Instituto Adolfo Lutz, Interdiciplinary Procedures Center, Strategic Laboratory | Claudio Tavares Sacchi, Claudia Regina Gonçalves, Erica Valessa Ramos Gomes |
| EPI_ISL_468315 | Hospital Municipal do Tatuape Carmino Caricchio | Instituto Adolfo Lutz, Interdiciplinary Procedures Center, Strategic Laboratory | Claudio Tavares Sacchi, Claudia Regina Gonçalves, Erica Valessa Ramos Gomes |
| EPI_ISL_468316 | UPA Vila Assis | Instituto Adolfo Lutz, Interdiciplinary Procedures Center, Strategic Laboratory | Claudio Tavares Sacchi, Claudia Regina Gonçalves, Erica Valessa Ramos Gomes |
| EPI_ISL_468318 | Hospital Universitario da USP | Instituto Adolfo Lutz, Interdiciplinary Procedures Center, Strategic Laboratory | Claudio Tavares Sacchi, Claudia Regina Gonçalves, Erica Valessa Ramos Gomes |
| EPI_ISL_468319 | Vigilancia Epidemiologica de São Bernardo do Campo | Instituto Adolfo Lutz, Interdiciplinary Procedures Center, Strategic Laboratory | Claudio Tavares Sacchi, Claudia Regina Gonçalves, Erica Valessa Ramos Gomes |
| EPI_ISL_468321 | Hospital Universitario da USP | Instituto Adolfo Lutz, Interdiciplinary Procedures Center, Strategic Laboratory | Claudio Tavares Sacchi, Claudia Regina Gonçalves, Erica Valessa Ramos Gomes |

| | | | |
|---|---|---|---|
| EPI_ISL_471539 | Hospital Universitario da USP Sao Paulo | Instituto Adolfo Lutz, Interdiciplinary Procedures Center, Strategic Laboratory | Claudio Tavares Sacchi, Claudia Regina Gonçalves, Erica Valessa Ramos Gomes |
| EPI_ISL_471541 | Hospital Geral Santa Marcelina | Instituto Adolfo Lutz, Interdiciplinary Procedures Center, Strategic Laboratory | Claudio Tavares Sacchi, Claudia Regina Gonçalves, Erica Valessa Ramos Gomes |
| EPI_ISL_471542 | Secretaria de Saude de Mogi das Cruzes | Instituto Adolfo Lutz, Interdiciplinary Procedures Center, Strategic Laboratory | Claudio Tavares Sacchi, Claudia Regina Gonçalves, Erica Valessa Ramos Gomes |
| EPI_ISL_471545 | Hospital Sao Paulo de Ensino da Unifesp | Instituto Adolfo Lutz, Interdiciplinary Procedures Center, Strategic Laboratory | Claudio Tavares Sacchi, Claudia Regina Gonçalves, Erica Valessa Ramos Gomes |
| EPI_ISL_471546 | AMA DR Jose Soares Hungria | Instituto Adolfo Lutz, Interdiciplinary Procedures Center, Strategic Laboratory | Claudio Tavares Sacchi, Claudia Regina Gonçalves, Erica Valessa Ramos Gomes |
| EPI_ISL_471548 | Hospital do Servidor Público Estadual Francisco Morato de Oliveira | Instituto Adolfo Lutz, Interdiciplinary Procedures Center, Strategic Laboratory | Claudio Tavares Sacchi, Claudia Regina Gonçalves, Erica Valessa Ramos Gomes |
| EPI_ISL_471549 | Hospital Municipal Carmen Prudente | Instituto Adolfo Lutz, Interdiciplinary Procedures Center, Strategic Laboratory | Claudio Tavares Sacchi, Claudia Regina Gonçalves, Erica Valessa Ramos Gomes |
| EPI_ISL_471552 | Hospital Sancta Maggiore | Instituto Adolfo Lutz, Interdiciplinary Procedures Center, Strategic Laboratory | Claudio Tavares Sacchi, Claudia Regina Gonçalves, Erica Valessa Ramos Gomes |
| EPI_ISL_471556 | Pronto Socorro Jose Ibrahin | Instituto Adolfo Lutz, Interdiciplinary Procedures Center, Strategic Laboratory | Claudio Tavares Sacchi, Claudia Regina Gonçalves, Erica Valessa Ramos Gomes |
| EPI_ISL_471562, EPI_ISL_471581 | Hosp. Municipal Prof. Dr. Alípio Corrêa Netto | Instituto Adolfo Lutz, Interdiciplinary Procedures Center, Strategic Laboratory | Claudio Tavares Sacchi, Claudia Regina Gonçalves, Erica Valessa Ramos Gomes |
| EPI_ISL_471647 | Hospital Municipal de Barueri Dr. Francisco Moran | Instituto Adolfo Lutz, Interdiciplinary Procedures Center, Strategic Laboratory | Claudio Tavares Sacchi, Claudia Regina Gonçalves, Erica Valessa Ramos Gomes |
| EPI_ISL_471648 | UBS e Pronto Socorro Jd. Jacira | Instituto Adolfo Lutz, Interdiciplinary Procedures Center, Strategic Laboratory | Claudio Tavares Sacchi, Claudia Regina Gonçalves, Erica Valessa Ramos Gomes |
| EPI_ISL_476282 | DB Diagnósticos do Brasil | Instituto de Medicina Tropical da Univesidade de São Paulo | Samples: Nelson Gaburo Jr; Sequencing: Ingra Morales Claro, Jaqueline Goes de Jesus, Erika Regina Manuli, Flavia Cristina da Silva Sales, Thais de Moura Coletti, Camila Alves Maia da Silva, Mariana Severo Ramundo, Giula Magalhaes Ferreira, Darlan da Silva Candido, Julien Theze, Nuno Faria, Ester Sabino |
| EPI_ISL_476341 | Laboratório de Patologia Clínica - UNICAMP | Laboratório de Estudos de Vírus Emergentes - UNICAMP | José Luiz Proença-Modena, Magnun Nueldo Nunes dos Santos, Angelica Schreiber, Julia Forato,Camila Simeoni, Marcilio Jorge Fumagalli, Mariene Ribeiro Amorim, Darlan da Silva Candido, Nuno Rodrigues Faria, Julien Theze, Luiz Gonzaga,Jaqueline Goes Jesus e William Marciel de Souza |
| EPI_ISL_476373 | Hospital da Clínicas da Faculdade de Medicina da Universidade de São Paulo | Instituto de Medicina Tropical da Univesidade de São Paulo | Samples: Ingra Morales Claro, Erika Regina Manuli, Cecilia Salete Alencar, Carolina S. Lazar, Silvia F. Costa; Sequencing: Ingra Morales Claro, Jaqueline Goes de Jesus, Erika Regina Manuli, Flavia Cristina da Silva Sales, Thais de Moura Coletti, Camila Alves Maia da Silva, Mariana Severo Ramundo, Giula Magalhaes Ferreira, Darlan da Silva Candido, Julien Theze, Nuno Faria, Ester Sabino |
| EPI_ISL_476395, EPI_ISL_476398 | Laboratório de Patologia Clínica - UNICAMP | Laboratório de Estudos de Vírus Emergentes - UNICAMP | José Luiz Proença-Modena, Magnun Nueldo Nunes dos Santos, Angelica Schreiber, Julia Forato,Camila Simeoni, Marcilio Jorge Fumagalli, Mariene Ribeiro Amorim, Darlan da Silva Candido, Nuno Rodrigues Faria, Julien Theze, Luiz Gonzaga,Jaqueline Goes Jesus e William Marciel de Souza |
| EPI_ISL_476445, EPI_ISL_476446, EPI_ISL_476469 | Hospital da Clínicas da Faculdade de Medicina da Universidade de São Paulo | Instituto de Medicina Tropical da Univesidade de São Paulo | Samples: Ingra Morales Claro, Erika Regina Manuli, Cecilia Salete Alencar, Carolina S. Lazar, Silvia F. Costa; Sequencing: Ingra Morales Claro, Jaqueline Goes de Jesus, Erika Regina Manuli, Flavia Cristina da Silva Sales, Thais de Moura Coletti, Camila Alves Maia da Silva, Mariana Severo Ramundo, Giula Magalhaes Ferreira, Darlan da Silva Candido, Julien Theze, Nuno Faria, Ester Sabino |
| EPI_ISL_486429 | unknown | Clinical Laboratory, Hospital Israelita Albert Einstein | Malta,F., Amgarten,D., Guedes,R.L., Santana,R.A., de Menezes,F.G., Mangueira,C.L. and Pinho,J.R. |
| EPI_ISL_492036 | Instituto de Biologia do Exército | Laboratório Metabolismo Macromolecular FirminoTorres de Castro, Instituto de Biofísica Carlos Chagas Filho, Universidade Federal do Rio de Janeiro | Bianca Catarina Azevedo Cabral, Aline Rosa Vianna de Souza , Marcos Dornelas-Ribeiro, Tatiana LS Nogueira, Nádia Vaez Gonçalves da Cruz, Caleb GM Santos, Elizabeth Valentin, Marcio da Costa Cipitelli, Virginia Sara Grancieri do Amaral, Rodrigo Soares de Moura Neto, Clarissa Damaso, Rosane Silva |
| EPI_ISL_500483 | Laboratório Central de Saúde Pública do Estado de Pernambuco (LACEN-PE) | WallauLab, Aggeu Magalhaes Institute | Marcelo Henrique Santos Paiva, Duschinka Ribeiro Duarte Guedes, Cássia Docena, Matheus Filgueira Bezerra, Filipe Zimmer Dezordi, Laís Ceschini Machado, Larissa Krokovsky, Elisama Helvecio, Alexandre Freitas da Silva, Luydson Richardson Silva Vasconcelos, Antonio Mauro Rezende, Severino Jefferson Ribeiro da Silva, Kamila Gaudêncio da Silva Sales, Bruna Santos Lima Figueiredo de Sá, Derciliano Lopes da Cruz, Claudio Eduardo Cavalcanti, Armando de Menezes Neto, Caroline Targino Alves da Silva, Renata Pessôa Germano Mendes, Maria Almerice Lopes da Silva, Tiago Gräf, Paola Cristina Resende, Gonzalo Bello, Michelle da Silva Barros, Wheverton Ricardo Correia do Nascimento, Rodrigo Moraes Loyo Arcoverde, Luciane Caroline Albuquerque Bezerra, Sinval Pinto Brandão Filho, Constância Flávia Junqueira Ayres, Gabriel Luz Wallau on behalf of the Fiocruz COVID-19 Genomic Surveillance Network |
| EPI_ISL_502875 | LACEN/PE | LABBE, Federal University of Pernambuco | WILSON JOSE DA SILVA JUNIOR, HEIDI LACERDA ALVES DA CRUZ, MARCOS DA SILVEIRA REGUEIRA NETO, BRUNO SAMPAIO, SERGIO DE SA LEITAO PAIVA JUNIOR, ZILDENE DE SOUSA SILVEIRA, MAIRA GALDINO DA ROCHA PITTA, MICHELLY CRISTINY PEREIRA, REGINALDO GONCALVES DE LIMA NETO, MARCOS ANTONIO DE MORAIS JUNIOR, ANTONIO CARLOS DE FREITAS, VALDIR DE QUEIROZ BALBINO. |
| EPI_ISL_513514, EPI_ISL_513532, EPI_ISL_513546, EPI_ISL_513557, EPI_ISL_513578 | Programa de Oncovirologia, Instituto Nacional de Câncer | Programa de Oncovirologia, Instituto Nacional de Câncer | Juliana D. Siqueira, Livia R. Goes, Brunna M. Alves, Claudia Cicala,James Arthos, João P.B. Viola, Andreia C. de Melo, Marcelo A. Soares |
| EPI_ISL_515520 | Hospital Municipal do Tatuape Carmino Caricchio | Instituto Adolfo Lutz, Interdiciplinary Procedures Center, Strategic Laboratory | Claudio Tavares Sacchi, Claudia Regina Gonçalves, Erica Valessa Ramos Gomes |
| EPI_ISL_515524 | PS Municipal Dr Lauro Ribas Braga | Instituto Adolfo Lutz, Interdiciplinary Procedures Center, Strategic Laboratory | Claudio Tavares Sacchi, Claudia Regina Gonçalves, Erica Valessa Ramos Gomes |
| EPI_ISL_515529 | Pronto Socorro Municipal Julio Tupy | Instituto Adolfo Lutz, Interdiciplinary Procedures Center, Strategic Laboratory | Claudio Tavares Sacchi, Claudia Regina Gonçalves, Erica Valessa Ramos Gomes |
| EPI_ISL_515541 | Hospital Montemagno | Instituto Adolfo Lutz, Interdiciplinary Procedures Center, Strategic Laboratory | Claudio Tavares Sacchi, Claudia Regina Gonçalves, Erica Valessa Ramos Gomes |
| EPI_ISL_515542 | Vigilância Epidemiológica de Leme | Instituto Adolfo Lutz, Interdiciplinary Procedures Center, Strategic Laboratory | Claudio Tavares Sacchi, Claudia Regina Gonçalves, Erica Valessa Ramos Gomes |
| EPI_ISL_515544 | Ama Dr Jose Soares Hungria | Instituto Adolfo Lutz, Interdiciplinary Procedures Center, Strategic Laboratory | Claudio Tavares Sacchi, Claudia Regina Gonçalves, Erica Valessa Ramos Gomes |
| EPI_ISL_515545 | Hospital Sao Paulo de Ensino da Unifesp | Instituto Adolfo Lutz, Interdiciplinary Procedures Center, Strategic Laboratory | Claudio Tavares Sacchi, Claudia Regina Gonçalves, Erica Valessa Ramos Gomes |
| EPI_ISL_515546 | Hospital Municipal do Tatuape Carmino Caricchio | Instituto Adolfo Lutz, Interdiciplinary Procedures Center, Strategic Laboratory | Claudio Tavares Sacchi, Claudia Regina Gonçalves, Erica Valessa Ramos Gomes |
| EPI_ISL_515547 | Centro Medico da Policia Militar do Estado de Sao Paulo | Instituto Adolfo Lutz, Interdiciplinary Procedures Center, Strategic Laboratory | Claudio Tavares Sacchi, Claudia Regina Gonçalves, Erica Valessa Ramos Gomes |

| | | | |
|---|---|---|---|
| EPI_ISL_515548 | Hospital Municipal Dr. Jose Soares Hungria | Instituto Adolfo Lutz, Interdiciplinary Procedures Center, Strategic Laboratory | Claudio Tavares Sacchi, Claudia Regina Gonçalves, Erica Valessa Ramos Gomes |
| EPI_ISL_515552 | Hospital Municipal do Tatuape Carmino Caricchio | Instituto Adolfo Lutz, Interdiciplinary Procedures Center, Strategic Laboratory | Claudio Tavares Sacchi, Claudia Regina Gonçalves, Erica Valessa Ramos Gomes |
| EPI_ISL_515553 | Hospital Municipal Dr. Ignacio Proença de Gouvea | Instituto Adolfo Lutz, Interdiciplinary Procedures Center, Strategic Laboratory | Claudio Tavares Sacchi, Claudia Regina Gonçalves, Erica Valessa Ramos Gomes |
| EPI_ISL_515554 | Pronto Socorro Municipal de Perus | Instituto Adolfo Lutz, Interdiciplinary Procedures Center, Strategic Laboratory | Claudio Tavares Sacchi, Claudia Regina Gonçalves, Erica Valessa Ramos Gomes |
| EPI_ISL_515555 | Hospital Geral de Vila Nova Cachoeirinha | Instituto Adolfo Lutz, Interdiciplinary Procedures Center, Strategic Laboratory | Claudio Tavares Sacchi, Claudia Regina Gonçalves, Erica Valessa Ramos Gomes |
| EPI_ISL_515559, EPI_ISL_515560 | Hospital Sao Paulo de Ensino da Unifesp | Instituto Adolfo Lutz, Interdiciplinary Procedures Center, Strategic Laboratory | Claudio Tavares Sacchi, Claudia Regina Gonçalves, Erica Valessa Ramos Gomes |
| EPI_ISL_515561 | Hospital Montemagno | Instituto Adolfo Lutz, Interdiciplinary Procedures Center, Strategic Laboratory | Claudio Tavares Sacchi, Claudia Regina Gonçalves, Erica Valessa Ramos Gomes |
| EPI_ISL_515562 | Hospital Municipal Doutor Alexandre Zaio | Instituto Adolfo Lutz, Interdiciplinary Procedures Center, Strategic Laboratory | Claudio Tavares Sacchi, Claudia Regina Gonçalves, Erica Valessa Ramos Gomes |
| EPI_ISL_515563 | Hospital Municipal Dr. Jose Soares Hungria | Instituto Adolfo Lutz, Interdiciplinary Procedures Center, Strategic Laboratory | Claudio Tavares Sacchi, Claudia Regina Gonçalves, Erica Valessa Ramos Gomes |
| EPI_ISL_515564 | Hosp. Municipal Prof. Dr. Alípio Corrêa Netto | Instituto Adolfo Lutz, Interdiciplinary Procedures Center, Strategic Laboratory | Claudio Tavares Sacchi, Claudia Regina Gonçalves, Erica Valessa Ramos Gomes |
| EPI_ISL_515565 | Hospital do Servidor Público Estadual Francisco Morato de Oliveira | Instituto Adolfo Lutz, Interdiciplinary Procedures Center, Strategic Laboratory | Claudio Tavares Sacchi, Claudia Regina Gonçalves, Erica Valessa Ramos Gomes |
| EPI_ISL_515566 | PS Municipal Dr Lauro Ribas Braga | Instituto Adolfo Lutz, Interdiciplinary Procedures Center, Strategic Laboratory | Claudio Tavares Sacchi, Claudia Regina Gonçalves, Erica Valessa Ramos Gomes |
| EPI_ISL_523955 | Hospital Municipal do Tatuape Carmino Caricchio | Instituto Adolfo Lutz, Interdiciplinary Procedures Center, Strategic Laboratory | Claudio Tavares Sacchi, Claudia Regina Gonçalves, Erica Valessa Ramos Gomes |
| EPI_ISL_523957 | Hospital Itamaraty | Instituto Adolfo Lutz, Interdiciplinary Procedures Center, Strategic Laboratory | Claudio Tavares Sacchi, Claudia Regina Gonçalves, Erica Valessa Ramos Gomes |
| EPI_ISL_523958 | Pronto Socorro Municipal de Perus | Instituto Adolfo Lutz, Interdiciplinary Procedures Center, Strategic Laboratory | Claudio Tavares Sacchi, Claudia Regina Gonçalves, Erica Valessa Ramos Gomes |
| EPI_ISL_523965 | Hospital do Servidor Público Estadual Francisco Morato de Oliveira | Instituto Adolfo Lutz, Interdiciplinary Procedures Center, Strategic Laboratory | Claudio Tavares Sacchi, Claudia Regina Gonçalves, Erica Valessa Ramos Gomes |
| EPI_ISL_523969 | Hospital Sao Paulo de Ensino da Unifesp | Instituto Adolfo Lutz, Interdiciplinary Procedures Center, Strategic Laboratory | Claudio Tavares Sacchi, Claudia Regina Gonçalves, Erica Valessa Ramos Gomes |
| EPI_ISL_523970 | Conjunto Hospitalar do Mandaqui | Instituto Adolfo Lutz, Interdiciplinary Procedures Center, Strategic Laboratory | Claudio Tavares Sacchi, Claudia Regina Gonçalves, Erica Valessa Ramos Gomes |
| EPI_ISL_523971 | Hospital Geral Santa Marcelina | Instituto Adolfo Lutz, Interdiciplinary Procedures Center, Strategic Laboratory | Claudio Tavares Sacchi, Claudia Regina Gonçalves, Erica Valessa Ramos Gomes |
| EPI_ISL_523974 | Hospital Municipal do Tatuape Carmino Caricchio | Instituto Adolfo Lutz, Interdiciplinary Procedures Center, Strategic Laboratory | Claudio Tavares Sacchi, Claudia Regina Gonçalves, Erica Valessa Ramos Gomes |
| EPI_ISL_523975 | UPA Tito Lopes | Instituto Adolfo Lutz, Interdiciplinary Procedures Center, Strategic Laboratory | Claudio Tavares Sacchi, Claudia Regina Gonçalves, Erica Valessa Ramos Gomes |
| EPI_ISL_523977 | Hosp. Municipal Prof. Dr. Alípio Corrêa Netto | Instituto Adolfo Lutz, Interdiciplinary Procedures Center, Strategic Laboratory | Claudio Tavares Sacchi, Claudia Regina Gonçalves, Erica Valessa Ramos Gomes |
| EPI_ISL_523978 | Hospital do Servidor Público Estadual Francisco Morato de Oliveira | Instituto Adolfo Lutz, Interdiciplinary Procedures Center, Strategic Laboratory | Claudio Tavares Sacchi, Claudia Regina Gonçalves, Erica Valessa Ramos Gomes |
| EPI_ISL_523980 | UPA Tito Lopes | Instituto Adolfo Lutz, Interdiciplinary Procedures Center, Strategic Laboratory | Claudio Tavares Sacchi, Claudia Regina Gonçalves, Erica Valessa Ramos Gomes |
| EPI_ISL_523981 | Hospital Sao Paulo de Ensino da Unifesp | Instituto Adolfo Lutz, Interdiciplinary Procedures Center, Strategic Laboratory | Claudio Tavares Sacchi, Claudia Regina Gonçalves, Erica Valessa Ramos Gomes |
| EPI_ISL_523982 | Hospital do Servidor Público Estadual Francisco Morato de Oliveira | Instituto Adolfo Lutz, Interdiciplinary Procedures Center, Strategic Laboratory | Claudio Tavares Sacchi, Claudia Regina Gonçalves, Erica Valessa Ramos Gomes |
| EPI_ISL_523983 | UPA Campo Limpo | Instituto Adolfo Lutz, Interdiciplinary Procedures Center, Strategic Laboratory | Claudio Tavares Sacchi, Claudia Regina Gonçalves, Erica Valessa Ramos Gomes |
| EPI_ISL_523984 | Ama Dr Jose Soares Hungria | Instituto Adolfo Lutz, Interdiciplinary Procedures Center, Strategic Laboratory | Claudio Tavares Sacchi, Claudia Regina Gonçalves, Erica Valessa Ramos Gomes |
| EPI_ISL_523985 | Hospital Municipal Dr. Benedicto Montenegro | Instituto Adolfo Lutz, Interdiciplinary Procedures Center, Strategic Laboratory | Claudio Tavares Sacchi, Claudia Regina Gonçalves, Erica Valessa Ramos Gomes |
| EPI_ISL_523986 | Ama Dr Jose Soares Hungria | Instituto Adolfo Lutz, Interdiciplinary Procedures Center, Strategic Laboratory | Claudio Tavares Sacchi, Claudia Regina Gonçalves, Erica Valessa Ramos Gomes |
| EPI_ISL_523988 | Hospital Sao Paulo de Ensino da Unifesp | Instituto Adolfo Lutz, Interdiciplinary Procedures Center, Strategic Laboratory | Claudio Tavares Sacchi, Claudia Regina Gonçalves, Erica Valessa Ramos Gomes |
| EPI_ISL_523989 | AMA Jardim Joamar | Instituto Adolfo Lutz, Interdiciplinary Procedures Center, Strategic Laboratory | Claudio Tavares Sacchi, Claudia Regina Gonçalves, Erica Valessa Ramos Gomes |
| EPI_ISL_523990 | AMA Jardim Peri | Instituto Adolfo Lutz, Interdiciplinary Procedures Center, Strategic Laboratory | Claudio Tavares Sacchi, Claudia Regina Gonçalves, Erica Valessa Ramos Gomes |
| EPI_ISL_524462 | Hospital Metropolitano | Instituto Adolfo Lutz, Interdiciplinary Procedures Center, Strategic Laboratory | Claudio Tavares Sacchi, Claudia Regina Gonçalves, Erica Valessa Ramos Gomes |
| EPI_ISL_524463 | Hospital Regional de Cotia | Instituto Adolfo Lutz, Interdiciplinary Procedures Center, Strategic Laboratory | Claudio Tavares Sacchi, Claudia Regina Gonçalves, Erica Valessa Ramos Gomes |

| | | | |
|---|---|---|---|
| EPI_ISL_524464 | Santa Casa de Santa Isabel | Instituto Adolfo Lutz, Interdiciplinary Procedures Center, Strategic Laboratory | Claudio Tavares Sacchi, Claudia Regina Gonçalves, Erica Valessa Ramos Gomes |
| EPI_ISL_524465 | PS Municipal Dr. Caetano Virgílio Neto | Instituto Adolfo Lutz, Interdiciplinary Procedures Center, Strategic Laboratory | Claudio Tavares Sacchi, Claudia Regina Gonçalves, Erica Valessa Ramos Gomes |
| EPI_ISL_524466 | PS Municipal Dr Lauro Ribas Braga | Instituto Adolfo Lutz, Interdiciplinary Procedures Center, Strategic Laboratory | Claudio Tavares Sacchi, Claudia Regina Gonçalves, Erica Valessa Ramos Gomes |
| EPI_ISL_524468 | Hospital Municipal Vereador Jose Storopolli | Instituto Adolfo Lutz, Interdiciplinary Procedures Center, Strategic Laboratory | Claudio Tavares Sacchi, Claudia Regina Gonçalves, Erica Valessa Ramos Gomes |
| EPI_ISL_524469 | Santa Casa de Misericordia de Sao Paulo | Instituto Adolfo Lutz, Interdiciplinary Procedures Center, Strategic Laboratory | Claudio Tavares Sacchi, Claudia Regina Gonçalves, Erica Valessa Ramos Gomes |
| EPI_ISL_524783, EPI_ISL_524785, EPI_ISL_524786, EPI_ISL_524787 | Evandro Chagas Institute | Evandro Chagas Institute | Santos, M.C.; Silva, A.M.; Junior, W.D.C.; Barbagelata, L.S.; Ferreira, J.A.; Sousa, E.M.A.; da Silva, P.S.; Resque, H.R; Martins, L.C.; Sousa Junior, E.C.;Viana, G.M.R |
| EPI_ISL_527856 | Hospital Municipal Prof. Waldomiro de Paula | Instituto Adolfo Lutz, Interdiciplinary Procedures Center, Strategic Laboratory | Claudio Tavares Sacchi, Claudia Regina Gonçalves, Erica Valessa Ramos Gomes |
| EPI_ISL_527857 | Hospital Regional Vale do Ribeira | Instituto Adolfo Lutz, Interdiciplinary Procedures Center, Strategic Laboratory | Claudio Tavares Sacchi, Claudia Regina Gonçalves, Erica Valessa Ramos Gomes |
| EPI_ISL_527859 | Hospital Municipal Vereador Jose Storopolli | Instituto Adolfo Lutz, Interdiciplinary Procedures Center, Strategic Laboratory | Claudio Tavares Sacchi, Claudia Regina Gonçalves, Erica Valessa Ramos Gomes |
| EPI_ISL_527860 | Hospital Municipal de Parelheiros Josanias Castanha Braga | Instituto Adolfo Lutz, Interdiciplinary Procedures Center, Strategic Laboratory | Claudio Tavares Sacchi, Claudia Regina Gonçalves, Erica Valessa Ramos Gomes |
| EPI_ISL_527861 | Hospital e Maternidade Celso Pierro | Instituto Adolfo Lutz, Interdiciplinary Procedures Center, Strategic Laboratory | Av. Dr. Arnaldo, 355 - Brazil, Cerqueira Cesar, São Paulo - SP, 01246-1301 |
| EPI_ISL_527862 | Hospital Municipal de Urgência | Instituto Adolfo Lutz, Interdiciplinary Procedures Center, Strategic Laboratory | Claudio Tavares Sacchi, Claudia Regina Gonçalves, Erica Valessa Ramos Gomes |
| EPI_ISL_527863 | Hospital Municipal do Tatuape Carmino Caricchio | Instituto Adolfo Lutz, Interdiciplinary Procedures Center, Strategic Laboratory | Claudio Tavares Sacchi, Claudia Regina Gonçalves, Erica Valessa Ramos Gomes |
| EPI_ISL_527865 | Hospital e Maternidade São Cristóvão | Instituto Adolfo Lutz, Interdiciplinary Procedures Center, Strategic Laboratory | Claudio Tavares Sacchi, Claudia Regina Gonçalves, Erica Valessa Ramos Gomes |
| EPI_ISL_527866 | PS Municipal Dr Lauro Ribas Braga | Instituto Adolfo Lutz, Interdiciplinary Procedures Center, Strategic Laboratory | Av. Dr. Arnaldo, 355 - Brazil, Cerqueira Cesar, São Paulo - SP, 01246-1301 |
| EPI_ISL_527868 | Hospital e Maternidade do Braz | Instituto Adolfo Lutz, Interdiciplinary Procedures Center, Strategic Laboratory | Claudio Tavares Sacchi, Claudia Regina Gonçalves, Erica Valessa Ramos Gomes |
| EPI_ISL_527870 | Hospital Municipal Mário Gatti | Instituto Adolfo Lutz, Interdiciplinary Procedures Center, Strategic Laboratory | Claudio Tavares Sacchi, Claudia Regina Gonçalves, Erica Valessa Ramos Gomes |
| EPI_ISL_534311 | UPA III 26 de Agosto | Instituto Adolfo Lutz, Interdiciplinary Procedures Center, Strategic Laboratory | Claudio Tavares Sacchi, Claudia Regina Gonçalves, Erica Valessa Ramos Gomes |
| EPI_ISL_534314 | Hospital Universitario da USP de SP | Instituto Adolfo Lutz, Interdiciplinary Procedures Center, Strategic Laboratory | Claudio Tavares Sacchi, Claudia Regina Gonçalves, Erica Valessa Ramos Gomes |
| EPI_ISL_534316 | OS Mun Santana Lauro Ribas Braga | Instituto Adolfo Lutz, Interdiciplinary Procedures Center, Strategic Laboratory | Claudio Tavares Sacchi, Claudia Regina Gonçalves, Erica Valessa Ramos Gomes |
| EPI_ISL_534317 | Hospital Geral de Itapevi | Instituto Adolfo Lutz, Interdiciplinary Procedures Center, Strategic Laboratory | Claudio Tavares Sacchi, Claudia Regina Gonçalves, Erica Valessa Ramos Gomes |
| EPI_ISL_534318 | Hospital Municipal Antonio Giglio | Instituto Adolfo Lutz, Interdiciplinary Procedures Center, Strategic Laboratory | Claudio Tavares Sacchi, Claudia Regina Gonçalves, Erica Valessa Ramos Gomes |
| EPI_ISL_534319, EPI_ISL_534320 | Hospital do Serv Pub ESTAFCO Morato de Oliveira | Instituto Adolfo Lutz, Interdiciplinary Procedures Center, Strategic Laboratory | Claudio Tavares Sacchi, Claudia Regina Gonçalves, Erica Valessa Ramos Gomes |
| EPI_ISL_534321 | PS e Maternidade Nair Fonseca Leitao Arantes | Instituto Adolfo Lutz, Interdiciplinary Procedures Center, Strategic Laboratory | Claudio Tavares Sacchi, Claudia Regina Gonçalves, Erica Valessa Ramos Gomes |
| EPI_ISL_534322 | PS Mun Julio Tupy | Instituto Adolfo Lutz, Interdiciplinary Procedures Center, Strategic Laboratory | Claudio Tavares Sacchi, Claudia Regina Gonçalves, Erica Valessa Ramos Gomes |
| EPI_ISL_534326 | Notre Dame Intermedica Saude AS | Instituto Adolfo Lutz, Interdiciplinary Procedures Center, Strategic Laboratory | Claudio Tavares Sacchi, Claudia Regina Gonçalves, Erica Valessa Ramos Gomes |
| EPI_ISL_541343, EPI_ISL_541344 | Laboratório Central de Saúde Pública do Estado do Paraná (LACEN-PR) | Laboratory of Respiratory Viruses and Measles, Oswaldo Cruz Institute, FIOCRUZ | Paola Resende, Luciana Appolinario, Fernando Motta, Anna Carolina Paixão, Ana Carolina Mendonça, Jonathan Lopes, Irina Riediger, Maria do Carmo Debur, Marilda Siqueira on behalf of the Fiocruz COVID-19 Genomic Surveillance Network |
| EPI_ISL_541354, EPI_ISL_541355 | Laboratory of Respiratory Viruses and Measles, Oswaldo Cruz Institute, FIOCRUZ | Laboratory of Respiratory Viruses and Measles, Oswaldo Cruz Institute, FIOCRUZ | Paola Resende, Luciana Appolinario, Fernando Motta, Anna Carolina Paixão, Ana Carolina Mendonça, Jonathan Lopes, Marilda Siqueira on behalf of the Fiocruz COVID-19 Genomic Surveillance Network |
| EPI_ISL_541359 | Laboratory of Respiratory Viruses and Measles, Oswaldo Cruz Institute, FIOCRUZ | Laboratory of Respiratory Viruses and Measles, Oswaldo Cruz Institute, FIOCRUZ | Paola Resende, Roxana Loayza, Cinthia Avila, Luciana Appolinario, Fernando Motta, Anna Carolina Paixao, Ana Carolina Mendonca, Marilda Siqueira on behalf of the Fiocruz COVID-19 Genomic Surveillance Network |
| EPI_ISL_541372, EPI_ISL_541386 | Laboratório Central de Saúde Pública do Estado de Sergipe (LACEN-SE) | Laboratory of Respiratory Viruses and Measles, Oswaldo Cruz Institute, FIOCRUZ | Paola Resende, Luciana Appolinario, Fernando Motta, Anna Carolina Paixão, Ana Carolina Mendonça, Jonathan Lopes, Clioma Santos, Marilda Siqueira on behalf of the Fiocruz COVID-19 Genomic Surveillance Network |
| EPI_ISL_547573 | Vigilância em Saúde de Cajamar | Instituto Adolfo Lutz, Interdiciplinary Procedures Center, Strategic Laboratory | Claudio Tavares Sacchi, Claudia Regina Gonçalves, Erica Valessa Ramos Gomes, Karoline Rodrigues Campos |
| EPI_ISL_547575 | SVO Jundiaí | Instituto Adolfo Lutz, Interdiciplinary Procedures Center, Strategic Laboratory | Claudio Tavares Sacchi, Claudia Regina Gonçalves, Erica Valessa Ramos Gomes, Karoline Rodrigues Campos |
| EPI_ISL_547576 | Secretaria Municipal de Saúde | Instituto Adolfo Lutz, Interdiciplinary Procedures Center, Strategic Laboratory | Claudio Tavares Sacchi, Claudia Regina Gonçalves, Erica Valessa Ramos Gomes, Karoline Rodrigues Campos |
| EPI_ISL_547579 | Santa Casa de Misericórdia de Araçatuba | Instituto Adolfo Lutz, Interdiciplinary Procedures Center, Strategic Laboratory | Claudio Tavares Sacchi, Claudia Regina Gonçalves, Erica Valessa Ramos Gomes, Karoline Rodrigues Campos |
| EPI_ISL_572371 | Laboratório Central de Saúde Pública do Estado de Pernambuco (LACEN-PE) | WallauLab, Aggeu Magalhaes Institute | Marcelo Henrique Santos Paiva, Duschinka Ribeiro Duarte Guedes, Cássia Docena, Matheus Filgueira Bezerra, Filipe Zimmer Dezordi, Laís Ceschini Machado, Larissa Krokovsky, Elisama Helvecio, Alexandre Freitas da Silva, Luydson Richardson Silva Vasconcelos, Antonio Mauro Rezende, Severino Jefferson Ribeiro da Silva, Kamila Gaudêncio da Silva Sales, Bruna Santos Lima Figueiredo de Sá, Derciliano Lopes da Cruz, Claudio Eduardo |

| | | | |
|---|---|---|---|
| | | | Cavalcanti, Armando de Menezes Neto, Caroline Targino Alves da Silva, Renata Pessôa Germano Mendes, Maria Almerice Lopes da Silva, Tiago Gräf, Paola Cristina Resende, Gonzalo Bello, Michelle da Silva Barros, Wheverton Ricardo Correia do Nascimento, Rodrigo Moraes Loyo Arcoverde, Luciane Caroline Albuquerque Bezerra, Sinval Pinto Brandão Filho, Constância Flávia Junqueira Ayres, Gabriel Luz Wallau on behalf of the Fiocruz COVID-19 Genomic Surveillance Network |
| EPI_ISL_574577 | Hospital Municipal Dr. Ignacio Proença de Gouvea | Instituto Adolfo Lutz, Interdiciplinary Procedures Center, Strategic Laboratory | Claudio Tavares Sacchi, Claudia Regina Gonçalves, Erica Valessa Ramos Gomes, Karoline Rodrigues Campos |
| EPI_ISL_574578 | Hospital Municipal Mário Gatti | Instituto Adolfo Lutz, Interdiciplinary Procedures Center, Strategic Laboratory | Claudio Tavares Sacchi, Claudia Regina Gonçalves, Erica Valessa Ramos Gomes, Karoline Rodrigues Campos |
| EPI_ISL_574579 | Hospital Municipal Dr. Ignacio Proença de Gouvea | Instituto Adolfo Lutz, Interdiciplinary Procedures Center, Strategic Laboratory | Claudio Tavares Sacchi, Claudia Regina Gonçalves, Erica Valessa Ramos Gomes, Karoline Rodrigues Campos |
| EPI_ISL_574580 | Hospital Cidade Tiradentes Carmen Prudente | Instituto Adolfo Lutz, Interdiciplinary Procedures Center, Strategic Laboratory | Claudio Tavares Sacchi, Claudia Regina Gonçalves, Erica Valessa Ramos Gomes, Karoline Rodrigues Campos |
| EPI_ISL_574583 | Secretaria Municipal de Saude de Jandira | Instituto Adolfo Lutz, Interdiciplinary Procedures Center, Strategic Laboratory | Claudio Tavares Sacchi, Claudia Regina Gonçalves, Erica Valessa Ramos Gomes, Karoline Rodrigues Campos |
| EPI_ISL_574588 | Hospital Estadual Sumare | Instituto Adolfo Lutz, Interdiciplinary Procedures Center, Strategic Laboratory | Claudio Tavares Sacchi, Claudia Regina Gonçalves, Erica Valessa Ramos Gomes, Karoline Rodrigues Campos |
| EPI_ISL_574589 | Hospital Municipal Dr. Jose Soares Hungria | Instituto Adolfo Lutz, Interdiciplinary Procedures Center, Strategic Laboratory | Claudio Tavares Sacchi, Claudia Regina Gonçalves, Erica Valessa Ramos Gomes, Karoline Rodrigues Campos |
| EPI_ISL_574590 | Unidade de Printo Atendimento UPA I Santa Isabel | Instituto Adolfo Lutz, Interdiciplinary Procedures Center, Strategic Laboratory | Claudio Tavares Sacchi, Claudia Regina Gonçalves, Erica Valessa Ramos Gomes, Karoline Rodrigues Campos |
| EPI_ISL_574591, EPI_ISL_574592 | Hospital Domingos Leonardo Ceravolo Presidente Prudente | Instituto Adolfo Lutz, Interdiciplinary Procedures Center, Strategic Laboratory | Claudio Tavares Sacchi, Claudia Regina Gonçalves, Erica Valessa Ramos Gomes, Karoline Rodrigues Campos |
| EPI_ISL_574594 | Hospital Escola da Universidade de Taubate | Instituto Adolfo Lutz, Interdiciplinary Procedures Center, Strategic Laboratory | Claudio Tavares Sacchi, Claudia Regina Gonçalves, Erica Valessa Ramos Gomes, Karoline Rodrigues Campos |
| EPI_ISL_574595 | Hospital Geral de Vila Penteado Dr. Jose Pamgella | Instituto Adolfo Lutz, Interdiciplinary Procedures Center, Strategic Laboratory | Claudio Tavares Sacchi, Claudia Regina Gonçalves, Erica Valessa Ramos Gomes, Karoline Rodrigues Campos |
| EPI_ISL_574597 | Secretaria Municipal de Saude de Jarinu | Instituto Adolfo Lutz, Interdiciplinary Procedures Center, Strategic Laboratory | Claudio Tavares Sacchi, Claudia Regina Gonçalves, Erica Valessa Ramos Gomes, Karoline Rodrigues Campos |
| EPI_ISL_574598 | Servico de Verificacao de Obito SVO | Instituto Adolfo Lutz, Interdiciplinary Procedures Center, Strategic Laboratory | Claudio Tavares Sacchi, Claudia Regina Gonçalves, Erica Valessa Ramos Gomes, Karoline Rodrigues Campos |
| EPI_ISL_583490 | Hospital Estadual Sumare | Instituto Adolfo Lutz, Interdiciplinary Procedures Center, Strategic Laboratory | Claudio Tavares Sacchi, Claudia Regina Gonçalves, Erica Valessa Ramos Gomes, Karoline Rodrigues Campos |
| EPI_ISL_583492 | Santa Casa Anna Cintra | Instituto Adolfo Lutz, Interdiciplinary Procedures Center, Strategic Laboratory | Claudio Tavares Sacchi, Claudia Regina Gonçalves, Erica Valessa Ramos Gomes, Karoline Rodrigues Campos |
| EPI_ISL_583494 | CS II Dr. Antonio Vicoso Moreira de Rezende Sumare | Instituto Adolfo Lutz, Interdiciplinary Procedures Center, Strategic Laboratory | Claudio Tavares Sacchi, Claudia Regina Gonçalves, Erica Valessa Ramos Gomes, Karoline Rodrigues Campos |
| EPI_ISL_583496 | UPA Jandira | Instituto Adolfo Lutz, Interdiciplinary Procedures Center, Strategic Laboratory | Claudio Tavares Sacchi, Claudia Regina Gonçalves, Erica Valessa Ramos Gomes, Karoline Rodrigues Campos |
| EPI_ISL_583497 | Complexo Hospitalar Ouro Verde de Campinas | Instituto Adolfo Lutz, Interdiciplinary Procedures Center, Strategic Laboratory | Claudio Tavares Sacchi, Claudia Regina Gonçalves, Erica Valessa Ramos Gomes, Karoline Rodrigues Campos |
| EPI_ISL_583498 | Hospital Municipal Dr. Waldemar Tebaldi | Instituto Adolfo Lutz, Interdiciplinary Procedures Center, Strategic Laboratory | Claudio Tavares Sacchi, Claudia Regina Gonçalves, Erica Valessa Ramos Gomes, Karoline Rodrigues Campos |
| EPI_ISL_583499 | Distrito Sanitario Sul Campinas | Instituto Adolfo Lutz, Interdiciplinary Procedures Center, Strategic Laboratory | Claudio Tavares Sacchi, Claudia Regina Gonçalves, Erica Valessa Ramos Gomes, Karoline Rodrigues Campos |
| EPI_ISL_583500 | Centro de Saude I Tacito Leite de Carvalho e Silva | Instituto Adolfo Lutz, Interdiciplinary Procedures Center, Strategic Laboratory | Claudio Tavares Sacchi, Claudia Regina Gonçalves, Erica Valessa Ramos Gomes, Karoline Rodrigues Campos |
| EPI_ISL_583501 | Hospital Estadual de CampanhaCOVID 19 Barradas | Instituto Adolfo Lutz, Interdiciplinary Procedures Center, Strategic Laboratory | Claudio Tavares Sacchi, Claudia Regina Gonçalves, Erica Valessa Ramos Gomes, Karoline Rodrigues Campos |
| EPI_ISL_583502 | Serv de Vig Sanitaria Epidemio e CTRL de Zoonoses Guaruja | Instituto Adolfo Lutz, Interdiciplinary Procedures Center, Strategic Laboratory | Claudio Tavares Sacchi, Claudia Regina Gonçalves, Erica Valessa Ramos Gomes, Karoline Rodrigues Campos |
| EPI_ISL_583503 | CTA Centro de Testagem e Aconselhamento | Instituto Adolfo Lutz, Interdiciplinary Procedures Center, Strategic Laboratory | Claudio Tavares Sacchi, Claudia Regina Gonçalves, Erica Valessa Ramos Gomes, Karoline Rodrigues Campos |
| EPI_ISL_583504, EPI_ISL_583505 | Casa de Saude Stella Maris | Instituto Adolfo Lutz, Interdiciplinary Procedures Center, Strategic Laboratory | Claudio Tavares Sacchi, Claudia Regina Gonçalves, Erica Valessa Ramos Gomes, Karoline Rodrigues Campos |
| EPI_ISL_603021 | Pronto Socorro Dr. Conrado Cesarino Nuvolini | Instituto Adolfo Lutz, Interdiciplinary Procedures Center, Strategic Laboratory | Claudio Tavares Sacchi, Claudia Regina Gonçalves, Erica Valessa Ramos Gomes, Karoline Rodrigues Campos |
| EPI_ISL_603023 | Vigilância em Saúde Visa Sul | Instituto Adolfo Lutz, Interdiciplinary Procedures Center, Strategic Laboratory | Claudio Tavares Sacchi, Claudia Regina Gonçalves, Erica Valessa Ramos Gomes, Karoline Rodrigues Campos |
| EPI_ISL_603024 | Santa Casa de Misericordia de Araçatuba | Instituto Adolfo Lutz, Interdiciplinary Procedures Center, Strategic Laboratory | Claudio Tavares Sacchi, Claudia Regina Gonçalves, Erica Valessa Ramos Gomes, Karoline Rodrigues Campos |
| EPI_ISL_603028 | Hospital Municipal Santa Ana | Instituto Adolfo Lutz, Interdiciplinary Procedures Center, Strategic Laboratory | Claudio Tavares Sacchi, Claudia Regina Gonçalves, Erica Valessa Ramos Gomes, Karoline Rodrigues Campos |
| EPI_ISL_603030 | Hospital Domingos Leonardo Ceravolo Presidente Prudente | Instituto Adolfo Lutz, Interdiciplinary Procedures Center, Strategic Laboratory | Claudio Tavares Sacchi, Claudia Regina Gonçalves, Erica Valessa Ramos Gomes, Karoline Rodrigues Campos |
| EPI_ISL_603033 | Vigilancia Epidemiologica de São Bernardo do Campo | Instituto Adolfo Lutz, Interdiciplinary Procedures Center, Strategic Laboratory | Claudio Tavares Sacchi, Claudia Regina Gonçalves, Erica Valessa Ramos Gomes, Karoline Rodrigues Campos |
| EPI_ISL_603034 | Departamento de Vigilância à Saúde | Instituto Adolfo Lutz, Interdiciplinary Procedures Center, Strategic Laboratory | Claudio Tavares Sacchi, Claudia Regina Gonçalves, Erica Valessa Ramos Gomes, Karoline Rodrigues Campos |
| EPI_ISL_603035 | Secretaria Municipal de Saúde | Instituto Adolfo Lutz, Interdiciplinary Procedures Center, Strategic Laboratory | Claudio Tavares Sacchi, Claudia Regina Gonçalves, Erica Valessa Ramos Gomes, Karoline Rodrigues Campos |
| EPI_ISL_603036 | Hospital Santa Ana | Instituto Adolfo Lutz, Interdiciplinary Procedures Center, | Claudio Tavares Sacchi, Claudia Regina Gonçalves, Erica Valessa Ramos Gomes, Karoline Rodrigues Campos |

| | | | |
|---|---|---|---|
| | | Strategic Laboratory | |
| EPI_ISL_603037 | Hospital Geral de Pedreira | Instituto Adolfo Lutz, Interdiciplinary Procedures Center, Strategic Laboratory | Claudio Tavares Sacchi, Claudia Regina Gonçalves, Erica Valessa Ramos Gomes, Karoline Rodrigues Campos |
| EPI_ISL_603038 | Santa Casa de Misericordia de Araçatuba | Instituto Adolfo Lutz, Interdiciplinary Procedures Center, Strategic Laboratory | Claudio Tavares Sacchi, Claudia Regina Gonçalves, Erica Valessa Ramos Gomes, Karoline Rodrigues Campos |
| EPI_ISL_623130 | Laboratorio de Virologia Molecular / UFRJ | Bioinformatics Laboratory / LNCC | Carolina M Voloch, Ronaldo S Francisco Jr, Luiz G P de Almeida, Otavio J. Brustolini, Cynthia C Cardoso, Alexandra L Gerber, Ana Paula de C Guimarães, Diana Mariani, Covid19-UFRJ Workgroup, Luís Cristóvão Pôrto, Renato S Aguiar, Terezinha M P P Castiñeiras, Orlando C. Ferreira, Amilcar Tanuri, Ana Tereza R de Vasconcelos |
| EPI_ISL_672705, EPI_ISL_672711, EPI_ISL_672719, EPI_ISL_672720, EPI_ISL_672748 | Institute of Tropical Medicine at the University of São Paulo (IMT-USP) | Laboratório de Parasitologia Médica - Instituto de Medicina Tropical - Universidade de São Paulo | Brazil-UK Centre for Arbovirus Discovery Diagnosis Genomics and Epidemiology (CADDE) Genomic Network - Instituto de Medicina Tropical |
| EPI_ISL_693195 | Hospital e Pronto Socorro Portinari | Instituto Adolfo Lutz, Interdiciplinary Procedures Center, Strategic Laboratory | Claudio Tavares Sacchi, Claudia Regina Gonçalves, Erica Valessa Ramos Gomes, Karoline Rodrigues Campos |
| EPI_ISL_693196 | Hospital Santa Clara | Instituto Adolfo Lutz, Interdiciplinary Procedures Center, Strategic Laboratory | Claudio Tavares Sacchi, Claudia Regina Gonçalves, Erica Valessa Ramos Gomes, Karoline Rodrigues Campos |
| EPI_ISL_693198 | Santa Casa de Misericordia de Sao Paulo - Hospital Central | Instituto Adolfo Lutz, Interdiciplinary Procedures Center, Strategic Laboratory | Claudio Tavares Sacchi, Claudia Regina Gonçalves, Erica Valessa Ramos Gomes, Karoline Rodrigues Campos |
| EPI_ISL_693199 | Hospital do Servidor Publico Estadual Francisco Morato de Oliveira | Instituto Adolfo Lutz, Interdiciplinary Procedures Center, Strategic Laboratory | Claudio Tavares Sacchi, Claudia Regina Gonçalves, Erica Valessa Ramos Gomes, Karoline Rodrigues Campos |
| EPI_ISL_693200 | Hospital e Maternidade Mairipora | Instituto Adolfo Lutz, Interdiciplinary Procedures Center, Strategic Laboratory | Claudio Tavares Sacchi, Claudia Regina Gonçalves, Erica Valessa Ramos Gomes, Karoline Rodrigues Campos |
| EPI_ISL_693201 | Hospital Sao Paulo de Ensino da Unifesp | Instituto Adolfo Lutz, Interdiciplinary Procedures Center, Strategic Laboratory | Claudio Tavares Sacchi, Claudia Regina Gonçalves, Erica Valessa Ramos Gomes, Karoline Rodrigues Campos |
| EPI_ISL_693202 | Pronto Socorro Municipal Prof. Joao Catarin Mezomo | Instituto Adolfo Lutz, Interdiciplinary Procedures Center, Strategic Laboratory | Claudio Tavares Sacchi, Claudia Regina Gonçalves, Erica Valessa Ramos Gomes, Karoline Rodrigues Campos |
| EPI_ISL_693203 | Hospital Municipal Doutor Arthur Ribeiro de Saboya | Instituto Adolfo Lutz, Interdiciplinary Procedures Center, Strategic Laboratory | Claudio Tavares Sacchi, Claudia Regina Gonçalves, Erica Valessa Ramos Gomes, Karoline Rodrigues Campos |
| EPI_ISL_693204 | Pronto Socorro Dr. Conrado Cesarino Nuvolini | Instituto Adolfo Lutz, Interdiciplinary Procedures Center, Strategic Laboratory | Claudio Tavares Sacchi, Claudia Regina Gonçalves, Erica Valessa Ramos Gomes, Karoline Rodrigues Campos |
| EPI_ISL_693205 | Hospital de Campanha Covid-19 Assis | Instituto Adolfo Lutz, Interdiciplinary Procedures Center, Strategic Laboratory | Claudio Tavares Sacchi, Claudia Regina Gonçalves, Erica Valessa Ramos Gomes, Karoline Rodrigues Campos |
| EPI_ISL_693206 | Hospital Municipal Mario Gatti | Instituto Adolfo Lutz, Interdiciplinary Procedures Center, Strategic Laboratory | Claudio Tavares Sacchi, Claudia Regina Gonçalves, Erica Valessa Ramos Gomes, Karoline Rodrigues Campos |
| EPI_ISL_693207 | Cs II Doutor Antonio Vicoso Moreira de Rezende | Instituto Adolfo Lutz, Interdiciplinary Procedures Center, Strategic Laboratory | Claudio Tavares Sacchi, Claudia Regina Gonçalves, Erica Valessa Ramos Gomes, Karoline Rodrigues Campos |
| EPI_ISL_693208, EPI_ISL_693209 | Hospital Municipal Antonio Giglio | Instituto Adolfo Lutz, Interdiciplinary Procedures Center, Strategic Laboratory | Claudio Tavares Sacchi, Claudia Regina Gonçalves, Erica Valessa Ramos Gomes, Karoline Rodrigues Campos |
| EPI_ISL_693210 | Pronto-Socorro Dr. Osmar Mesquita | Instituto Adolfo Lutz, Interdiciplinary Procedures Center, Strategic Laboratory | Claudio Tavares Sacchi, Claudia Regina Gonçalves, Erica Valessa Ramos Gomes, Karoline Rodrigues Campos |
| EPI_ISL_693211 | Santa Casa de Misericordia e Maternidade | Instituto Adolfo Lutz, Interdiciplinary Procedures Center, Strategic Laboratory | Claudio Tavares Sacchi, Claudia Regina Gonçalves, Erica Valessa Ramos Gomes, Karoline Rodrigues Campos |
| EPI_ISL_693212 | Santa Casa de Misericordia de Braganca Paulista | Instituto Adolfo Lutz, Interdiciplinary Procedures Center, Strategic Laboratory | Claudio Tavares Sacchi, Claudia Regina Gonçalves, Erica Valessa Ramos Gomes, Karoline Rodrigues Campos |
| EPI_ISL_693214 | Unidade de Pronto Atendimento Central de Caraguatatuba | Instituto Adolfo Lutz, Interdiciplinary Procedures Center, Strategic Laboratory | Claudio Tavares Sacchi, Claudia Regina Gonçalves, Erica Valessa Ramos Gomes, Karoline Rodrigues Campos |
| EPI_ISL_693215 | Secretaria Municipal de Saúde de Iracemapolis | Instituto Adolfo Lutz, Interdiciplinary Procedures Center, Strategic Laboratory | Claudio Tavares Sacchi, Claudia Regina Gonçalves, Erica Valessa Ramos Gomes, Karoline Rodrigues Campos |
| EPI_ISL_693216, EPI_ISL_693217 | Unidade de Vigilância Epiudemiológica de Araras | Instituto Adolfo Lutz, Interdiciplinary Procedures Center, Strategic Laboratory | Claudio Tavares Sacchi, Claudia Regina Gonçalves, Erica Valessa Ramos Gomes, Karoline Rodrigues Campos |
| EPI_ISL_693220 | Laboratório Municipal de Piracicaba | Instituto Adolfo Lutz, Interdiciplinary Procedures Center, Strategic Laboratory | Claudio Tavares Sacchi, Claudia Regina Gonçalves, Erica Valessa Ramos Gomes, Karoline Rodrigues Campos |
| EPI_ISL_693221 | Secretaria Municipal de Saúde de Birigui | Instituto Adolfo Lutz, Interdiciplinary Procedures Center, Strategic Laboratory | Claudio Tavares Sacchi, Claudia Regina Gonçalves, Erica Valessa Ramos Gomes, Karoline Rodrigues Campos |
| EPI_ISL_693223, EPI_ISL_693224 | Laboratório Municipal de Piracicaba | Instituto Adolfo Lutz, Interdiciplinary Procedures Center, Strategic Laboratory | Claudio Tavares Sacchi, Claudia Regina Gonçalves, Erica Valessa Ramos Gomes, Karoline Rodrigues Campos |
| EPI_ISL_693225 | Ubs Vila Rosa - Olímpia Gomes De Almeida | Instituto Adolfo Lutz, Interdiciplinary Procedures Center, Strategic Laboratory | Claudio Tavares Sacchi, Claudia Regina Gonçalves, Erica Valessa Ramos Gomes, Karoline Rodrigues Campos |
| EPI_ISL_693226 | Unidade de Pronto Atendimento Sao José | Instituto Adolfo Lutz, Interdiciplinary Procedures Center, Strategic Laboratory | Claudio Tavares Sacchi, Claudia Regina Gonçalves, Erica Valessa Ramos Gomes, Karoline Rodrigues Campos |
| EPI_ISL_693228 | Secretaria Municipal de Sorocaba | Instituto Adolfo Lutz, Interdiciplinary Procedures Center, Strategic Laboratory | Claudio Tavares Sacchi, Claudia Regina Gonçalves, Erica Valessa Ramos Gomes, Karoline Rodrigues Campos |
| EPI_ISL_693229 | Hospital 8 de Maio | Instituto Adolfo Lutz, Interdiciplinary Procedures Center, Strategic Laboratory | Claudio Tavares Sacchi, Claudia Regina Gonçalves, Erica Valessa Ramos Gomes, Karoline Rodrigues Campos |
| EPI_ISL_693230 | Hospital e Pronto Socorro Portinari | Instituto Adolfo Lutz, Interdiciplinary Procedures Center, Strategic Laboratory | Claudio Tavares Sacchi, Claudia Regina Gonçalves, Erica Valessa Ramos Gomes, Karoline Rodrigues Campos |
| EPI_ISL_693231 | Pronto Socorro Municipal de Santa Branca | Instituto Adolfo Lutz, Interdiciplinary Procedures Center, Strategic Laboratory | Claudio Tavares Sacchi, Claudia Regina Gonçalves, Erica Valessa Ramos Gomes, Karoline Rodrigues Campos |
| EPI_ISL_693232 | Hospital e Pronto Socorro Portinari | Instituto Adolfo Lutz, Interdiciplinary Procedures Center, Strategic Laboratory | Claudio Tavares Sacchi, Claudia Regina Gonçalves, Erica Valessa Ramos Gomes, Karoline Rodrigues Campos |
| EPI_ISL_693233 | Hospital Santa Cruz | Instituto Adolfo Lutz, Interdiciplinary Procedures Center, Strategic Laboratory | Claudio Tavares Sacchi, Claudia Regina Gonçalves, Erica Valessa Ramos Gomes, Karoline Rodrigues Campos |

| | | | |
|---|---|---|---|
| EPI_ISL_693234 | Upa Vereador Jose Da Rocha Goncalves | Instituto Adolfo Lutz, Interdiciplinary Procedures Center, Strategic Laboratory | Claudio Tavares Sacchi, Claudia Regina Gonçalves, Erica Valessa Ramos Gomes, Karoline Rodrigues Campos |
| EPI_ISL_693235 | Casmi Centro Atendimento Saude da Mulher e Infancia | Instituto Adolfo Lutz, Interdiciplinary Procedures Center, Strategic Laboratory | Claudio Tavares Sacchi, Claudia Regina Gonçalves, Erica Valessa Ramos Gomes, Karoline Rodrigues Campos |
| EPI_ISL_693236 | Hospital Santa Marcelina Sao Paulo | Instituto Adolfo Lutz, Interdiciplinary Procedures Center, Strategic Laboratory | Claudio Tavares Sacchi, Claudia Regina Gonçalves, Erica Valessa Ramos Gomes, Karoline Rodrigues Campos |
| EPI_ISL_693237 | UPA Santa Isabel | Instituto Adolfo Lutz, Interdiciplinary Procedures Center, Strategic Laboratory | Claudio Tavares Sacchi, Claudia Regina Gonçalves, Erica Valessa Ramos Gomes, Karoline Rodrigues Campos |
| EPI_ISL_693238, EPI_ISL_693239 | Secao Centro de Diagnostico Secedi | Instituto Adolfo Lutz, Interdiciplinary Procedures Center, Strategic Laboratory | Claudio Tavares Sacchi, Claudia Regina Gonçalves, Erica Valessa Ramos Gomes, Karoline Rodrigues Campos |
| EPI_ISL_693240 | Centro de Vigilância a Saude de Diadema | Instituto Adolfo Lutz, Interdiciplinary Procedures Center, Strategic Laboratory | Claudio Tavares Sacchi, Claudia Regina Gonçalves, Erica Valessa Ramos Gomes, Karoline Rodrigues Campos |
| EPI_ISL_693241 | Hospital e Maternidade Sao Lucas | Instituto Adolfo Lutz, Interdiciplinary Procedures Center, Strategic Laboratory | Claudio Tavares Sacchi, Claudia Regina Gonçalves, Erica Valessa Ramos Gomes, Karoline Rodrigues Campos |
| EPI_ISL_693242 | Centro de Vigilância a Saude de Diadema | Instituto Adolfo Lutz, Interdiciplinary Procedures Center, Strategic Laboratory | Claudio Tavares Sacchi, Claudia Regina Gonçalves, Erica Valessa Ramos Gomes, Karoline Rodrigues Campos |
| EPI_ISL_693243 | Laboratório Municipal de Piracicaba | Instituto Adolfo Lutz, Interdiciplinary Procedures Center, Strategic Laboratory | Claudio Tavares Sacchi, Claudia Regina Gonçalves, Erica Valessa Ramos Gomes, Karoline Rodrigues Campos |
| EPI_ISL_693244 | Centro Médico da Polícia Militar do Estado de Sao Paulo | Instituto Adolfo Lutz, Interdiciplinary Procedures Center, Strategic Laboratory | Claudio Tavares Sacchi, Claudia Regina Gonçalves, Erica Valessa Ramos Gomes, Karoline Rodrigues Campos |
| EPI_ISL_693245 | UPA Santa Isabel | Instituto Adolfo Lutz, Interdiciplinary Procedures Center, Strategic Laboratory | Claudio Tavares Sacchi, Claudia Regina Gonçalves, Erica Valessa Ramos Gomes, Karoline Rodrigues Campos |
| EPI_ISL_708530 | Secretaria Municipal de Saude de Fernandópolis | Instituto Adolfo Lutz, Interdiciplinary Procedures Center, Strategic Laboratory | Claudio Tavares Sacchi, Claudia Regina Gonçalves, Erica Valessa Ramos Gomes, Carlos Henrique Camargo, Karoline Rodrigues Campos, Fernanda Modesto Tolentino Binhardi, Maricelia Navarro Pinheiro Flores, Marcia Maria Costa Nunes Soares, Janaina Other Martins Montanha |
| EPI_ISL_717807, EPI_ISL_717808, EPI_ISL_717810, EPI_ISL_717811, EPI_ISL_717812, EPI_ISL_717813, EPI_ISL_717814, EPI_ISL_717815, EPI_ISL_717818, EPI_ISL_717819, EPI_ISL_717820, EPI_ISL_717821, EPI_ISL_717822, EPI_ISL_717823, EPI_ISL_717824, EPI_ISL_717825, EPI_ISL_717826, EPI_ISL_717827, EPI_ISL_717828, EPI_ISL_717829, EPI_ISL_717830 | | | |
| see above | Laboratorio de Virologia Molecular / UFRJ | Bioinformatics Laboratory / LNCC | Carolina M Voloch, Ronaldo da Silva F Jr, Luiz G P de Almeida, Cynthia C Cardoso, Otavio Bustrolini, Alexandra L Gerber, Ana Paula de C Guimarães, Diana Mariani, Andréa Cony Cavalcanti, Claudia dos Santos Rodrigues, Terezinha M P P Castiñeira, Amilcar Tanuri, Ana Tereza R de Vasconcelos |
| EPI_ISL_729801, EPI_ISL_729803, EPI_ISL_729805, EPI_ISL_729806, EPI_ISL_729808, EPI_ISL_729813, EPI_ISL_729840, EPI_ISL_729845, EPI_ISL_729852, EPI_ISL_729853, EPI_ISL_729854, EPI_ISL_729856, EPI_ISL_729861 | | | |
| see above | Laboratório Central de Saúde Pública do Estado do Rio Grande do Sul (LACEN-RS) | Laboratory of Respiratory Viruses and Measles, Oswaldo Cruz Institute, FIOCRUZ | Paola Resende, Luciana Appolinario, Fernando Motta, Anna Carolina Paixão, Ana Carolina Mendonça, Tatiana Schaffer Gregianini, Marilda Tereza Mar da Rosa, Marilda Siqueira on behalf of the Fiocruz COVID-19 Genomic Surveillance Network |
| EPI_ISL_735396 | Hospital de Camplanha COVID 19 SER | Instituto Adolfo Lutz, Interdiciplinary Procedures Center, Strategic Laboratory | Claudio Tavares Sacchi, Claudia Regina Gonçalves, Erica Valessa Ramos Gomes, Karoline Rodrigues Campos |
| EPI_ISL_735397 | Unidade Respiratória Nova Hortolandia | Instituto Adolfo Lutz, Interdiciplinary Procedures Center, Strategic Laboratory | Claudio Tavares Sacchi, Claudia Regina Gonçalves, Erica Valessa Ramos Gomes, Karoline Rodrigues Campos |
| EPI_ISL_735398 | Laboratorio Fleury | Instituto Adolfo Lutz, Interdiciplinary Procedures Center, Strategic Laboratory | Claudio Tavares Sacchi, Claudia Regina Gonçalves, Erica Valessa Ramos Gomes, Karoline Rodrigues Campos |
| EPI_ISL_735399 | Hospital Municiplal Dr Ignacio de gouvea | Instituto Adolfo Lutz, Interdiciplinary Procedures Center, Strategic Laboratory | Claudio Tavares Sacchi, Claudia Regina Gonçalves, Erica Valessa Ramos Gomes, Karoline Rodrigues Campos |
| EPI_ISL_735400 | Instituto Adolfo Lutz - Regional de Santos | Instituto Adolfo Lutz, Interdiciplinary Procedures Center, Strategic Laboratory | Claudio Tavares Sacchi, Claudia Regina Gonçalves, Erica Valessa Ramos Gomes, Karoline Rodrigues Campos |
| EPI_ISL_735401, EPI_ISL_735402, EPI_ISL_735403, EPI_ISL_735404 | Instituto Adolfo Lutz - Regional de Rio Claro | Instituto Adolfo Lutz, Interdiciplinary Procedures Center, Strategic Laboratory | Claudio Tavares Sacchi, Claudia Regina Gonçalves, Erica Valessa Ramos Gomes, Karoline Rodrigues Campos |
| EPI_ISL_735405 | Secretaria Minucipal de Saude de Birigui | Instituto Adolfo Lutz, Interdiciplinary Procedures Center, Strategic Laboratory | Claudio Tavares Sacchi, Claudia Regina Gonçalves, Erica Valessa Ramos Gomes, Karoline Rodrigues Campos |
| EPI_ISL_735406 | Unidade de Pronto Atendimento UPA I Sta Isabel | Instituto Adolfo Lutz, Interdiciplinary Procedures Center, Strategic Laboratory | Claudio Tavares Sacchi, Claudia Regina Gonçalves, Erica Valessa Ramos Gomes, Karoline Rodrigues Campos |
| EPI_ISL_735408 | COVID 19 Centro de Combate ao Coronavirus CCC Jandira | Instituto Adolfo Lutz, Interdiciplinary Procedures Center, Strategic Laboratory | Claudio Tavares Sacchi, Claudia Regina Gonçalves, Erica Valessa Ramos Gomes, Karoline Rodrigues Campos |
| EPI_ISL_735409 | Unidade de Pronto Atendimento Carlos Lourenco | Instituto Adolfo Lutz, Interdiciplinary Procedures Center, Strategic Laboratory | Claudio Tavares Sacchi, Claudia Regina Gonçalves, Erica Valessa Ramos Gomes, Karoline Rodrigues Campos |
| EPI_ISL_735411 | Centro de Vigilancia a Saude de Diadema | Instituto Adolfo Lutz, Interdiciplinary Procedures Center, Strategic Laboratory | Claudio Tavares Sacchi, Claudia Regina Gonçalves, Erica Valessa Ramos Gomes, Karoline Rodrigues Campos |
| EPI_ISL_735412 | Hospital e Pronto Socorro Portinari | Instituto Adolfo Lutz, Interdiciplinary Procedures Center, Strategic Laboratory | Claudio Tavares Sacchi, Claudia Regina Gonçalves, Erica Valessa Ramos Gomes, Karoline Rodrigues Campos |
| EPI_ISL_735413 | Militello Centro de Diagnosticos e Biopesquisa Clinica | Instituto Adolfo Lutz, Interdiciplinary Procedures Center, Strategic Laboratory | Claudio Tavares Sacchi, Claudia Regina Gonçalves, Erica Valessa Ramos Gomes, Karoline Rodrigues Campos |
| EPI_ISL_735417 | Unidade de Pronto Atendimento de Agenor de Campos | Instituto Adolfo Lutz, Interdiciplinary Procedures Center, Strategic Laboratory | Claudio Tavares Sacchi, Claudia Regina Gonçalves, Erica Valessa Ramos Gomes, Karoline Rodrigues Campos |
| EPI_ISL_735418 | Hospital Regional do Vale do Paraiba | Instituto Adolfo Lutz, Interdiciplinary Procedures Center, Strategic Laboratory | Claudio Tavares Sacchi, Claudia Regina Gonçalves, Erica Valessa Ramos Gomes, Karoline Rodrigues Campos |
| EPI_ISL_735419 | UBS Alvarenga | Instituto Adolfo Lutz, Interdiciplinary Procedures Center, Strategic Laboratory | Claudio Tavares Sacchi, Claudia Regina Gonçalves, Erica Valessa Ramos Gomes, Karoline Rodrigues Campos |
| EPI_ISL_735421 | UBS Sta Terezinha | Instituto Adolfo Lutz, Interdiciplinary Procedures Center, Strategic Laboratory | Claudio Tavares Sacchi, Claudia Regina Gonçalves, Erica Valessa Ramos Gomes, Karoline Rodrigues Campos |
| EPI_ISL_735422 | UBS Dematchi | Instituto Adolfo Lutz, Interdiciplinary Procedures Center, Strategic Laboratory | Claudio Tavares Sacchi, Claudia Regina Gonçalves, Erica Valessa Ramos Gomes, Karoline Rodrigues Campos |
| EPI_ISL_735424, EPI_ISL_735426 | Centro de Vigilancia a Saude de Diadema | Instituto Adolfo Lutz, Interdiciplinary Procedures Center, Strategic Laboratory | Claudio Tavares Sacchi, Claudia Regina Gonçalves, Erica Valessa Ramos Gomes, Karoline Rodrigues Campos |
| EPI_ISL_735428, EPI_ISL_735429, | Hospital Nipo Brasileiro | Instituto Adolfo Lutz, Interdiciplinary Procedures Center, | Claudio Tavares Sacchi, Claudia Regina Gonçalves, Erica Valessa Ramos Gomes, Karoline Rodrigues Campos |

| | | | |
|---|---|---|---|
| EPI_ISL_735431 | | Strategic Laboratory | |
| EPI_ISL_735433 | Posto de Atendimento Saude Cidade Pasc Cajati | Instituto Adolfo Lutz, Interdiciplinary Procedures Center, Strategic Laboratory | Claudio Tavares Sacchi, Claudia Regina Gonçalves, Erica Valessa Ramos Gomes, Karoline Rodrigues Campos |
| EPI_ISL_755640 | Instituto Adolfo Lutz - Central | Instituto Adolfo Lutz, Interdiciplinary Procedures Center, Strategic Laboratory | Claudio Tavares Sacchi, Claudia Regina Gonçalves, Erica Valessa Ramos Gomes, Karoline Rodrigues Campos |
| EPI_ISL_755641 | Instituto Adolfo Lutz - Regional de Santo Andre | Instituto Adolfo Lutz, Interdiciplinary Procedures Center, Strategic Laboratory | Claudio Tavares Sacchi, Claudia Regina Gonçalves, Erica Valessa Ramos Gomes, Karoline Rodrigues Campos |
| EPI_ISL_755643 | Instituto Adolfo Lutz - Central | Instituto Adolfo Lutz, Interdiciplinary Procedures Center, Strategic Laboratory | Claudio Tavares Sacchi, Claudia Regina Gonçalves, Erica Valessa Ramos Gomes, Karoline Rodrigues Campos |
| EPI_ISL_755644 | Lab LOC - Itapecerica da Serra | Instituto Adolfo Lutz, Interdiciplinary Procedures Center, Strategic Laboratory | Claudio Tavares Sacchi, Claudia Regina Gonçalves, Erica Valessa Ramos Gomes, Karoline Rodrigues Campos |
| EPI_ISL_755647 | Instituto Adolfo Lutz - Regional de Santo Andre | Instituto Adolfo Lutz, Interdiciplinary Procedures Center, Strategic Laboratory | Claudio Tavares Sacchi, Claudia Regina Gonçalves, Erica Valessa Ramos Gomes, Karoline Rodrigues Campos |
| EPI_ISL_755648, EPI_ISL_755650 | Instituto Adolfo Lutz - Regional de Taubate | Instituto Adolfo Lutz, Interdiciplinary Procedures Center, Strategic Laboratory | Claudio Tavares Sacchi, Claudia Regina Gonçalves, Erica Valessa Ramos Gomes, Karoline Rodrigues Campos |
| EPI_ISL_755654 | Instituto Adolfo Lutz - Central | Instituto Adolfo Lutz, Interdiciplinary Procedures Center, Strategic Laboratory | Claudio Tavares Sacchi, Claudia Regina Gonçalves, Erica Valessa Ramos Gomes, Karoline Rodrigues Campos |
| EPI_ISL_755655 | Instituto Adolfo Lutz - Regional de Campinas | Instituto Adolfo Lutz, Interdiciplinary Procedures Center, Strategic Laboratory | Claudio Tavares Sacchi, Claudia Regina Gonçalves, Erica Valessa Ramos Gomes, Karoline Rodrigues Campos |
| EPI_ISL_770555, EPI_ISL_770558, EPI_ISL_770562, EPI_ISL_770569, EPI_ISL_770572, EPI_ISL_770573, EPI_ISL_770576, EPI_ISL_770577, EPI_ISL_770582, EPI_ISL_770585, EPI_ISL_770586, EPI_ISL_770588, EPI_ISL_770590, EPI_ISL_770597, EPI_ISL_770599, EPI_ISL_770600, EPI_ISL_770601, EPI_ISL_770608, EPI_ISL_770609, EPI_ISL_770610, EPI_ISL_770611, EPI_ISL_770614, EPI_ISL_770615, EPI_ISL_770623, EPI_ISL_770626, EPI_ISL_770627, EPI_ISL_770629 | | | |
| see above | Laboratório de Microbiologia Molecular - Universidade FEEVALE | Bioinformatics Laboratory / LNCC | Felipe Benites, Fernando Rosado Spilki, Alana Witt Hansen, Juliane Deise Fleck, Juliana Schons, Meriane Demoliner, Ana Karolina Eisen Antunes, Fagner Henrique Heldt, Larissa Mallmann, Bruna Hermann, Ana Luiza Ziulkoski, Vyctoria Goes, Karoline Schallenberger, Matheus Nunes Weber, Paula Rodrigues de Almeida, Alessandra Pavan Lamarca da Silva, Ronaldo da Silva F Jr , Luiz G P de Almeida, Alexandra L Gerber , Ana Paula de C Guimarães,Ana Tereza R de Vasconcelos |
| EPI_ISL_776750, EPI_ISL_776752, EPI_ISL_776753, EPI_ISL_776755, EPI_ISL_776756 | Instituto Adolfo Lutz - Central | Instituto Adolfo Lutz, Interdiciplinary Procedures Center, Strategic Laboratory | Claudio Tavares Sacchi, Claudia Regina Gonçalves, Erica Valessa Ramos Gomes, Karoline Rodrigues Campos |
| EPI_ISL_776757, EPI_ISL_776758 | Instituto Adolfo Lutz - Regional de Marilia | Instituto Adolfo Lutz, Interdiciplinary Procedures Center, Strategic Laboratory | Claudio Tavares Sacchi, Claudia Regina Gonçalves, Erica Valessa Ramos Gomes, Karoline Rodrigues Campos |
| EPI_ISL_776761 | Instituto Adolfo Lutz - Central | Instituto Adolfo Lutz, Interdiciplinary Procedures Center, Strategic Laboratory | Claudio Tavares Sacchi, Claudia Regina Gonçalves, Erica Valessa Ramos Gomes, Karoline Rodrigues Campos |
| EPI_ISL_776765, EPI_ISL_776766 | Instituto Adolfo Lutz - Regional de Santo Andre | Instituto Adolfo Lutz, Interdiciplinary Procedures Center, Strategic Laboratory | Claudio Tavares Sacchi, Claudia Regina Gonçalves, Erica Valessa Ramos Gomes, Karoline Rodrigues Campos |
| EPI_ISL_776767 | Instituto Adolfo Lutz - Regional de Marilia | Instituto Adolfo Lutz, Interdiciplinary Procedures Center, Strategic Laboratory | Claudio Tavares Sacchi, Claudia Regina Gonçalves, Erica Valessa Ramos Gomes, Karoline Rodrigues Campos |
| EPI_ISL_776768 | Instituto Adolfo Lutz - Regional de Aracatuba | Instituto Adolfo Lutz, Interdiciplinary Procedures Center, Strategic Laboratory | Claudio Tavares Sacchi, Claudia Regina Gonçalves, Erica Valessa Ramos Gomes, Karoline Rodrigues Campos |
| EPI_ISL_776769 | Instituto Adolfo Lutz - Regional de Santo Andre | Instituto Adolfo Lutz, Interdiciplinary Procedures Center, Strategic Laboratory | Claudio Tavares Sacchi, Claudia Regina Gonçalves, Erica Valessa Ramos Gomes, Karoline Rodrigues Campos |
| EPI_ISL_779156, EPI_ISL_779160, EPI_ISL_779161, EPI_ISL_779162, EPI_ISL_779163, EPI_ISL_779165, EPI_ISL_779166, EPI_ISL_779167, EPI_ISL_779168 | Laboratório de Microbiologia Molecular - Universidade FEEVALE | Bioinformatics Laboratory / LNCC | Felipe Benites, Fernando Rosado Spilki, Alana Witt Hansen, Juliane Deise Fleck, Juliana Schons, Meriane Demoliner, Ana Karolina Eisen Antunes, Fagner Henrique Heldt, Larissa Mallmann, Bruna Hermann, Ana Luiza Ziulkoski, Vyctoria Goes, Karoline Schallenberger, Matheus Nunes Weber, Paula Rodrigues de Almeida, Alessandra Pavan Lamarca da Silva, Ronaldo da Silva F Jr , Luiz G P de Almeida, Alexandra L Gerber , Ana Paula de C Guimarães,Ana Tereza R de Vasconcelos |
| EPI_ISL_792101 | Instituto Adolfo Lutz - Central | Instituto Adolfo Lutz, Interdiciplinary Procedures Center, Strategic Laboratory | Claudio Tavares Sacchi, Claudia Regina Gonçalves, Erica Valessa Ramos Gomes, Karoline Rodrigues Campos |
| EPI_ISL_792103 | Instituto Adolfo Lutz - Regional de Santo Andre | Instituto Adolfo Lutz, Interdiciplinary Procedures Center, Strategic Laboratory | Claudio Tavares Sacchi, Claudia Regina Gonçalves, Erica Valessa Ramos Gomes, Karoline Rodrigues Campos |
| EPI_ISL_792104, EPI_ISL_792106, EPI_ISL_792107, EPI_ISL_792108, EPI_ISL_792109, EPI_ISL_792110, EPI_ISL_792111, EPI_ISL_792112, EPI_ISL_792113, EPI_ISL_792114 | Instituto Adolfo Lutz - Central | Instituto Adolfo Lutz, Interdiciplinary Procedures Center, Strategic Laboratory | Claudio Tavares Sacchi, Claudia Regina Gonçalves, Erica Valessa Ramos Gomes, Karoline Rodrigues Campos |
| EPI_ISL_792115, EPI_ISL_792116 | Instituto Adolfo Lutz - Regional de Taubate | Instituto Adolfo Lutz, Interdiciplinary Procedures Center, Strategic Laboratory | Claudio Tavares Sacchi, Claudia Regina Gonçalves, Erica Valessa Ramos Gomes, Karoline Rodrigues Campos |
| EPI_ISL_792605, EPI_ISL_792631, EPI_ISL_792633 | Laboratório Central de Saúde Pública do Estado da Paraíba (LACEN-PB) | Laboratory of Respiratory Viruses and Measles, Oswaldo Cruz Institute, FIOCRUZ | Paola Resende, Luciana Appolinario, Fernando Motta, Anna Carolina Paixao, Ana Carolina Mendonca, João Felipe Bezerra, Romero Henrique Teixeira de Vasconcelos, Dalane Loudal Florentino Teixeira, Thiago Franco de Oliveira Carneiro, Marilda Siqueira on behalf of the Fiocruz COVID-19 Genomic Surveillance Network |
| EPI_ISL_792643 | Laboratório Central de Saúde Pública do Estado de Alagoas (LACEN-AL) | Laboratory of Respiratory Viruses and Measles, Oswaldo Cruz Institute, FIOCRUZ | Paola Resende, Luciana Appolinario, Fernando Motta, Anna Carolina Paixao, Ana Carolina Mendonca, Anderson Brandao Leite, Marilda Siqueira on behalf of the Fiocruz COVID-19 Genomic Surveillance Network |
| EPI_ISL_792647, EPI_ISL_792649, EPI_ISL_792653, EPI_ISL_792654 | Laboratório Central de Saúde Pública do Estado do Paraná (LACEN-PR) | Laboratory of Respiratory Viruses and Measles, Oswaldo Cruz Institute, FIOCRUZ | Paola Resende, Luciana Appolinario, Fernando Motta, Anna Carolina Paixao, Ana Carolina Mendonca, Maria do Carmo Debur, Irina Nastassja Riediger, Marilda Siqueira on behalf of the Fiocruz COVID-19 Genomic Surveillance Network |
| EPI_ISL_801386, EPI_ISL_801387, EPI_ISL_801388, EPI_ISL_801389, EPI_ISL_801390, EPI_ISL_801391, EPI_ISL_801392, EPI_ISL_801393, EPI_ISL_801394, EPI_ISL_801395, EPI_ISL_801396 | | | |
| see above | Laboratorio de Ecologia de Doencas Transmissiveis na Amazonia, Instituto Leonidas e Maria Deane - Fiocruz Amazonia | Laboratorio de Ecologia de Doencas Transmissiveis na Amazonia, Instituto Leonidas e Maria Deane - Fiocruz Amazonia | Valdinete Nascimento, Victor Souza, André Corado, Fernanda Nascimento, George Silva, Ágatha Costa, Debora Duarte, Luciana Gonçalves, Maria Júlia Brandão, Michele Jesus, Felipe Naveca on behalf of the Fiocruz COVID-19 Genomic Surveillance Network |
| EPI_ISL_801397, EPI_ISL_801398, EPI_ISL_801399, EPI_ISL_801400, EPI_ISL_801401, EPI_ISL_801402, EPI_ISL_801403 | Laboratório Central de Saúde Pública do Estado do Amazonas (LACEN-AM) | Laboratorio de Ecologia de Doencas Transmissiveis na Amazonia, Instituto Leonidas e Maria Deane - Fiocruz Amazonia | Valdinete Nascimento, Victor Souza, André Corado, Fernanda Nascimento, George Silva, Ágatha Costa, Debora Duarte, Luciana Gonçalves, Maria Júlia Brandão, Michele Jesus, Felipe Naveca on behalf of the Fiocruz COVID-19 Genomic Surveillance Network |
| EPI_ISL_831645, EPI_ISL_831660, EPI_ISL_831688, EPI_ISL_831689, | Laboratório de Microbiologia Molecular - Universidade FEEVALE | Universidade Federal de Ciências da Saúde de Porto Alegre | Vinícius Bonetti Franceschi, Amanda de Menezes Mayer, Gabriel Dickin Caldana, Carla Andretta Moreira Neves, Patrícia Aline Gröhs Ferrareze, Gabriela Bettella Cybis, Ricardo Ariel Zimerman, Lívia Kmetzsch, Fernando Rosado Spilki, Claudia Elizabeth Thompson |

| | | | |
|---|---|---|---|
| EPI_ISL_831938, EPI_ISL_832009, EPI_ISL_832011 | | | |
| EPI_ISL_833131 | Laboratorio de Ecologia de Doencas Transmissiveis na Amazonia, Instituto Leonidas e Maria Deane - Fiocruz Amazonia | Laboratorio de Ecologia de Doencas Transmissiveis na Amazonia, Instituto Leonidas e Maria Deane - Fiocruz Amazonia | Valdinete Nascimento, Victor Souza, André Corado, Fernanda Nascimento, George Silva, Ágatha Costa, Debora Duarte, Karina Pessoa, Matilde Mejía, Luciana Gonçalves, Maria Júlia Brandão, Michele Jesus, Felipe Naveca on behalf of the Fiocruz COVID-19 Genomic Surveillance Network |
| EPI_ISL_833152, EPI_ISL_833153, EPI_ISL_833154 | Instituto Adolfo Lutz - Central | Instituto Adolfo Lutz, Interdiciplinary Procedures Center, Strategic Laboratory | Claudio Tavares Sacchi, Claudia Regina Gonçalves, Erica Valessa Ramos Gomes, Karoline Rodrigues Campos |
| EPI_ISL_833156 | Instituto Adolfo Lutz - Regional de Sorocaba | Instituto Adolfo Lutz, Interdiciplinary Procedures Center, Strategic Laboratory | Claudio Tavares Sacchi, Claudia Regina Gonçalves, Erica Valessa Ramos Gomes, Karoline Rodrigues Campos |
| EPI_ISL_833157 | Instituto Adolfo Lutz - Regional de Santo Andre | Instituto Adolfo Lutz, Interdiciplinary Procedures Center, Strategic Laboratory | Claudio Tavares Sacchi, Claudia Regina Gonçalves, Erica Valessa Ramos Gomes, Karoline Rodrigues Campos |
| EPI_ISL_833162 | Lab LOC - Itapecerica da Serra | Instituto Adolfo Lutz, Interdiciplinary Procedures Center, Strategic Laboratory | Claudio Tavares Sacchi, Claudia Regina Gonçalves, Erica Valessa Ramos Gomes, Karoline Rodrigues Campos |
| EPI_ISL_833164 | Secretaria Municipal de Saude de Santa Barbara d'oeste | Instituto Adolfo Lutz, Interdiciplinary Procedures Center, Strategic Laboratory | Claudio Tavares Sacchi, Claudia Regina Gonçalves, Erica Valessa Ramos Gomes, Karoline Rodrigues Campos |
| EPI_ISL_833165 | Hospital Samaritano | Instituto Adolfo Lutz, Interdiciplinary Procedures Center, Strategic Laboratory | Claudio Tavares Sacchi, Claudia Regina Gonçalves, Erica Valessa Ramos Gomes, Karoline Rodrigues Campos |
| EPI_ISL_833168 | DB Diagnosticos do Brasil | Instituto Adolfo Lutz, Interdiciplinary Procedures Center, Strategic Laboratory | Claudio Tavares Sacchi, Claudia Regina Gonçalves, Erica Valessa Ramos Gomes, Karoline Rodrigues Campos |
| EPI_ISL_836978 | Irmandade da Santa Casa de Misericordia de Lorena | Instituto Adolfo Lutz, Interdiciplinary Procedures Center, Strategic Laboratory | Claudio Tavares Sacchi, Claudia Regina Gonçalves, Erica Valessa Ramos Gomes, Karoline Rodrigues Campos |
| EPI_ISL_837053 | UBS Darcy Alves e Robalinho | Instituto Adolfo Lutz, Interdiciplinary Procedures Center, Strategic Laboratory | Claudio Tavares Sacchi, Claudia Regina Gonçalves, Erica Valessa Ramos Gomes, Karoline Rodrigues Campos |
| EPI_ISL_837054 | UBS Jose Sabino Ferreira | Instituto Adolfo Lutz, Interdiciplinary Procedures Center, Strategic Laboratory | Claudio Tavares Sacchi, Claudia Regina Gonçalves, Erica Valessa Ramos Gomes, Karoline Rodrigues Campos |
| EPI_ISL_848562, EPI_ISL_848563, EPI_ISL_848565, EPI_ISL_848566, EPI_ISL_848571, EPI_ISL_848582, EPI_ISL_848583, EPI_ISL_848585, EPI_ISL_848587, EPI_ISL_848588, EPI_ISL_848589, EPI_ISL_848590, EPI_ISL_848592, EPI_ISL_848593, EPI_ISL_848595, EPI_ISL_848611, EPI_ISL_848615, EPI_ISL_848617, EPI_ISL_848618, EPI_ISL_848619, EPI_ISL_848620, EPI_ISL_848621, EPI_ISL_848622, EPI_ISL_848623, EPI_ISL_848624, EPI_ISL_848628 | | | |
| see above | Evandro Chagas Institute | Evandro Chagas Institute | Santos, M.C.; Silva, A.M.; Junior, W.D.C.; Barbagelata, L.S.; Ferreira, J.A.; Sousa, E.M.A.; da Silva, P.S.; Pinheiro, K.C.; L.C.; Sousa Junior, E.C. |
| EPI_ISL_861242 | Instituto de Biotecnologia - UNESP-Botucatu-SP | Instituto de Biotecnologia - UNESP-Botucatu-SP | Leila Sabrina Ullmann; Fábio Sossai Possebon, Camila Dantas Malossi, Paula Rahal, Paulo Inacio da Costa, João Pessoa Araújo Jr. |
| EPI_ISL_861625, EPI_ISL_861626, EPI_ISL_861627 | Instituto Adolfo Lutz - Central | Instituto Adolfo Lutz, Interdiciplinary Procedures Center, Strategic Laboratory | Claudio Tavares Sacchi, Claudia Regina Gonçalves, Erica Valessa Ramos Gomes, Karoline Rodrigues Campos |
| EPI_ISL_861628 | Laboratorio Municipal de Guarulhos | Instituto Adolfo Lutz, Interdiciplinary Procedures Center, Strategic Laboratory | Claudio Tavares Sacchi, Claudia Regina Gonçalves, Erica Valessa Ramos Gomes, Karoline Rodrigues Campos |
| EPI_ISL_861629, EPI_ISL_861630, EPI_ISL_861631, EPI_ISL_861632, EPI_ISL_861633, EPI_ISL_861634 | Instituto Adolfo Lutz - Central | Instituto Adolfo Lutz, Interdiciplinary Procedures Center, Strategic Laboratory | Claudio Tavares Sacchi, Claudia Regina Gonçalves, Erica Valessa Ramos Gomes, Karoline Rodrigues Campos |
| EPI_ISL_861636 | Hospital Geral de Sao Mateus São Paulo | Instituto Adolfo Lutz, Interdiciplinary Procedures Center, Strategic Laboratory | Claudio Tavares Sacchi, Claudia Regina Gonçalves, Erica Valessa Ramos Gomes, Karoline Rodrigues Campos |
| EPI_ISL_861639 | Hospital Sao Paulo de Ensino da Unifesp | Instituto Adolfo Lutz, Interdiciplinary Procedures Center, Strategic Laboratory | Claudio Tavares Sacchi, Claudia Regina Gonçalves, Erica Valessa Ramos Gomes, Karoline Rodrigues Campos |
| EPI_ISL_861640, EPI_ISL_861641 | Hospital Municipal Dr. Moyses Deutsch | Instituto Adolfo Lutz, Interdiciplinary Procedures Center, Strategic Laboratory | Claudio Tavares Sacchi, Claudia Regina Gonçalves, Erica Valessa Ramos Gomes, Karoline Rodrigues Campos |
| EPI_ISL_861643 | Instituto Adolfo Lutz - Central | Instituto Adolfo Lutz, Interdiciplinary Procedures Center, Strategic Laboratory | Claudio Tavares Sacchi, Claudia Regina Gonçalves, Erica Valessa Ramos Gomes, Karoline Rodrigues Campos |
| EPI_ISL_861644 | Hospital Santa Virginia | Instituto Adolfo Lutz, Interdiciplinary Procedures Center, Strategic Laboratory | Claudio Tavares Sacchi, Claudia Regina Gonçalves, Erica Valessa Ramos Gomes, Karoline Rodrigues Campos |
| EPI_ISL_861645 | Hospital e Pronto Socorro Comunitario Vila Iolanda | Instituto Adolfo Lutz, Interdiciplinary Procedures Center, Strategic Laboratory | Claudio Tavares Sacchi, Claudia Regina Gonçalves, Erica Valessa Ramos Gomes, Karoline Rodrigues Campos |
| EPI_ISL_861646, EPI_ISL_861647 | Hospital Santa Marcelina Sao Paulo | Instituto Adolfo Lutz, Interdiciplinary Procedures Center, Strategic Laboratory | Claudio Tavares Sacchi, Claudia Regina Gonçalves, Erica Valessa Ramos Gomes, Karoline Rodrigues Campos |
| EPI_ISL_861648 | Hospital e Pronto Socorro Portinari | Instituto Adolfo Lutz, Interdiciplinary Procedures Center, Strategic Laboratory | Claudio Tavares Sacchi, Claudia Regina Gonçalves, Erica Valessa Ramos Gomes, Karoline Rodrigues Campos |
| EPI_ISL_861649 | Hospital Renascença Campinas | Instituto Adolfo Lutz, Interdiciplinary Procedures Center, Strategic Laboratory | Claudio Tavares Sacchi, Claudia Regina Gonçalves, Erica Valessa Ramos Gomes, Karoline Rodrigues Campos |
| EPI_ISL_861650 | Hospital Santa Marcelina Sao Paulo | Instituto Adolfo Lutz, Interdiciplinary Procedures Center, Strategic Laboratory | Claudio Tavares Sacchi, Claudia Regina Gonçalves, Erica Valessa Ramos Gomes, Karoline Rodrigues Campos |
| EPI_ISL_861652 | AMA Wamberto Dias da Costa | Instituto Adolfo Lutz, Interdiciplinary Procedures Center, Strategic Laboratory | Claudio Tavares Sacchi, Claudia Regina Gonçalves, Erica Valessa Ramos Gomes, Karoline Rodrigues Campos |
| EPI_ISL_861654, EPI_ISL_861655 | Hospital Santa Marcelina Sao Paulo | Instituto Adolfo Lutz, Interdiciplinary Procedures Center, Strategic Laboratory | Claudio Tavares Sacchi, Claudia Regina Gonçalves, Erica Valessa Ramos Gomes, Karoline Rodrigues Campos |
| EPI_ISL_861656 | UPA de Jandira | Instituto Adolfo Lutz, Interdiciplinary Procedures Center, Strategic Laboratory | Claudio Tavares Sacchi, Claudia Regina Gonçalves, Erica Valessa Ramos Gomes, Karoline Rodrigues Campos |
| EPI_ISL_861657 | Hospital e Maternidade Sino Brasileiro | Instituto Adolfo Lutz, Interdiciplinary Procedures Center, Strategic Laboratory | Claudio Tavares Sacchi, Claudia Regina Gonçalves, Erica Valessa Ramos Gomes, Karoline Rodrigues Campos |
| EPI_ISL_861658 | Hospital Municipal Antônio Giglio | Instituto Adolfo Lutz, Interdiciplinary Procedures Center, Strategic Laboratory | Claudio Tavares Sacchi, Claudia Regina Gonçalves, Erica Valessa Ramos Gomes, Karoline Rodrigues Campos |
| EPI_ISL_861659, EPI_ISL_861660, EPI_ISL_861661 | PS e Maternidade Nair Fonseca Leitao Arantes | Instituto Adolfo Lutz, Interdiciplinary Procedures Center, Strategic Laboratory | Claudio Tavares Sacchi, Claudia Regina Gonçalves, Erica Valessa Ramos Gomes, Karoline Rodrigues Campos |
| EPI_ISL_861663 | Instituto Adolfo Lutz - Central | Instituto Adolfo Lutz, Interdiciplinary Procedures Center, Strategic Laboratory | Claudio Tavares Sacchi, Claudia Regina Gonçalves, Erica Valessa Ramos Gomes, Karoline Rodrigues Campos |

| | | | |
|---|---|---|---|
| EPI_ISL_861666 | PSF Dr. Antonio Pires de Almeida | Instituto Adolfo Lutz, Interdiciplinary Procedures Center, Strategic Laboratory | Claudio Tavares Sacchi, Claudia Regina Gonçalves, Erica Valessa Ramos Gomes, Karoline Rodrigues Campos |
| EPI_ISL_861667 | Instituto Adolfo Lutz - Regional de Rio Claro | Instituto Adolfo Lutz, Interdiciplinary Procedures Center, Strategic Laboratory | Claudio Tavares Sacchi, Claudia Regina Gonçalves, Erica Valessa Ramos Gomes, Karoline Rodrigues Campos |
| EPI_ISL_861669 | Lab LOC - Itapecerica da Serra | Instituto Adolfo Lutz, Interdiciplinary Procedures Center, Strategic Laboratory | Claudio Tavares Sacchi, Claudia Regina Gonçalves, Erica Valessa Ramos Gomes, Karoline Rodrigues Campos |
| EPI_ISL_861671 | Hospital Municipal Prefeito Waldemar Costa Filho | Instituto Adolfo Lutz, Interdiciplinary Procedures Center, Strategic Laboratory | Claudio Tavares Sacchi, Claudia Regina Gonçalves, Erica Valessa Ramos Gomes, Karoline Rodrigues Campos |
| EPI_ISL_861673 | PA Novo Osasco | Instituto Adolfo Lutz, Interdiciplinary Procedures Center, Strategic Laboratory | Claudio Tavares Sacchi, Claudia Regina Gonçalves, Erica Valessa Ramos Gomes, Karoline Rodrigues Campos |
| EPI_ISL_861680 | Hospital e Pronto Socorro Portinari | Instituto Adolfo Lutz, Interdiciplinary Procedures Center, Strategic Laboratory | Claudio Tavares Sacchi, Claudia Regina Gonçalves, Erica Valessa Ramos Gomes, Karoline Rodrigues Campos |
| EPI_ISL_861682 | UPA Vila Santa Catarina | Instituto Adolfo Lutz, Interdiciplinary Procedures Center, Strategic Laboratory | Claudio Tavares Sacchi, Claudia Regina Gonçalves, Erica Valessa Ramos Gomes, Karoline Rodrigues Campos |
| EPI_ISL_861867, EPI_ISL_861868, EPI_ISL_861873, EPI_ISL_861875, EPI_ISL_861876, EPI_ISL_861879, EPI_ISL_861885, EPI_ISL_861886, EPI_ISL_861890, EPI_ISL_861892, EPI_ISL_861894, EPI_ISL_861895, EPI_ISL_861896, EPI_ISL_861900, EPI_ISL_861901, EPI_ISL_861902, EPI_ISL_861903, EPI_ISL_861905, EPI_ISL_861906, EPI_ISL_861909, EPI_ISL_861912, EPI_ISL_861913 | | | |
| see above | LATE - Laboratório de Técnicas Especiais - Hospital Israelita Albert Einstein | LATE - Laboratório de Técnicas Especiais - Hospital Israelita Albert Einstein | Deyvid Amgarten, Fernanda de Mello Malta, Raquel Riyuzo, Ana Paula Moreira Salles, Pedro Henrique Sebe Rodrigues, João Renato Rebello Pinho |
| EPI_ISL_861914 | Genomika Einstein | LATE - Laboratório de Técnicas Especiais - Hospital Israelita Albert Einstein | Deyvid Amgarten, Fernanda de Mello Malta, Raquel Riyuzo, Ana Paula Moreira Salles, Pedro Henrique Sebe Rodrigues, João Bosco Oliveira Filho, João Renato Rebello Pinho |
| EPI_ISL_875540, EPI_ISL_875541, EPI_ISL_875542, EPI_ISL_875543, EPI_ISL_875544, EPI_ISL_875545, EPI_ISL_875546, EPI_ISL_875547, EPI_ISL_875548, EPI_ISL_875549, EPI_ISL_875550 | | | |
| see above | Instituto de Biotecnologia - UNESP-Botucatu-SP | Instituto de Biotecnologia - UNESP-Botucatu-SP | Leila Sabrina Ullmann; Fábio Sossai Possebon, Camila Dantas Malossi, Paula Rahal, Paulo Inacio da Costa, João Pessoa Araújo Jr. |
| EPI_ISL_882658 | Secretaria Municipal de Saude | Instituto Adolfo Lutz, Interdiciplinary Procedures Center, Strategic Laboratory | Claudio Tavares Sacchi, Claudia Regina Gonçalves, Erica Valessa Ramos Gomes, Karoline Rodrigues Campos |
| EPI_ISL_882659 | Centro de Triagem Covid19 | Instituto Adolfo Lutz, Interdiciplinary Procedures Center, Strategic Laboratory | Claudio Tavares Sacchi, Claudia Regina Gonçalves, Erica Valessa Ramos Gomes, Karoline Rodrigues Campos |
| EPI_ISL_882660 | Hospital Municipal Prefeito Waldemar Costa Filho | Instituto Adolfo Lutz, Interdiciplinary Procedures Center, Strategic Laboratory | Claudio Tavares Sacchi, Claudia Regina Gonçalves, Erica Valessa Ramos Gomes, Karoline Rodrigues Campos |
| EPI_ISL_882661, EPI_ISL_882662 | Hospital de Santa Barbara de Goias | Instituto Adolfo Lutz, Interdiciplinary Procedures Center, Strategic Laboratory | Claudio Tavares Sacchi, Claudia Regina Gonçalves, Erica Valessa Ramos Gomes, Karoline Rodrigues Campos |
| EPI_ISL_882665 | Unidade de Pronto Atendimento Dra Zilda Arns | Instituto Adolfo Lutz, Interdiciplinary Procedures Center, Strategic Laboratory | Claudio Tavares Sacchi, Claudia Regina Gonçalves, Erica Valessa Ramos Gomes, Karoline Rodrigues Campos |
| EPI_ISL_882672 | Hospital Municipal Dr. Guido Guida | Instituto Adolfo Lutz, Interdiciplinary Procedures Center, Strategic Laboratory | Claudio Tavares Sacchi, Claudia Regina Gonçalves, Erica Valessa Ramos Gomes, Karoline Rodrigues Campos |
| EPI_ISL_888671, EPI_ISL_888672 | Instituto de Biotecnologia - UNESP-Botucatu-SP | Instituto de Biotecnologia - UNESP-Botucatu-SP | Leila Sabrina Ullmann; Fábio Sossai Possebon, Camila Dantas Malossi, Paula Rahal, Paulo Inacio da Costa, João Pessoa Araújo Jr. |
| EPI_ISL_906065 | Day Hospital de Ermelino Matarazzo | Instituto Adolfo Lutz, Interdiciplinary Procedures Center, Strategic Laboratory | Claudio Tavares Sacchi, Claudia Regina Gonçalves, Erica Valessa Ramos Gomes, Karoline Rodrigues Campos |
| EPI_ISL_906066 | Hospital Nipo Brasileiro | Instituto Adolfo Lutz, Interdiciplinary Procedures Center, Strategic Laboratory | Claudio Tavares Sacchi, Claudia Regina Gonçalves, Erica Valessa Ramos Gomes, Karoline Rodrigues Campos |
| EPI_ISL_906067 | PS e Maternidade Nair Fonseca Leitao Arantes | Instituto Adolfo Lutz, Interdiciplinary Procedures Center, Strategic Laboratory | Claudio Tavares Sacchi, Claudia Regina Gonçalves, Erica Valessa Ramos Gomes, Karoline Rodrigues Campos |
| EPI_ISL_918512 | LACEN - Laboratório Central de Saúde Pública do Amazonas | Evandro Chagas Institute | Santos, M.C.; Silva, A.M.; Junior, W.D.C.; Barbagelata, L.S.; Ferreira, J.A.; Sousa, E.M.A.; da Silva, P.S.; Pinheiro, K.C.; L.C.; Sousa Junior, E.C. |
| EPI_ISL_918515 | LACEN - Laboratório Central de Saúde Pública do Para | Evandro Chagas Institute | Santos, M.C.; Silva, A.M.; Junior, W.D.C.; Barbagelata, L.S.; Ferreira, J.A.; Sousa, E.M.A.; da Silva, P.S.; Pinheiro, K.C.; L.C.; Sousa Junior, E.C. |
| EPI_ISL_918518 | Evandro Chagas Institute | Evandro Chagas Institute | Santos, M.C.; Silva, A.M.; Junior, W.D.C.; Barbagelata, L.S.; Ferreira, J.A.; Sousa, E.M.A.; da Silva, P.S.; Pinheiro, K.C.; L.C.; Sousa Junior, E.C. |
| EPI_ISL_918550 | LACEN - Laboratório Central de Saúde Pública do Para | Evandro Chagas Institute | Santos, M.C.; Silva, A.M.; Junior, W.D.C.; Barbagelata, L.S.; Ferreira, J.A.; Sousa, E.M.A.; da Silva, P.S.; Pinheiro, K.C.; L.C.; Sousa Junior, E.C. |
| EPI_ISL_925916, EPI_ISL_926446 | LACEN - Laboratório Central de Saúde Pública do Amazonas | Evandro Chagas Institute Virology | Santos, M.C.; Silva, A.M.; Junior, W.D.C.; Barbagelata, L.S.; Ferreira, J.A.; Sousa, E.M.A.; da Silva, P.S.; Pinheiro, K.C.; L.C.; Sousa Junior, E.C. |
| EPI_ISL_930857 | Central Laboratory of Public Health of Rio Grande do Sul (Lacen-RS) | State Center for Health Surveillance of the Health Department of the State of Rio Grande do Sul (CEVS/SES-RS) | Barcellos R, Campos A, Dornelles C, Godinho F, Gonzalez A, Gregianini T, Molina C, Salvato R, Schaurich A, |
| EPI_ISL_940608 | Laboratório Sao Lucas | Instituto Adolfo Lutz, Interdiciplinary Procedures Center, Strategic Laboratory | Claudio Tavares Sacchi, Claudia Regina Gonçalves, Erica Valessa Ramos Gomes, Karoline Rodrigues Campos |
| EPI_ISL_942898 | Central Laboratory of Public Health of Rio Grande do Sul (Lacen-RS) | State Center for Health Surveillance of the Health Department of the State of Rio Grande do Sul (CEVS/SES-RS) | Barcellos R, Campos A, Crescente L, Da Silva A, Dornelles C, Fonseca V, Garay L, Godinho F, Gonzalez A, Gregianini T, Molina C, Salvato R, Schaurich A |
| EPI_ISL_943581, EPI_ISL_943584, EPI_ISL_943606, EPI_ISL_943609 | Central Laboratory of Public Health of Rio Grande do Sul (Lacen-RS) | State Center for Health Surveillance of the Health Department of the State of Rio Grande do Sul (CEVS/SES-RS) | Aline Campos, Amanda da Silva, Anelise Schaurich, Claudia Dornelles, Cynthia Molina, Fernanda Godinho, Lara Crescente, Leticia Garay, Regina Barcellos, Richard Salvato, Tatiana Gregianini, Vagner Fonseca |
| EPI_ISL_943974, EPI_ISL_943975, EPI_ISL_943976, EPI_ISL_943977, EPI_ISL_943978, EPI_ISL_943979, EPI_ISL_943981, EPI_ISL_943983, EPI_ISL_943985 | LACEN do Estado de Tocantins | Instituto Adolfo Lutz, Interdiciplinary Procedures Center, Strategic Laboratory | Claudio Tavares Sacchi, Claudia Regina Gonçalves, Erica Valessa Ramos Gomes, Karoline Rodrigues Campos |
| EPI_ISL_943988 | LACEN do Estado de Goias | Instituto Adolfo Lutz, Interdiciplinary Procedures Center, Strategic Laboratory | Claudio Tavares Sacchi, Claudia Regina Gonçalves, Erica Valessa Ramos Gomes, Karoline Rodrigues Campos |
| EPI_ISL_943991 | LACEN do Estado de Tocantins | Instituto Adolfo Lutz, Interdiciplinary Procedures Center, Strategic Laboratory | Claudio Tavares Sacchi, Claudia Regina Gonçalves, Erica Valessa Ramos Gomes, Karoline Rodrigues Campos |
| EPI_ISL_977471 | Instituto Adolfo Lutz - Regional de Presidente Prudente | Instituto Adolfo Lutz, Interdiciplinary Procedures Center, Strategic Laboratory | Claudio Tavares Sacchi, Claudia Regina Gonçalves, Erica Valessa Ramos Gomes, Karoline Rodrigues Campos |
| EPI_ISL_977472, EPI_ISL_977473, EPI_ISL_977474 | Instituto Adolfo Lutz Central | Instituto Adolfo Lutz, Interdiciplinary Procedures Center, Strategic Laboratory | Claudio Tavares Sacchi, Claudia Regina Gonçalves, Erica Valessa Ramos Gomes, Karoline Rodrigues Campos |
| EPI_ISL_977475 | Instituto Adolfo Lutz - Regional de Presidente Prudente | Instituto Adolfo Lutz, Interdiciplinary Procedures Center, Strategic Laboratory | Claudio Tavares Sacchi, Claudia Regina Gonçalves, Erica Valessa Ramos Gomes, Karoline Rodrigues Campos |

| | | | |
|---|---|---|---|
| EPI_ISL_977476, EPI_ISL_977477 | Instituto Adolfo Lutz Central | Instituto Adolfo Lutz, Interdiciplinary Procedures Center, Strategic Laboratory | Claudio Tavares Sacchi, Claudia Regina Gonçalves, Erica Valessa Ramos Gomes, Karoline Rodrigues Campos |
| EPI_ISL_977478, EPI_ISL_977480, EPI_ISL_977481 | Instituto Adolfo Lutz - Regional de Presidente Prudente | Instituto Adolfo Lutz, Interdiciplinary Procedures Center, Strategic Laboratory | Claudio Tavares Sacchi, Claudia Regina Gonçalves, Erica Valessa Ramos Gomes, Karoline Rodrigues Campos |
| EPI_ISL_977483, EPI_ISL_977484 | Instituto Adolfo Lutz Central | Instituto Adolfo Lutz, Interdiciplinary Procedures Center, Strategic Laboratory | Claudio Tavares Sacchi, Claudia Regina Gonçalves, Erica Valessa Ramos Gomes, Karoline Rodrigues Campos |
| EPI_ISL_977485 | Instituto Adolfo Lutz - Regional de Presidente Prudente | Instituto Adolfo Lutz, Interdiciplinary Procedures Center, Strategic Laboratory | Claudio Tavares Sacchi, Claudia Regina Gonçalves, Erica Valessa Ramos Gomes, Karoline Rodrigues Campos |
| EPI_ISL_977487 | Instituto Adolfo Lutz Central | Instituto Adolfo Lutz, Interdiciplinary Procedures Center, Strategic Laboratory | Claudio Tavares Sacchi, Claudia Regina Gonçalves, Erica Valessa Ramos Gomes, Karoline Rodrigues Campos |
| EPI_ISL_977488 | Instituto Adolfo Lutz - Regional de Presidente Prudente | Instituto Adolfo Lutz, Interdiciplinary Procedures Center, Strategic Laboratory | Claudio Tavares Sacchi, Claudia Regina Gonçalves, Erica Valessa Ramos Gomes, Karoline Rodrigues Campos |
| EPI_ISL_978498, EPI_ISL_978506, EPI_ISL_978515, EPI_ISL_978517, EPI_ISL_978518, EPI_ISL_978525, EPI_ISL_978529 | Central Public Health Laboratory - LACEN -Bahia, Salvador, Brazil | Central Public Health Laboratory - LACEN -Bahia, Salvador, Brazil | Stephane Tosta, Luciana Oliveira, Vanessa Nardy,Patrícia Cajado,Marcela Gómez, Breno Dominguez, Jaqueline Gomes, Vagner Fonseca,Marta Giovanetti,Luiz Alcantara, Felicidade Pereira, Arabela Leal |
| EPI_ISL_983868 | Central Laboratory of Public Health of Rio Grande do Sul (Lacen-RS) | State Center for Health Surveillance of the Health Department of the State of Rio Grande do Sul (CEVS/SES-RS) | Aline Campos, Cynthia Molina, Lara Crescente, Leticia Garay, Ludmila Fiorenzano Baethgen, Richard Salvato, Tatiana Gregianini |
| EPI_ISL_984242 | Instituto Adolfo Lutz Central | Instituto Adolfo Lutz, Interdiciplinary Procedures Center, Strategic Laboratory | Claudio Tavares Sacchi, Claudia Regina Gonçalves, Erica Valessa Ramos Gomes, Karoline Rodrigues Campos |
| EPI_ISL_984243 | Instituto Adolfo Lutz - Regional de Marilia | Instituto Adolfo Lutz, Interdiciplinary Procedures Center, Strategic Laboratory | Claudio Tavares Sacchi, Claudia Regina Gonçalves, Erica Valessa Ramos Gomes, Karoline Rodrigues Campos |
| EPI_ISL_984246 | Instituto Adolfo Lutz Central | Instituto Adolfo Lutz, Interdiciplinary Procedures Center, Strategic Laboratory | Claudio Tavares Sacchi, Claudia Regina Gonçalves, Erica Valessa Ramos Gomes, Karoline Rodrigues Campos |
| EPI_ISL_984248, EPI_ISL_984253, EPI_ISL_984254 | IAL Regional de Marilia | Instituto Adolfo Lutz, Interdiciplinary Procedures Center, Strategic Laboratory | Claudio Tavares Sacchi, Claudia Regina Gonçalves, Erica Valessa Ramos Gomes, Karoline Rodrigues Campos |
| EPI_ISL_984263 | IAL Regional de Bauru | Instituto Adolfo Lutz, Interdiciplinary Procedures Center, Strategic Laboratory | Claudio Tavares Sacchi, Claudia Regina Gonçalves, Erica Valessa Ramos Gomes, Karoline Rodrigues Campos |
| EPI_ISL_985170 | Instituto Adolfo Lutz - Regional de Presidente Prudente | Instituto Adolfo Lutz, Interdiciplinary Procedures Center, Strategic Laboratory | Claudio Tavares Sacchi, Claudia Regina Gonçalves, Erica Valessa Ramos Gomes, Karoline Rodrigues Campos |
| EPI_ISL_985171, EPI_ISL_985172, EPI_ISL_985173 | Instituto Adolfo Lutz - Regional de Taubate | Instituto Adolfo Lutz, Interdiciplinary Procedures Center, Strategic Laboratory | Claudio Tavares Sacchi, Claudia Regina Gonçalves, Erica Valessa Ramos Gomes, Karoline Rodrigues Campos |
| EPI_ISL_985175 | Instituto Adolfo Lutz Central | Instituto Adolfo Lutz, Interdiciplinary Procedures Center, Strategic Laboratory | Claudio Tavares Sacchi, Claudia Regina Gonçalves, Erica Valessa Ramos Gomes, Karoline Rodrigues Campos |
| EPI_ISL_985178 | Lab Loc - Itapecerica da Serra | Instituto Adolfo Lutz, Interdiciplinary Procedures Center, Strategic Laboratory | Claudio Tavares Sacchi, Claudia Regina Gonçalves, Erica Valessa Ramos Gomes, Karoline Rodrigues Campos |

We gratefully acknowledge the following Authors from the Originating laboratories responsible for obtaining the specimens, as well as the Submitting laboratories where the genome data were generated and shared via GISAID, on which this research is based.

All Submitters of data may be contacted directly via www.gisaid.org

Authors are sorted alphabetically.

| Accession ID | Originating Laboratory | Submitting Laboratory | Authors |
|---|---|---|---|
| EPI_ISL_1019095 | Lighthouse Lab in Alderley Park | Wellcome Sanger Institute for the COVID-19 Genomics UK (COG-UK) Consortium | Jacquelyn Wynn, Mairead Hyland, The Lighthouse Lab in Alderley Park and Alex Alderton, Roberto Amato, Jeffrey Barrett, Sonia Goncalves, Ewan Harrison, David K. Jackson, Ian Johnston, Dominic Kwiatkowski, Cordelia Langford, John Sillitoe on behalf of the Wellcome Sanger Institute COVID-19 Surveillance Team |
| EPI_ISL_1038930 | New Mexico Department of Health Scientific Laboratory | New Mexico Department of Health Scientific Laboratory | Ellie Johnson, Anastacia Griego-Fisher, D'eldra Malone, Jennifer Benoit |
| EPI_ISL_1041312, EPI_ISL_1041411 | Pandemic Response Lab - NYC | Pandemic Response Lab, R&D | Henry Lee, Michael Hammerling, Melissa Hopkins, Cybill del Castillo, William Ward, Pradeep Bugga, Haiping Hao, Jon Laurent |
| EPI_ISL_1061891 | Lighthouse Lab in Alderley Park | Wellcome Sanger Institute for the COVID-19 Genomics UK (COG-UK) Consortium | Jacquelyn Wynn, Mairead Hyland, The Lighthouse Lab in Alderley Park and Alex Alderton, Roberto Amato, Jeffrey Barrett, Sonia Goncalves, Ewan Harrison, David K. Jackson, Ian Johnston, Dominic Kwiatkowski, Cordelia Langford, John Sillitoe on behalf of the Wellcome Sanger Institute COVID-19 Surveillance Team |
| EPI_ISL_1076422 | SYNLAB - Laboratoire J. Collard | GIGA Medical Genomics | Keith Durkin, Maria Artesi, Sébastien Bontems, Raphaël Boreux, Bouchra Boujemla, Nathalie Renotte, Cécile Meex, Pierrette Melin, Marie-Pierre Hayette, Vincent Bours |
| EPI_ISL_1098236 | Pandemic Response Lab - NYC | Pandemic Response Lab, R&D | Henry Lee, Michael Hammerling, Melissa Hopkins, Cybill del Castillo, Shinyoung Clair Kang, William Ward, Pradeep Bugga, Haiping Hao, Jon Laurent |
| EPI_ISL_1113129 | SYNLAB - Laboratoire J. Collard | GIGA Medical Genomics | Keith Durkin, Maria Artesi, Sébastien Bontems, Raphaël Boreux, Bouchra Boujemla, Nathalie Renotte, Cécile Meex, Pierrette Melin, Marie-Pierre Hayette, Vincent Bours |
| EPI_ISL_1163469 | Laboratory Corporation of America | Respiratory Viruses Branch, Division of Viral Diseases, Centers for Disease Control and Prevention | Peter W. Cook, Dakota Howard, Dhwani Batra, Ben L. Rambo-Martin, Minoo Agarwal, Eyad Almasri Debbie Boles, Ayla Burns, Nuthawin Charoensri, Oren Cohen, Susan Countryman, Mary Ann Cristobal, Bobbi Croy, Suzanne Dale, Hrushikesh Deshmukh, Amanda Douglas, Vincent Drouillon, Marcia Eisenberg, Howard Engler, Rama Ghatti, Prashant Gupta, Susan Hicks, Jake Humphrey, Lax Iyer, Manoj Jain, Mohan Kolli, Brian Krueger, Tim Kuphal, Stanley Letovsky, Michael Levandoski, Craig Lukasik, Jonathan Meltzer, Brian Norvell, Mindy Nye, Scott Parker, Christos Petropoulos, John Pruitt, Steven Ragan, Scott Ryan, Mike Sapeta, Jana Schroth, Suresh Babu Selvaraju, Goran Stevovic, Amanda Suchanek, Andrea Throop, Lyndon Tilson, Thomas Urban, Joe Voshell, Kimberly Wagner, Jonathan Williams, Mary Williamson, Qian Zeng, Tricia Zwiefelhofer, Clinton R. Paden, Suxiang Tong, Duncan MacCannell |
| EPI_ISL_1176584, EPI_ISL_1176646, EPI_ISL_1176708 | Department of Pathology, University of Cambridge | COVID-19 Genomics UK (COG-UK) Consortium | Aminu S. Jahun, Yasmin Chaudhry, Iliana Georgana, Myra Hosmillo, Rhys Izuagbe, William L. Hamilton, Martin D. Curran, Surendra Parmar, Ian Goodfellow |
| EPI_ISL_1176828 | Virology Department, Sheffield Teaching Hospitals NHS Foundation Trust/Department of Infection, Immunity and Cardiovascular Disease, The Medical School, University of Sheffield | COVID-19 Genomics UK (COG-UK) Consortium | Thushan de Silva, Matthew Parker, Nikki Smith, Adri Angyal, Rebecca Brown, Luke Green, Rachel Tucker, Paul Parsons, Danielle Groves, Katie Johnson, Laura Carrilero, Alex Keeley, Dave Partridge, Matthew Wyles, Benjamin Lindsey, Mehmet Yavuz, Mohammad Raza, Cariad Evans |
| EPI_ISL_1186053 | New Mexico Department of Health Scientific Laboratory | New Mexico Department of Health Scientific Laboratory | Ellie Johnson, Anastacia Griego-Fisher, D'eldra Malone, Jennifer Benoit |
| EPI_ISL_1187232 | Lighthouse Lab in Cambridge | Wellcome Sanger Institute for the COVID-19 Genomics UK (COG-UK) Consortium | Rob Howes, The Lighthouse Lab in Cambridge and Alex Alderton, Roberto Amato, Jeffrey Barrett, Sonia Goncalves, Ewan Harrison, David K. Jackson, Ian Johnston, Dominic Kwiatkowski, Cordelia Langford, John Sillitoe on behalf of the Wellcome Sanger Institute COVID-19 Surveillance Team |
| EPI_ISL_1195189 | Viollier AG | Department of Biosystems Science and Engineering, ETH Zürich | Chaoran Chen, Sarah Nadeau, Ivan Topolsky, Emmanouil Dermitzakis, Keith Harshman, Ioannis Xenarios, Henri Pegeot, Lorenzo Cerutti, Deborah Penet, Philipp Jablonski, Lara Fuhrmann, David Dreifuss, Katharina Jahn, Christiane Beckmann, Maurice Redondo, Olivier Kobel, Christoph Noppen, Sophie Seidel, Noemie Santamaria de Souza, Niko Beerenwinkel, Tanja Stadler |
| EPI_ISL_1269276 | Helix/Illumina | Centers for Disease Control and Prevention Division of Viral Diseases, Pathogen Discovery | Peter W. Cook, Dakota Howard, Dhwani Batra, Ben L. Rambo-Martin, Eileen de Feo, Jan Antico, Christine Tran, Matthew Tolentino, Shannon Wickline, Kim Gietzen, Brad Sickler, Jingtao Liu, Eric Allen, Phil Febbo, Summer Galloway, Nicole L. Washington, Simon White, Geraint Levan, Kelly Schiabor Barrett, Elizabeth Cirulli, Alexandre Bolze, Ary Ascencio, Charlotte Rivera-Garcia, Ryan Cho, Jason Nguyen, Sherry Wang, Jimmy Ramirez, Tyler Cassens, Efren Sandoval, Magnus Isaksson, William Lee, David Becker, Marc Laurent, James Lu, Clinton R. Paden, Suxiang Tong, Duncan MacCannell |
| EPI_ISL_1369108 | Lab voor klinische biologie | Lab voor klinische biologie | Marija Janevska, Hannelore Hamerlinck, Bruno Verhasselt |
| EPI_ISL_1382711 | KU Leuven, Rega Institute, Clinical and Epidemiological Virology | KU Leuven, Rega Institute, Clinical and Epidemiological Virology | Tony Wawina-Bokalanga, Bert Vanmechelen, Joan Marti-Carerras, Piet Maes |
| EPI_ISL_1460806 | Helix/Illumina | Centers for Disease Control and Prevention Division of Viral Diseases, Pathogen Discovery | Dakota Howard, Dhwani Batra, Peter W. Cook, Kara Moser, Adrian Paskey, Jason Caravas, Benjamin Rambo-Martin, Shatavia Morrison, Christopher Gulvick, Scott Sammons, Yvette Unoarumhi, Darlene Wagner, Matthew Schmerer, Eileen de Feo, Jan Antico, Christine Tran, Matthew Tolentino, Shannon Wickline, Kim Gietzen, Brad Sickler, Jingtao Liu, Eric Allen, Phil Febbo, Nicole L. Washington, Simon White, Geraint Levan, Kelly Schiabor Barrett, Elizabeth Cirulli, Alexandre Bolze, Ary Ascencio, Charlotte Rivera-Garcia, Ryan Cho, Jason Nguyen, Sherry Wang, Jimmy Ramirez, Tyler Cassens, Efren Sandoval, Magnus Isaksson, William Lee, David Becker, Marc Laurent, James Lu, Clinton R. Paden, Duncan MacCannell |
| EPI_ISL_1462890 | Laboratory Corporation of America | Centers for Disease Control and Prevention Division of Viral Diseases, Pathogen Discovery | Dakota Howard, Dhwani Batra, Peter W. Cook, Kara Moser, Adrian Paskey, Jason Caravas, Benjamin Rambo-Martin, Shatavia Morrison, Christopher Gulvick, Scott Sammons, Yvette Unoarumhi, Darlene Wagner, Matthew Schmerer, Minoo Agarwal, Eyad Almasri, Debbie Boles, Ayla Burns, Nuthawin Charoensri, Oren Cohen, Susan Countryman, Mary Ann Cristobal, Bobbi Croy, Suzanne Dale, Hrushikesh Deshmukh, Amanda Douglas, Vincent Drouillon, Marcia Eisenberg, Howard Engler, Rama Ghatti, Prashant Gupta, Susan Hicks, Jake Humphrey, Lax Iyer, Manoj Jain, Mohan Kolli, Brian Krueger, Tim Kuphal, Stanley Letovsky, Michael Levandoski, Craig Lukasik, Jonathan Meltzer, Brian Norvell, Mindy Nye, Scott Parker, Christos Petropoulos, John Pruitt, Steven Ragan, Scott Ryan, Mike Sapeta, Jana Schroth, Suresh Babu Selvaraju, Goran Stevovic, Amanda Suchanek, Andrea Throop, Lyndon Tilson, Thomas Urban, Joe Voshell, Kimberly Wagner, Jonathan Williams, Mary Williamson, Qian Zeng, Tricia Zwiefelhofer, Clinton R. Paden, Duncan MacCannell |
| EPI_ISL_1471516 | Pandemic Response Lab - NYC | Pandemic Response Lab, R&D | Henry Lee, Michael Hammerling, Melissa Hopkins, Cybill del Castillo, Shinyoung Clair Kang, William Ward, Pradeep Bugga, Sol Rey, Dylan Law, Haiping Hao, Jon Laurent |
| EPI_ISL_1520165 | Northwestern Medicine | Illinois Department of Public Health - Chicago Lab | Vineet K. Dhiman, Ira Heimler |
| EPI_ISL_1527453 | GA Department of Public Health | GA Department of Public Health | Stacy Reeves, Jonathan Edwards, Cynthia Dixey, Tonia Parrott, Aliyah Fields, Taylor Smith |
| EPI_ISL_1581351 | Helix/Illumina | Centers for Disease Control and Prevention Division of Viral Diseases, Pathogen Discovery | Dakota Howard, Dhwani Batra, Peter W. Cook, Kara Moser, Adrian Paskey, Jason Caravas, Benjamin Rambo-Martin, Shatavia Morrison, Christopher Gulvick, Scott Sammons, Yvette Unoarumhi, Darlene Wagner, Matthew Schmerer, Eileen de Feo, Jan Antico, Christine Tran, Matthew Tolentino, Shannon Wickline, Kim Gietzen, Brad Sickler, Jingtao Liu, Eric Allen, Phil Febbo, Nicole L. Washington, Simon White, Geraint Levan, Kelly Schiabor Barrett, Elizabeth Cirulli, Alexandre Bolze, Ary Ascencio, Charlotte Rivera-Garcia, Ryan Cho, Jason Nguyen, Sherry Wang, Jimmy Ramirez, Tyler Cassens, Efren Sandoval, Magnus Isaksson, William Lee, David Becker, Marc Laurent, James Lu, Clinton R. Paden, Duncan MacCannell |
| EPI_ISL_1581920, EPI_ISL_1582076 | Quest Diagnostics Incorporated | Centers for Disease Control and Prevention Division of Viral Diseases, Pathogen Discovery | Dakota Howard, Dhwani Batra, Peter W. Cook, Kara Moser, Adrian Paskey, Jason Caravas, Benjamin Rambo-Martin, Shatavia Morrison, Christopher Gulvick, Scott Sammons, Yvette Unoarumhi, Darlene Wagner, Matthew Schmerer, S. H. Rosenthal, A. Gerasimova, R. M. Kagan, B. Anderson, M. Hua, Y. Liu, L.E. Bernstein, K.E. Livingston, A. Perez, I. A. Shlyakhter, R. V. Rolando, R. Owen, P. Tanpaiboon, F. Lacbawan, Clinton R. Paden, Duncan MacCannell |

| | | | |
|---|---|---|---|
| EPI_ISL_1694523 | Infinity Biologix | Centers for Disease Control and Prevention Division of Viral Diseases, Pathogen Discovery | Dakota Howard, Dhwani Batra, Peter W. Cook, Kara Moser, Adrian Paskey, Jason Caravas, Benjamin Rambo-Martin, Shatavia Morrison, Christopher Gulvick, Scott Sammons, Yvette Unoarumhi, Darlene Wagner, Matthew Schmerer, Christian Bixby, Yihe Wang, Jonathan Schultz, Chirayu Goswami, Russ Hager, Robin Grimwood, Clinton R. Paden, Duncan MacCannell |
| EPI_ISL_1788474 | LBM DE LA SAUVEGARDE | CNR Virus des Infections Respiratoires - France SUD | Antonin Bal, Gregory Destras, Gwendolyne Burfin, Hadrien Regue, Quentin Semanas, Martine Valette, Bruno Lina, Laurence Josset |
| EPI_ISL_1796674 | Helix/Illumina | Centers for Disease Control and Prevention Division of Viral Diseases, Pathogen Discovery | Dakota Howard, Dhwani Batra, Peter W. Cook, Kara Moser, Adrian Paskey, Jason Caravas, Benjamin Rambo-Martin, Shatavia Morrison, Christopher Gulvick, Scott Sammons, Yvette Unoarumhi, Darlene Wagner, Matthew Schmerer, Eileen de Feo, Jan Antico, Christine Tran, Matthew Tolentino, Shannon Wickline, Kim Gietzen, Brad Sickler, Jingtao Liu, Eric Allen, Phil Febbo, Nicole L. Washington, Simon White, Geraint Levan, Kelly Schiabor Barrett, Elizabeth Cirulli, Alexandre Bolze, Ary Ascencio, Charlotte Rivera-Garcia, Ryan Cho, Jason Nguyen, Sherry Wang, Jimmy Ramirez, Tyler Cassens, Efren Sandoval, Magnus Isaksson, William Lee, David Becker, Marc Laurent, James Lu, Clinton R. Paden, Duncan MacCannell |
| EPI_ISL_1805190 | Maryland Genomics, Institute for Genome Sciences, University of Maryland School of Medicine | Maryland Genomics, Institute for Genome Sciences, University of Maryland School of Medicine | Tallon, Luke J; Sadzewicz, Lisa D; Humphrys, Mike; Ott, Sandra; Roussey, Holly; Mehta, Aditya; Vavikolanu, Kranthi; Fraser, Claire M; Ravel, Jacques |
| EPI_ISL_1840673 | Helix/Illumina | Centers for Disease Control and Prevention Division of Viral Diseases, Pathogen Discovery | Dakota Howard, Dhwani Batra, Peter W. Cook, Kara Moser, Adrian Paskey, Jason Caravas, Benjamin Rambo-Martin, Shatavia Morrison, Christopher Gulvick, Scott Sammons, Yvette Unoarumhi, Darlene Wagner, Matthew Schmerer, Eileen de Feo, Jan Antico, Christine Tran, Matthew Tolentino, Shannon Wickline, Kim Gietzen, Brad Sickler, Jingtao Liu, Eric Allen, Phil Febbo, Nicole L. Washington, Simon White, Geraint Levan, Kelly Schiabor Barrett, Elizabeth Cirulli, Alexandre Bolze, Ary Ascencio, Charlotte Rivera-Garcia, Ryan Cho, Jason Nguyen, Sherry Wang, Jimmy Ramirez, Tyler Cassens, Efren Sandoval, Magnus Isaksson, William Lee, David Becker, Marc Laurent, James Lu, Clinton R. Paden, Duncan MacCannell |
| EPI_ISL_1922657 | Maryland Genomics, Institute for Genome Sciences, University of Maryland School of Medicine | Maryland Genomics, Institute for Genome Sciences, University of Maryland School of Medicine | Tallon, Luke J; Sadzewicz, Lisa D; Humphrys, Mike; Ott, Sandra; Roussey, Holly; Mehta, Aditya; Vavikolanu, Kranthi; Fraser, Claire M; Ravel, Jacques |
| EPI_ISL_1939070 | Universidad de León | SeqCOVID-SPAIN consortium/IBV(CSIC) | Ana Carvajal, Vicente Martín, Héctor Argüello, Juan M. Fregeneda, Tania Fernández-Villa, Antonio J. Molina and SeqCOVID-SPAIN consortium |
| EPI_ISL_1962939 | Dutch COVID-19 response team | National Institute for Public Health and the Environment (RIVM) | Adam Meijer, Harry Vennema, Dirk Eggink, Jeroen Cremer, Sharon van den Brink, Bas van der Veer, AnneMarie van den Brandt, Lisa Wijsman, Kim Freriks, Ryanne Jaarsma, Euníce Then, Lynn Aarts, Sanne Bos, Melissa van Tuil, Linda van de Nes, Sjoerd Kuiling, James Groot, Florian Zwagemaker, Dennis Schmitz, Annelies Kroneman, Karim Hajji, Chantal Reusken, on behalf of the national COVID-19 response team |
| EPI_ISL_2099708 | National Centre for Disease Control | CDFD -INSACOG | To be addded later |
| EPI_ISL_2316007 | Universitätsklinikum Köln; Institut für Virologie | Robert Koch Institute | unknown |
| EPI_ISL_2321569 | Emory University | Centers for Disease Control and Prevention Division of Viral Diseases, Pathogen Discovery | Mili Sheth, Sarah Nobles, Jasmine Padilla, Mark Burroughs, Shoshona Le, Katie Dillon, Peter Cook, Clinton R. Paden, Dhwani Batra, Krista Queen, Kristen Knipe, Dakota Howard, Yvette Unoarumhi, Darlene Wagner, Matthew Schmerer, Ben L. Rambo-Martin, Kristine Lacek, Sam Shepard, Alison Laufer Halpin, Dave Wentworth, Vivien Dugan, Suxiang Tong, Justin Lee |
| EPI_ISL_2341980, EPI_ISL_2342927 | NCCS, Pune | Institute of Life Sciences - INSACOG | Sunil K. Raghav, Safal Walia, Arup Ghosh, Atimukta Jha, Amol M. Kanampalliwar, Omprakash Shiriwas, Sana Fatma, Shifu Aggarwal, Rupesh Dash, Rajeeb Swain, Punit Prasad, INSACOG Consortium, Ajay Parida |
| EPI_ISL_2427517, EPI_ISL_2427520, EPI_ISL_2427528, EPI_ISL_2427532, EPI_ISL_2427535, EPI_ISL_2427540, EPI_ISL_2427542, EPI_ISL_2427550, EPI_ISL_2427557, EPI_ISL_2427568, EPI_ISL_2427575, EPI_ISL_2427605, EPI_ISL_2427607, EPI_ISL_2427608, EPI_ISL_2427609, EPI_ISL_2427613, EPI_ISL_2427616, EPI_ISL_2427617, EPI_ISL_2427619, EPI_ISL_2427624, EPI_ISL_2427627, EPI_ISL_2427631, EPI_ISL_2427632, EPI_ISL_2427641, EPI_ISL_2427645, EPI_ISL_2427650, EPI_ISL_2427655, EPI_ISL_2427658, EPI_ISL_2427662, EPI_ISL_2427679, EPI_ISL_2427680, EPI_ISL_2427685, EPI_ISL_2427691, EPI_ISL_2427699, EPI_ISL_2427716, EPI_ISL_2427718, EPI_ISL_2427723, EPI_ISL_2427726, EPI_ISL_2427745, EPI_ISL_2427774 | | | |
| see above | Laboratorio de Biología Molecular Médica Uruguaya | Departments of Pathology and Medicine, New York University School of Medicine | Maria Victoria Elizondo, Maria Noel Zubillaga, Gonzalo Manrique, Cecilia Sorhouet, Maria Cristina Mogdasy, Paul Zappile, Dacia Dimartino, Christian Marier, Adriana Heguy |
| EPI_ISL_2517017 | Molecular Microbiology, Washington University of St. Louis | Molecular Microbiology, Washington University of St. Louis | Wang,D., Handley,S. |
| EPI_ISL_2597320, EPI_ISL_2597389 | Hospital of the University of Pennsylvania Molecular Pathology Lab | Bushman Lab - University of Pennsylvania | John K. Everett, Kyle Rodino, Shantan Reddy, Aoife M. Roche, Young Hwang, Scott Sherrill-Mix, Samantha A. Whiteside, Jevon Graham-Wooten, Layla A. Khatib, Ayannah S. Fitzgerald, Arupa Ganguly, Mike Feldman, Brendan Kelly, Ronald G. Collman and Frederic Bushan |
| EPI_ISL_2754030, EPI_ISL_2754031, EPI_ISL_2754032, EPI_ISL_2754073 | Sanatorio Americano | Institut Pasteur de Montevideo | Natalia Rego,Tamara Fernández-Calero,Ighor Arantes,Verónica Noya,Daiana Mir,Mariana Brandes,Juan Zanetti,Mailen Arleo,Emiliano Pereira,Tania Possi,Odhille Chappos,Lucia Bilbao,Natalia Reyes,Melissa Duquía,Matías Victoria,Pía Techera,María José Benítez-Galeano,Luciana Griffero,Mauricio Méndez,Belén González,Pablo Smircich,Andres Lizasoain,Matías Castells,Matías Salvo,Rodney Colina,Cecilia Alonso,Gonzalo Bello,Lucía Spangenberg |
| EPI_ISL_654986 | Lighthouse Lab in Glasgow | Wellcome Sanger Institute for the COVID-19 Genomics UK (COG-UK) Consortium | Harper VanSteenhouse, Yumi Kasai, David Gray, Carol Clugston, Anna Dominiczak and Alex Alderton, Roberto Amato, Sonia Goncalves, Ewan Harrison, David K. Jackson, Ian Johnston, Dominic Kwiatkowski, Cordelia Langford, John Sillitoe on behalf of the Wellcome Sanger Institute COVID-19 Surveillance Team |
| EPI_ISL_656403, EPI_ISL_656489 | Lighthouse Lab in Alderley Park | Wellcome Sanger Institute for the COVID-19 Genomics UK (COG-UK) Consortium | Jacquelyn Wynn, Mairead Hyland, The Lighthouse Lab in Alderley Park and Alex Alderton, Roberto Amato, Sonia Goncalves, Ewan Harrison, David K. Jackson, Ian Johnston, Dominic Kwiatkowski, Cordelia Langford, John Sillitoe on behalf of the Wellcome Sanger Institute COVID-19 Surveillance Team |
| EPI_ISL_661471, EPI_ISL_668167 | Lighthouse Lab in Glasgow | Wellcome Sanger Institute for the COVID-19 Genomics UK (COG-UK) Consortium | Harper VanSteenhouse, Yumi Kasai, David Gray, Carol Clugston, Anna Dominiczak and Alex Alderton, Roberto Amato, Sonia Goncalves, Ewan Harrison, David K. Jackson, Ian Johnston, Dominic Kwiatkowski, Cordelia Langford, John Sillitoe on behalf of the Wellcome Sanger Institute COVID-19 Surveillance Team |
| EPI_ISL_674666, EPI_ISL_674682 | Lighthouse Lab in Milton Keynes | Wellcome Sanger Institute for the COVID-19 Genomics UK (COG-UK) Consortium | The Lighthouse Lab in Milton Keynes and Alex Alderton, Roberto Amato, Sonia Goncalves, Ewan Harrison, David K. Jackson, Ian Johnston, Dominic Kwiatkowski, Cordelia Langford, John Sillitoe on behalf of the Wellcome Sanger Institute COVID-19 Surveillance Team |
| EPI_ISL_702452 | Lighthouse Lab in Alderley Park | Wellcome Sanger Institute for the COVID-19 Genomics UK (COG-UK) Consortium | Jacquelyn Wynn, Mairead Hyland, The Lighthouse Lab in Alderley Park and Alex Alderton, Roberto Amato, Sonia Goncalves, Ewan Harrison, David K. Jackson, Ian Johnston, Dominic Kwiatkowski, Cordelia Langford, John Sillitoe on behalf of the Wellcome Sanger Institute COVID-19 Surveillance Team |
| EPI_ISL_704363, EPI_ISL_704542 | Lighthouse Lab in Glasgow | Wellcome Sanger Institute for the COVID-19 Genomics UK (COG-UK) Consortium | Harper VanSteenhouse, Yumi Kasai, David Gray, Carol Clugston, Anna Dominiczak and Alex Alderton, Roberto Amato, Sonia Goncalves, Ewan Harrison, David K. Jackson, Ian Johnston, Dominic Kwiatkowski, Cordelia Langford, John Sillitoe on behalf of the Wellcome Sanger Institute COVID-19 Surveillance Team |
| EPI_ISL_708889 | Lighthouse Lab in Milton Keynes | Wellcome Sanger Institute for the COVID-19 Genomics UK (COG-UK) Consortium | The Lighthouse Lab in Milton Keynes and Alex Alderton, Roberto Amato, Sonia Goncalves, Ewan Harrison, David K. Jackson, Ian Johnston, Dominic Kwiatkowski, Cordelia Langford, John Sillitoe on behalf of the Wellcome Sanger Institute COVID-19 Surveillance Team |
| EPI_ISL_717702 | Area of Virology, Serology and Virology Division (SAViD), New South Wales Health Pathology Randwick | Virology Research Laboratory; Area of Virology, Serology and Virology Division (SAViD), New South Wales Health Pathology Randwick | Foster, C.; Au, J.; Ruiz Silva, M.; Deveson, I.; Bull, R.; Van Hal, S.; Rawlinson, W. |
| EPI_ISL_719372 | Lighthouse Lab in Cambridge | Wellcome Sanger Institute for the COVID-19 Genomics UK (COG-UK) Consortium | Rob Howes, The Lighthouse Lab in Cambridge and Alex Alderton, Roberto Amato, Sonia Goncalves, Ewan Harrison, David K. Jackson, Ian Johnston, Dominic Kwiatkowski, Cordelia Langford, John Sillitoe on behalf of the Wellcome Sanger Institute COVID-19 Surveillance Team |
| EPI_ISL_720256 | Lighthouse Lab in Milton Keynes | Wellcome Sanger Institute for the COVID-19 Genomics UK (COG-UK) Consortium | The Lighthouse Lab in Milton Keynes and Alex Alderton, Roberto Amato, Sonia Goncalves, Ewan Harrison, David K. Jackson, Ian Johnston, Dominic Kwiatkowski, Cordelia Langford, John Sillitoe on behalf of the Wellcome Sanger Institute COVID-19 Surveillance Team |
| EPI_ISL_720929 | Lighthouse Lab in Glasgow | Wellcome Sanger Institute for the COVID-19 Genomics UK (COG-UK) Consortium | Harper VanSteenhouse, Yumi Kasai, David Gray, Carol Clugston, Anna Dominiczak and Alex Alderton, Roberto Amato, Sonia Goncalves, Ewan Harrison, David K. Jackson, Ian Johnston, Dominic Kwiatkowski, Cordelia Langford, John Sillitoe on behalf of the Wellcome Sanger Institute COVID-19 Surveillance Team |
| EPI_ISL_727818 | Oxford Viromics, NDM, University of Oxford; Oxford University Hospitals; Basingstoke and North Hampshire Hospital | COVID-19 Genomics UK (COG-UK) Consortium | Tanya Golubchik, David Bonsall, George Macintyre, Amy Trebes, Mariateresa de Cesare, Catrin Moore, Alex Mobbs, Anita Justice, Robert Shaw, Monique Andersson, Timothy Peto, Emma Wise, Nathan Moore, Jessica Lynch, Nick Cortes, Matilde Mori, Stephen Kidd, David Buck, John Todd, Christophe Fraser |
| EPI_ISL_735576 | Lighthouse Lab in Milton Keynes | Wellcome Sanger Institute for the COVID-19 Genomics UK (COG-UK) Consortium | The Lighthouse Lab in Milton Keynes and Alex Alderton, Roberto Amato, Sonia Goncalves, Ewan Harrison, David K. Jackson, Ian Johnston, Dominic Kwiatkowski, Cordelia Langford, John Sillitoe on behalf of the Wellcome Sanger Institute COVID-19 Surveillance Team |
| EPI_ISL_736533 | Lighthouse Lab in Glasgow | Wellcome Sanger Institute for the COVID-19 Genomics UK (COG-UK) Consortium | Harper VanSteenhouse, Yumi Kasai, David Gray, Carol Clugston, Anna Dominiczak and Alex Alderton, Roberto Amato, Sonia Goncalves, Ewan Harrison, David K. Jackson, Ian Johnston, Dominic Kwiatkowski, Cordelia Langford, John Sillitoe on behalf of the Wellcome Sanger Institute COVID-19 Surveillance |

| | | | |
|---|---|---|---|
| | | | Team |
| EPI_ISL_740868 | South Eastern Area Laboratory Services (SEALS) | NSW Health Pathology - Institute of Clinical Pathology and Medical Research; Westmead Hospital; University of Sydney | CIDM-PH et al. |
| EPI_ISL_761219 | Lighthouse Lab in Alderley Park | Wellcome Sanger Institute for the COVID-19 Genomics UK (COG-UK) Consortium | Jacquelyn Wynn, Mairead Hyland, The Lighthouse Lab in Alderley Park and Alex Alderton, Roberto Amato, Sonia Goncalves, Ewan Harrison, David K. Jackson, Ian Johnston, Dominic Kwiatkowski, Cordelia Langford, John Sillitoe on behalf of the Wellcome Sanger Institute COVID-19 Surveillance Team |
| EPI_ISL_777460, EPI_ISL_778221 | Lighthouse Lab in Cambridge | Wellcome Sanger Institute for the COVID-19 Genomics UK (COG-UK) Consortium | Rob Howes, The Lighthouse Lab in Cambridge and Alex Alderton, Roberto Amato, Sonia Goncalves, Ewan Harrison, David K. Jackson, Ian Johnston, Dominic Kwiatkowski, Cordelia Langford, John Sillitoe on behalf of the Wellcome Sanger Institute COVID-19 Surveillance Team |
| EPI_ISL_778371, EPI_ISL_778550 | Lighthouse Lab in Milton Keynes | Wellcome Sanger Institute for the COVID-19 Genomics UK (COG-UK) Consortium | The Lighthouse Lab in Milton Keynes and Alex Alderton, Roberto Amato, Sonia Goncalves, Ewan Harrison, David K. Jackson, Ian Johnston, Dominic Kwiatkowski, Cordelia Langford, John Sillitoe on behalf of the Wellcome Sanger Institute COVID-19 Surveillance Team |
| EPI_ISL_779685 | Pathogen Genomics Center, National Institute of Infectious Diseases | Pathogen Genomics Center, National Institute of Infectious Diseases | Tsuyoshi Sekizuka, Kentaro Itokawa, Rina Tanaka, Masanori Hashino, Makoto Kuroda |
| EPI_ISL_804356 | Respiratory Virus Unit, National Infection Service, Public Health England | COVID-19 Genomics UK (COG-UK) Consortium | PHE Covid Sequencing Team |
| EPI_ISL_816841 | Bioinformatics and Biostatistics Lab, Advanced Sequencing Facility | COVID-19 Genomics UK (COG-UK) Consortium | Aengus Stewart,Jerome Nicod,Chelsea Sawyer,Laura Cubitt,Harshil Patel,Margaret Crawford |
| EPI_ISL_825572 | Respiratory Virus Unit, National Infection Service, Public Health England | COVID-19 Genomics UK (COG-UK) Consortium | PHE Covid Sequencing Team |
| EPI_ISL_834664 | Lighthouse Lab in Glasgow | Wellcome Sanger Institute for the COVID-19 Genomics UK (COG-UK) Consortium | Harper VanSteenhouse, Yumi Kasai, David Gray, Carol Clugston, Anna Dominiczak and Alex Alderton, Roberto Amato, Sonia Goncalves, Ewan Harrison, David K. Jackson, Ian Johnston, Dominic Kwiatkowski, Cordelia Langford, John Sillitoe on behalf of the Wellcome Sanger Institute COVID-19 Surveillance Team |
| EPI_ISL_880986, EPI_ISL_885889 | Lighthouse Lab in Alderley Park | Wellcome Sanger Institute for the COVID-19 Genomics UK (COG-UK) Consortium | Jacquelyn Wynn, Mairead Hyland, The Lighthouse Lab in Alderley Park and Alex Alderton, Roberto Amato, Sonia Goncalves, Ewan Harrison, David K. Jackson, Ian Johnston, Dominic Kwiatkowski, Cordelia Langford, John Sillitoe on behalf of the Wellcome Sanger Institute COVID-19 Surveillance Team |
| EPI_ISL_890322 | KU Leuven, Rega Institute, Clinical and Epidemiological Virology | KU Leuven, Rega Institute, Clinical and Epidemiological Virology | Tony Wawina-Bokalanga, Bert Vanmechelen, Joan Marti-Carerras, Piet Maes |
| EPI_ISL_908794, EPI_ISL_908890 | Lighthouse Lab in Cambridge | Wellcome Sanger Institute for the COVID-19 Genomics UK (COG-UK) Consortium | Rob Howes, The Lighthouse Lab in Cambridge and Alex Alderton, Roberto Amato, Sonia Goncalves, Ewan Harrison, David K. Jackson, Ian Johnston, Dominic Kwiatkowski, Cordelia Langford, John Sillitoe on behalf of the Wellcome Sanger Institute COVID-19 Surveillance Team |
| EPI_ISL_913777 | KU Leuven, Rega Institute, Clinical and Epidemiological Virology | KU Leuven, Rega Institute, Clinical and Epidemiological Virology | Tony Wawina-Bokalanga, Bert Vanmechelen, Joan Marti-Carerras, Piet Maes |
| EPI_ISL_917080 | Lighthouse Lab in Alderley Park | Wellcome Sanger Institute for the COVID-19 Genomics UK (COG-UK) Consortium | Jacquelyn Wynn, Mairead Hyland, The Lighthouse Lab in Alderley Park and Alex Alderton, Roberto Amato, Sonia Goncalves, Ewan Harrison, David K. Jackson, Ian Johnston, Dominic Kwiatkowski, Cordelia Langford, John Sillitoe on behalf of the Wellcome Sanger Institute COVID-19 Surveillance Team |
| EPI_ISL_936643 | Northwestern Memorial Hospital | Ozer Lab | Ramon Lorenzo-Redondo, Lacy M. Simons, Chad J. Achenbach, Lawrence J. Jennings, Michael G. Ison, Judd F. Hultquist, Egon A. Ozer |
| EPI_ISL_946464 | Lighthouse Lab in Alderley Park | Wellcome Sanger Institute for the COVID-19 Genomics UK (COG-UK) Consortium | Jacquelyn Wynn, Mairead Hyland, The Lighthouse Lab in Alderley Park and Alex Alderton, Roberto Amato, Sonia Goncalves, Ewan Harrison, David K. Jackson, Ian Johnston, Dominic Kwiatkowski, Cordelia Langford, John Sillitoe on behalf of the Wellcome Sanger Institute COVID-19 Surveillance Team |
| EPI_ISL_957689 | Lighthouse Lab in Cambridge | Wellcome Sanger Institute for the COVID-19 Genomics UK (COG-UK) Consortium | Rob Howes, The Lighthouse Lab in Cambridge and Alex Alderton, Roberto Amato, Sonia Goncalves, Ewan Harrison, David K. Jackson, Ian Johnston, Dominic Kwiatkowski, Cordelia Langford, John Sillitoe on behalf of the Wellcome Sanger Institute COVID-19 Surveillance Team |
| EPI_ISL_958825 | Lighthouse Lab in Milton Keynes | Wellcome Sanger Institute for the COVID-19 Genomics UK (COG-UK) Consortium | The Lighthouse Lab in Milton Keynes and Alex Alderton, Roberto Amato, Sonia Goncalves, Ewan Harrison, David K. Jackson, Ian Johnston, Dominic Kwiatkowski, Cordelia Langford, John Sillitoe on behalf of the Wellcome Sanger Institute COVID-19 Surveillance Team |
| EPI_ISL_963376 | Lighthouse Lab in Glasgow | Wellcome Sanger Institute for the COVID-19 Genomics UK (COG-UK) Consortium | Harper VanSteenhouse, Yumi Kasai, David Gray, Carol Clugston, Anna Dominiczak and Alex Alderton, Roberto Amato, Sonia Goncalves, Ewan Harrison, David K. Jackson, Ian Johnston, Dominic Kwiatkowski, Cordelia Langford, John Sillitoe on behalf of the Wellcome Sanger Institute COVID-19 Surveillance Team |
| EPI_ISL_990891 | Lighthouse Lab in Alderley Park | Wellcome Sanger Institute for the COVID-19 Genomics UK (COG-UK) Consortium | Jacquelyn Wynn, Mairead Hyland, The Lighthouse Lab in Alderley Park and Alex Alderton, Roberto Amato, Sonia Goncalves, Ewan Harrison, David K. Jackson, Ian Johnston, Dominic Kwiatkowski, Cordelia Langford, John Sillitoe on behalf of the Wellcome Sanger Institute COVID-19 Surveillance Team |
